# Effectiveness and efficacy of long-lasting insecticidal nets for malaria control in Africa: Systematic review and meta-analysis of randomized controlled trials

**DOI:** 10.1101/2024.07.31.24311306

**Authors:** Dereje Bayisa Demissie, Getahun Fetensa Hirko, Tilahun Desta, Firew Tiruneh Tiyare

## Abstract

**Background:** Malaria, a parasitic disease that is transmitted by the bite of a female Anopheles mosquito, can also be contracted through exposure to infected blood products or congenitally. Long-lasting insecticidal nets have significantly reduced the malaria burden in the past decade and this malaria prevalence reduction has been achieved through the upgrading of pyrethroid long-lasting insecticidal nets (LLINs), but the reduction has stopped due to pyrethroid fast resistance. The World Health Organization (WHO) recommends new LLINs with two active ingredients for areas with malaria vectors resistant to pyrethroids. Malaria control in Africa faces threat from pyrethroid resistance, prompting the development of new long-lasting insecticidal nets (LLINs) with dual active ingredients to interrupt transmission in pyrethroid-resistant areas. A study aimed to evaluate the effectiveness and efficacy of different mixtures of active-ingredient LLINs compared to standard pyrethroid LLINs against pyrethroid-resistant malaria vectors showed no reduction in the density of Mansonia spp. in the pyrethroid pyriproxyfen LLIN arm outdoors. Therefore, the objective of this systematic review and meta-analysis is to pool estimates of the effectiveness and efficacy of pyriproxyfen, chlorfenapyr, and piperonyl butoxide long-lasting insecticidal nets (LLINs) with pyrethroid-only LLINs for malaria control in African. This study also evaluated the effectiveness and efficacy of chlorfenapyr, and piperonyl butoxide long-lasting insecticidal nets compared to pyriproxyfen LLINs.

**Method:** The protocol was registered in PROSPERO with the protocol number: CRD42024499800. This review used Cochrane methodology to assess risk of bias and evaluate evidence quality. It included cluster randomized or prospective clinical trials comparing long-lasting insecticidal nets (LLINs) of Pyriproxyfen, Chlorfenapyr, and/or Piperonyl Butoxide for malaria control (test arm) and pyrethroid-only standard LLINs (control arm) for malaria control. Three reviewers independently read each preprint or publication and extracted relevant data from individual studies. The search was conducted from 2000 to 2024, and meta-analysis was performed using Excel and STATA 17. The extracted data from eligible studies were pooled using the random effects model and expressed as a risk ratio (RR) with a 95% confidence interval (CI).

**Result:** A total of 11 cluster randomized controlled trials with 21,916 households, 1,145,035 people, and 34,327 children across all of the studies reporting sample size. This study found that the pooled prevalence of post-intervention malaria infection among children using chlorfenapyr, piperonyl butoxide, and pyriproxyfen long-lasting insecticidal nets was 25.58 per 100 children, 32.38 per 100 children, and 33.70 per 100 children, respectively, compared to the control group/pyrethroid-only long-lasting insecticidal nets of 40.84% per 100 children in Africa, which is higher in the control group.

The study found that the post-intervention pooled mean indoor vector density per household per night in the control group/pyrethroid-only long-lasting insecticidal nets was higher than in the intervention groups, with pyrethroid-only nets having the highest density at 8.04 per household per night, compared to other insecticidal nets (7.74 per 100 households in pyriproxyfen, 5.53 per 100 households in chlorfenapyr, and the lowest 1.9 per 100 households per night in piperonyl butoxide) in Africa.

The study determined that the post-intervention pooled sporozoite rate per mosquito in the control group/pyrethroid-only long-lasting insecticidal nets was almost two to three times higher than in the intervention groups, with pyrethroid-only nets having the highest sporozoite rate per mosquito at 227 per 100 anopheles, compared to other interventional long-lasting insecticidal nets (165 per 100 anopheles in pyriproxyfen, 172 per 100 anopheles in piperonyl butoxide, and the lowest 79 per 100 anopheles in chlorfenapyr) in Africa.

A meta-analysis found that pyriproxyfen (PPF) long-lasting insecticidal nets (LLINs) effectively reduce indoor vector density by 1%, entomological inoculation rate by 7%, and sporozoite rate of malaria parasites by 15% compared to pyrethroid-only LLINs in Africa, despite no significant difference in malaria infection, case incidence, and anemia reduction among children.

The study found that piperonyl butoxide (PBO) long-lasting insecticidal nets (LLINs) are highly effective and efficacious in reducing malaria infection by 1%, case incidence by 2%, and anaemia by 3% among children, as well as reducing indoor vector density by 3%, the mean entomological inoculation rate by 12%, and the sporozoite rate by 10% in Africa as compared to pyrethroid-only LLINs in Africa.

The study found that chlorfenapyr (CFP) long-lasting insecticidal nets (LLINs) are highly effective and efficacious in reducing malaria infection by 1%, case incidence by 1%, and anaemia by 4% among children, as well as reducing indoor vector density by 4%, the inoculation rate by 23%, and the sporozoite rate by 9% in Africa as compared to pyrethroid-only LLINs in Africa.

The study compared the effectiveness and efficacy of chlorfenapyr (CFP) and pyriproxyfen long-lasting insecticidal nets in Africa. Results showed that CFP nets were highly effective, and efficacious in reducing malaria infection, case incidence, indoor vector density, inoculation rate, and sporozoite rate by 1%, 15%, and 7%, respectively, compared to pyriproxyfen nets long-lasting insecticidal nets for malaria control in Africa.

The evidence evaluating the effectiveness and efficacy of piperonyl butoxide (PBO) compared with pyriproxyfen long-lasting insecticidal nets found that piperonyl butoxide (PBO) long-lasting insecticidal nets (LLINs) are highly effective and efficacious in reducing malaria infection by 0.0%, case incidence by 2% among children, indoor vector density by 4%, inoculation rate by 5%, and sporozoite rate by 1% in piperonyl butoxide (PBO) as compared to pyriproxyfen long-lasting insecticidal nets for malaria control in Africa. Critical appraisal of individual randomized control trials revealed that 100% of the studies scored high quality, and Cochrane methodology was used to assess the risk of bias and evaluate evidence quality, which was graded as high. This research provides a very good indication of the likely effect. The likelihood that the effect will be substantially different is low.

**Conclusion:** This generated evidence was evaluated the effectiveness and efficacy of pyriproxyfen, chlorfenapyr, and piperonyl butoxide long-lasting insecticidal nets against the pyrethroid-only LLINs.

This study found that PYR-only LLINs (control arm) had higher pooled prevalence of malaria infection, case incidence, anaemia, mean indoor vector density, inoculationrate, and sporozoite rate as compared to intervention group (PPF, CFP, and PBO LLINs

The evidence generated from this meta-analysis reveals that pyriproxyfen (PPF) long-lasting insecticidal nets (LLINs) have no significant difference in malaria infection, case incidence, or anemia reduction among children as compared to pyrethroid-only LLINs. However, this study found that Pyriproxyfen (PPF) LLINs effectively and efficaciously reduce indoor vector density, entomological inoculationrate, and sporozoite rate of malaria parasites compared to pyrethroid-only LLINs.

The study found that chlorfenapyr (CFP) and piperonyl butoxide (PBO) long-lasting insecticidal nets (LLINs) are highly effective and efficacious in reducing malaria infection, case incidence, and anaemia among children, as well as reducing indoor vector density, inoculation rate, and sporozoite rate in Africa as compared to pyrethroid-only LLINs.

The evidence generated found that piperonyl butoxide (PBO) long-lasting insecticidal nets effectively and efficaciously reduce indoor vector density, entomological inoculation rate, and sporozoite rate of malaria parasites compared to Pyriproxyfen (PPF) LLINs, but no significant difference was found in malaria infection reduction among children who use piperonyl butoxide (PBO) versus Pyriproxyfen (PPF) long-lasting insecticidal nets in Africa.

The study found that chlorfenapyr (CFP) long-lasting insecticidal nets (LLINs) are highly effective and superiorly efficacious in reducing malaria infection, case incidence, and anemia among children, as well as reducing mean indoor vector density, mean entomological inoculation rate, and sporozoite rate compared to pyriproxyfen (PPF) long-lasting insecticidal nets (LLINs) in Africa. Therefore, policymakers and health planners should give a great deal of emphasis on addressing the effectiveness, efficacy, and resistance management of long-lasting insecticidal nets (LLINs) as part of their current public health agenda to eliminate malaria.

## Introduction

Over the past 20 years, significant investment in long-lasting insecticidal nets, indoor residual spraying, and artemisinin-based therapies has led to the averted of 1.5 billion malaria cases and 6.6 million deaths (1). Long-lasting insecticidal nets (LLINs) are the foundation of malaria control but resistance of mosquito vectors to pyrethroids threatens their effectiveness (2). The World Health Organization (WHO) reported that vector resistance to insecticides has been observed in 88 malaria endemic countries from 2010 to 2020(3). Globally, resistance to pyrethroids was 64% of sites reporting resistance and with high intensity resistance was detected in 27 countries and 293 sites. Monitoring of insecticide resistance has been adjusted to align with new procedures, including new discriminating concentrations and procedures for chlorfenapyr, clothianidin, transfluthrin, flupyradifurone, and pyriproxyfen (3). Pyrethroid (PY) long-lasting insecticidal nets (LLINs) are the primary malaria control method in sub-Saharan Africa, contributing significantly to the decline in malaria morbidity and mortality. However, the rapid spread of PY resistance in vector populations has hindered the stagnation of this decline (4).

Long-lasting insecticidal nets (LLINs) effect may vary based on type of vector of mosquito species(5). Malaria prevalence reduction has been undertaken by the up grading of pyrethroid long-lasting insecticidal nets (LLINs), however, the decrement of malaria burden stopped which may related with pyrethroid fast resistance (6). World health Organization described as piperonyl butoxide( PBO) LLINs are more effective against malaria than non-PBO LLINs in case of resistance to pyrethroids is high (2). Evidences are indicating that outdoors, no reduction in the density of Mansonia spp was observed in the pyrethroid pyriproxyfen LLIN arm(7).

Malaria control in sub-Saharan Africa faces threat from pyrethroid resistance, prompting the development of new long-lasting insecticidal nets (LLINs) with dual active ingredients to interrupt transmission in pyrethroid-resistant areas (3). A randomize control trial study found that Chlorfenapyr pyrethroid LLINs were the most effective intervention against the main malaria vector, a funestus sl, over three years of community use. Piperonyl-butoxide pyrethroid LLINs had a sustained effect for two years. The other vector, an arabiensis, was not controlled by any LLINs. Additional vector control tools and strategies are needed for further malaria transmission reduction (8). A study in Southern Mali found that deltamethrin + PBO LLINs reduced An. gambiae sporozoite rates during high malaria transmission seasons, but no improvement in parity rates or indoor resting densities. Combination nets may be more effective in areas with mixed function oxidases (9).

New long-lasting insecticidal nets (LLINs) combining insecticide mixtures with various modes of action could potentially regain malaria control in sub-Saharan Africa following resurgence in transmission. A randomized control trial study found that chlorfenapyr-pyrethroid LLINs provided greater protection from malaria than pyrethroid-only LLINs over two years in areas with pyrethroid-resistant mosquitoes but Pyriproxyfen-pyrethroid LLINs provided protection similar to pyrethroid-only LLINs. The study confirms the importance of chlorfenapyr as an LLIN treatment for malaria control in pyrethroid-resistant areas (10). However, more active ingredients are needed for long-term resistance management and effective vector control strategies.

A cluster-randomised controlled trial in southern Benin found that chlorfenapyr-pyrethroid LLINs provided greater protection from malaria over two years than pyrethroid-only LLINs in areas with pyrethroid-resistant mosquitoes. However, Pyriproxyfen-pyrethroid LLINs provided similar protection with pyrethroid-only LLINs. The study confirms the importance of chlorfenapyr as an LLIN treatment for malaria control, but further investigation is needed for effective vector control strategies (10). Another a third-year post-distribution secondary analysis of a cluster randomised controlled trial in southern Benin found that the pyriproxyfen-pyrethroid LLIN group did not provide superior protection against malaria cases compared to the standard LLIN group, and the chlorfenapyr-pyrethroid LLIN group also did not offer superior protection against malaria cases or infections (11). A cluster randomized trial in Tanzania found that malaria incidence was consistently lower in too-torn PBO-PY LLIN and Chlorfenapyr-PY LLIN compared to intact PY-only LLIN during the first year of follow-up. In year 2, incidence was only significantly lower in intact Chlorfenapyr-PY LLIN compared to intact PY LLIN (12). A systematic review and meta-analysis found that chlorfenapyr-pyrethroid insecticide-treated nets are more effective in reducing malaria case incidence and parasite prevalence than pyrethroid-only ITNs. However, only chlorfenapyr-pyrethroid ITNs showed a reduction in these outcomes compared to pyrethroid-PBO ITNs (13). Hence, researchers have followed several approaches to systematically measure the processes of different long-lasting insecticidal nets (LLINs) effectiveness and efficacy for malaria control. These inconsistent approaches to measuring the effectiveness and efficacy of long-lasting insecticidal nets (LLINs) pose challenges to programmers and policymakers. Hence, it is important to obtain a pooled estimate of the effectiveness and efficacy of long-lasting insecticidal nets (LLINs) by comparing different long-lasting insecticidal nets (LLINs) over a range of 6 months to 36 months post-interventions distribution of long-lasting insecticidal nets (LLINs). However, to the best of our knowledge, no systematic review or meta-analysis has been conducted to estimate the pooled effectiveness and efficacy of long-lasting insecticidal nets (LLINs). Therefore, considering the scarcity of evidence on the effectiveness and efficacy of pyriproxyfen, chlorfenapyr, and piperonyl butoxide long-lasting insecticidal nets (LLINs) with pyrethroid-only LLINs for malaria control, therefore, this study aimed to fill this evidence gap. Hence, programmers and policymakers rely on the evidence from their businesses. Furthermore, researchers can gain insights into another research question to further study long-lasting insecticidal nets (LLINs). This systematic review and meta-analysis aimed to enable governments, policymakers, health professionals, and populations at risks of malaria infections to inform themselves about the importance of effectiveness and efficacy long-lasting insecticidal nets (LLINs) and to evaluate changes and trends in the effectiveness and efficacy of pyriproxyfen, chlorfenapyr, and piperonyl butoxide long-lasting insecticidal nets (LLINs) with pyrethroid-only LLINs and the study also compares the effectiveness of chlorfenapyr and piperonyl butoxide LLINs compared to pyriproxyfen LLINs for malaria control over time.

## The objectives of this review were

To conduct a systematic review of long-lasting insecticidal nets (LLINs) effectiveness and efficacy for malaria control

To conduct a meta-analysis to pooled estimates of the effectiveness and efficacy of pyriproxyfen, chlorfenapyr, and piperonyl butoxide long-lasting insecticidal nets (LLINs) with pyrethroid-only LLINs for malaria control in Africa.

To compare the effectiveness and efficacy of pyriproxyfen, chlorfenapyr, and piperonyl butoxide LLINs with pyrethroid-only LLINs for malaria control in Africa.

To evaluate the effectiveness and efficacy of chlorfenapyr and piperonyl butoxide long-lasting insecticidal nets compared to pyriproxyfen LLINs for malaria control in Africa.

## Methods

The protocol of this systematic review and meta-analysis was registered in PROSPERO on January24, 2024 (registration number: CRD42024499800). Available from: https://www.crd.york.ac.uk/prospero/display_record.php?ID=CRD42024499800. This systematic review used Cochrane methodology for systematic reviews of interventional studies to assess risk of bias and evaluate evidence quality (14). The analysis, interpretation, and reporting included a risk of bias assessment using the Cochrane Risk of Bias tool, which assigns studies as having a low, unclear, or high risk of bias. Quality of evidence and strength of recommendation were based on the Grades of Recommendation, Assessment, Development, and Evaluation (GRADE) approach, which involves consideration of methodological quality, directness of evidence, heterogeneity, precision of effect estimates, and publication bias (15).

## Literature search strategy

A systematic literature search was done to identify relevant articles from online databases PubMed, MEDLINE, Embase, and the Cochrane Central Register of Controlled Trials’ database (CENTRAL) for retrieving randomized control trials comparing the effectiveness and efficacy of pyriproxyfen, chlorfenapyr, and piperonyl butoxide long-lasting insecticidal nets (LLINs) compared with pyrethroid-only LLINs for malaria control in Africa. The studies were searched from the following databases, without restriction on the date of publication among all age groups and children aged 5 years or younger were eligible to receive interventions of long-lasting insecticidal nets (LLINs): MEDLINE, Hinari, Scopes, PubMed CINAHL, PopLine, MedNar, Embase, the Cochrane Library, JBI Library, Web of Science, and Google Scholar and reference list of selected articles were also screened for identifying additional potentially eligible studies. Example of search string in PubMed: (((((((((((((((((" Effectiveness AND ("cost-effectiveness analysis"[MeSH] OR "cost effectiveness AND against ("malaria"[MeSH] AND pyriproxyfen "[MeSH] OR, chlorfenapyr "[MeSH] OR, and piperonyl butoxide long-lasting insecticidal nets (LLINs) "[MeSH] OR dual-active-ingredient AND longlasting AND insecticidal AND "nets AND LLINs AND (compared pyrethroid-only AND LLINs or (bednet OR bednets OR insecticide treated net OR insecticide treated nets OR insecticide treated bednet OR insecticide treated bednets OR insecticide treated bed net OR insecticide treated bed nets OR itn s OR itn OR itns OR llin* OR long-lasting insecticidal net OR long lasting insecticidal nets OR long lasting insecticide net OR long lasting insecticide nets OR mesh OR meshes OR net OR nets OR netting) Or Insecticides, Pyrethrins Piperonyl Butoxide )))))) ( Supplementary File searching annex ).

### Study eligibility

This review included randomized control trials or cluster randomized control trials or prospective clinical trials comparing long-lasting insecticidal nets (LLINs) of Pyriproxyfen, Chlorfenapyr, and/or Piperonyl Butoxide for malaria control (test arm) and pyrethroid-only standard LLINs (control arm) for malaria control. The study involved all studies including children aged 6 months to 14 years, permanently residing in a selected household, and an adult caregiver using long-lasting insecticidal nets (LLINs) for malaria control.

### Participants/ peoples

Studies conducted in adults and children who are residents of a region with ongoing malaria transmission and have been provided with long-lasting insecticidal nets (LLINs)-treated net in Africa were eligible for this review.

### Study design

Only cluster randomised and non-randomised cluster-controlled studies that included more than one cluster per arm were considered for this review. Non-randomised controlled study designs were only considered for inclusion when there was a comparison/ control group present. There were no exclusion rules based on any buffer period (i.e., when participants act as their own controls) or length of intervention or timing of measurement of outcomes. All observational studies and modelling studies were excluded.

### Intervention(s)

The interventions of interest are all studies assessed the effectiveness or efficacy of pyriproxyfen, chlorfenapyr, and piperonyl butoxide long-lasting insecticidal nets (LLINs) compared to the standard care (Pyrethroid-only long-lasting insecticidal nets (LLINs) for malaria control in context of Africa

### Comparator(s)/control

The standard care of Pyrethroid-only long-lasting insecticidal nets (LLINs) treated for malaria control in context of Africa

### Main outcome(s)

The selection of outcome measures for this systematic review was based on the outcomes of long-lasting insecticidal nets (LLINs) for malaria control effectiveness and efficacy in terms of reducing and prevented malaria infection prevalence in selected children, anaemia prevalence in children aged 6 months to 4 years, entomological characteristics (mean indoor vectors per household per night, mean entomological inoculation rate per household per night ), malaria case incidence in children (aged 6 months to 10 years, vector density, and sporozoite rate. Which were estimated by the total number of efficacy and effectiveness against malaria, insecticide resistance and malaria prevalence who have pyrethroid-only LLINs used standard treatment divided by a total number of patients multiplied by 100. The additional variable will be the factors associated with efficacy, effectiveness against malaria, and malaria prevalence, which will be reported as risk difference based on the binary outcome from the original studies.

### Operational definition of outcomes

The primary outcome was the prevalence of malaria infection (positive rapid diagnostic test) in children aged 6 months to 14 years at 24 months post-intervention (distribution of LLINs); secondary comparisons of this outcome were made at 12 and 18 months.

Malaria infection incidence–Defined as parasitaemia with or without symptoms, over a population at risk or person-time. Detected through passive or active surveillance.

Secondary outcomes were the incidence of malaria cases (temperature ≥37·5°C or fever within 48 h and positive rapid diagnostic test) in children aged 6 months to 10 years, measured over 24 months of follow-up; Incidence of severe disease–Defined as hospitalisation with parasitaemia, over a population at risk or person-time.

Malaria case incidence rate–Defined as [malaria] symptoms plus [malaria] parasitaemia, over a population at risk or person-time.

The primary malaria transmission outcome was the entomological inoculation rate defined as the number of mosquito vectors tested positive for malaria over 24 months

Prevalence of anaemia–Defined by study thresholds of anaemia.

The prevalence of moderate and severe anaemia (defined as haemoglobin concentration <8 g/dL) in children 6 months to 4 years measured at 12, 18, and 24 months

Studies containing data on entomological outcomes were only included in this review where data for epidemiological outcomes were also reported.

These outcomes were only listed during data extraction and have not formed the basis of any outcome reporting.

- Entomological inoculation rate (EIR)–Defined as the number of infective bites received per person per unit of time.
- Sporozoite rate–Percentage of female Anopheles mosquitoes with sporozoites in the salivary glands.

Sporozoite rate (proportion of vectors infected with malaria parasite),

- Anopheline density–Number of female anopheline mosquitoes in relation to the number of specified shelters or hosts or to a given period sampled, specifying the methods of collection. Vector-density (mean number of malaria vectors collected per house per night)

Other secondary outcomes, including malaria prevalence, anaemia, and entomological inoculation rate at 30 and 36 months post-intervention (13, 16).

### Setting

Studies conducted in countries with ongoing malaria transmission were considered for this review. The presence of other background interventions did not impact on study eligibility if they were present in both arms equally. Studies where additional malaria interventions are considered standard of care were included if interventions (both malaria and non-malaria) were balanced between intervention and control arms. All identified and retrieved studies were from Africa content only.

### Data Extraction

Data were extracted independently by two reviewers using a standardized form with pre- defined criteria as set out by the protocol, and checked and verified by a third reviewer. It included cluster randomized or prospective clinical trials comparing long-lasting insecticidal nets (LLINs) of Pyriproxyfen, Chlorfenapyr, and/or Piperonyl Butoxide for malaria control (test arm) and pyrethroid-only standard LLINs (control arm) for malaria control. The criteria according to which data were extracted comprised: participant demographics (mean age, Household, total number of cluster, total population, number of selected children, malaria infection and anaemia prevalence in selected children); intervention (long-lasting insecticidal nets (LLINs) of Pyriproxyfen, Chlorfenapyr, and/or Piperonyl Butoxide) and comparator details (pyrethroid-only standard LLINs for malaria control intervention); design features (cluster randomized or prospective clinical trials [cRCT] design comparing long-lasting insecticidal nets, study setting, malaria infection and cases incidence, entomological and vector density, and sporozoite rate outcome measures); and outcomes (post-treatment risk ratio (RR) with 95% confidence interval (CI) for malaria controls and other intervention(long-lasting insecticidal nets (LLINs) treatments at the end of treatment and follow-up).

### Data synthesis and statistical analysis

The data was reported in accordance with Preferred Reporting of Systematic Reviews and Meta-Analyses (PRISMA), with the updated guidance of the RISMA 2020 statement (17). We verified the appropriateness of each datum prior to analysis. Pooled estimates were calculated using the STATA Version 17 software (STATA Corporation, College Station, Texas, USA). The extracted data from eligible studies were pooled using the random effects model and expressed as a risk ratio (RR) with a 95% confidence interval (CI). Both random- and fixed- impact methods were used to measure the pooled estimates. The pooled estimates were computed using “metaprop” using a sample size as a weight (wgt) variable with 95% CIs. Pooled estimates were computed using random-effects models and weighted using the inverse variance method in the presence of high heterogeneity among studies.

Heterogeneity was assessed using the *Q*, *I*2 and Tau (τ) statistics. The *I*2 statistic was used to indicate the percentage of overall variability attributable to between-study heterogeneity, categorized into low (25%), moderate (50%), and substantial (75%) groupings according to guidelines reported by Higgins (18). Tau was reported to provide a robust estimate of the variance in true effect sizes (*SD*), which is not susceptible to influences from number and precision of included studies (as can be the case for Q and *I*2). Any p-value <0.05 was considered as statistically significant. Sensitivity analysis, subgroup analysis, and publication bias was also assessed as appropriate. Subgroup analyses were performed using different parameters (country, AOR, Intervention types, duration of follow up,). Forest plots, summary tables, and text are used to present the findings of this study.

### Quality assessment

The quality of the study was assessed using the revised Joanna Briggs Institute (JBI) critical appraisal tool for the assessment of risk of bias for randomized controlled trials (19) and the results were graded as low, medium, or high if the quality score was < 60%, 60–80%, or > 80%, respectively. We inspected the funnel plot and conducted Egger’s regression tests to assess publication bias (20) (Supplementary file JBI).

### Publication bias

The likelihood of publication bias was evaluated using four methods to prevent overreliance on one approach. These comprised the following: (1) Funnel plots of standardized mean differences plotted against standard errors which were visually observed to detect possible asymmetry; (2) (21) Trim and Fill imputation was used to predict the adjusted combined effect size (ES) taking account of publication bias; (3) Egger’s regression was utilized as a formal statistical assessment of potential publication bias by regressing standardized effect estimates onto a measure of precision(22, 23); and (4) Rosenthal fail-safe *N* calculation which estimates the number of additional studies with an ES of zero required to turn the overall effect insignificant (24). In addition, Egger’s weighted regression and Begg’s tests will be used to check publication bias. The statistical significance of publication bias was declared at a P-value of less than 0.05.

### GRADE Analysis

In the meta-analysis, the quality of the evidence was assessed for each comparison using the Grading of Recommendations Assessment, Development, and Evaluation (GRADE) tool (25).

Meta-analytic comparisons were graded by three of the authors to reach a consensus quality rating (high, moderate, low, or very low quality) based on five domains; (1) risk of bias in the individual included studies, (2) inconsistency, (3) indirectness of treatment estimate effects, (4) imprecision, and (5) publication bias ( Evidence Grading Supplementary).

## Results

### Selection of studies

In the initial search, 952 studies were obtained from databases and supplementary grey literature sources. First, 409 studies were excluded because of duplication. Then, of the 543 studies, 543 were screened using titles and abstracts, and 487 were removed. Finally, the full texts of 56 studies were assessed for eligibility (2, 6, 8–12, 16, 26–73). Of the 56 studies, 39 were excluded due to inconsistent results, and 6 were protocol RCTs. Finally, 11 eligible studies were used in the final analysis of the current systematic review and meta-analysis (2, 6, 8, 10–12, 16, 27–30) see details in figure 1.

**Figure 1:**
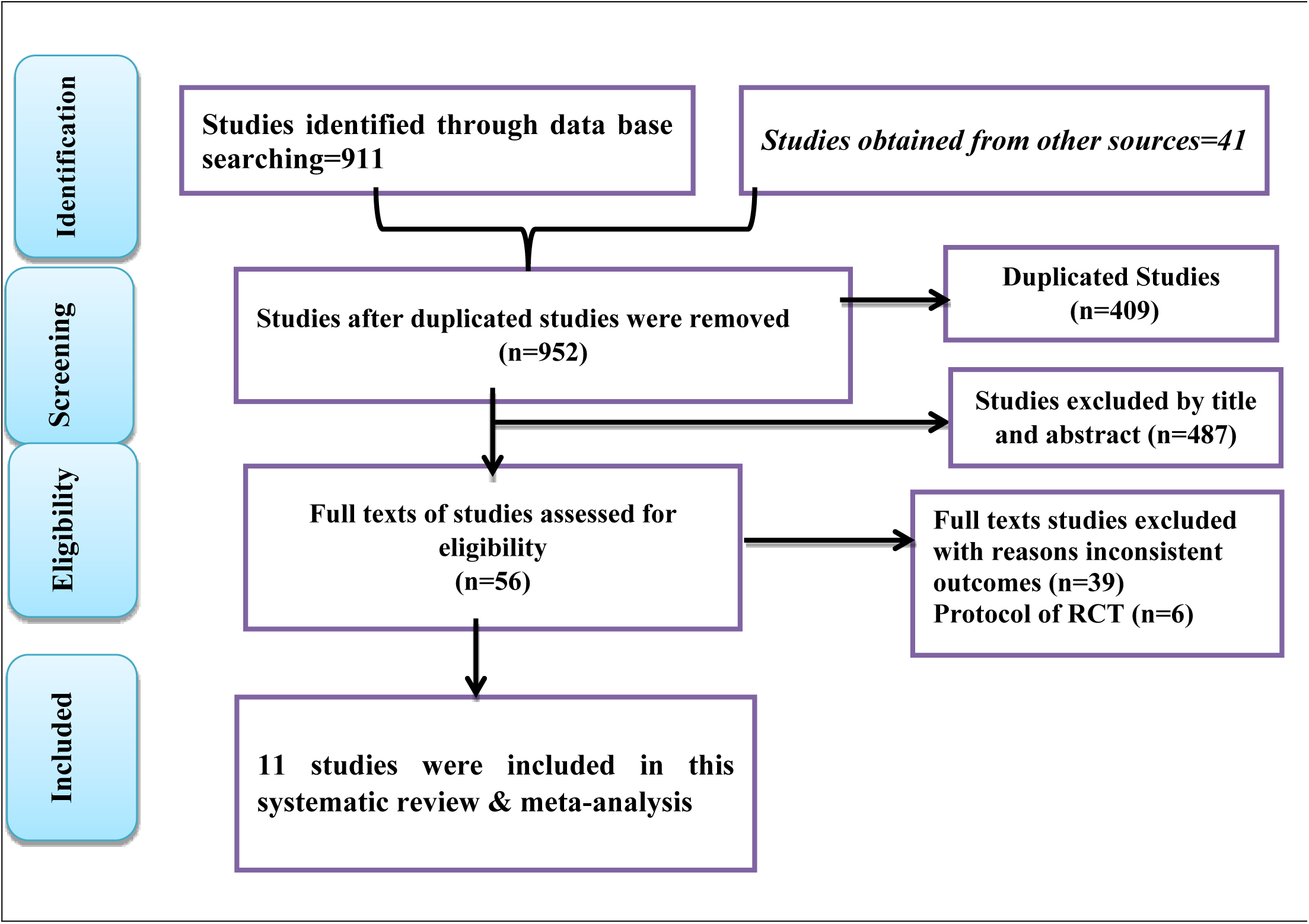
PRISMA flow chart showing the study selection process

### Study Characteristics

The study included 11 studies, involving data from over 21,916 households, 1,145,035 people in core and buffer areas, and 34,327 children across all of the studies reporting sample size were included in the final analysis, and all identified and retrieved studies were from Africa content only. Of these studies the majority six from Tanzania (6, 8, 12, 16, 28, 29), and the rest were two equally from Benin (10, 11), and Uganda (2, 30), and one from Kenya (27) respectively.

In summary, these studies were conducted in 4 African countries, and all studies were cluster randomized control trials or randomized control trials. Critical appraisal of randomized control trials revealed that 100% of the studies scored high quality, and Cochrane methodology was used to assess the risk of bias and evaluate evidence quality, which was graded as high. This research provides a very good indication of the likely effect. The likelihood that the effect will be substantially different is low **(Evidence Grading Supplementary).**

### Pooled baseline Malaria infection prevalence in selected children using different long-lasting insecticidal nets (LLINs) as malaria control in Africa

**Pooled effectiveness and efficacy of Pyriproxyfen long-lasting insecticidal nets (LLINs) versus pyrethroid-only LLINs malaria infection reduction**

Forest plots showed that malaria infection risk reduction among children using the Pyriproxyfen intervention group versus pyrethroid-only long-lasting insecticidal nets (LLINs) standard treatment revealed no significant difference (RR = -0.00 with 95%CI -0.04, 0.03). Figure 1 reveals that there is no significant difference in malaria prevalence reduction between treatment and control groups. Furthermore, the included studies were homogeneous, with an I2 of 0.00. See details in Figure 2.

**Figure 2:**
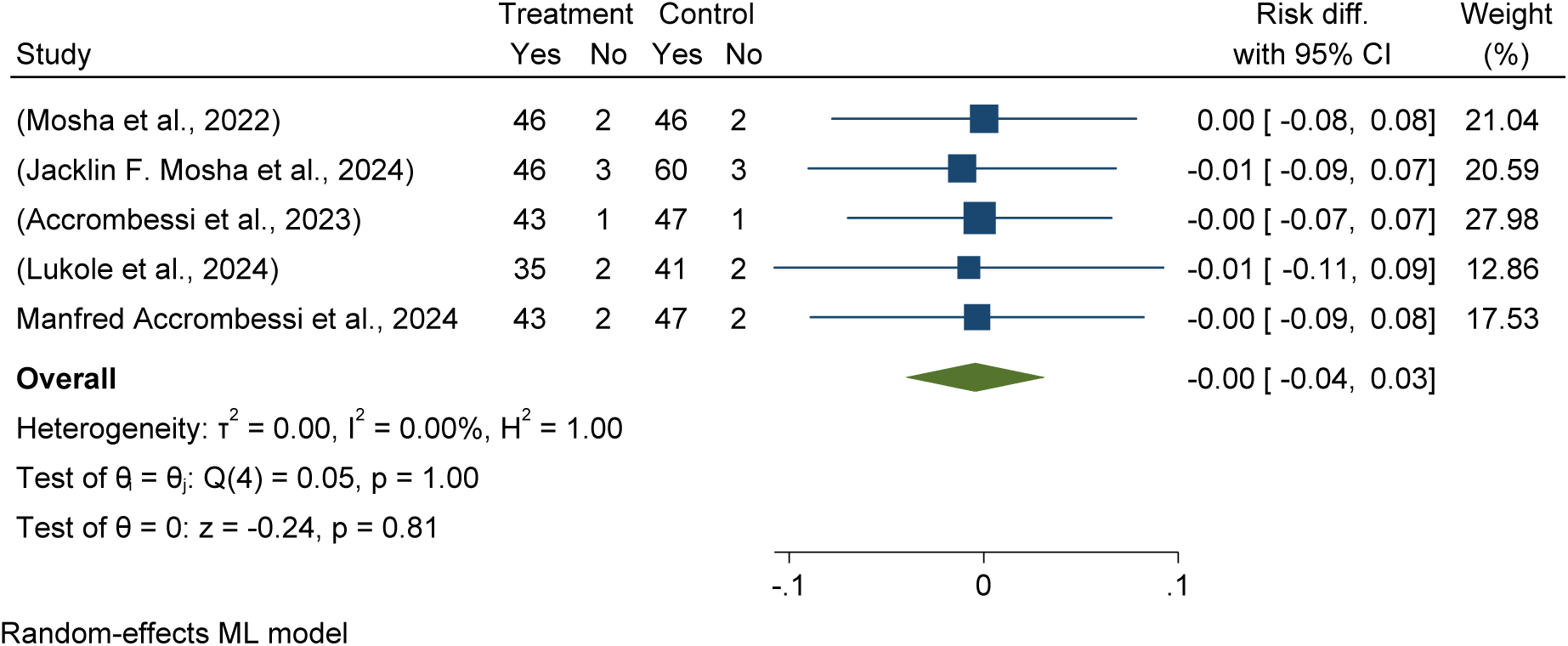
Illustrates the effectiveness and efficacy of pyriproxyfen long-lasting insecticidal nets (LLINs) in malaria infection risk reduction among children compared to pyrethroid-only LLINs for malaria control in Africa 2024.

### Pooled prevalence of malaria infection among children using pyriproxyfen long-lasting insecticidal nets (LLINs) versus pyrethroid-only LLINs

This randomized control trial meta-analysis found the estimated pooled prevalence of malaria infection among children using pyriproxyfen long-lasting insecticidal nets (LLINs) treatment groups were 42.8 per 100 children with (95% CI: 39.47–46.12%) and 47.80% (37.77%, 57.84%) in the pyrethroid-only LLINs control group (see details in Figure 3 and Figure 8).

**Figure 3:**
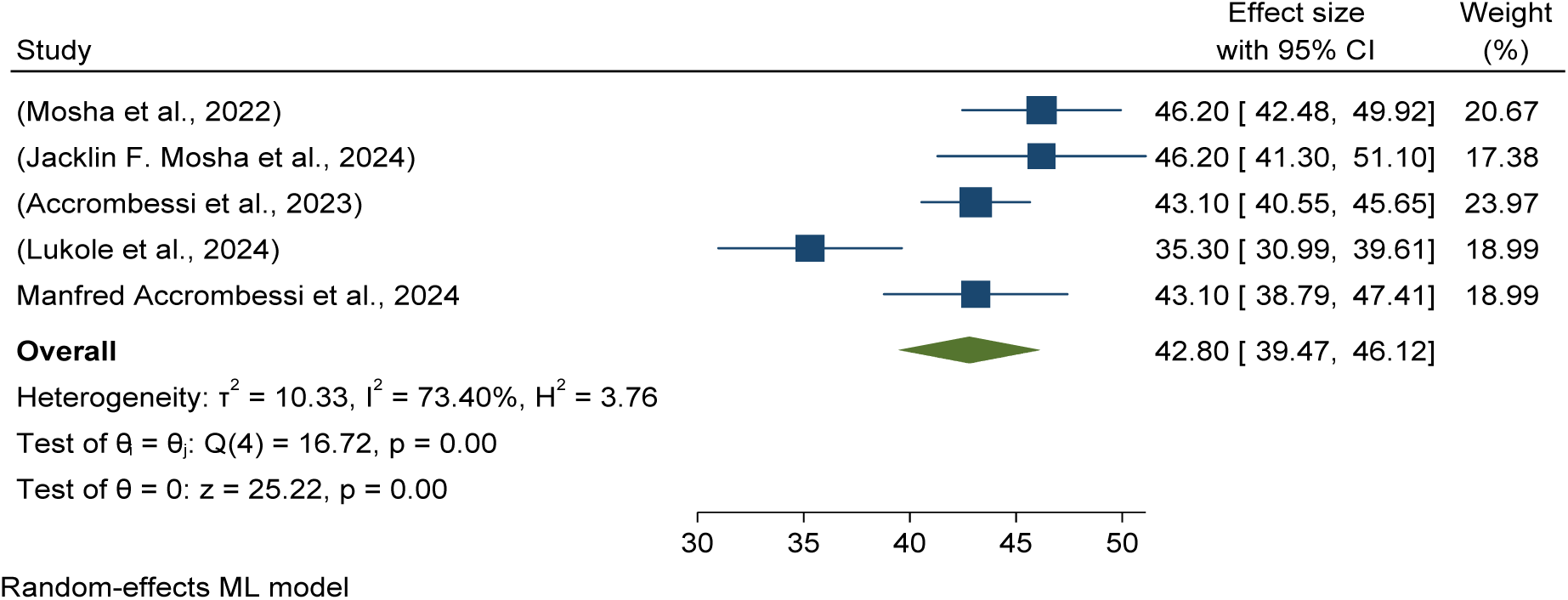
Forest plots showed pooled prevalence of malaria among children using pyriproxyfen long-lasting insecticidal nets (LLINs) for malaria control in Africa in 2024.

### Pooled effectiveness and efficacy of chlorfenapyr long-lasting insecticidal nets (LLINs) versus pyrethroid-only LLINs malaria infection reduction

Forest plots showed that malaria infection risk reduction among children using chlorfenapyr long-lasting insecticidal nets (LLINs) treatment groups reduced the risk of malaria infection by 1% compared to standard or pyrethroid-only LLINs (RR = -0.01 with a 95%CI of -0.04–0.03) see details in Figure 4.

**Figure 4:**
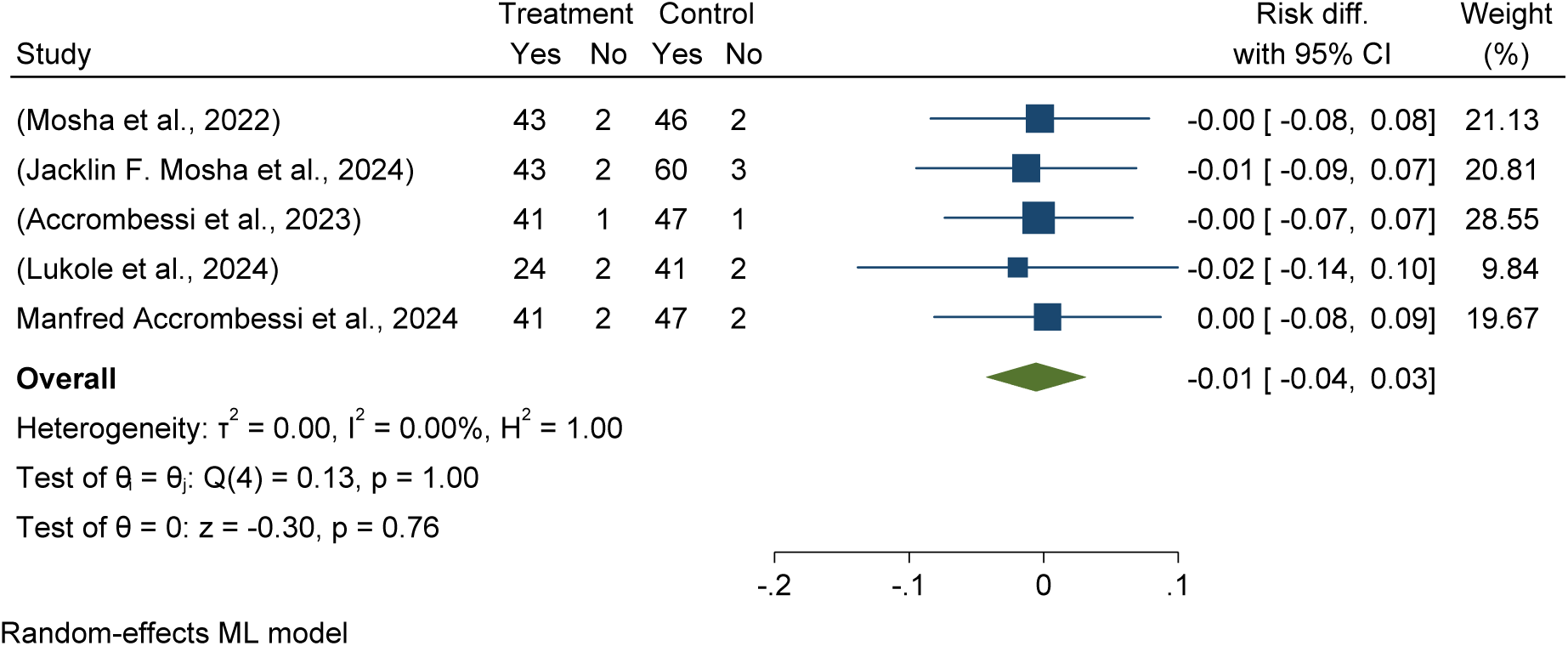
illustrates the effectiveness and efficacy of chlorfenapyr long-lasting insecticidal nets (LLINs) in malaria infection risk reduction among children compared to pyrethroid-only LLINs for malaria control in Africa 2024.

### Pooled prevalence of malaria infection among children using chlorfenapyr long lasting insecticidal nets (LLINs) versus pyrethroid-only LLINs

This meta-analysis of randomized control trials found that the pooled prevalence of malaria infection among children using chlorfenapyr long-lasting insecticidal nets was 38.09 per 100 with (95% CI: 39.47–46.12%), while pyrethroid-only LLINs had a nearly 50% (47.80 with 95% CI 37.77%, 57.84%) pooled prevalence malaria infection among children using pyrethroid-only long-lasting insecticidal nets (LLINs) (see details in Figure 5 and Figure 8).

**Figure 5:**
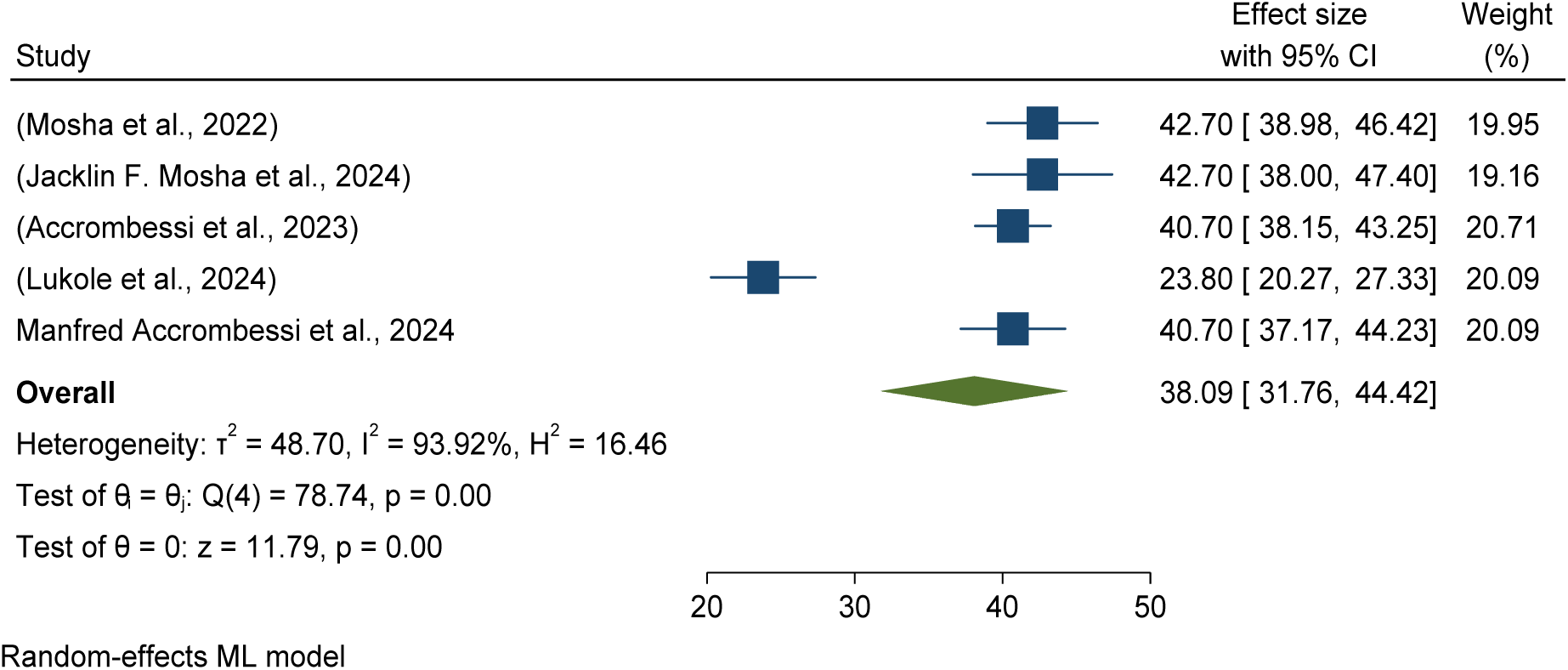
Forest plots showed pooled prevalence of malaria infection among children using chlorfenapyr long-lasting insecticidal nets (LLINs) for malaria control in Africa 2024.

### Pooled effectiveness and efficacy of Piperonyl butoxide long-lasting insecticidal nets (LLINs) versus pyrethroid-only LLINs malaria infection reduction

Forest plots showed that malaria infection risk reduction among children using the Piperonyl butoxide treatment group versus pyrethroid-only long-lasting insecticidal nets (LLINs) standard treatment revealed no significant difference (RR = -0.00 with 95%CI -0.03, 0.02).

The study shows no significant difference in malaria prevalence reduction between treatment and control groups, and the included studies were homogeneous with an I2 of 0.00. See details in Figure 6.

**Figure 6:**
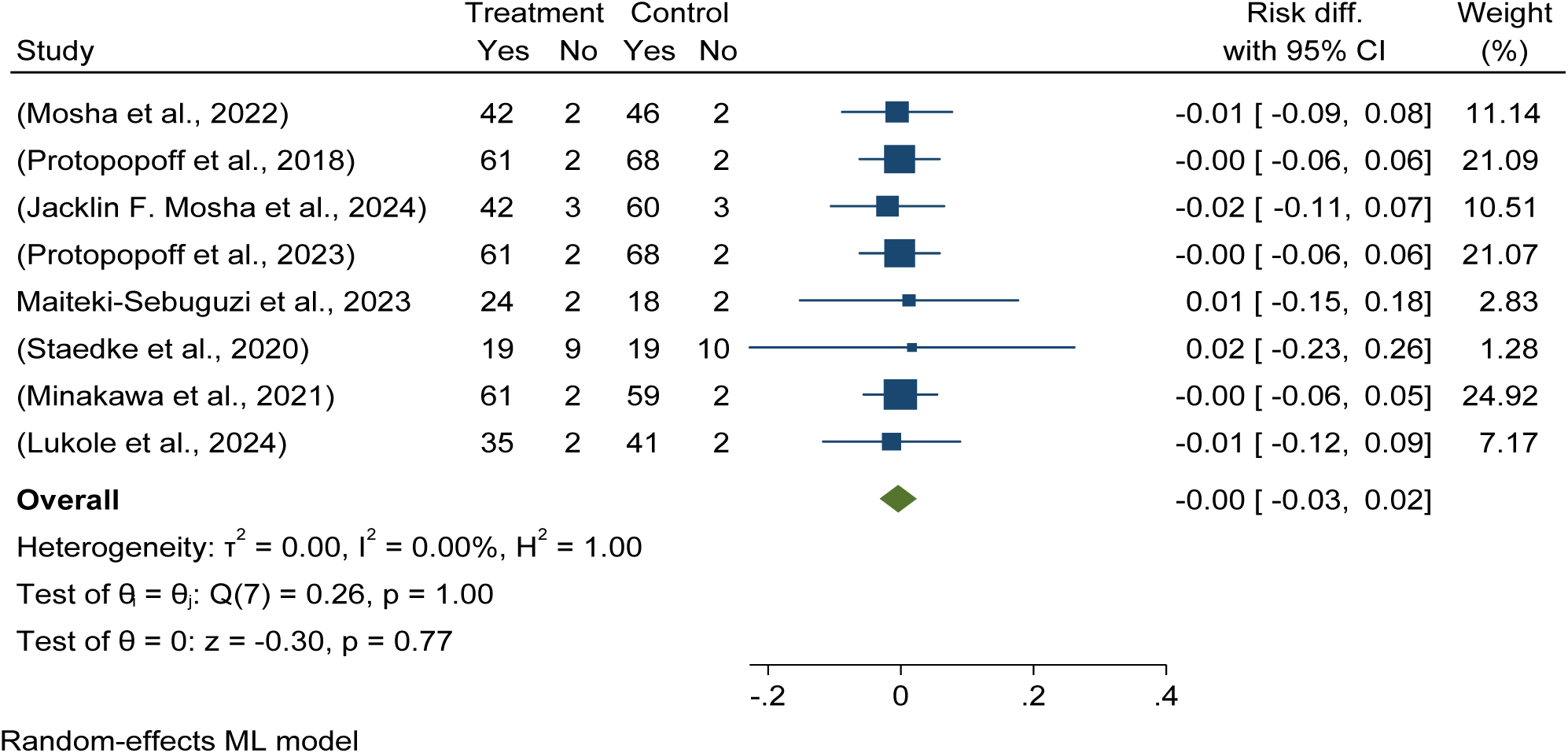
Forest plot illustrates the effectiveness and efficacy of Piperonyl butoxide long-lasting insecticidal nets (LLINs) in malaria infection risk reduction among children compared to pyrethroid-only LLINs for malaria control in Africa 2024

### Pooled prevalence of malaria infection among children using Piperonyl butoxide long lasting insecticidal nets (LLINs) versus pyrethroid-only LLINs

This randomized control trial meta-analysis determined the estimated pooled prevalence of malaria infection among children using Piperonyl butoxide long-lasting insecticidal nets (LLINs) were 43.91 per 100 children with (95% CI: 33.43%, 54.39%), while pyrethroid-only

LLINs had a nearly 50% (47.80 with 95% CI 37.77%, 57.84%) pooled prevalence malaria infection among children using pyrethroid-only long-lasting insecticidal nets (LLINs) (see details in Figure 7 and Figure 8).

**Figure 7:**
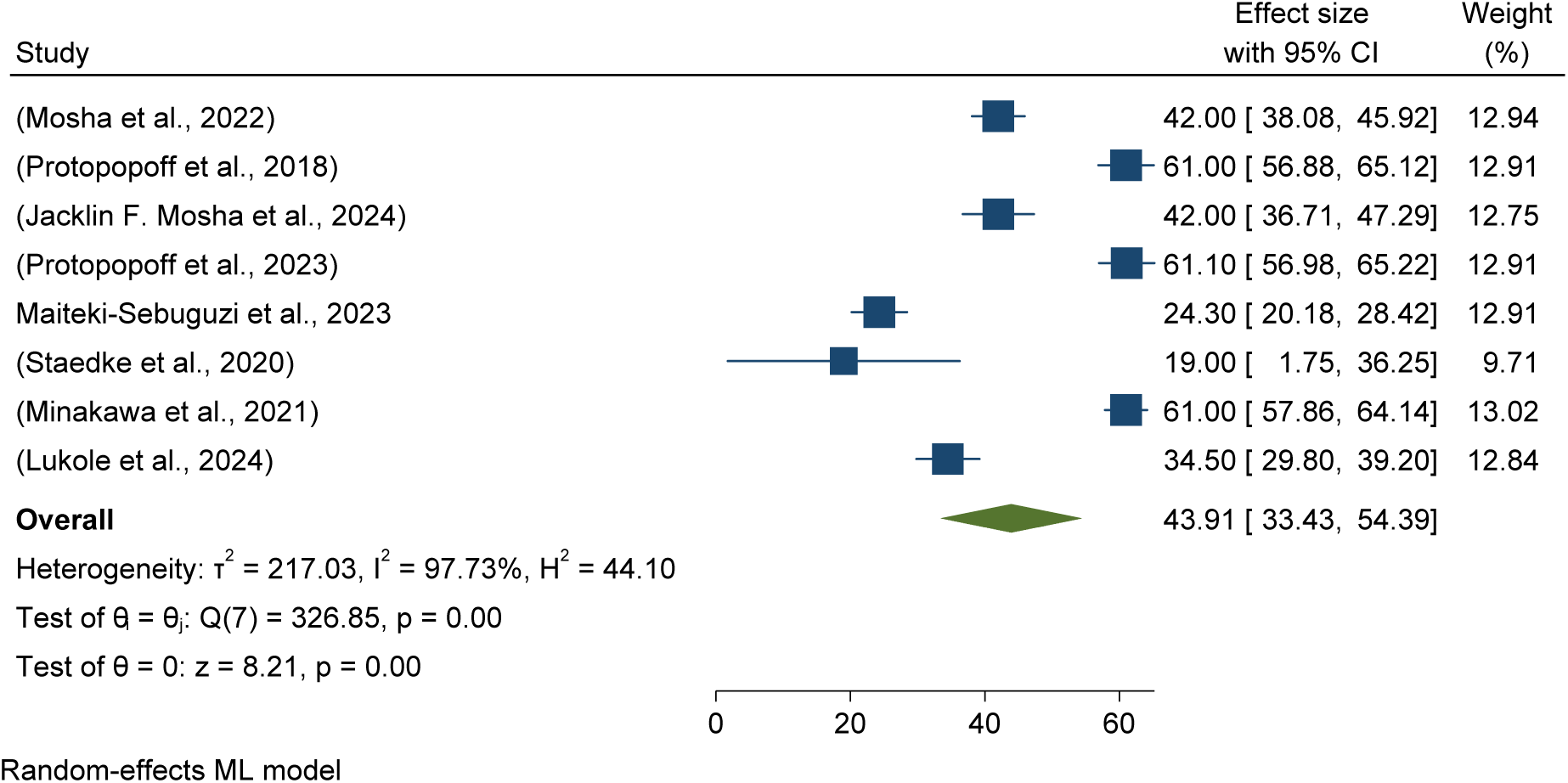
Forest plots showed pooled prevalence of malaria infection among children using c Piperonyl butoxide long-lasting insecticidal nets (LLINs) for malaria control in Africa 2024.

**Figure 7:**
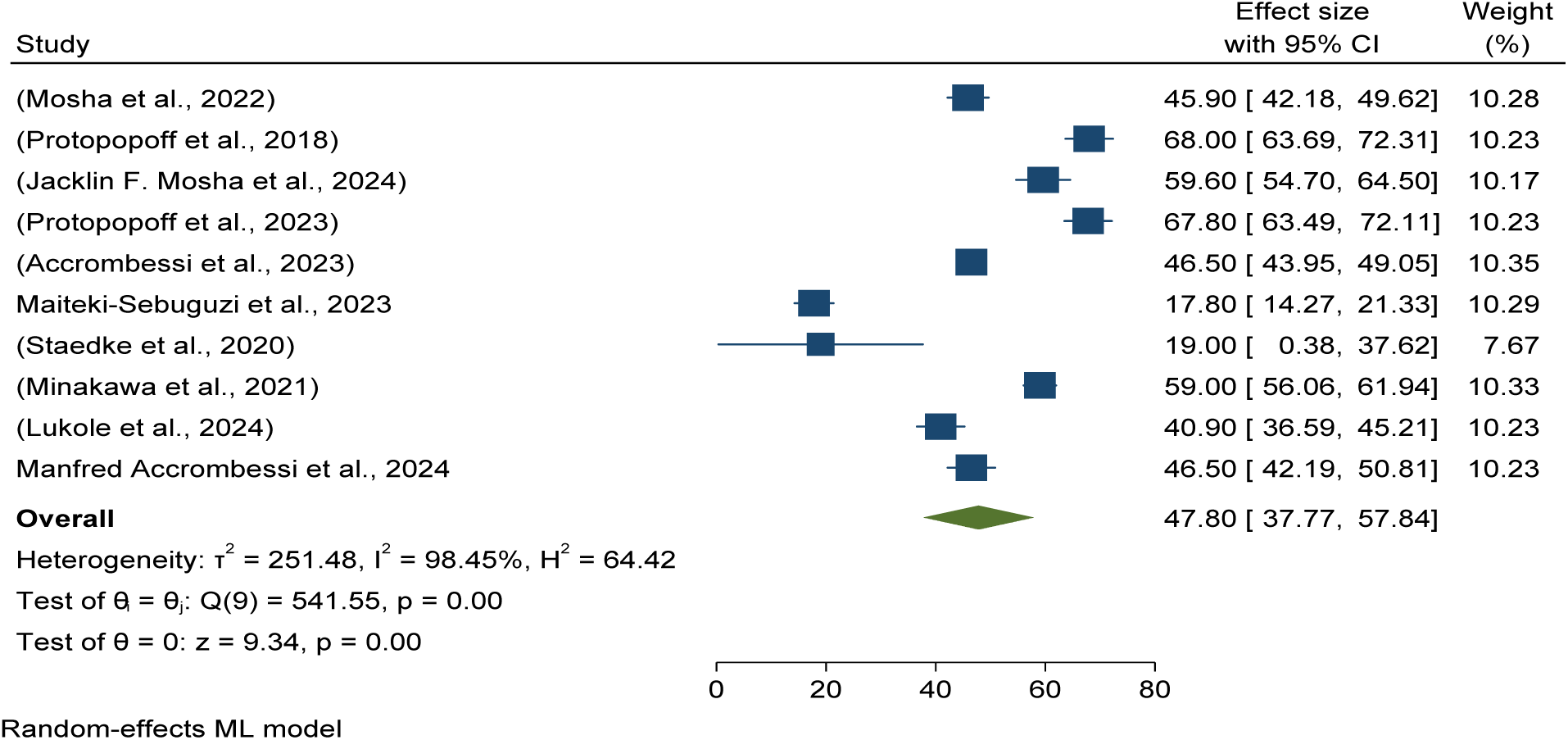
Forest plots showed pooled prevalence of malaria infection among children using pyrethroid-only long-lasting insecticidal nets (LLINs) for malaria control in Africa 2024

**Figure 8:**
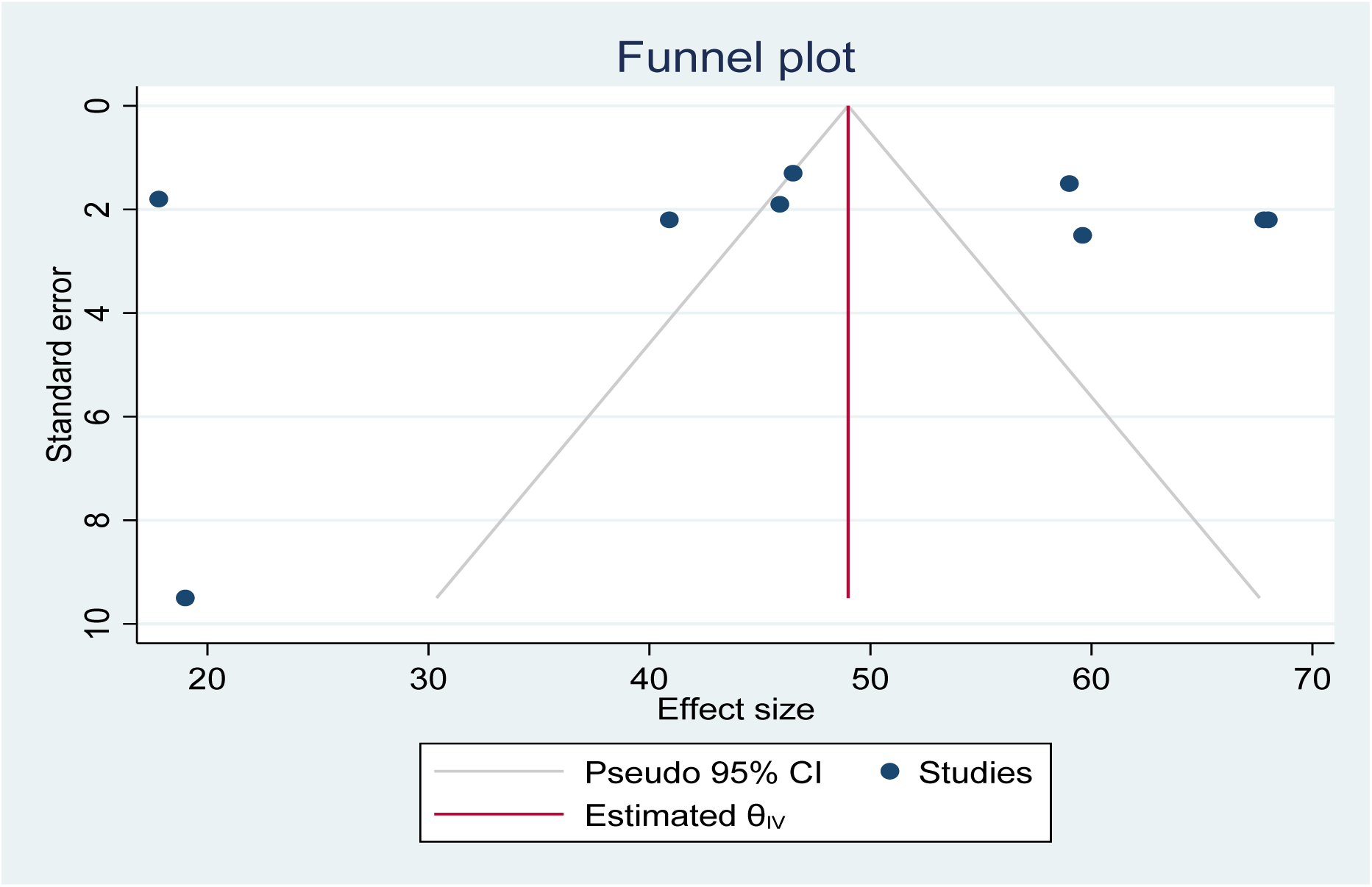
Funnel plot showing the distribution of included studies pooled malaria infection risk reduction among children using pyrethroid-only long-lasting insecticidal nets (LLINs) for malaria control in Africa in 2024.

### Pooled prevalence of malaria infection among children using Pyrethroid-only long lasting insecticidal nets (LLINs)

This randomized control trial meta-analysis determined the estimated pooled prevalence of malaria infection among children using pyrethroid-only long-lasting insecticidal nets (LLINs) control group were 47.80 per 100 children with (95% CI: 37.77%, 57.84%) in Africa (See details Figure 8).

This funnel plot indicates as there is no publication bias among included studies as it is symmetrical distributed. Hence, the funnel plot seemed symmetric and the Egger’s regression test (PV= 0.1460), which confirmed no publication bias among included studies (Figure 8).

### Pooled baseline Anaemia prevalence in children aged 6 months to 4 years using different long-lasting insecticidal nets (LLINs) as malaria control in Africa

**Pooled effectiveness and efficacy of Pyriproxyfen long-lasting insecticidal nets (LLINs) versus pyrethroid-only LLINs for anaemia reduction in children**

Forest plots showed that no difference in anaemia risk reduction among children aged 6 months to 4 years using pyriproxyfen long-lasting insecticidal nets versus pyrethroid-only LLINs standard treatment, with the included studies were homogeneous(I2 = 0.01%). See details in Figure 9.

**Figure 9:**
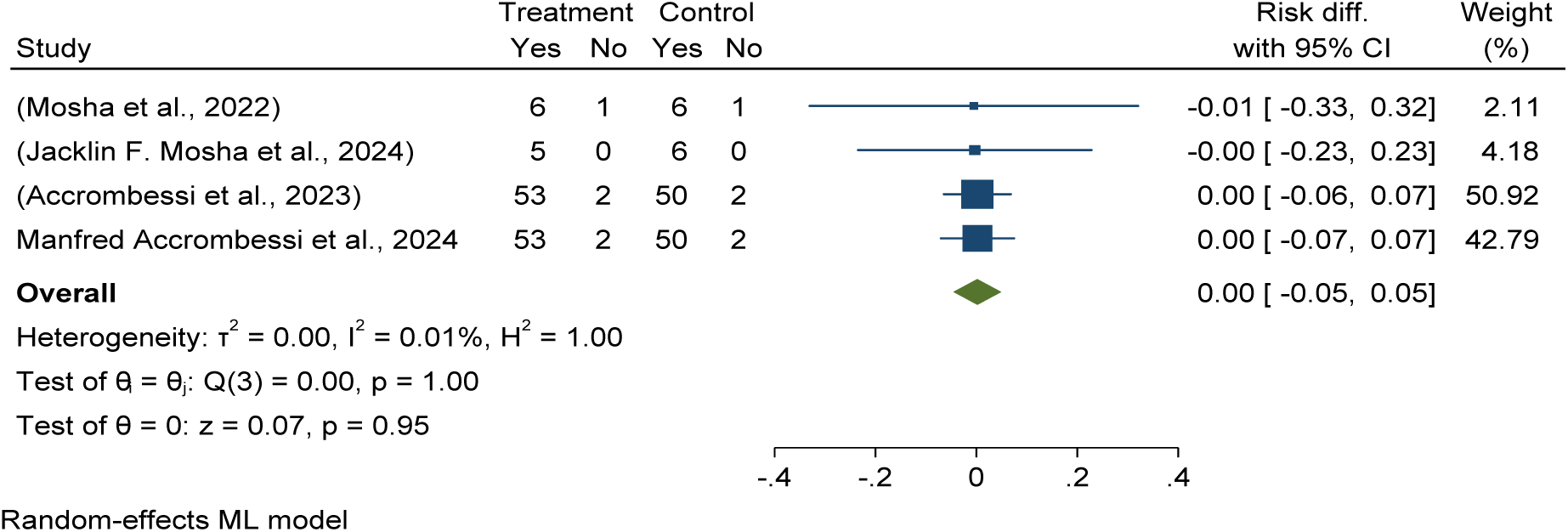
illustrates the effectiveness and efficacy of pyriproxyfen long-lasting insecticidal nets (LLINs) with no difference in anaemia risk reduction among children aged 6 months to 4 years compared to pyrethroid-only LLINs in Africa in 2024.

### Pooled baseline anaemia prevalence in children aged 6 months to 4 years using pyriproxyfen long-lasting insecticidal nets (LLINs) versus pyrethroid-only LLINs

This randomized control trial meta-analysis found the estimated pooled prevalence of Anaemia among children aged 6 months to 4 years using pyriproxyfen long-lasting insecticidal nets (LLINs) treatment groups were 29.28 per 100 children with (95% CI: 5.81–52.75%) and 25.18% (12.78%, 37.58%) in the pyrethroid-only LLINs control group (see details in Figure 10 and Figure 15).

**Figure 10:**
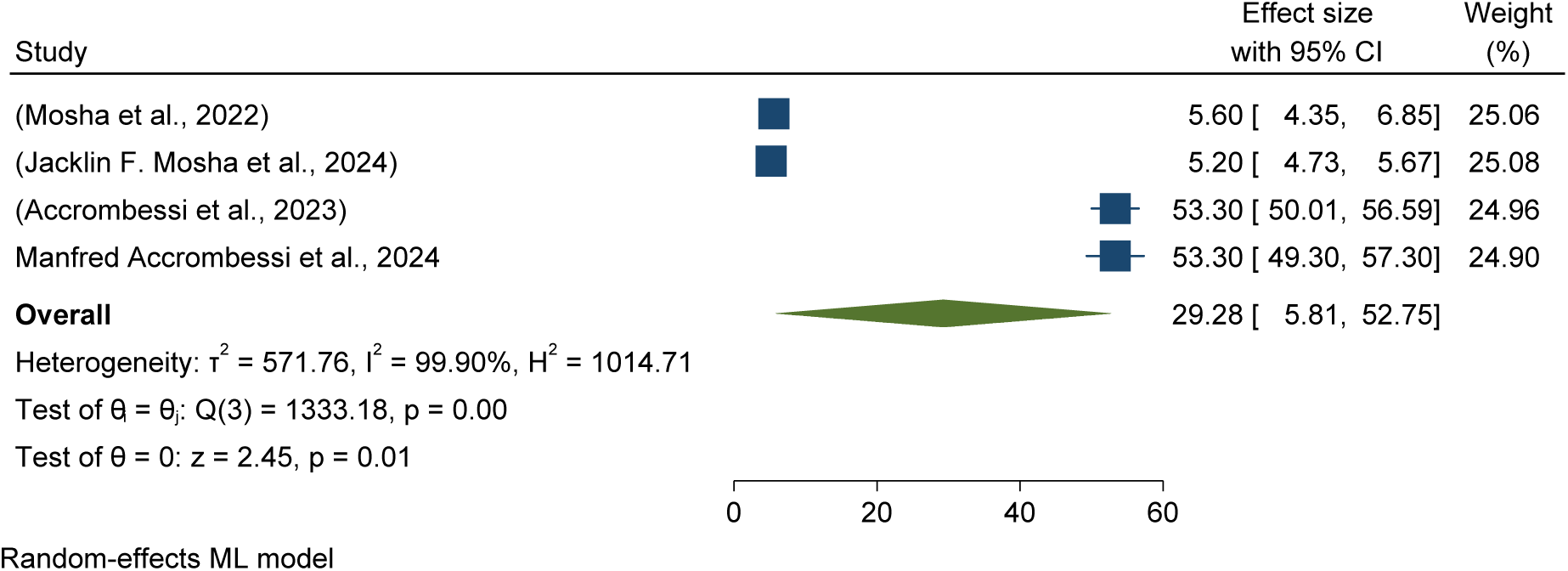
Forest plots showed pooled prevalence of anaemia among children aged 6 months to 4 years using pyriproxyfen long-lasting insecticidal nets (LLINs) for malaria control in Africa in 2024.

### Pooled effectiveness and efficacy of chlorfenapyr long-lasting insecticidal nets (LLINs) versus pyrethroid-only LLINs for anaemia reduction in children

Forest plots showed that anaemia risk reduction among children aged 6 months to 4 years using chlorfenapyr long-lasting insecticidal nets (LLINs) treatment groups was reduced by 1% compared to standard or pyrethroid-only LLINs (RR = -0.01 with a 95%CI of -0.05–0.03).see details in Figure 11.

**Figure 11:**
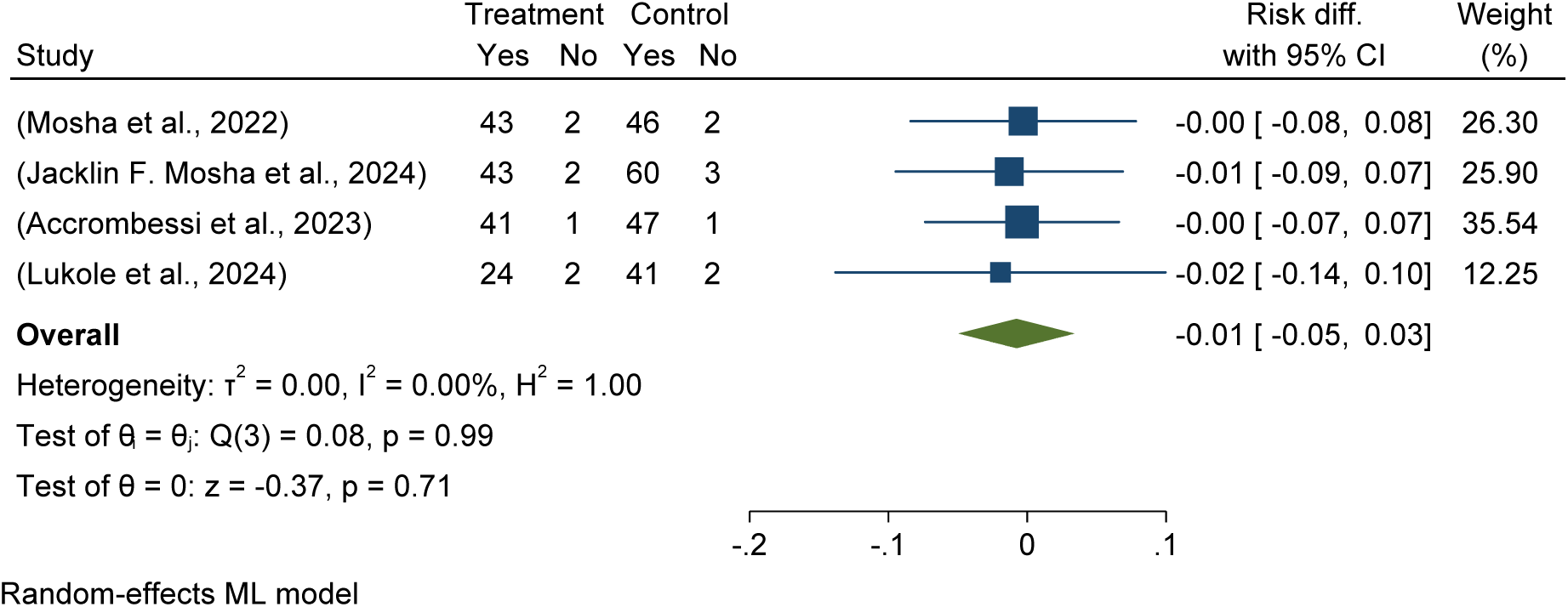
Forest plot shows the effectiveness and efficacy of chlorfenapyr long-lasting insecticidal nets (LLINs) in anaemia risk reduction among children aged 6 months to 4 years compared to pyrethroid-only LLINs in Africa in 2024.

### Pooled prevalence of anaemia among children using chlorfenapyr long lasting insecticidal nets (LLINs) versus pyrethroid-only LLINs

The study found that the pooled prevalence of anaemia among children aged 6 months to 4 years using chlorfenapyr long-lasting insecticidal nets was 29.36 per 100 children, compared to 25.18% in the control/pyrethroid-only group (see details in Figures 12 and 15).

**Figure 12:**
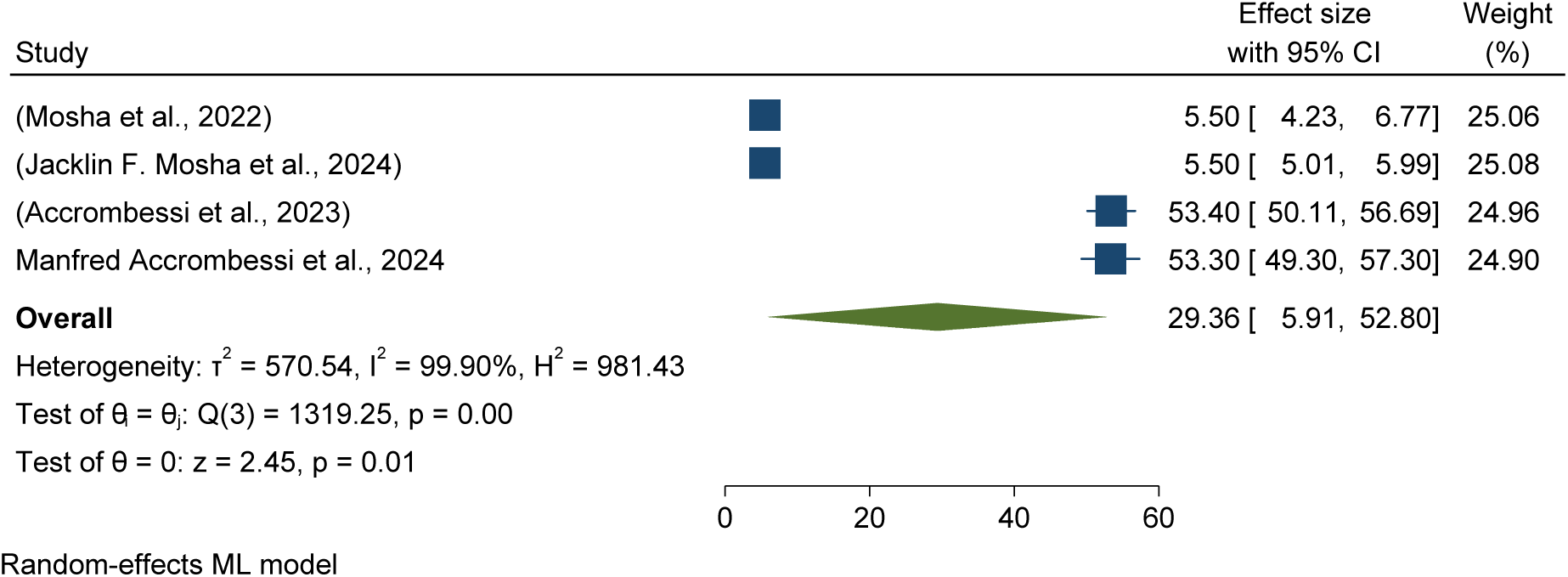
Forest plots showed a pooled prevalence of anaemia among children using chlorfenapyr long-lasting insecticidal nets (LLINs) for malaria control in Africa in 2024.

### Pooled effectiveness and efficacy of Piperonyl butoxide long-lasting insecticidal nets (LLINs) versus pyrethroid-only LLINs for anaemia reduction in children

Forest plots showed that anaemia risk reduction among children aged 6 months to 4 years using Piperonyl butoxide long-lasting insecticidal nets (LLINs) treatment groups was reduced by 2% compared to standard or pyrethroid-only LLINs (RR = -0.02 with a 95%CI of -0.07, 0.04).see details in Figure 13.

**Figure 13:**
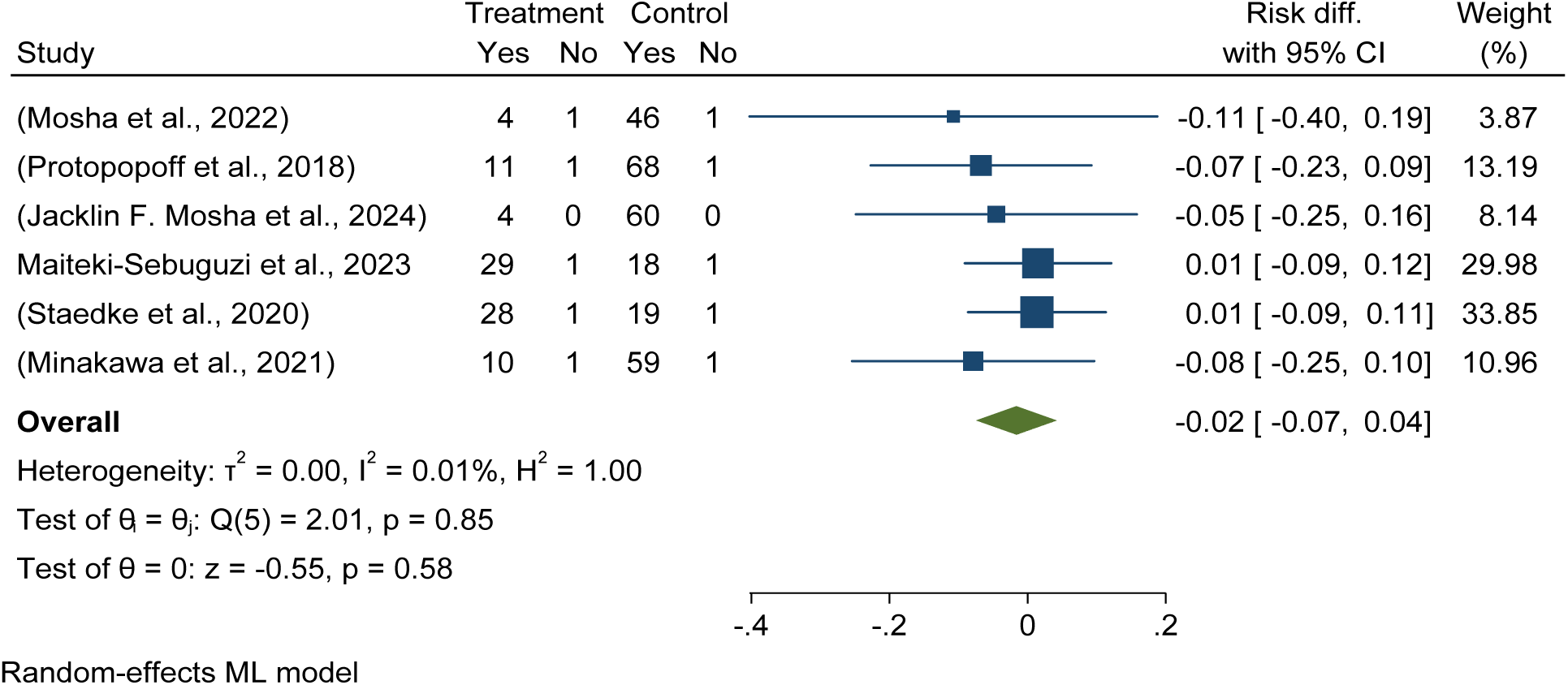
Forest lot shows the effectiveness and efficacy of Piperonyl butoxide long-lasting insecticidal nets (LLINs) in anaemia risk reduction among children aged 6 months to 4 years compared to pyrethroid-only LLINs in Africa in 2024.

### Pooled prevalence of anaemia among children using piperonyl butoxide long lasting insecticidal nets (LLINs) versus pyrethroid-only LLINs

The study found that the pooled prevalence of anaemia among children aged 6 months to 4 years using piperonyl butoxide long-lasting insecticidal nets was 14.31 per 100 children, compared to 25.18% in the control/pyrethroid-only group (see details in Figures 14 and 15).

**Figure 14:**
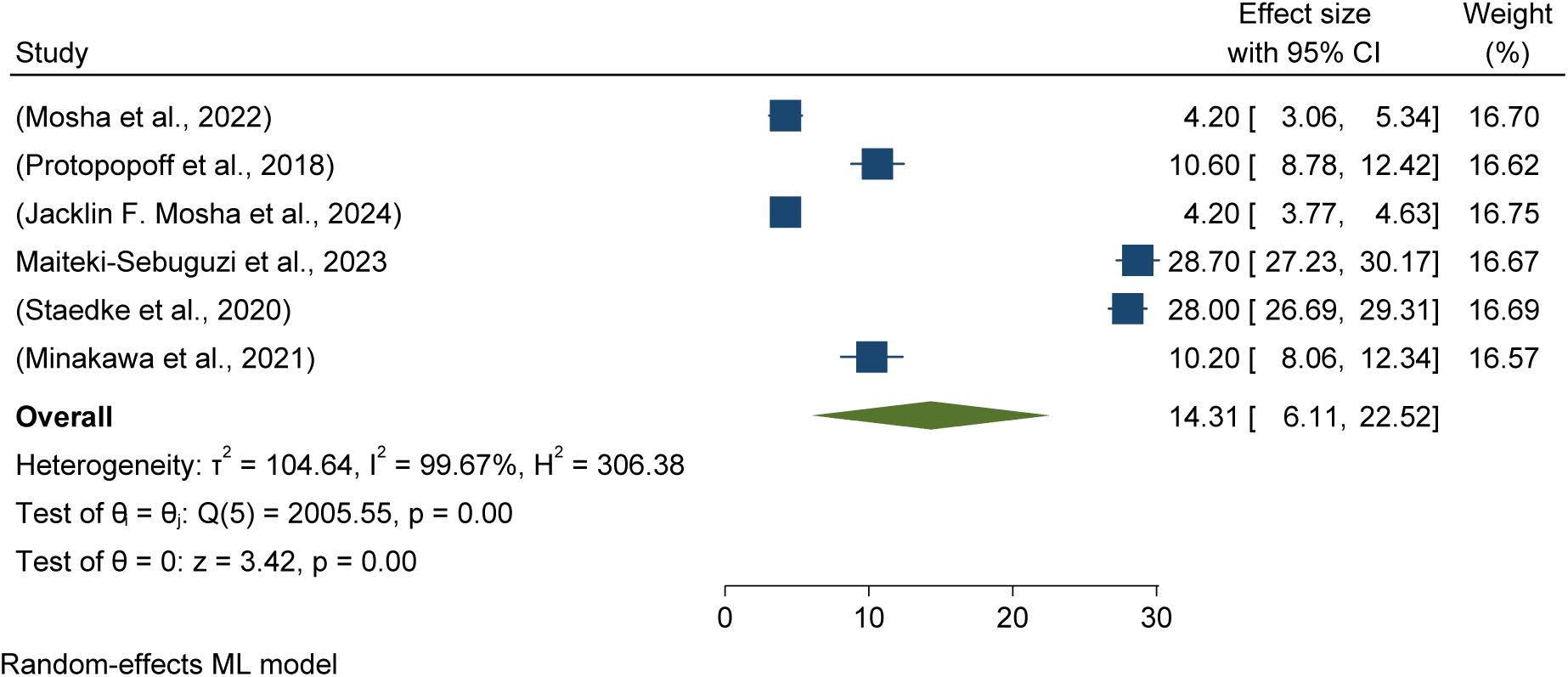
Forest plots showed a pooled prevalence of anaemia among children using piperonyl butoxide long-lasting insecticidal nets (LLINs) for malaria control in Africa in 2024.

**Figure 15:**
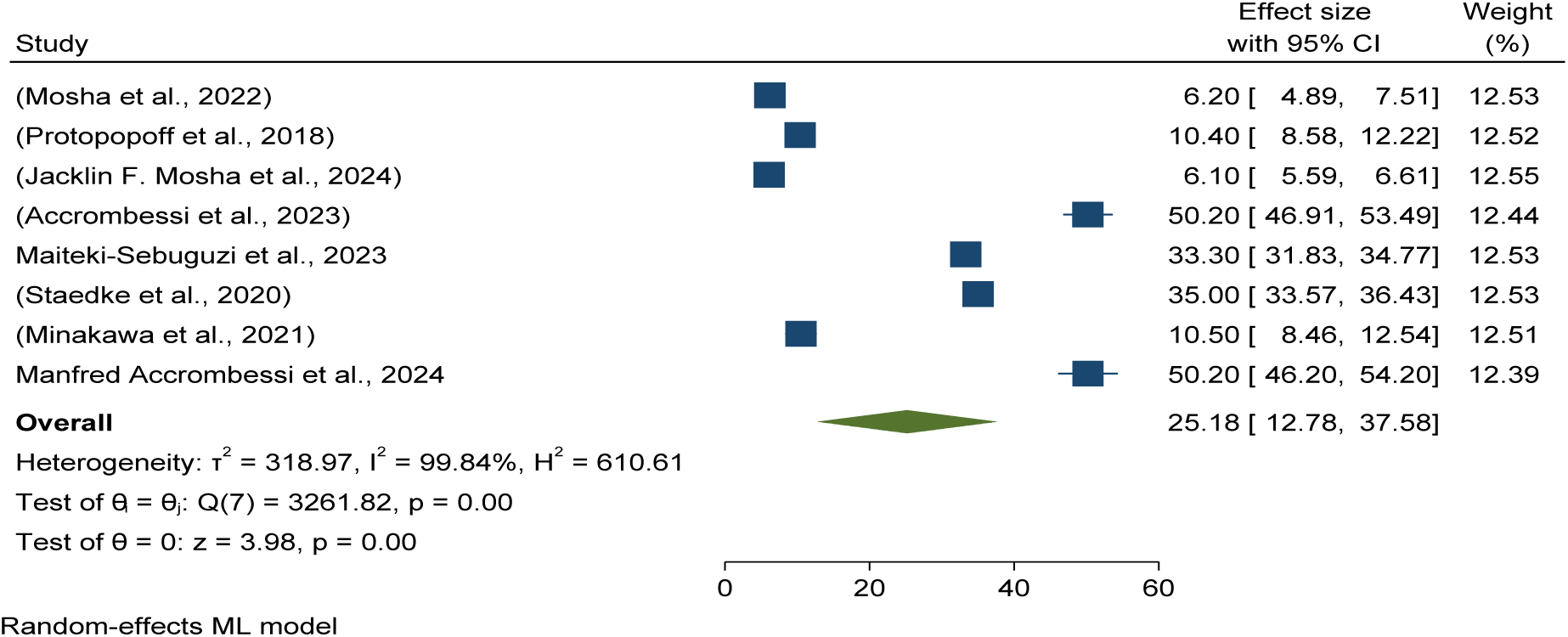
Forest plots showed a pooled prevalence of anaemia among children using pyrethroid-only long lasting insecticidal nets (LLINs) for malaria control in Africa in 2024.

### Pooled prevalence of anaemia among children using Pyrethroid-only long lasting insecticidal nets (LLINs)

The study found that the pooled prevalence of anaemia among children aged 6 months to 4 years using pyrethroid-only long lasting insecticidal nets (LLINs) nets was 25.18 per 100 children with (95% CI: 12.78%, 37.58%) in Africa 2024 (See details in Figure 15).

### Baseline pooled mean indoor vectors density per household per night

This meta-analysis determined entomological outcomes effectiveness and efficacy in terms of mean indoor vector density per household per night reduction by using different long-lasting insecticidal nets (LLINs) as malaria control in Africa.

Forest plots showed that no difference in mean indoor vector density per household per night reduction using pyriproxyfen long-lasting insecticidal nets versus pyrethroid-only LLINs standard treatment, with the included studies were homogeneous (I2 = 0.00%). This indicates no additional effectiveness or efficacy in the mean indoor vector density in the study area. See details in Figure 16.

**Figure 16:**
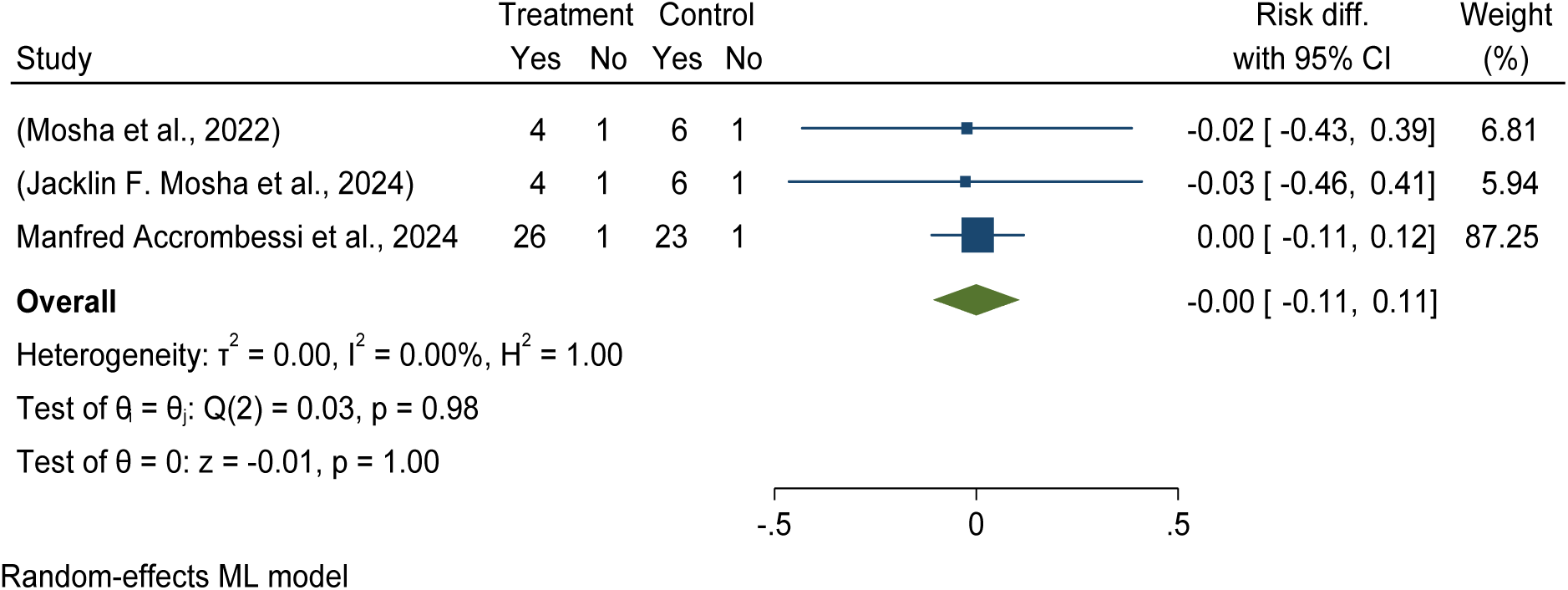
Forest plot shows the effectiveness and efficacy of pyriproxyfen long-lasting insecticidal nets (LLINs) with no difference in reducing mean indoor vector density per household per night compared to pyrethroid-only LLINs in Africa in 2024.

### Pooled prevalence of mean indoor vectors density per household per night using pyriproxyfen long lasting insecticidal nets (LLINs)

This meta-analysis found that pyriproxyfen long-lasting insecticidal nets (LLINs) did not significantly reduce indoor vector density per household per night compared to the standard/control group. The pooled prevalence of mean indoor vector density was 11.28 per household per night in pyriproxyfen LLINs, compared to 12.78 per household per night in pyrethroid-only LLINs (see details in Figures 17 and 22).

**Figure 17:**
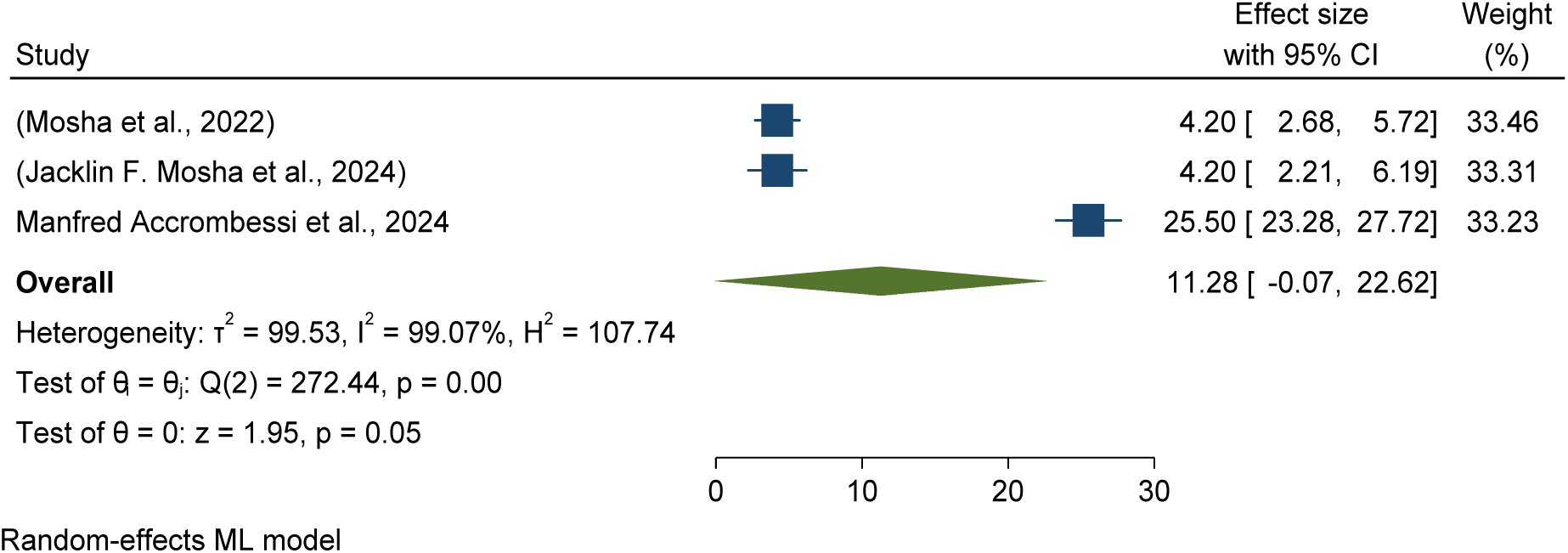
Forest plots showed baseline pooled mean indoor vectors density per household per night using pyriproxyfen long lasting insecticidal nets (LLINs) for malaria control in Africa in 2024.

### Pooled prevalence of mean indoor vectors density per household per night using chlorfenapyr long lasting insecticidal nets (LLINs)

Figure 18 Forest plot shows the effectiveness and efficacy of chlorfenapyr long-lasting insecticidal nets (LLINs) intervention decreases the risk of mean indoor vector density per household per night by 2% when controlled with the standard care/pyrethroid-only LLINs in Africa in 2024. Therefore, it effective intervention which have potential bring down the risk of malaria by 2%.

**Figure 18:**
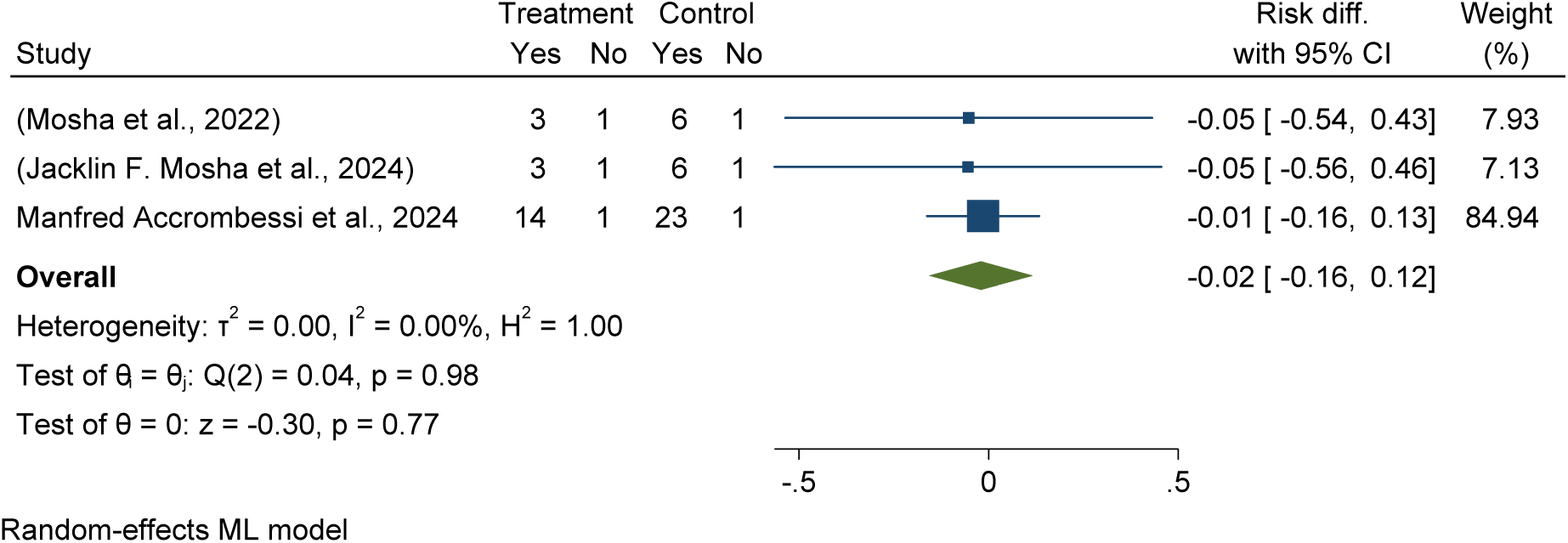
Forest plot shows the effectiveness and efficacy of *chlorfenapyr* long-lasting insecticidal nets (LLINs) with no difference in reducing mean indoor vector density per household per night compared to pyrethroid-only LLINs in Africa in 2024

### Pooled prevalence of mean indoor vectors density per household per night using chlorfenapyr long lasting insecticidal nets (LLINs)

The pooled prevalence of mean indoor vector density was 6.51per household per night in chlorfenapyr LLINs, compared to 12.78 per household per night in pyrethroid-only LLINs (see details in Figures 19 and 22).

**Figure 19:**
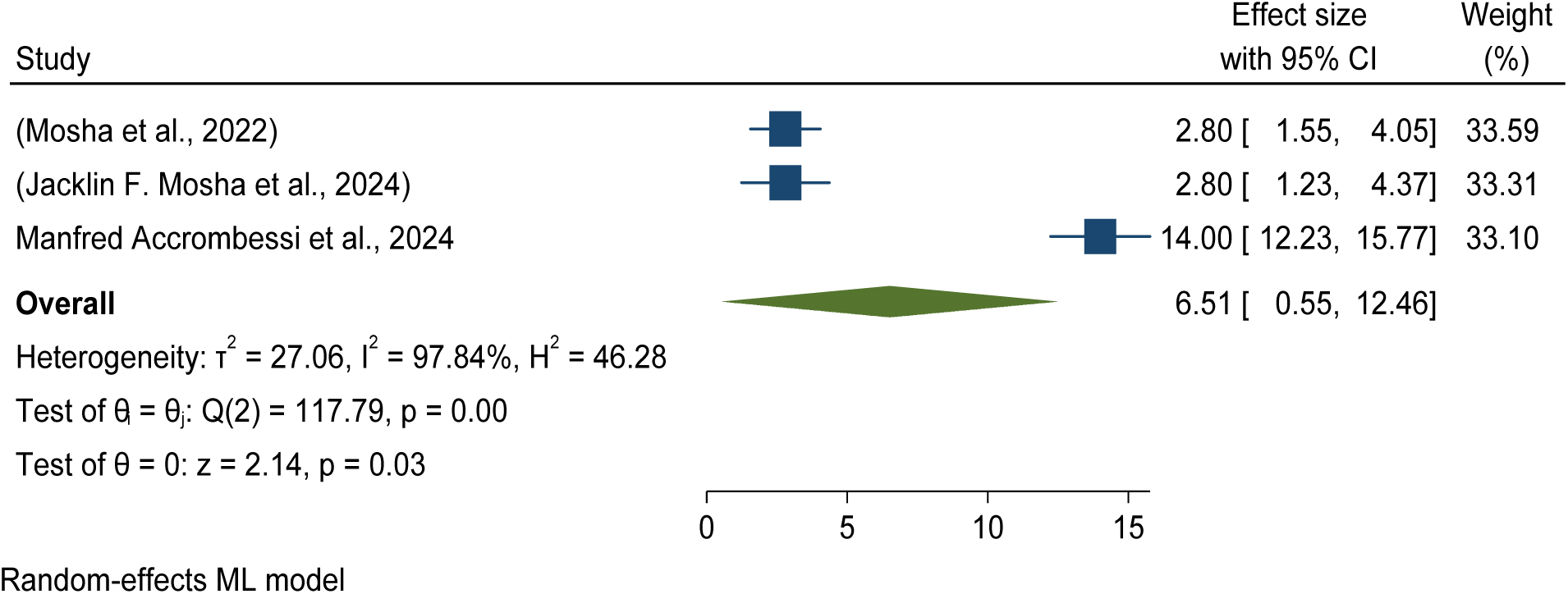
Forest plots showed baseline pooled mean indoor vectors density per household per night using chlorfenapyr LLINs for malaria control in Africa in 2024.

**Figure 20:**
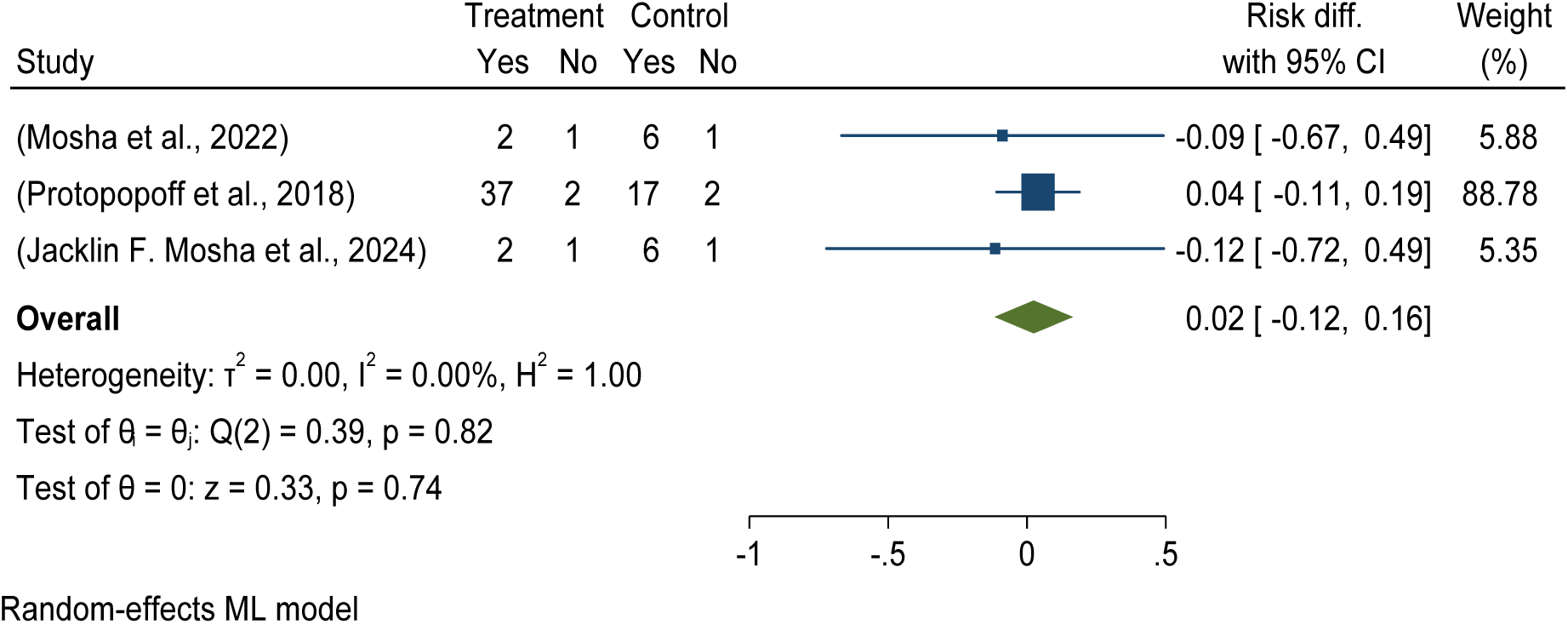
Forest plot shows the effectiveness and efficacy of piperonyl butoxide LLINs in reducing mean indoor vector density per household per night compared to pyrethroid-only LLINs in Africa in 2024

### Effectiveness and efficacy of *piperonyl butoxide* long-lasting insecticidal nets (LLINs) with no difference in reducing mean indoor vector density per household per night compared to pyrethroid-only LLINs

The Forest plot demonstrates that piperonyl butoxide LLINs significantly reduce mean indoor vector density in Africa by 2% compared to standard care/pyrethroid-only LLINs in 2024.

### Pooled prevalence of mean indoor vectors density per household per night using piperonyl butoxide LLINs

The pooled prevalence of mean indoor vector density was 13.49 per household per night in piperonyl butoxide LLINs compared to 12.78 per household per night in pyrethroid-only LLINs (see details in Figures 21 and 22).

**Figure 21:**
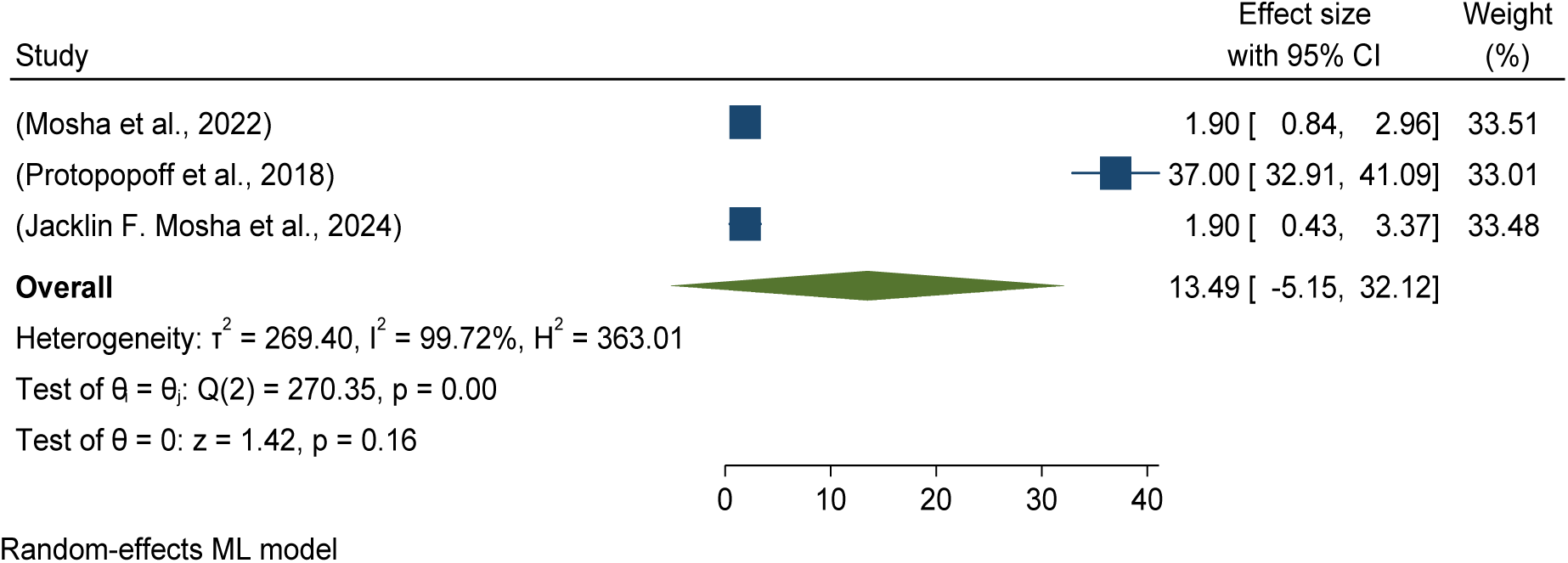
Forest plots showed baseline pooled mean indoor vectors density per household per night using piperonyl butoxide LLINs for malaria control in Africa in 2024.

**Figure 22:**
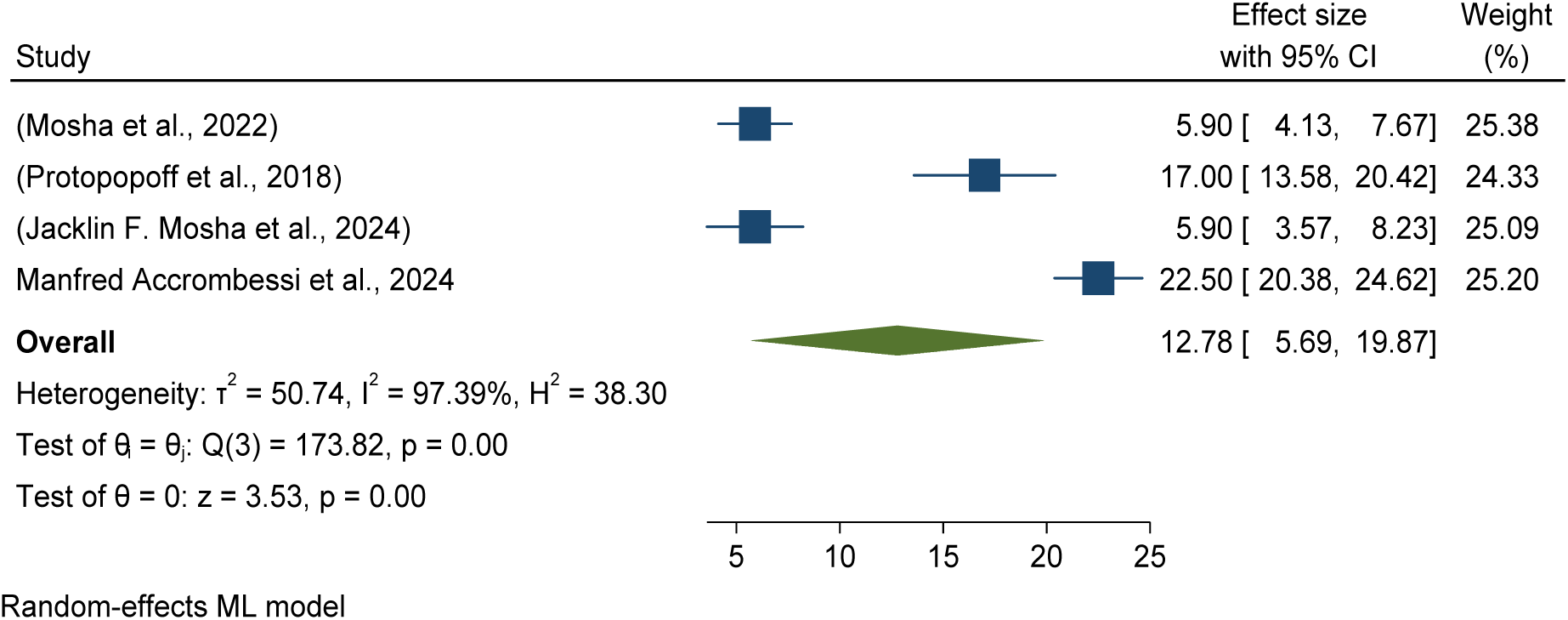
Forest plots showed baseline pooled mean indoor vectors density per household per night using pyrethroid-only LLINs for malaria control in Africa in 2024.

### Pooled prevalence of mean indoor vectors density per household per night using pyrethroid-only LLINs

The study found that the pooled prevalence of mean indoor vector density using pyrethroid- only long lasting insecticidal nets (LLINs) nets was 12.78 per household per night with (95% CI: 5.69, 19.87%) in Africa 2024 (See details in Figure 22).

### Baseline sporozoite rate is the proportion of vectors infected with malaria parasite Pooled effectiveness and efficacy of pyriproxyfen long-lasting insecticidal nets (LLINs) versus pyrethroid-only LLINs for sporozoite rate reduction

Forest plots showed that pyriproxyfen intervention effectively reduces the sporozoite rate by 2% compared to standard or pyrethroid-only LLINs (RR = -0.02 with a 95%CI of -0.37, 0.33). See details in Figure 23.

**Figure 23:**
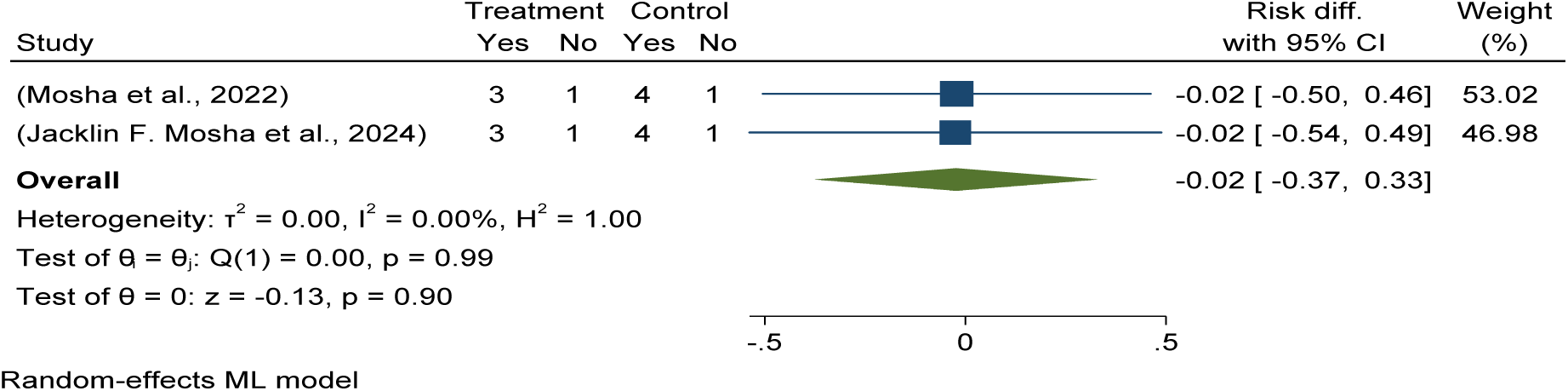
Forest plot shows the effectiveness and efficacy of pyriproxyfen long-lasting insecticidal nets (LLINs) in sporozoite rate reduction compared to pyrethroid-only LLINs in Africa in 2024.

### Pooled prevalence of sporozoite rate among pyriproxyfen LLINs intervention

The pooled prevalence of sporozoite rate was 3.30 in pyriproxyfen LLIN intervention use compared to 4.57 in pyrethroid-only LLINs standard. (See details in Figures 24 and 28).

**Figure 24:**
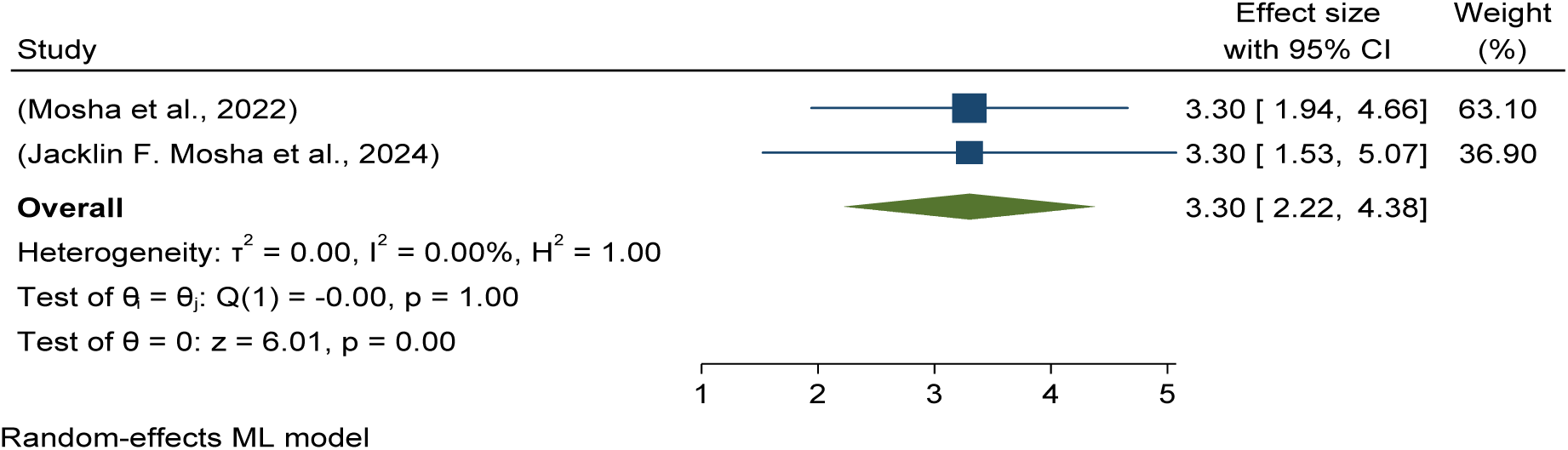
Forest plots showed baseline pooled sporozoite rate among pyriproxyfen LLINs intervention for malaria control in Africa in 2024.

### Pooled effectiveness and efficacy of chlorfenapyr long-lasting insecticidal nets (LLINs) versus pyrethroid-only LLINs for sporozoite rate reduction

This study found that chlorfenapyr intervention effectively reduces the sporozoite rate by 5% as compared to standard or pyrethroid-only LLINs, as shown in Figure 25.

**Figure 25:**
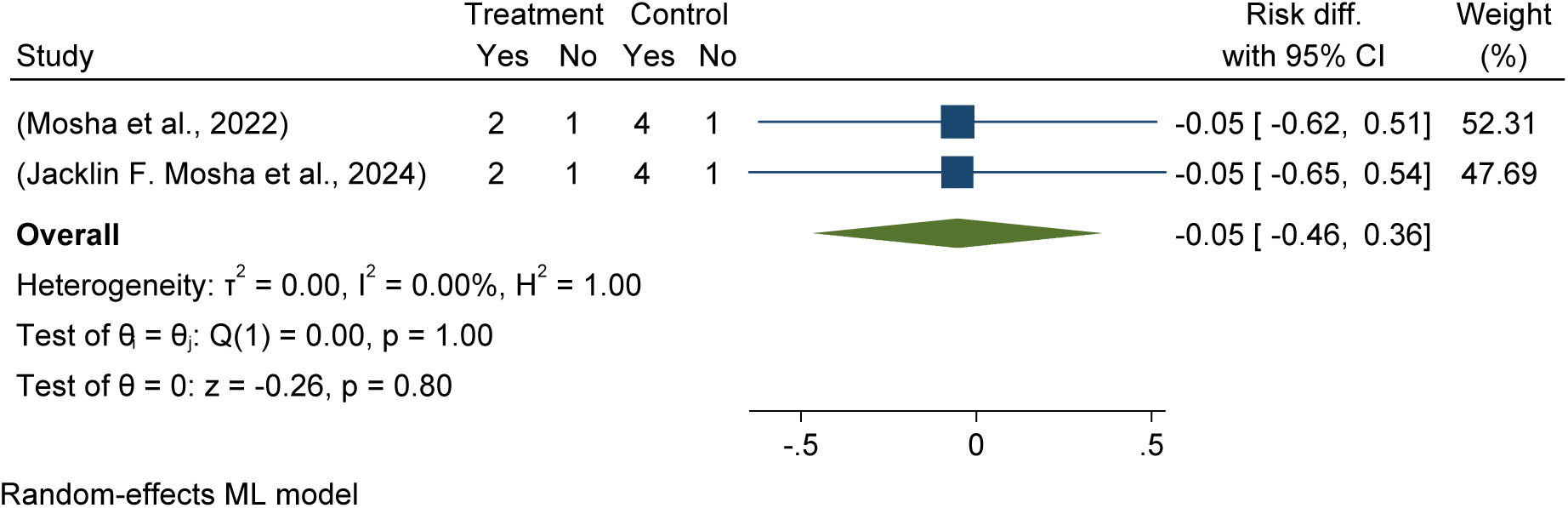
Forest plot shows the effectiveness and efficacy of chlorfenapyr long-lasting insecticidal nets (LLINs) in sporozoite rate reduction compared to pyrethroid-only LLINs in Africa in 2024.

### Pooled prevalence of sporozoite rate among chlorfenapyr LLINs intervention

The pooled prevalence of sporozoite rate was 2.20 in chlorfenapyr LLIN intervention use compared to 4.57 in pyrethroid-only LLINs standard. (See details in Figures 26 and 28).

**Figure 26:**
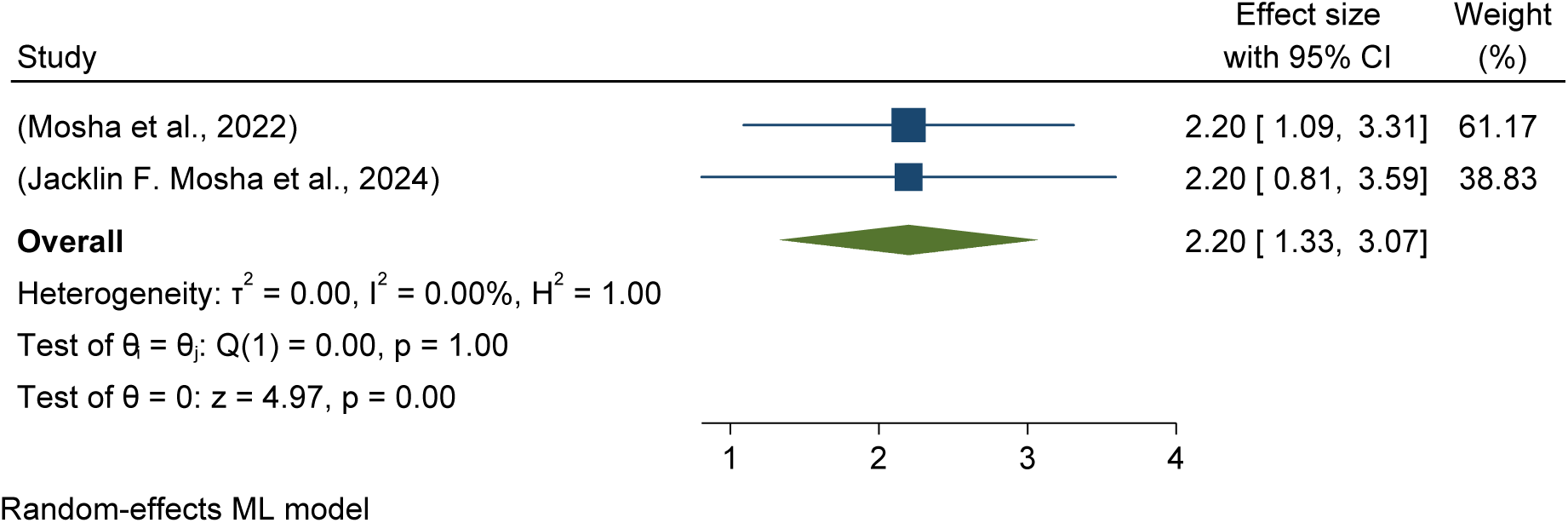
Forest plots showed baseline pooled sporozoite rate among chlorfenapyr LLINs intervention for malaria control in Africa in 2024.

### Pooled prevalence of sporozoite rate among Piperonyl butoxide LLINs intervention

This study found that Piperonyl butoxide intervention effectively reduces the sporozoite rate by 2% as compared to standard or pyrethroid-only LLINs, as shown in Figure 26.

### Pooled prevalence of sporozoite rate among Piperonyl butoxide LLINs intervention

The pooled prevalence of sporozoite rate was 3.50 in Piperonyl butoxide LLIN intervention use compared to 4.57 in pyrethroid-only LLINs standard. (See details in Figures 27 and 29).

**Figure 27:**
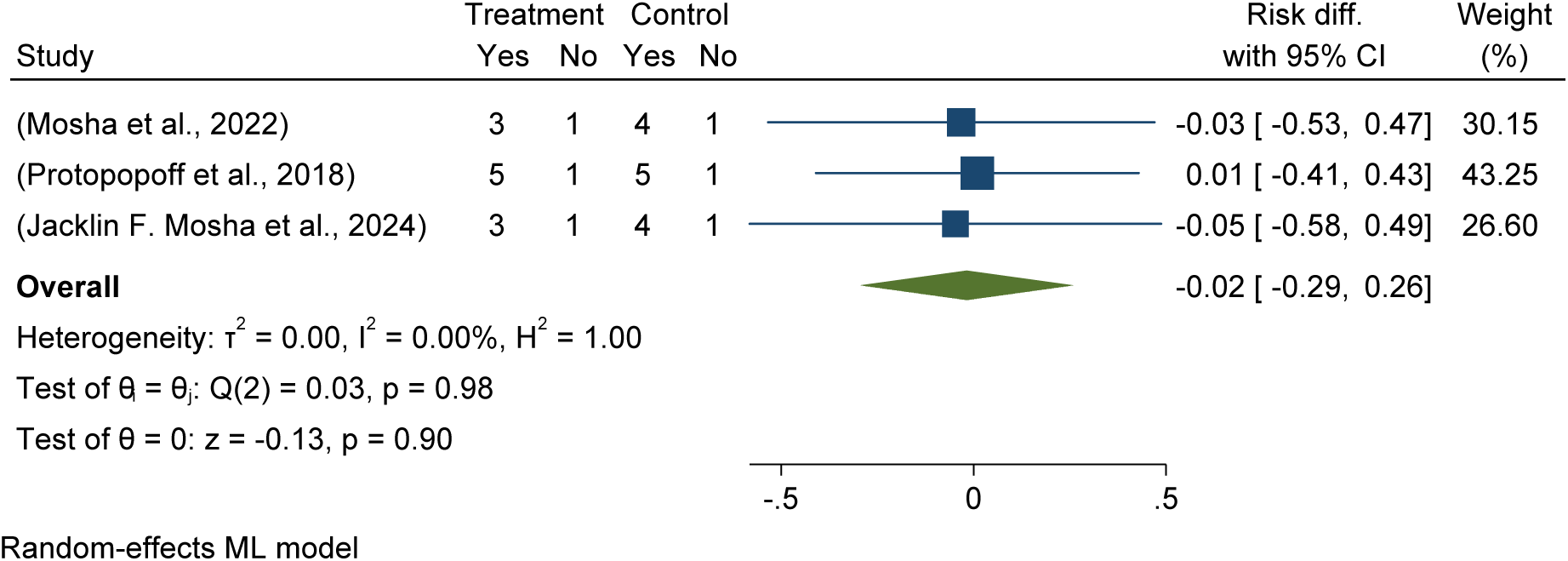
Forest plot shows the effectiveness and efficacy of Piperonyl butoxide long-lasting insecticidal nets (LLINs) in sporozoite rate reduction compared to pyrethroid-only LLINs in Africa in 2024.

**Figure 28:**
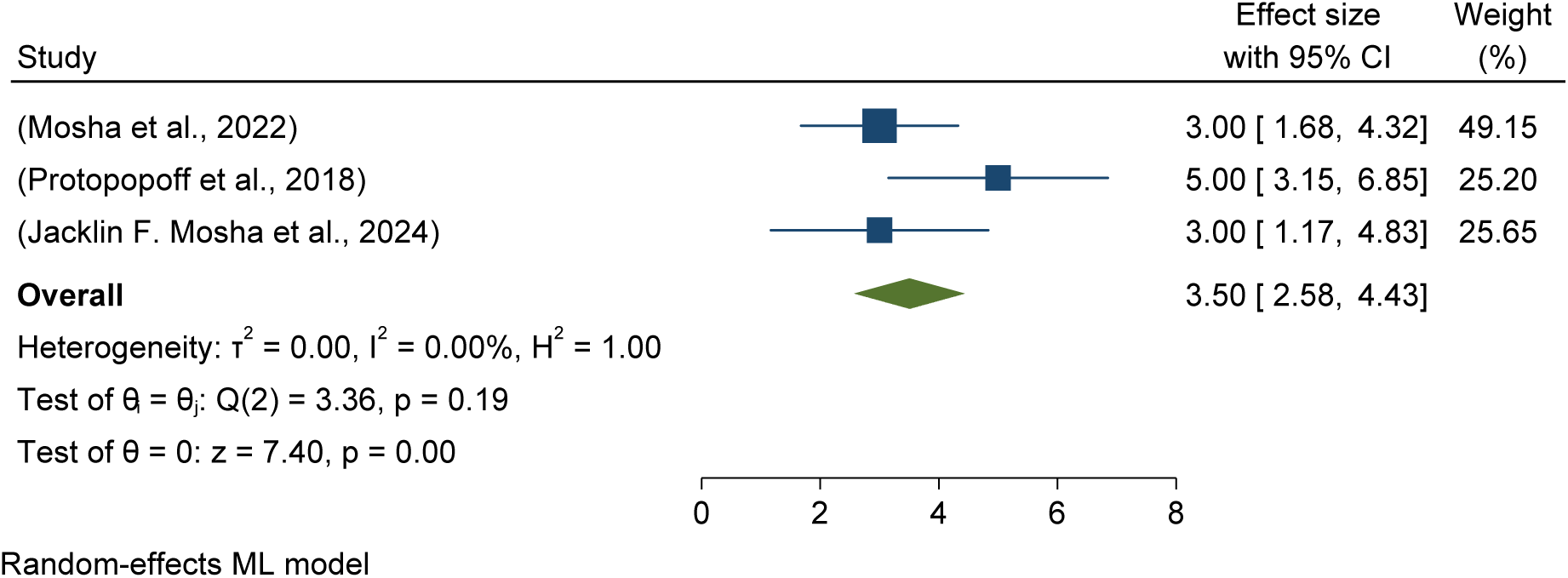
Forest plots showed baseline pooled sporozoite rate among Piperonyl butoxide LLINs intervention for malaria control in Africa in 2024.

**Figure 29:**
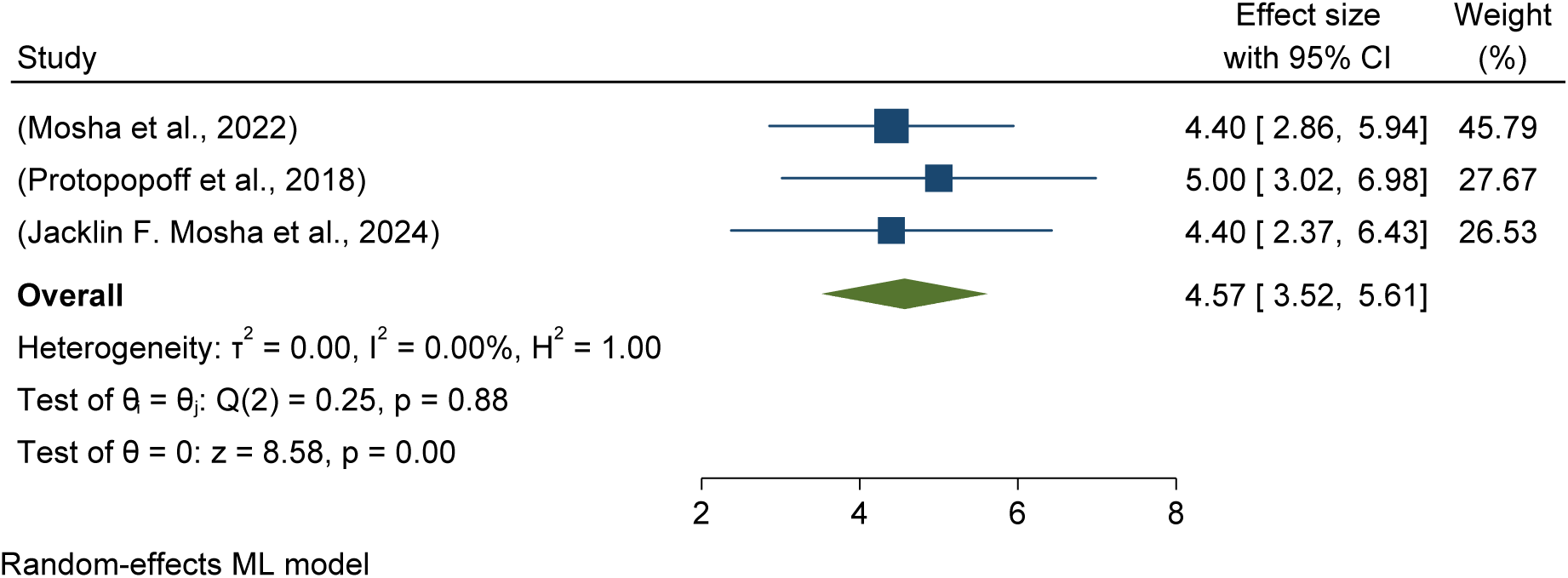
Forest plots showed baseline pooled sporozoite rate among Pyrethroid-only LLINs intervention for malaria control in Africa in 2024.

### Pooled prevalence of sporozoite rate among Pyrethroid-only LLINs intervention

The pooled prevalence of sporozoite rate was 4.57 in pyrethroid-only LLINs standard. (See details in Figure 29).

### Mean entomological inoculation rate per household per night (MEIR)

This meta-analysis determined entomological outcomes effectiveness and efficacy interterms of mean entomological inoculation rate per household per night (MEIR) reduction and reported that the Pyriproxyfen LLINs intervention was reduced by 8% as compared to the standard or control of the Pyrethroid-only group (AOR = -0.08, 95% CI = -1.03, 0.87). See details in Figure 30.

**Figure 30:**
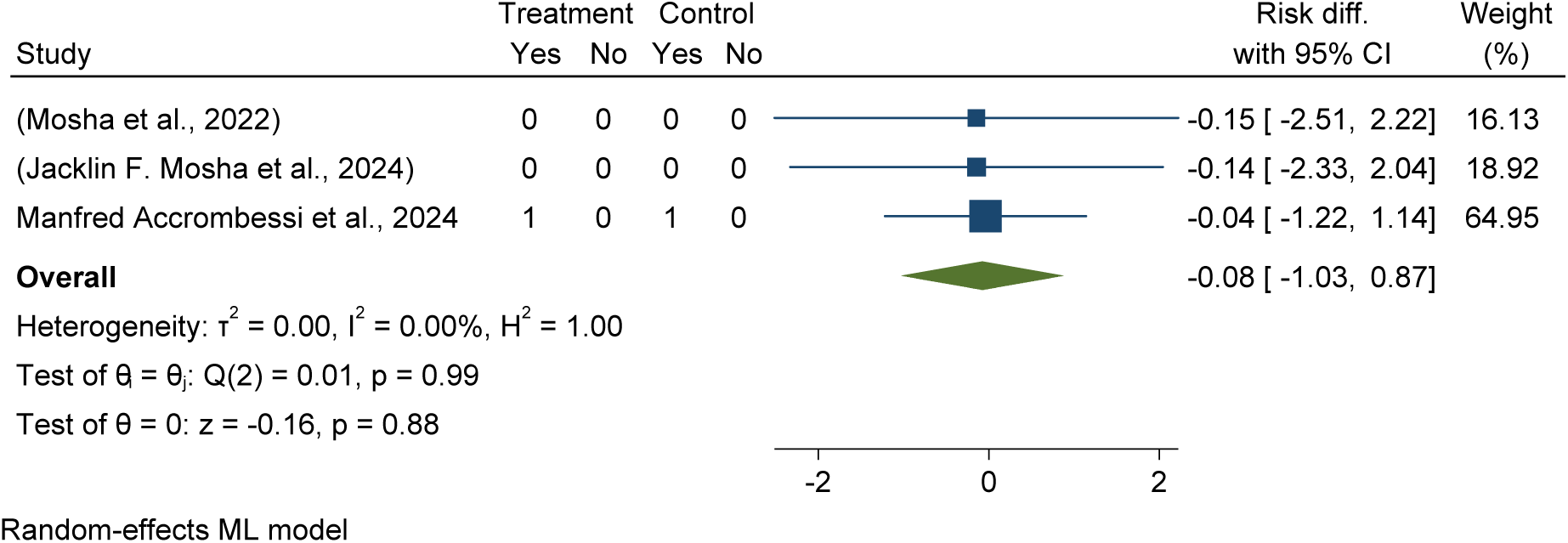
Forest plot shows the effectiveness and efficacy of *Pyriproxyfen* long-lasting insecticidal nets (LLINs) in mean entomological inoculation rate per household per night (MEIR) reduction compared to pyrethroid-only LLINs in Africa in 2024.

This study determined the pooled mean entomological inoculation rate per night (23 per 100 households per night with a 95% CI of 0.01, 0.48) in the Pyriproxyfen LLINs intervention versus 56 per 100 households per night in the Pyrethroid-only group with a 95% CI of 0.23, 0.88). (See details in Figures 31 and 36).

**Figure 31:**
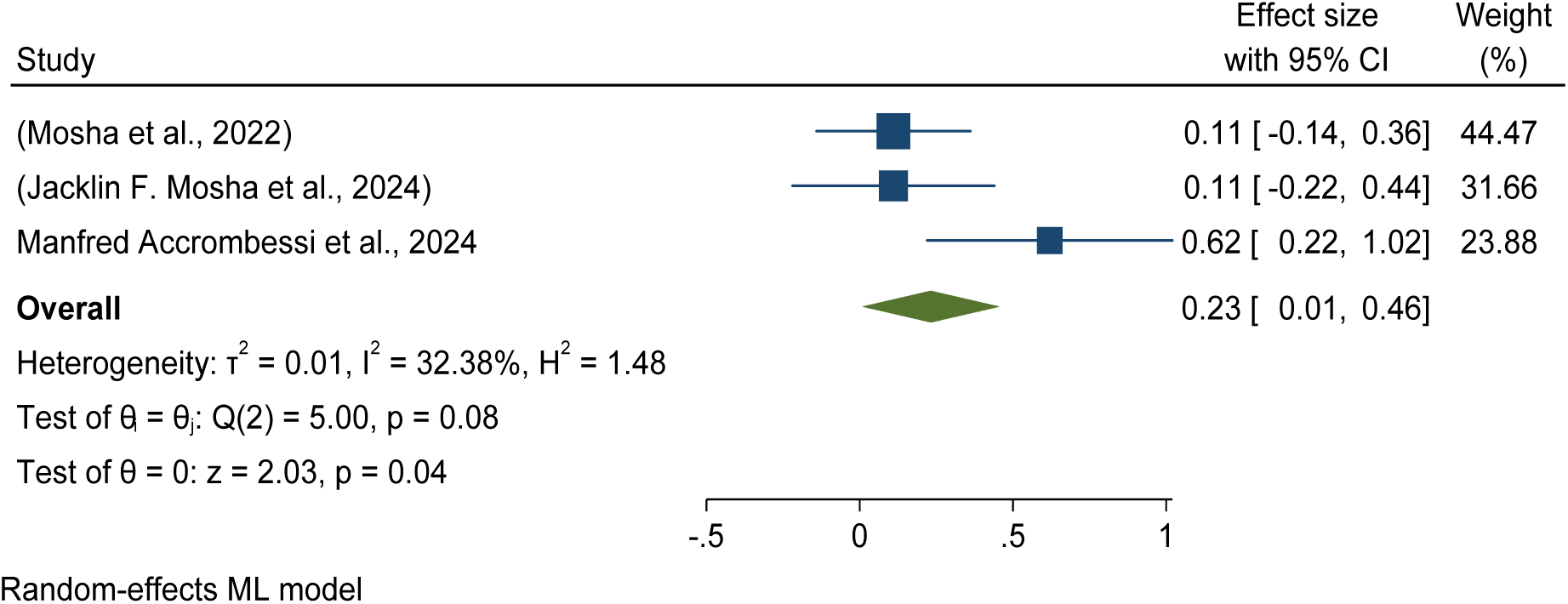
Forest plots showed baseline pooled mean entomological inoculation rate per household per night (MEIR) among Pyriproxyfen long-lasting insecticidal nets (LLINs) intervention for malaria control in Africa in 2024.

This meta-analysis determined entomological outcomes effectiveness and efficacy in terms of mean per household per night inoculation rate reduction and reported that the chlorfenapyr group was reduced by 12% as compared to the standard or control of the pyrethroid-only group (AOR = -.12, 95% CI = -1.21, 0.97). See details in Figure 32.

**Figure 32:**
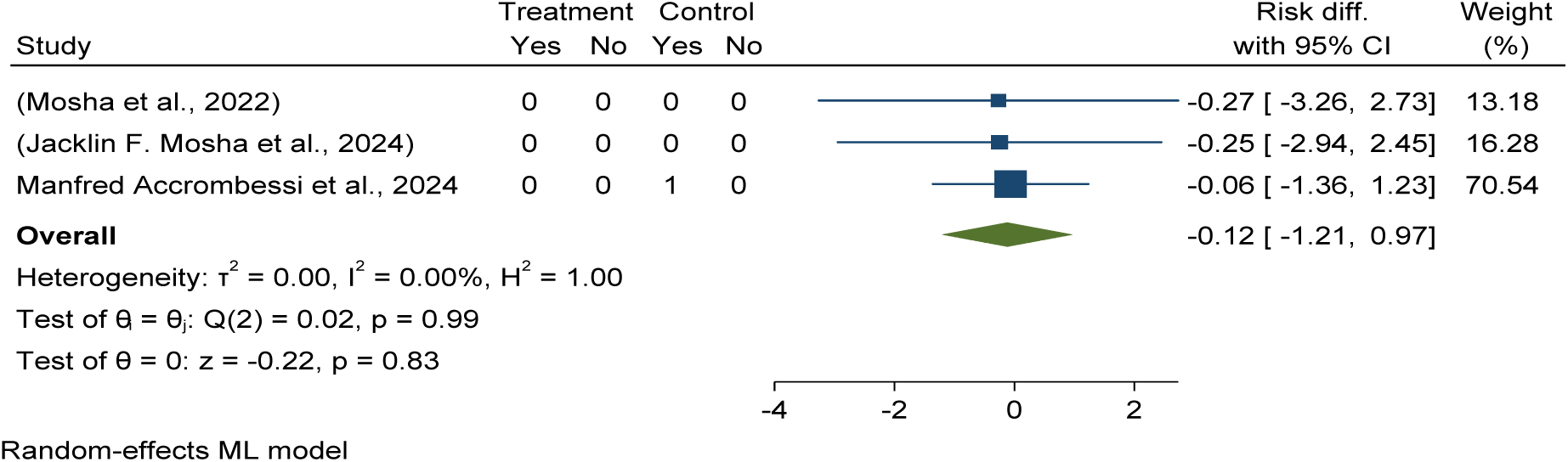
Forest plot shows the effectiveness and efficacy of *chlorfenapyr* long-lasting insecticidal nets (LLINs) in mean entomological inoculation rate per household per night (MEIR) reduction compared to pyrethroid-only LLINs in Africa in 2024.

This study determined the pooled mean entomological inoculation rate per night (8 per 100 households per night with a 95% CI of (-0.03, 0.20) in the chlorfenapyr LLINs intervention versus 56 per 100 households per night in the Pyrethroid-only group with a 95% CI of 0.23, 0.88). (See details in Figures 33 and 36).

**Figure 33:**
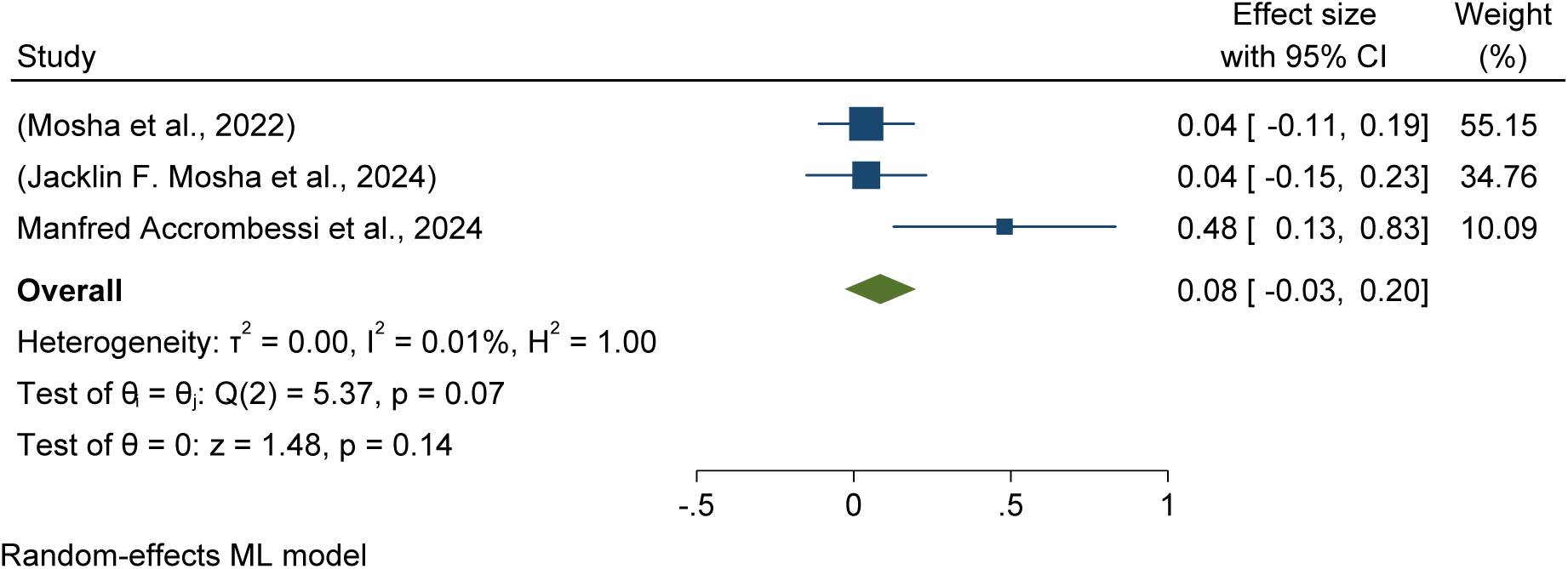
Forest plots showed baseline pooled mean entomological inoculation rate per household per night (MEIR) among Pyriproxyfen long-lasting insecticidal nets (LLINs) intervention for malaria control in Africa in 2024.

This meta-analysis determined entomological outcomes effectiveness and efficacy in terms of mean per household per night inoculation rate reduction and reported that the Piperonyl butoxide group was reduced by 21% as compared to the standard or control of the pyrethroid-only group (AOR = -.21, 95% CI = -1.95, 1.53). See details in Figure 34.

**Figure 34:**
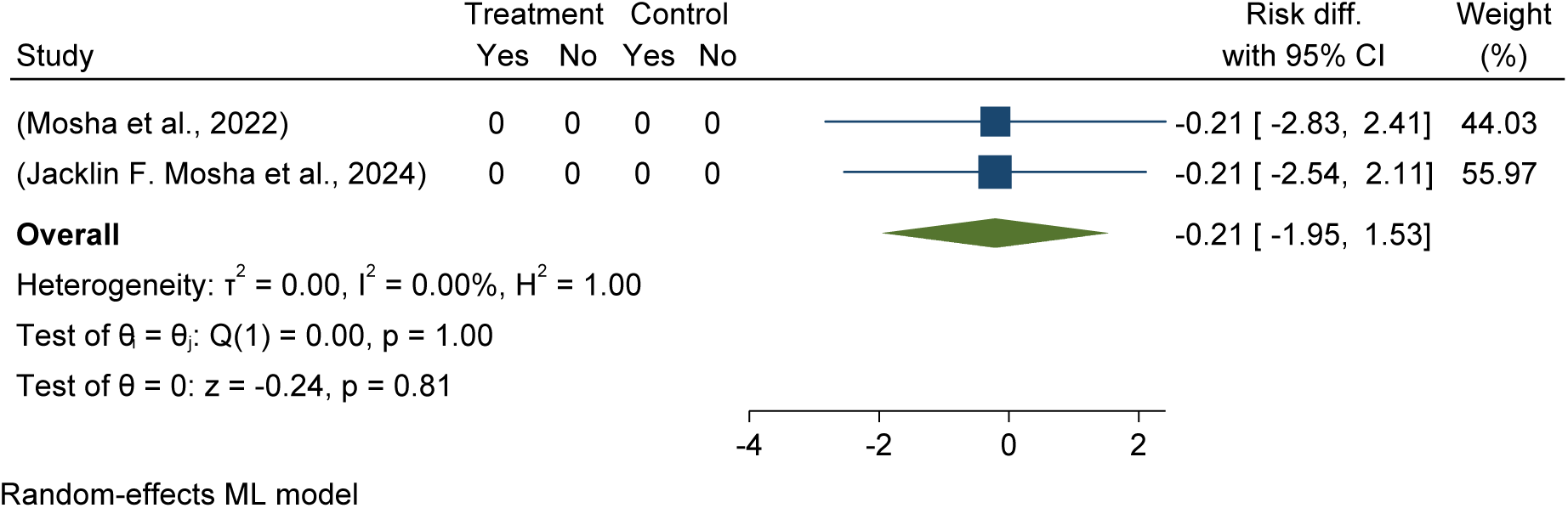
Forest plots showed baseline pooled mean entomological inoculation rate per household per night (MEIR) among *Piperonyl butoxide* long-lasting insecticidal nets (LLINs) intervention for malaria control in Africa in 2024

This study determined the pooled mean entomological inoculation rate per night (7 per 100 households per night with a 95% CI of (-0.10, 0.24) in the Piperonyl butoxide LLINs intervention versus 56 per 100 households per night in the Pyrethroid-only group with a 95% CI of 0.23, 0.88). (See details in Figures 35 and 36).

**Figure 35:**
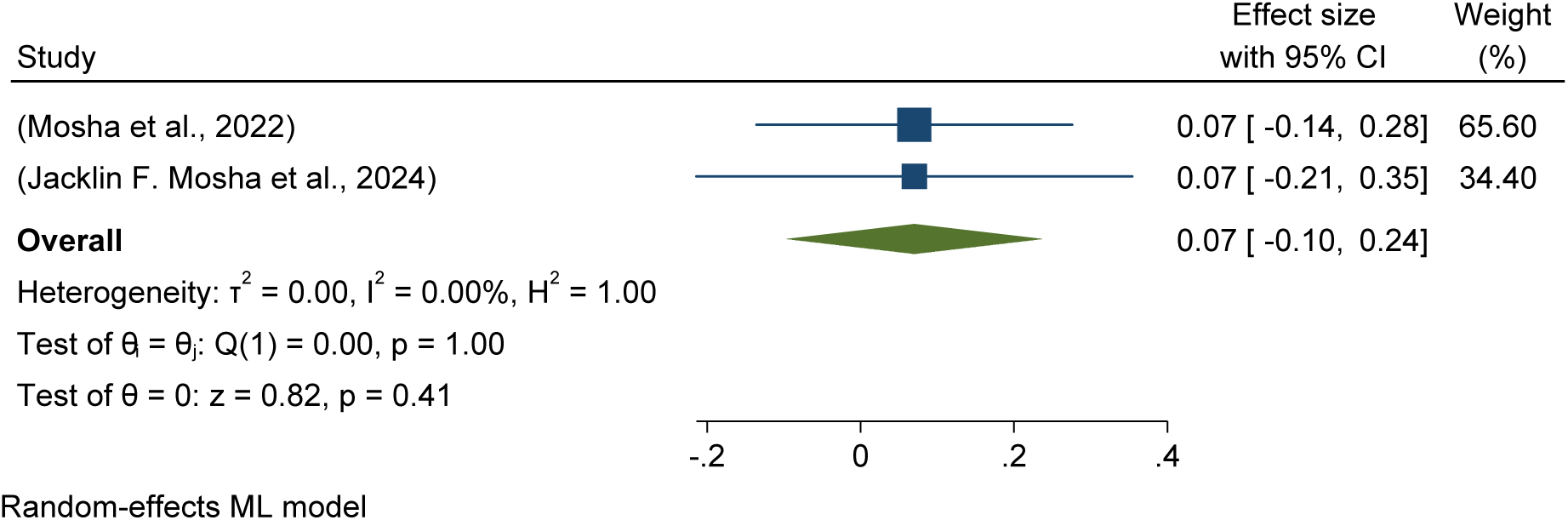
Forest plots showed baseline pooled mean entomological inoculation rate per household per night (MEIR) among *Piperonyl butoxide* long-lasting insecticidal nets (LLINs) intervention for malaria control in Africa in 2024.

**Figure 36:**
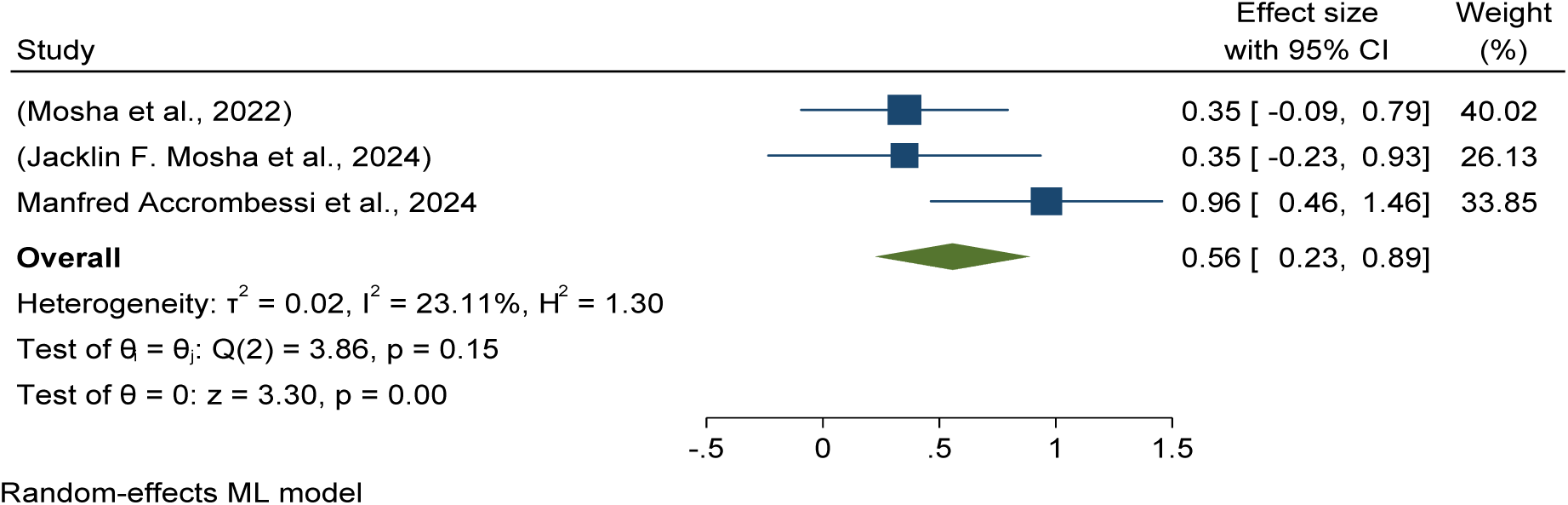
Forest plots showed baseline pooled mean entomological inoculation rate per household per night (MEIR) among *pyrethroid-only* long-lasting insecticidal nets (LLINs) intervention for malaria control in Africa in 2024.

This study determined the pooled mean entomological inoculation rate per night (56 per 100 households per night in the standard or control of the pyrethroid-only group with a 95% CI of 0.23, 0.88). (See details in Figure 36).

### Pooled post-intervention effectiveness and efficacy of Pyriproxyfen long-lasting insecticidal nets (LLINs) versus pyrethroid-only LLINs malaria infection reduction in Africa

This study evaluated the post-intervention effectiveness and efficacy of Pyriproxyfen long-lasting insecticidal nets (LLINs) in malaria infection risk reduction in children over different durations (6, 12, 18, 24, and 36 months) as compared placebo/standard pyrethroid-only LLINs revealed that had no difference in reducing risk of infection with (RR = -0.00 with 95%CI - 0.03,0.02). See details in Figure 37.

**Figure 37:**
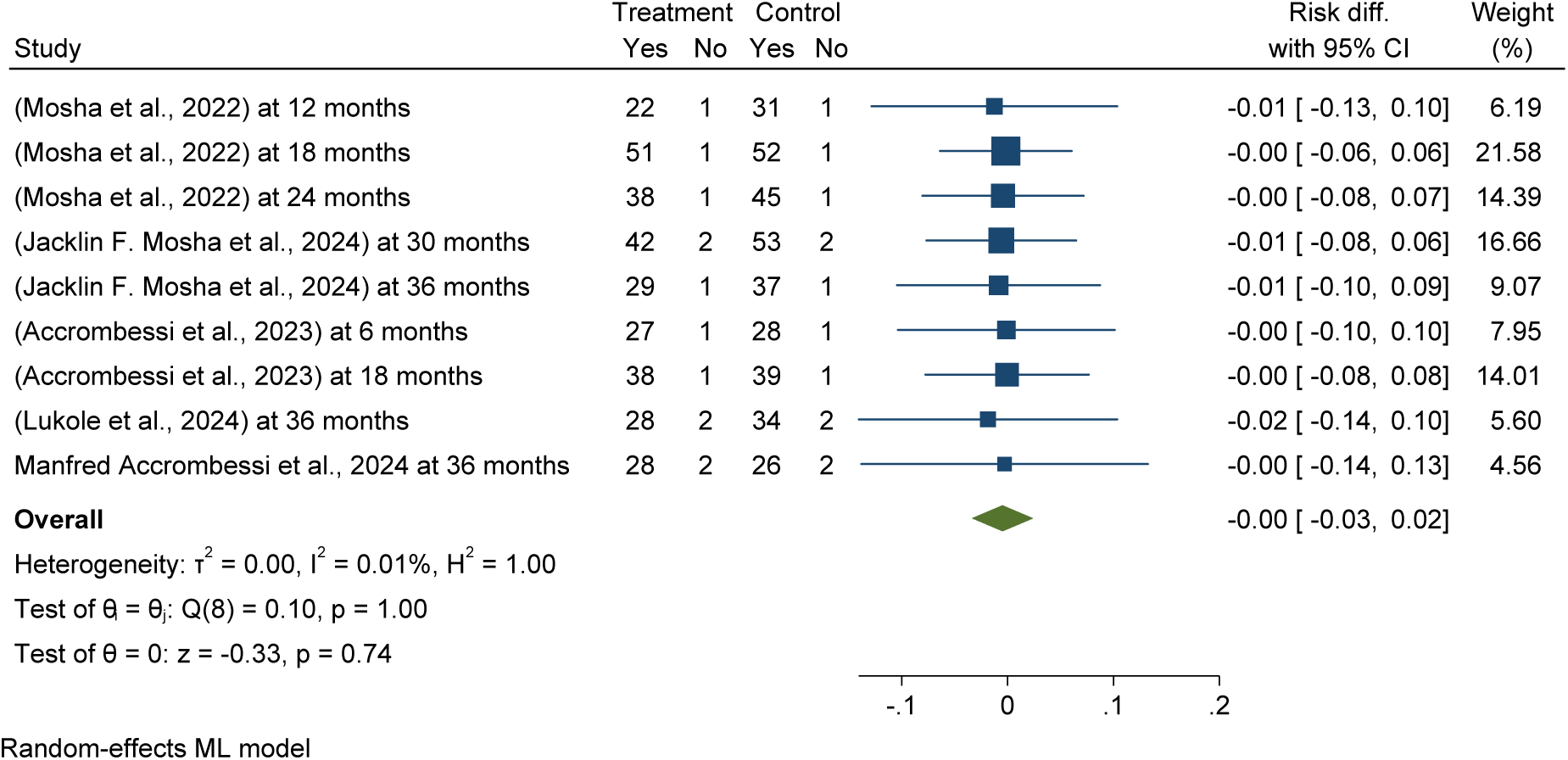
Forest plots shows *Pooled post-intervention effectiveness and efficacy of Pyriproxyfen long-lasting insecticidal nets (LLINs) versus pyrethroid-only LLINs malaria infection reduction in Africa 2024*.

The Subgroup analysis of post-intervention follow up effectiveness and efficacy of pyriproxyfen long-lasting insecticidal nets (LLINs) showed a significant reduction in malaria infection among children over different durations, with a 1% reduction at twelve months and 1% at 36 months post-distribution of LLINs, compared to pyrethroid-only LLINs (ARR = - 0.01, 95% CI = -0.08, 0.08), but has no difference in overall effectives and efficacy. See details in Figure 38

**Figure 38:**
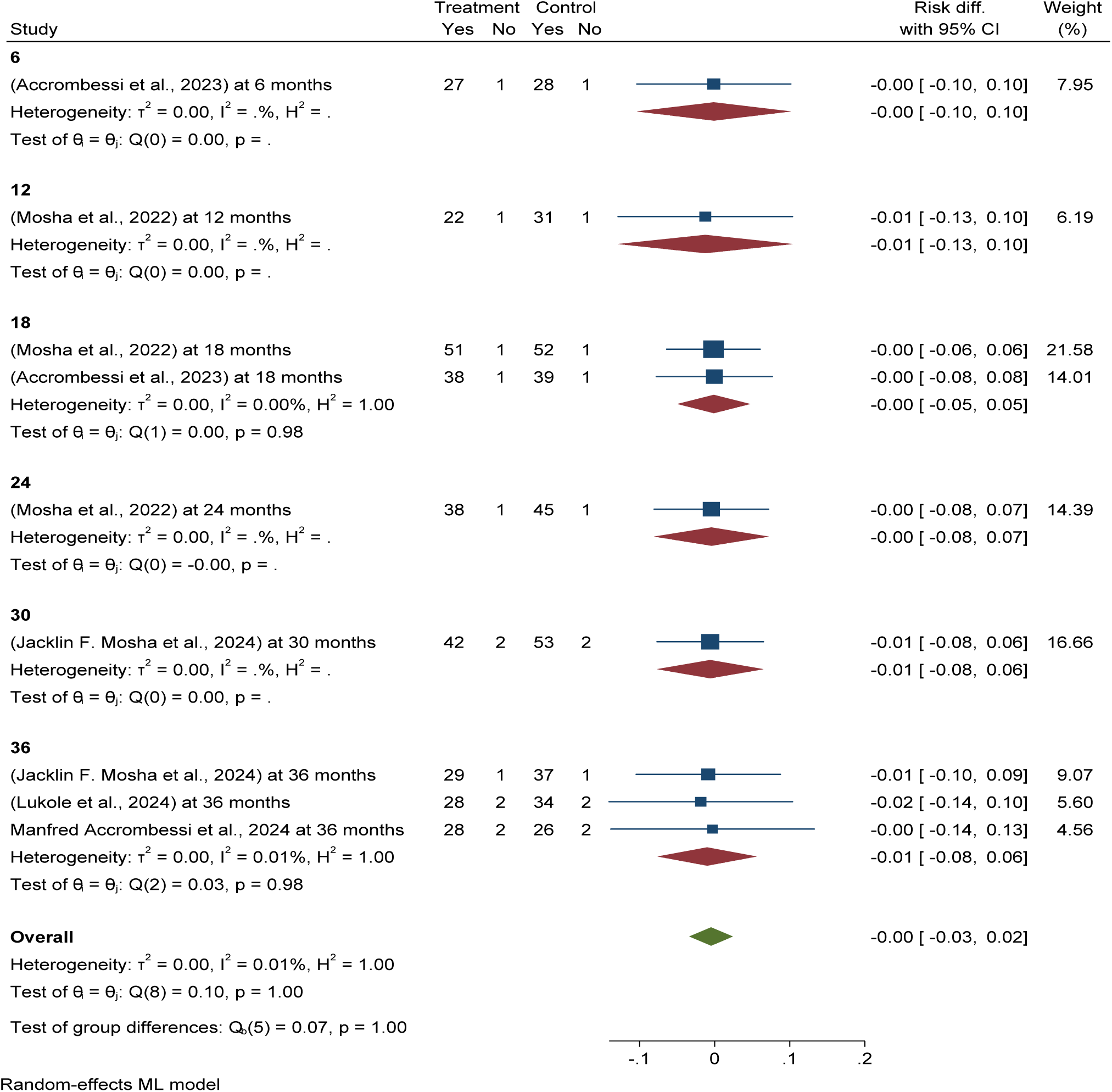
Forest plots shows s*ubgroup analysis of post-intervention follow up effectiveness and efficacy of Pyriproxyfen long-lasting insecticidal nets (LLINs) versus pyrethroid-only LLINs malaria infection reduction in Africa 2024*.

**Figure 39:**
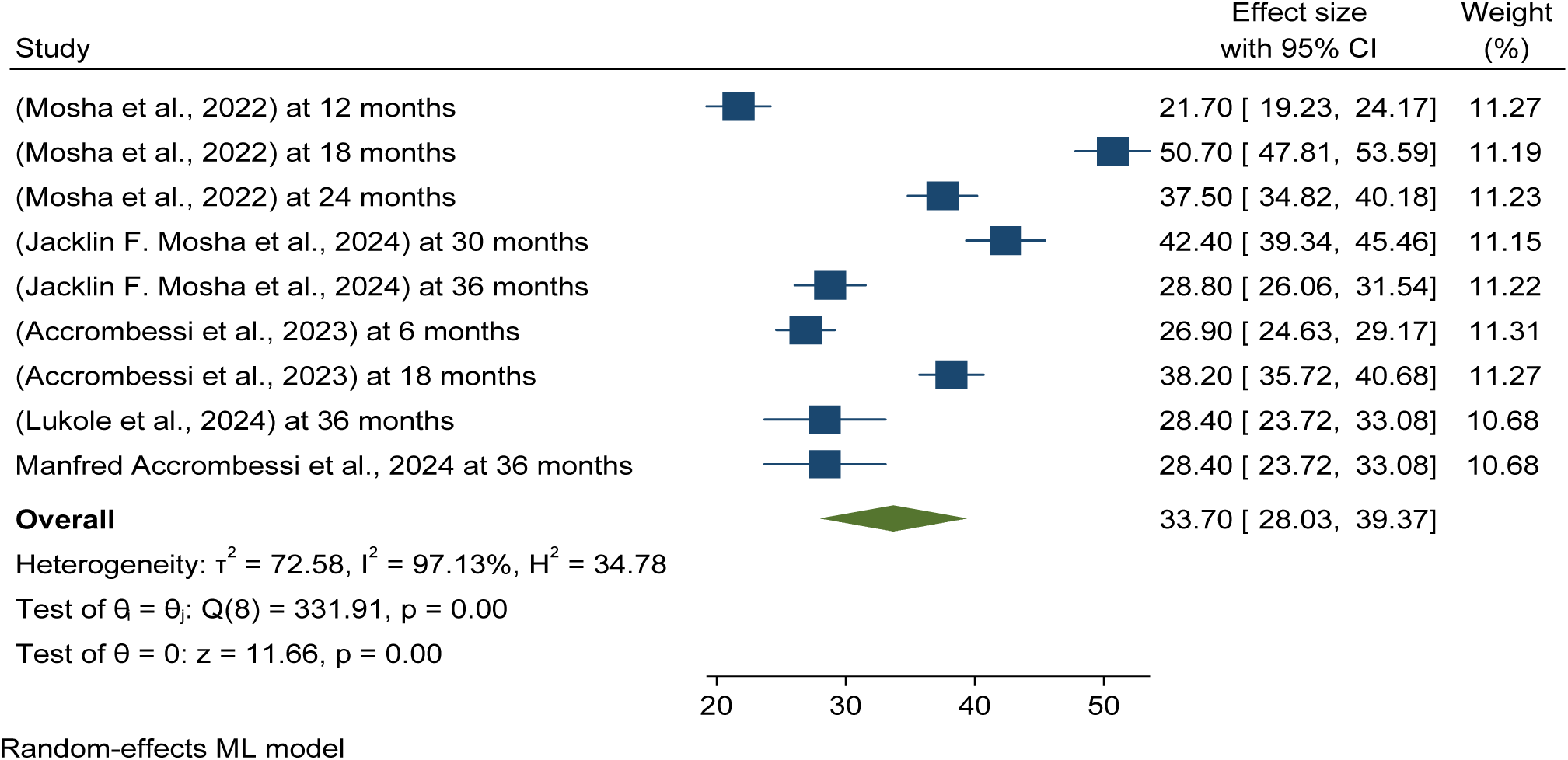
Forest plots shows *Pooled post-intervention malaria infection prevalence in selected children using pyriproxyfen long-lasting insecticidal nets (LLINs) as malaria control in Africa 2024*

### Pooled post-intervention Malaria infection prevalence in selected children using different long-lasting insecticidal nets (LLINs) as malaria control in Africa

This randomized control trial meta-analysis found that the pooled prevalence of post-intervention malaria infection among children using pyriproxyfen long-lasting insecticidal nets (LLINs) over different durations (6, 12, 18, 24, and 36 months) was 33.70 per 100 children (95% CI: 28.03–39.37%), while it was higher in the control group/pyrethroid-only LLINs (40.84% per 100 children) (95% CI: 32.45%, 49.22%) in Africa (see details in Figure 38 and Figure 49).

Publication bias was checked using funnel plots looking at symmetrical distribution, and it was objectively verified using Egger’s regression test, which revealed that there was no publication bias (p < 0.174) (Figure 40).

**Figure 40:**
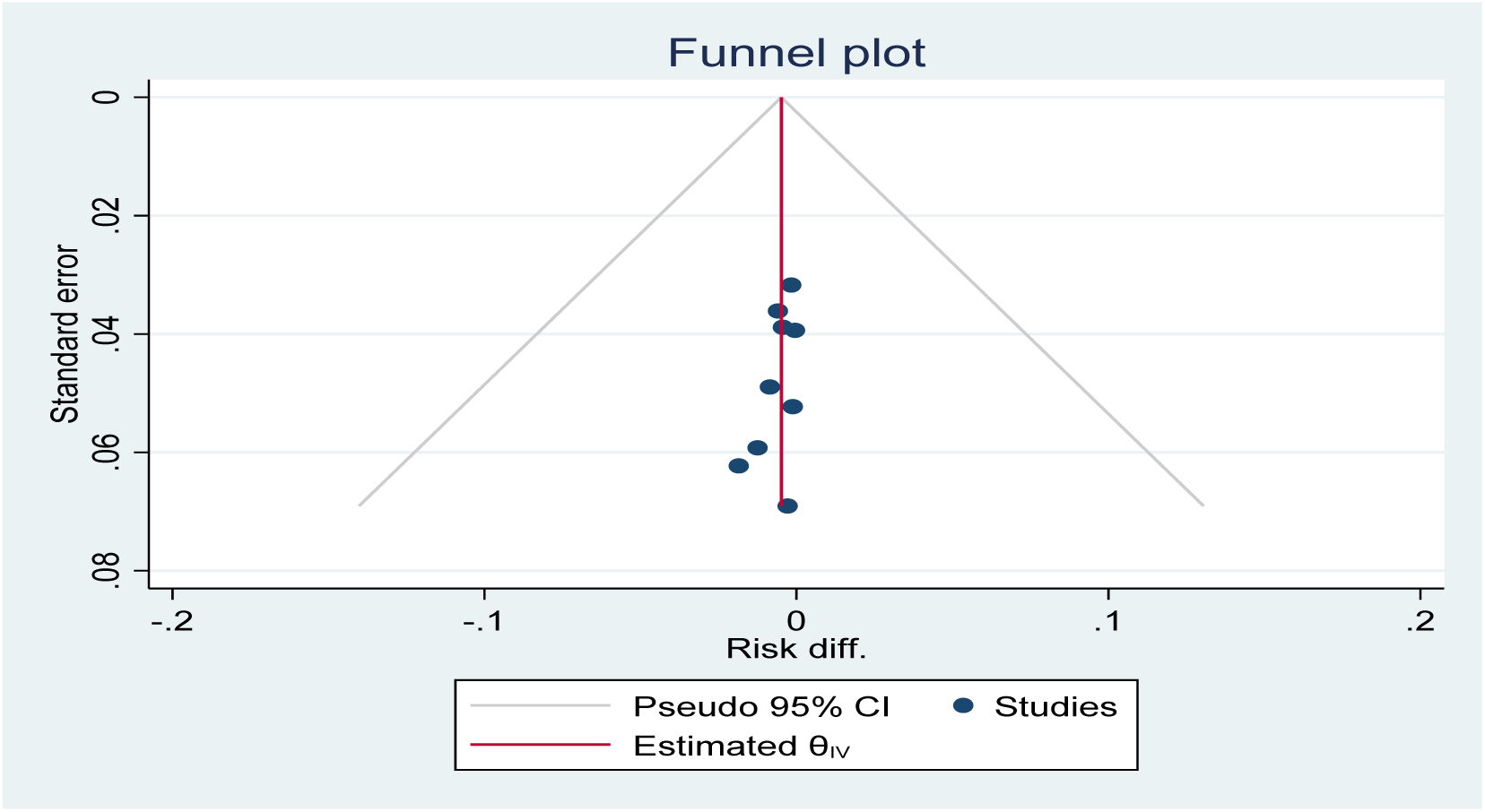
Funnel plot showing the distribution of included studies pooled malaria infection risk reduction among children using Pyriproxyfen long-lasting insecticidal nets (LLINs) versus pyrethroid-only LLINs for malaria control in Africa in 2024.

### Pooled post-intervention effectiveness and efficacy of chlorfenapyr long-lasting insecticidal nets (LLINs) versus pyrethroid-only LLINs malaria infection reduction in Africa

This study determined that post intervention effectiveness and efficacy of chlorfenapyr long-lasting insecticidal nets (LLINs) in malaria infection risk reduction in children over different durations (6, 12, 18, 24, and 36 months) was reduced by 1% as compared to the standard or control of the pyrethroid-only group (ARR = -0.01, 95% CI = -0.04, 0.02). See details in Figure 41.

**Figure 41:**
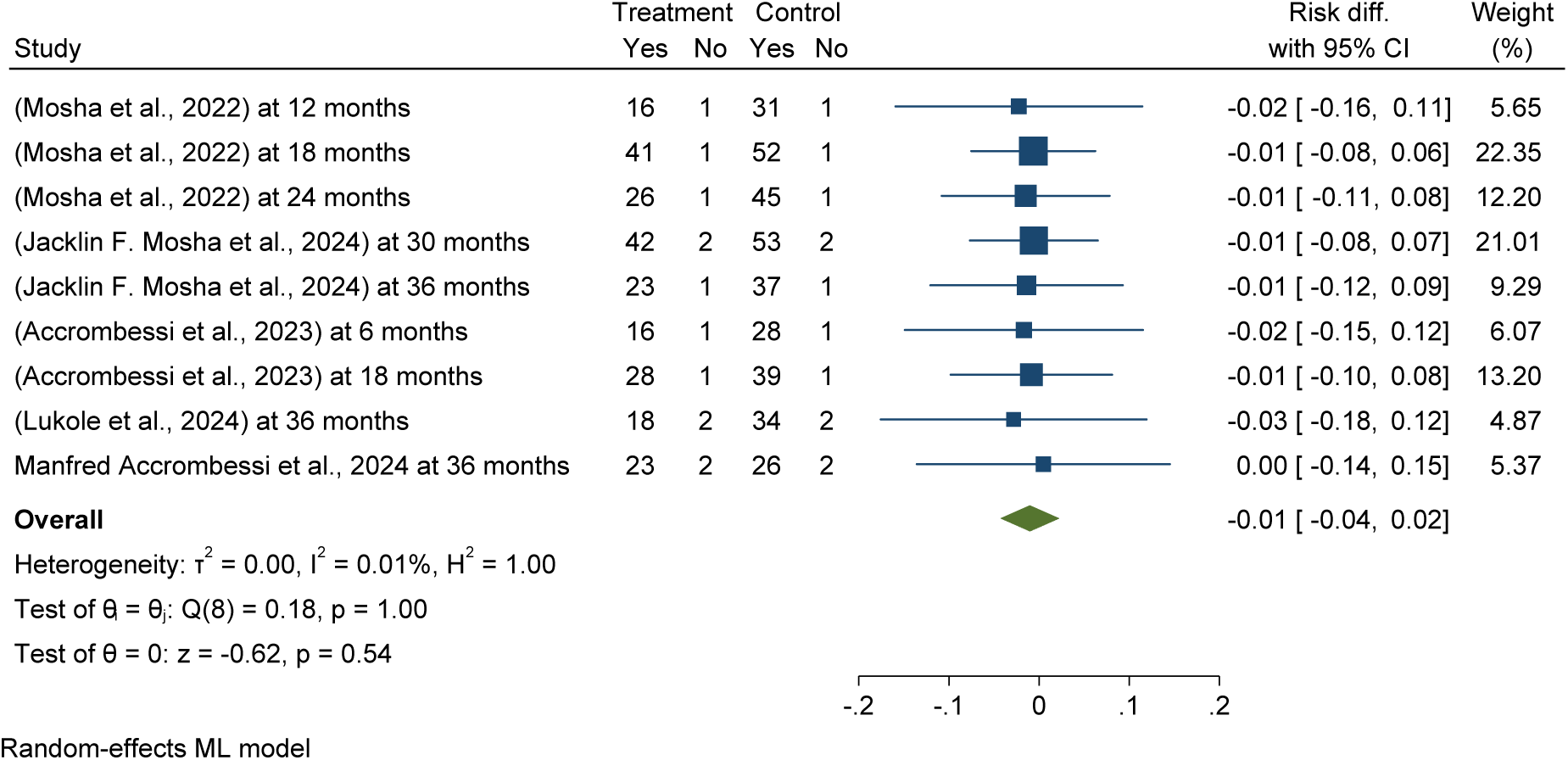
Forest plots shows *Pooled post-intervention effectiveness and efficacy of chlorfenapyr long-lasting insecticidal nets (LLINs) versus pyrethroid-only LLINs malaria infection reduction in Africa 2024*

The Subgroup analysis of post-intervention follow up effectiveness and efficacy of chlorfenapyr long-lasting insecticidal nets (LLINs) showed a significant reduction in malaria infection among children over different durations, with a 2% reduction at twelve months and 1% at 36 months post-distribution of LLINs, compared to pyrethroid-only LLINs (ARR = - 0.01, 95% CI = -0.04, 0.02). See details in Figure 42.

**Figure 42:**
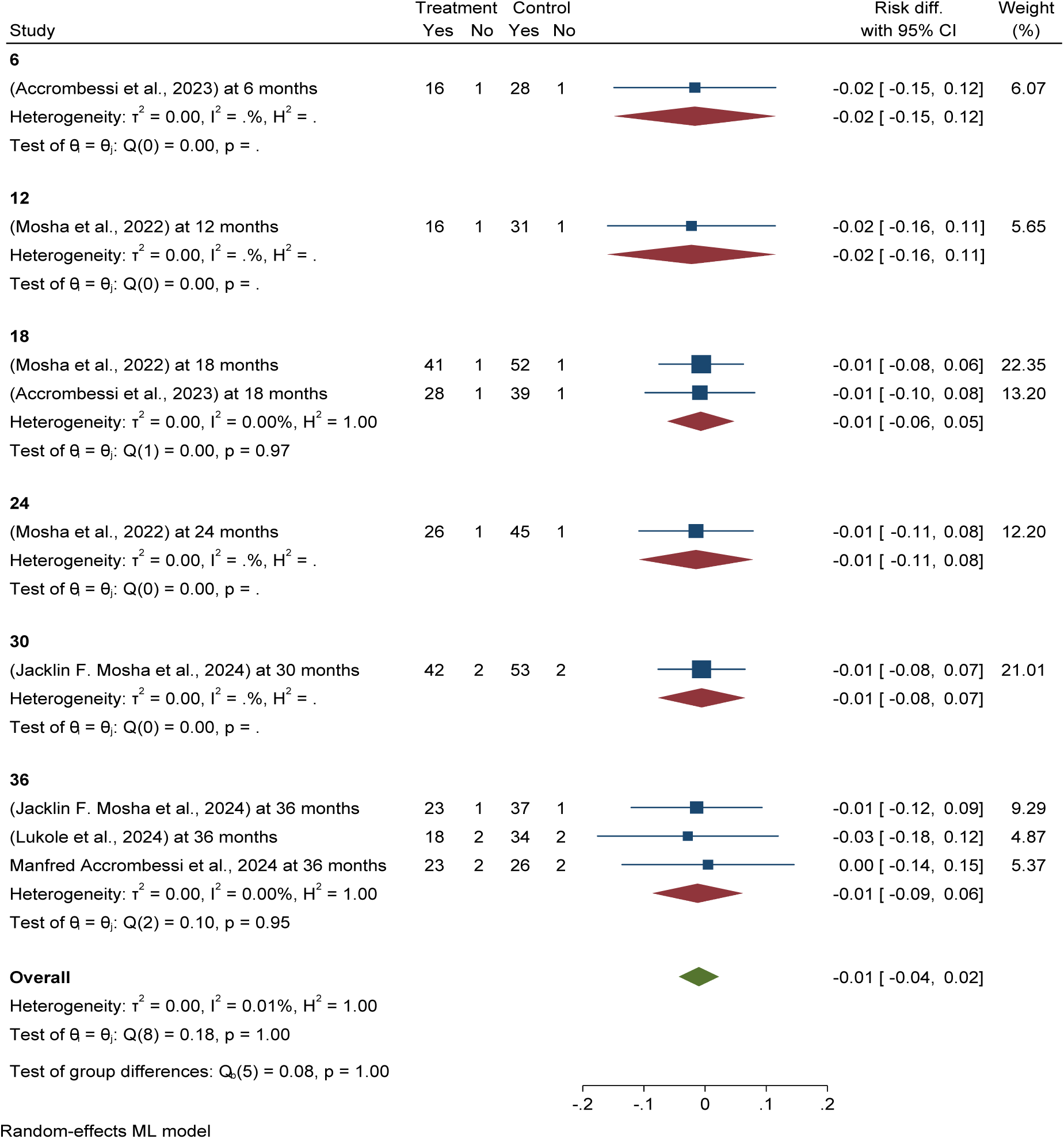
Forest plots shows s*ubgroup analysis of post-intervention follow up effectiveness and efficacy of **chlorfenapyr** long-lasting insecticidal nets (LLINs) versus pyrethroid-only LLINs malaria infection reduction in Africa 2024*.

### Pooled post-intervention malaria infection prevalence in selected children using chlorfenapyr long-lasting insecticidal nets (LLINs) as malaria control in Africa

This randomized control trial meta-analysis found that the pooled prevalence of post-intervention malaria infection among children using chlorfenapyr long-lasting insecticidal nets (LLINs) over different durations (6, 12, 18, 24, and 36 months) was 25.58 per 100 children (95% CI: 19.52–31.64%), while it was doubled in the control group/pyrethroid-only LLINs (40.84% per 100 children) (95% CI: 32.45%, 49.22%) in Africa (see details in Figure 43 and Figure 49).

**Figure 43:**
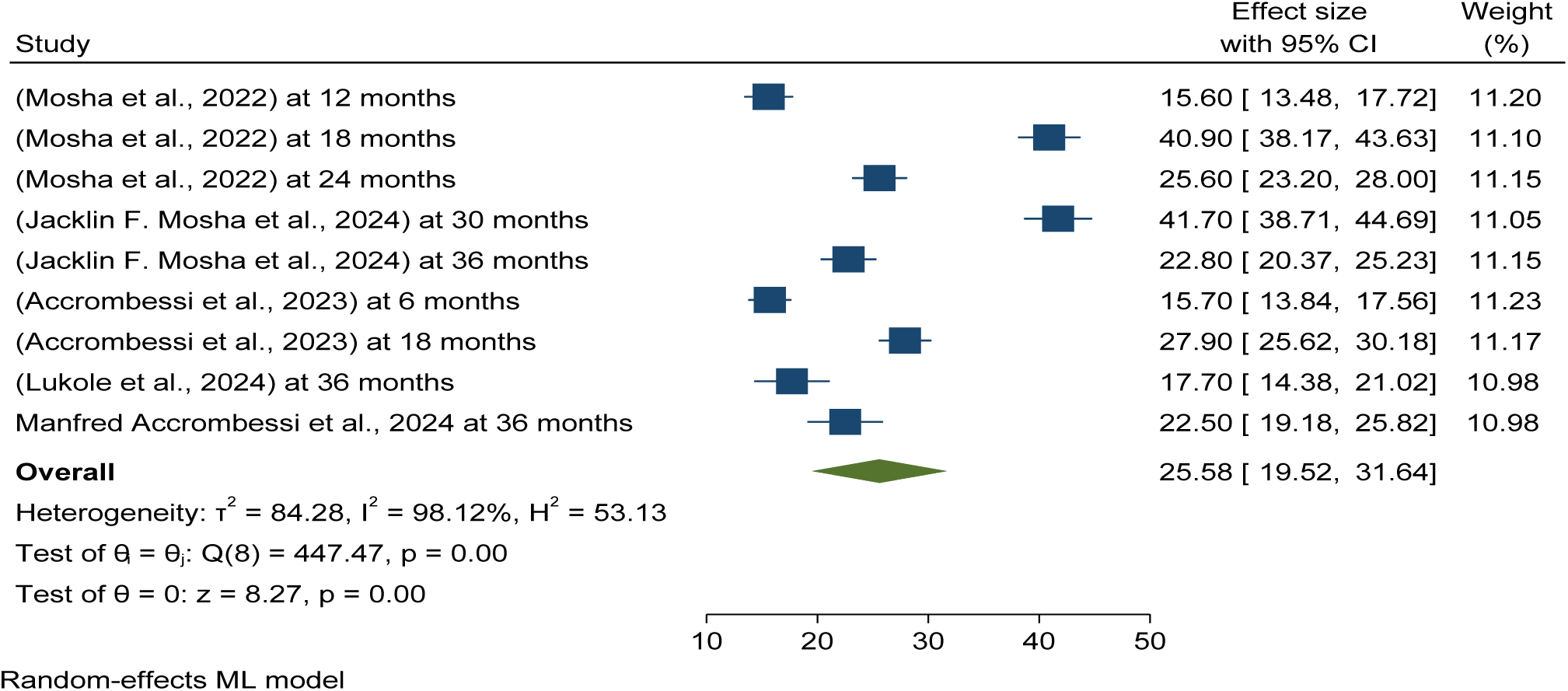
Forest plots shows *Pooled post-intervention malaria infection prevalence in selected children using chlorfenapyr long-lasting insecticidal nets (LLINs) as malaria control in Africa 2024*

Publication bias was checked using funnel plots looking at symmetrical distribution, and it was objectively verified using Egger’s regression test, which revealed that there was no publication bias (p < 0.9125) (Figure 44).

**Figure 44:**
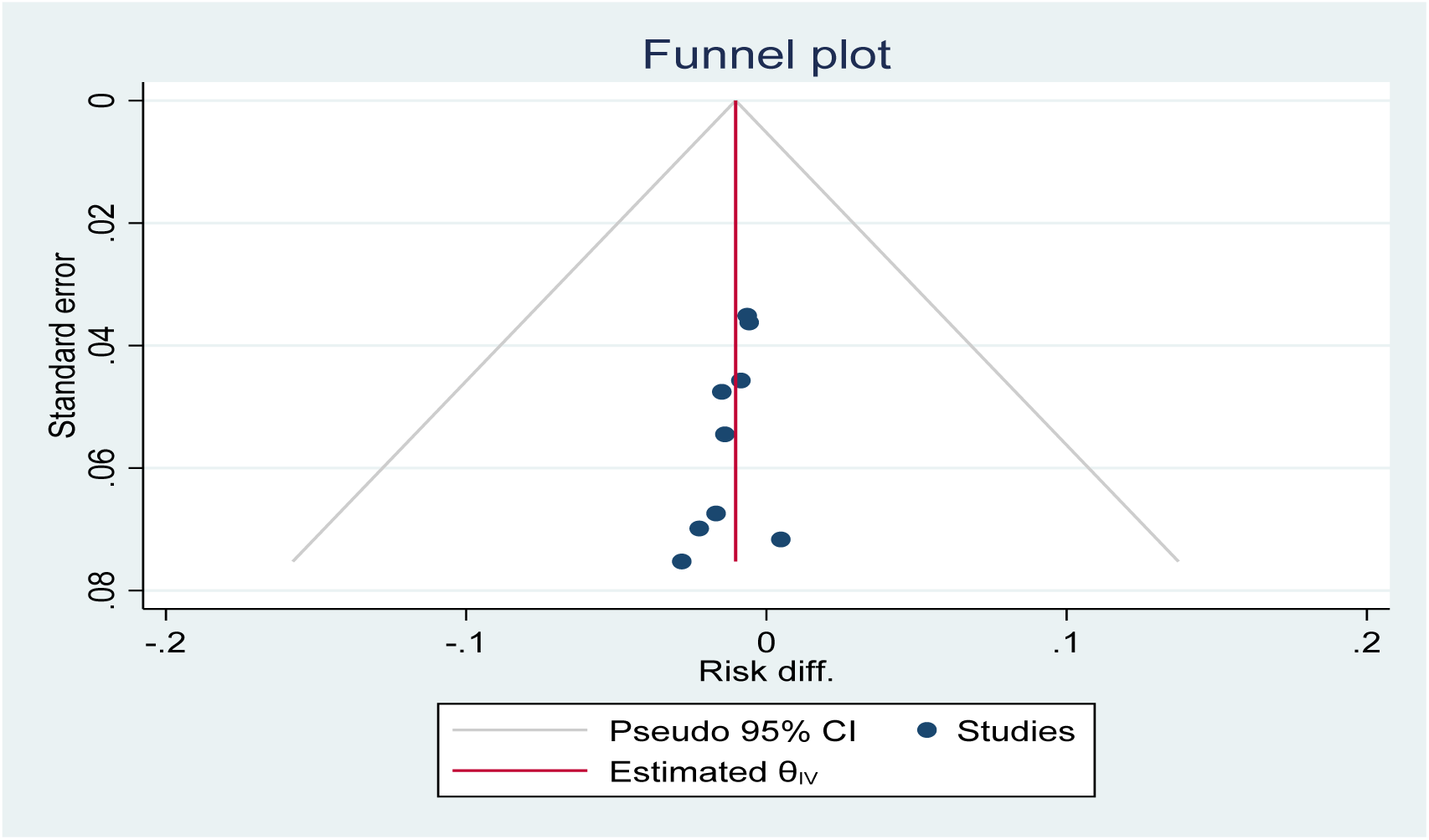
Funnel plot showing the distribution of included studies pooled malaria infection risk reduction among children using chlorfenapyr long-lasting insecticidal nets (LLINs) versus pyrethroid-only LLINs for malaria control in Africa in 2024.

### Pooled post-intervention effectiveness and efficacy of Piperonyl butoxide long-lasting insecticidal nets (LLINs) versus pyrethroid-only LLINs malaria infection reduction in Africa

This study determined that post intervention effectiveness and efficacy of piperonyl butoxide long-lasting insecticidal nets (LLINs) in malaria infection risk reduction in children over different durations (6, 12, 18, 24, and 36 months) was reduced by 1% as compared to the standard or control of the pyrethroid-only group (ARR = -0.01, 95% CI = --0.02, 0.01). See details in Figure 45.

**Figure 45:**
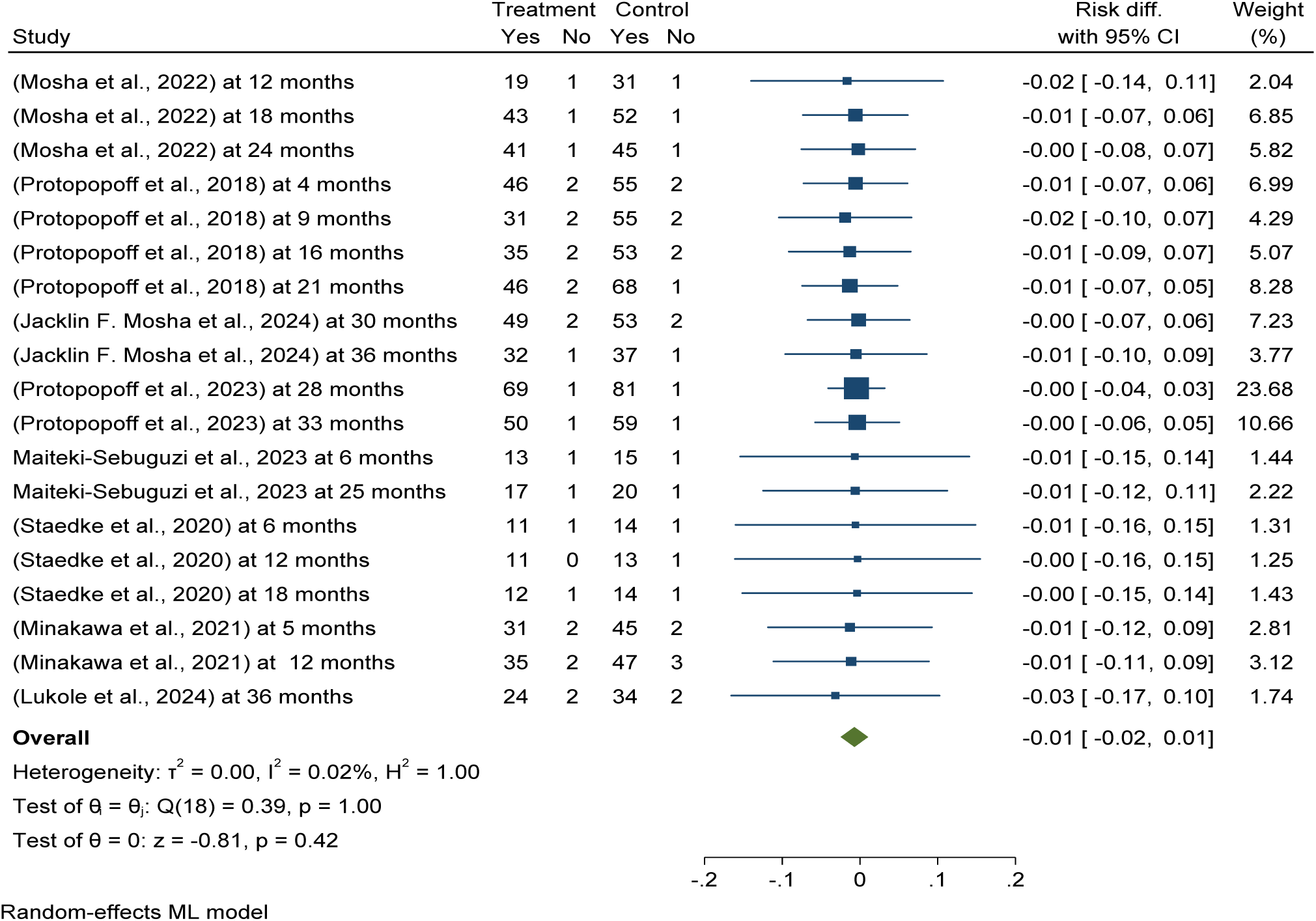
Forest plots shows *Pooled post-intervention effectiveness and efficacy of Piperonyl butoxide long-lasting insecticidal nets (LLINs) versus pyrethroid-only LLINs malaria infection reduction in Africa 2024*

The Subgroup analysis of post-intervention follow up effectiveness and efficacy of Piperonyl butoxide long-lasting insecticidal nets (LLINs) showed a significant reduction in malaria infection among children over different durations, with a 2% reduction at nine months and 1% at 36 months post-distribution of LLINs, compared to pyrethroid-only LLINs (ARR = -0.01, 95% CI = -0.02, 0.01). See details in Figure 46.

**Figure 46:**
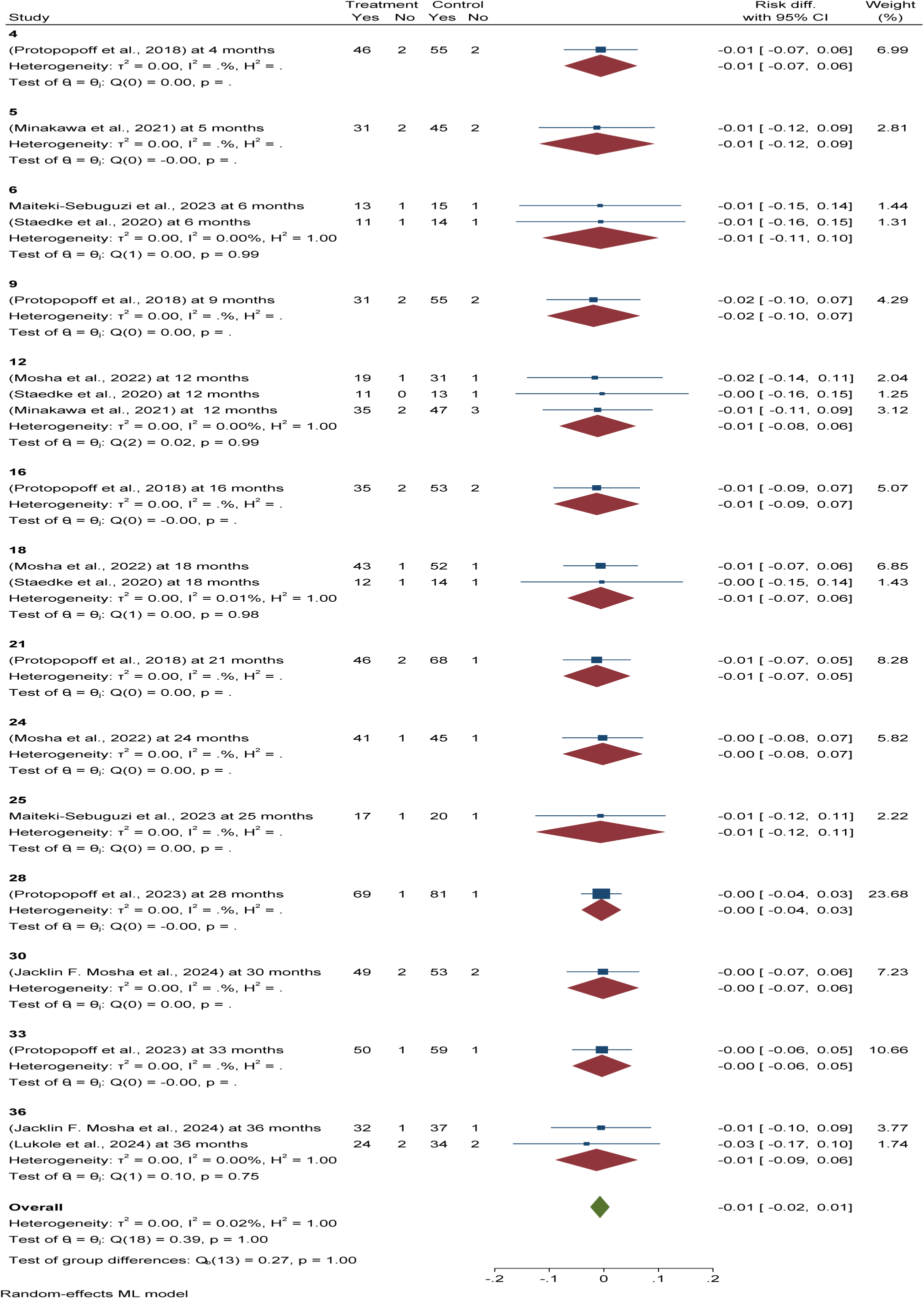
Forest plots shows s*ubgroup analysis of post-intervention follow up effectiveness and efficacy of piperonyl butoxide long-lasting insecticidal nets (LLINs) versus pyrethroid-only LLINs malaria infection reduction in Africa 2024*.

### Pooled post-intervention malaria infection prevalence in selected children using piperonyl butoxide long-lasting insecticidal nets (LLINs) in Africa

This randomized control trial meta-analysis found that the pooled prevalence of post-intervention malaria infection among children using piperonyl butoxide long-lasting insecticidal nets (LLINs) over different durations (6, 12, 18, 24, and 36 months) was 32.38 per 100 children (95% CI: 25.27–39.50%), while it was slightly higher in the control group/pyrethroid-only LLINs (40.84% per 100 children) (95% CI: 32.45%, 49.22%) in Africa (see details in Figure 47 and Figure 49).

**Figure 47:**
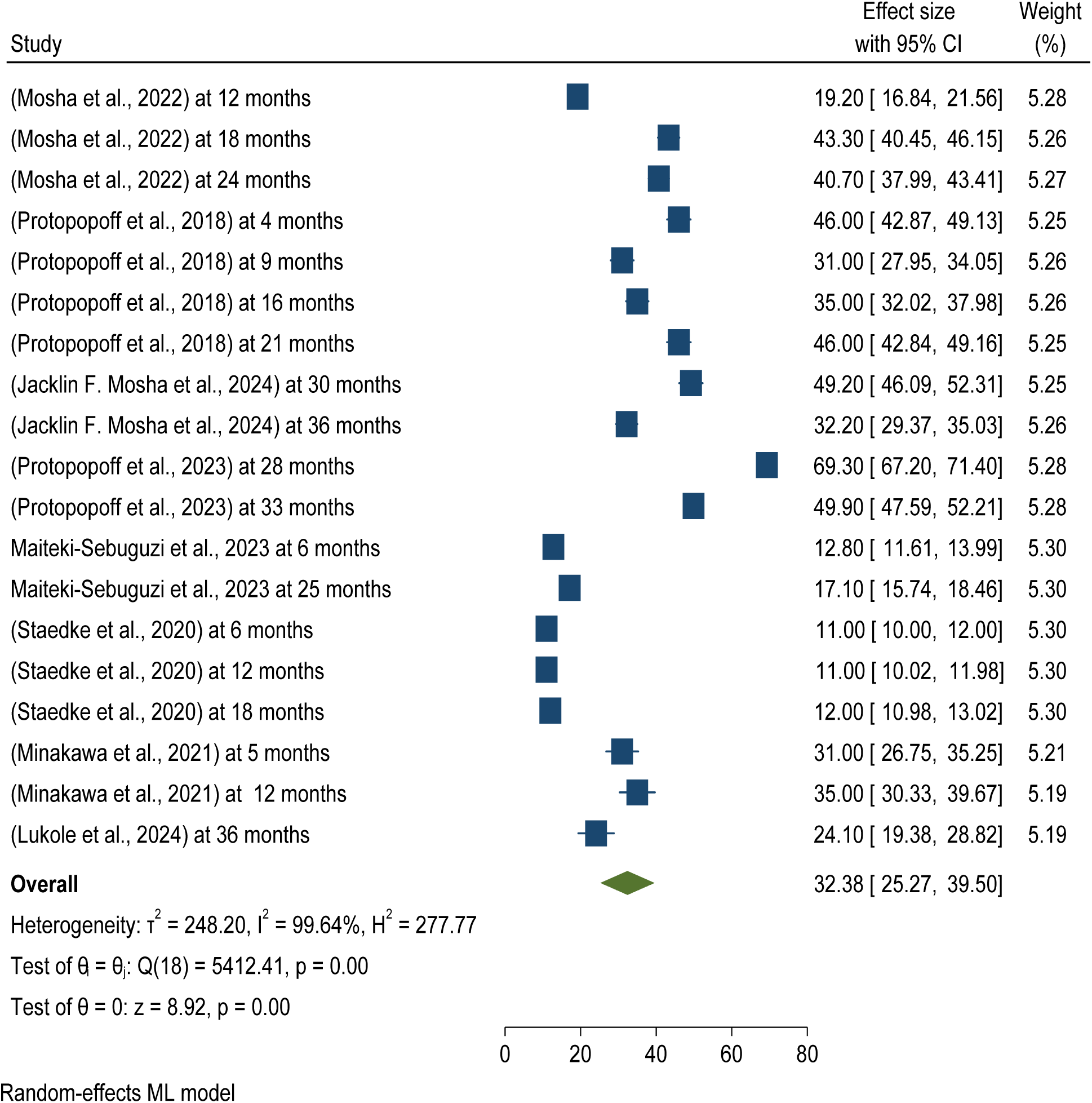
Forest plots shows Pooled post-intervention malaria infection prevalence in selected children using piperonyl butoxide long-lasting insecticidal nets (LLINs) in Africa 2024

Publication bias was checked using funnel plots looking at symmetrical distribution, and it was objectively verified using Egger’s regression test, which revealed that there was no publication bias (p < 0.847) (Figure 48).

**Figure 48:**
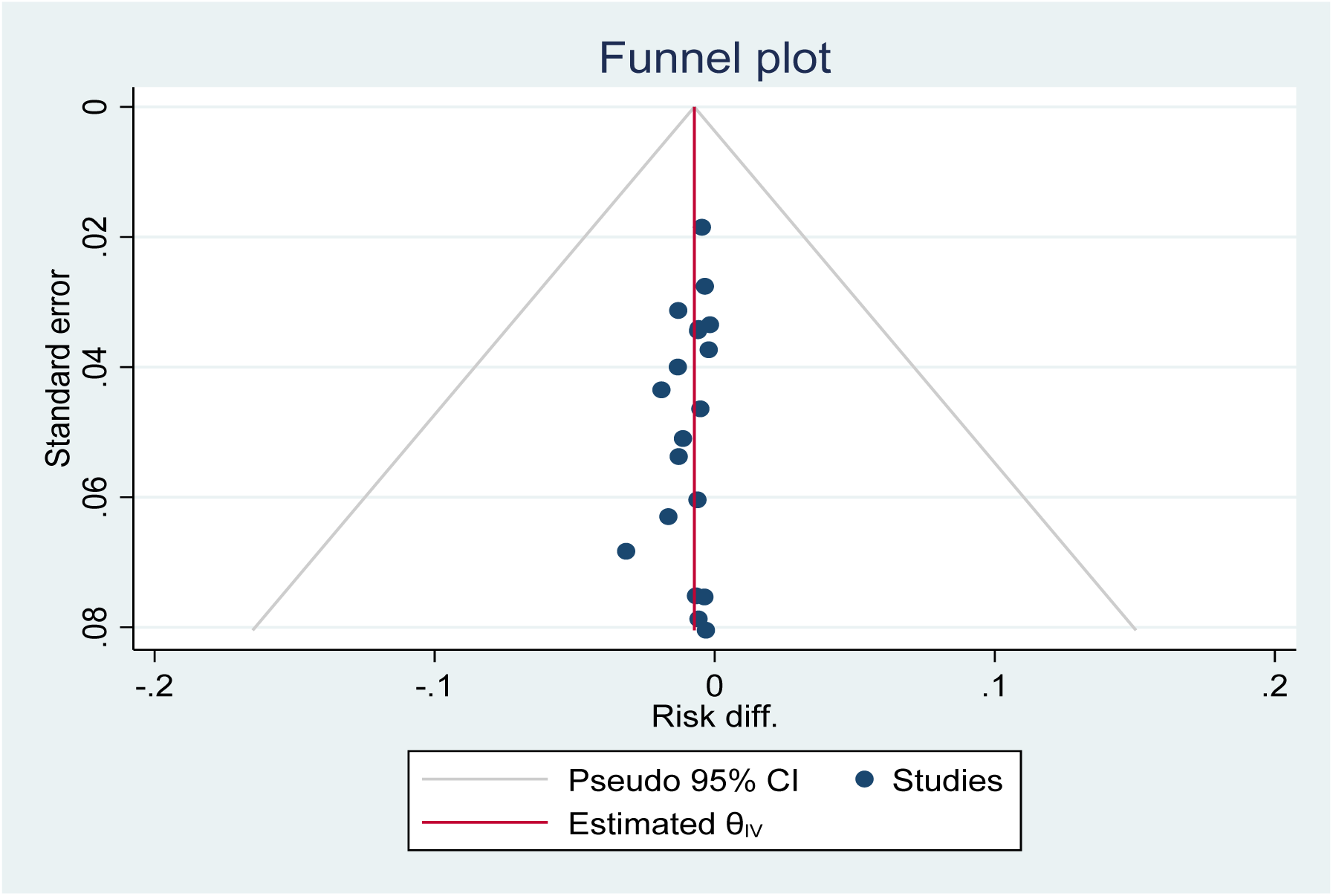
*Funnel plot showing the distribution of included studies pooled malaria infection risk reduction among children using* piperonyl butoxide *long-lasting insecticidal nets (LLINs) versus pyrethroid-only LLINs for malaria control in Africa in 2024*.

**Figure 49:**
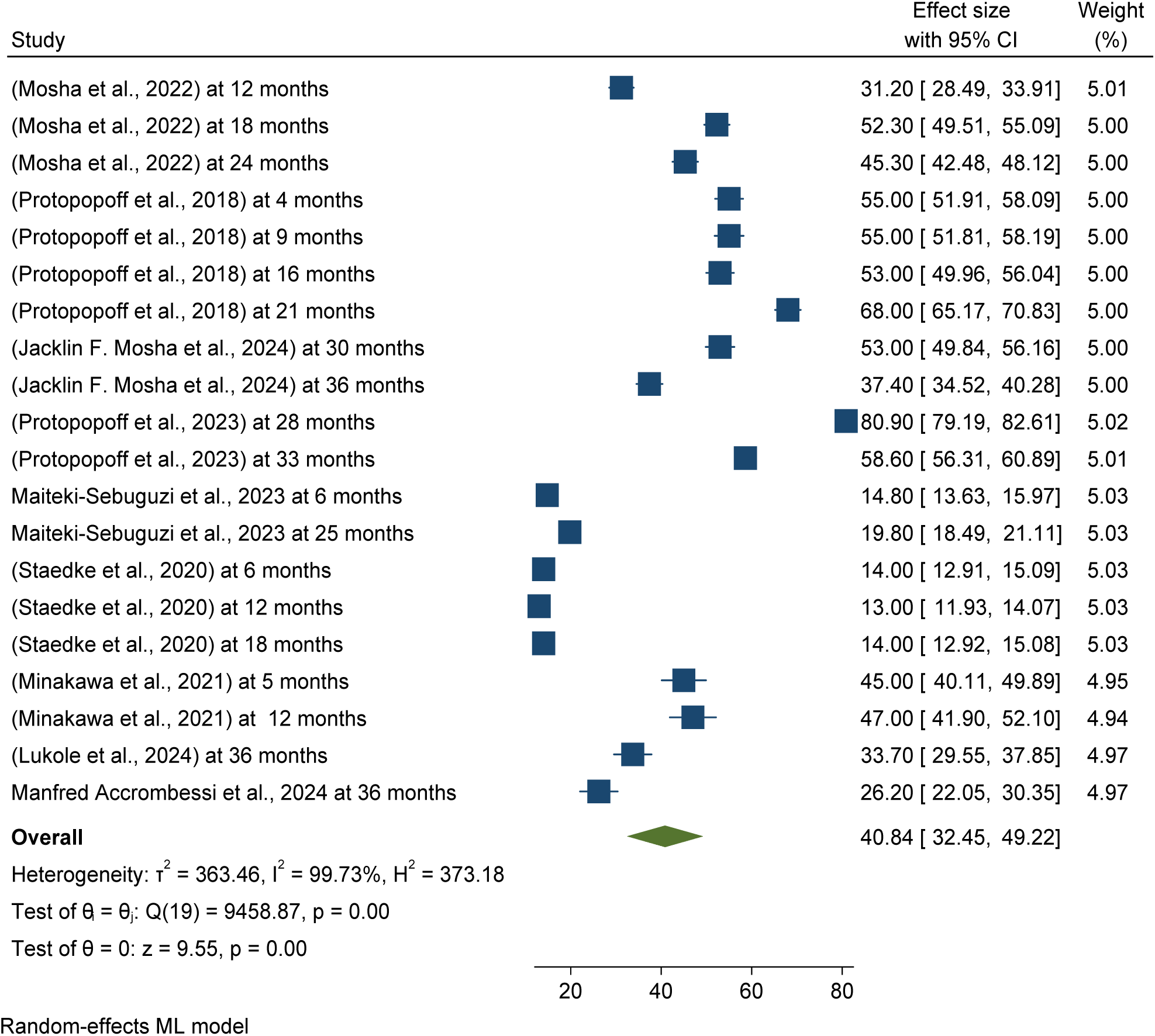
Forest plots shows *pooled post-intervention malaria infection prevalence in selected children using pyrethroid-only long-lasting insecticidal nets (LLINs) or control group in Africa 2024*

### Pooled post-intervention malaria infection prevalence in selected children using pyrethroid-only **long-lasting insecticidal nets (LLINs) or** control group **in Africa**

This randomized control trial meta-analysis found that the pooled prevalence of post-intervention malaria infection among children using pyrethroid-only LLINs or control group was 40.84% per 100 children) (95% CI: 32.45%, 49.22%) in Africa (see details in Figure 49).

Publication bias was checked using funnel plots looking at symmetrical distribution, and it was objectively verified using Egger’s regression test, which revealed that there was no publication bias (p < 0.066) (Figure 50).

**Figure 50:**
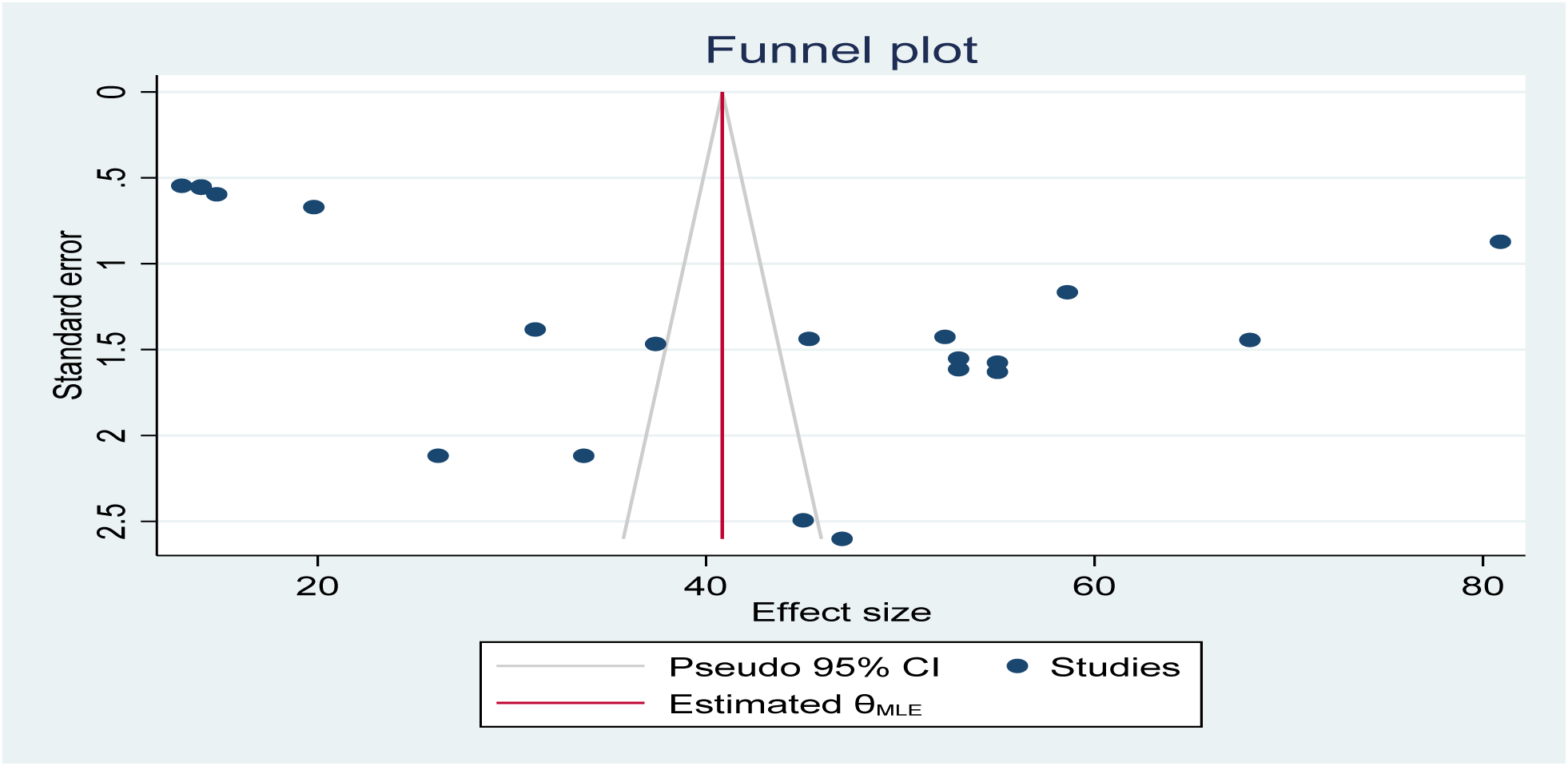
Funnel plot showing the distribution of included studies pooled malaria infection risk reduction among children using pyrethroid-only long-lasting insecticidal nets (LLINs) for malaria control in Africa in 2024

### Pooled post-intervention effectiveness and efficacy of Pyriproxyfen long-lasting insecticidal nets (LLINs) versus pyrethroid-only LLINs Malaria case incidence reduction in children (aged 6 months to 10 years in Africa

The study assessed the post-intervention effectiveness and efficacy of Pyriproxyfen long-lasting insecticidal nets (LLINs) in reducing malaria case incidence in children aged 6 months to 10 years over various durations compared to placebo/standard pyrethroid-only LLINs. Results showed no significant difference in reducing malaria case incidence (RR = -0.00). See details in Figure 51.

**Figure 51:**
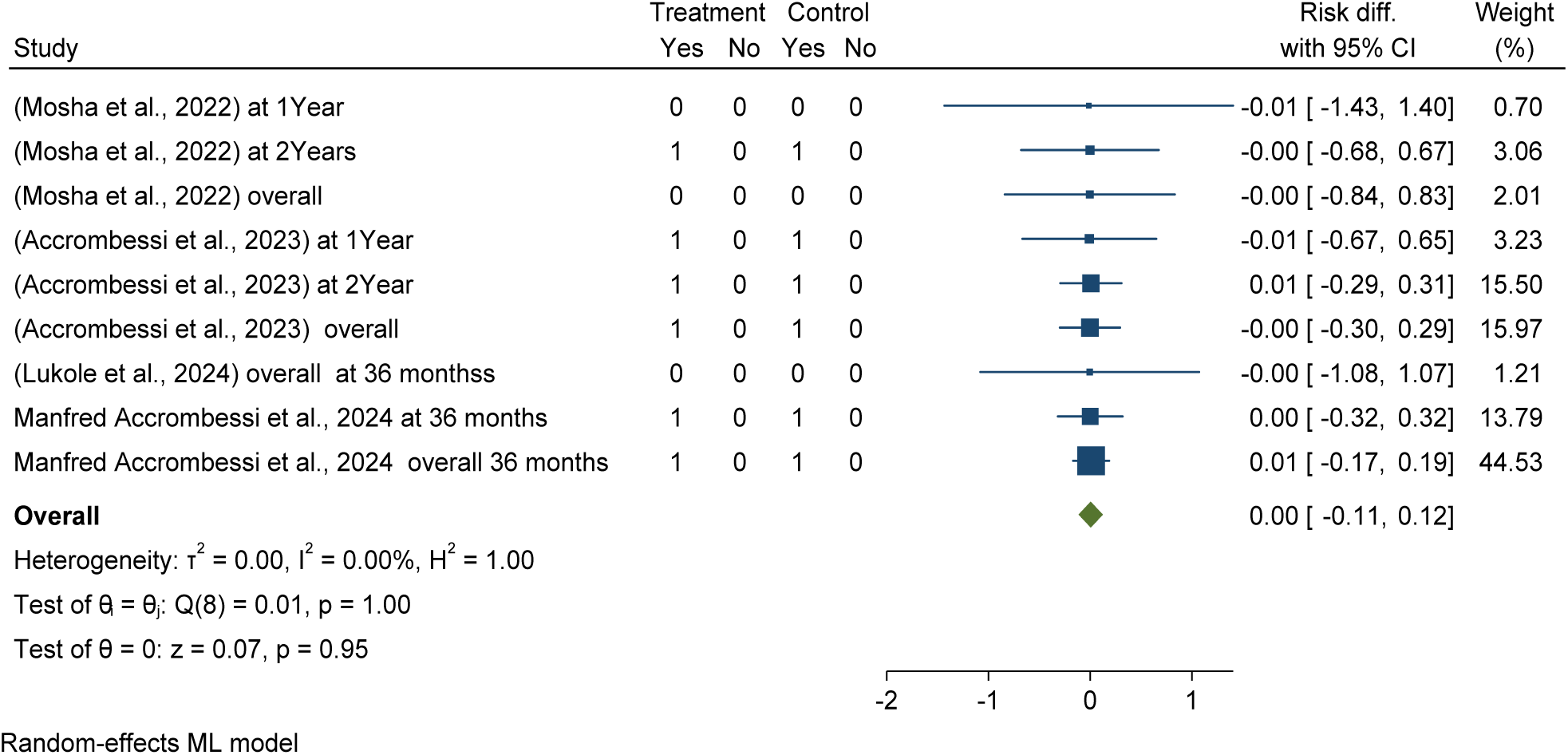
Forest plots shows Pooled post-intervention effectiveness and efficacy of Pyriproxyfen long-lasting insecticidal nets (LLINs) versus pyrethroid-only LLINs Malaria case incidence reduction in children (aged 6 months to 10 years in Africa

Publication bias was checked using funnel plots looking at symmetrical distribution, and it was objectively verified using Egger’s regression test, which revealed that there was no publication bias (p < 0.066) (Figure 52).

**Figure 52:**
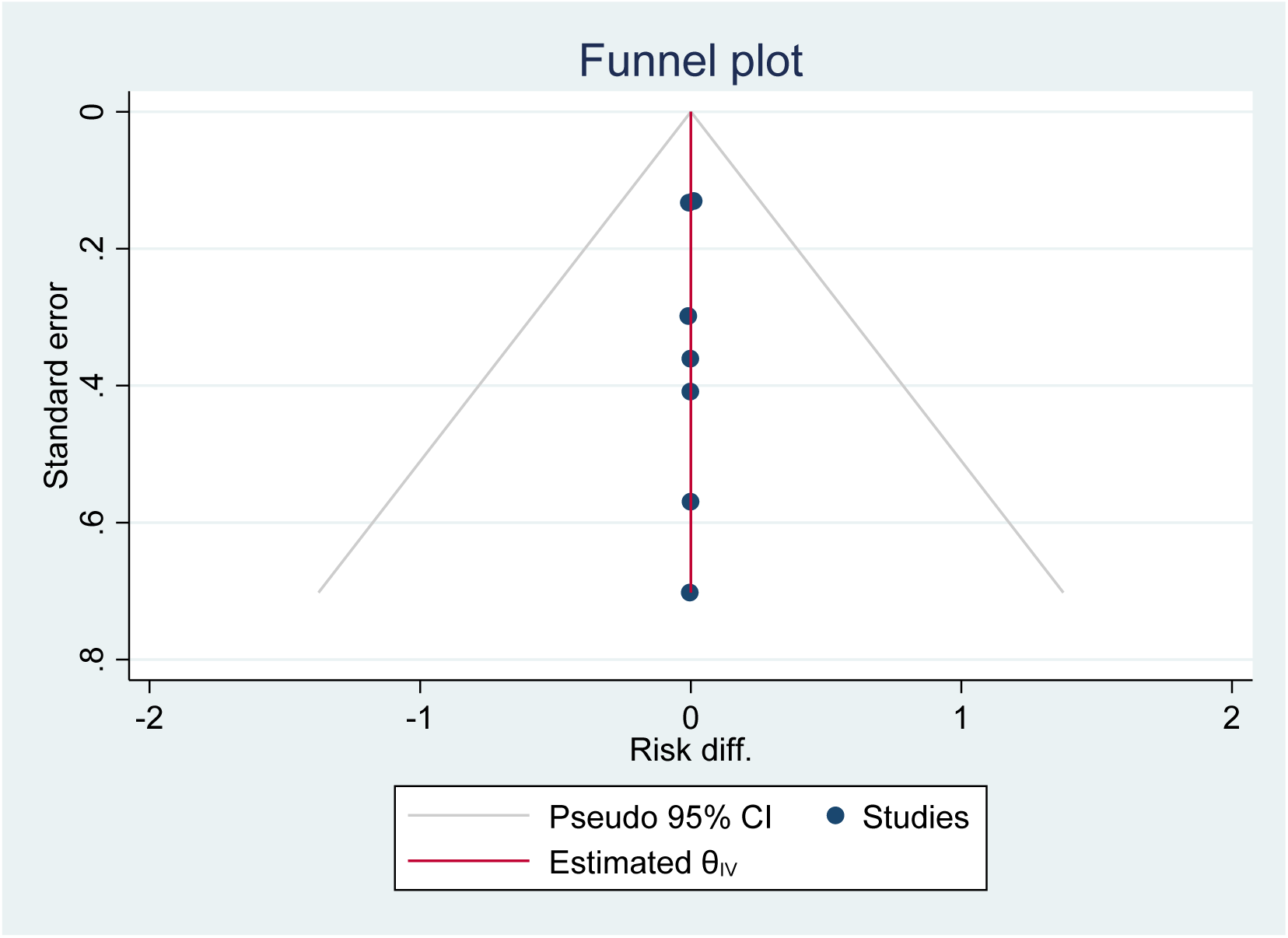
Funnel plot showing the distribution of included studies pooled effectiveness and efficacy of Pyriproxyfen long-lasting insecticidal nets (LLINs) versus pyrethroid-only LLINs Malaria case incidence reduction in children (aged 6 months to 10 years in Africa in 2024

### Pooled post-intervention Malaria case incidence reduction in children (aged 6 months to 10 years using Pyriproxyfen long-lasting insecticidal nets (LLINs) versus pyrethroid-only LLINs in Africa

This randomized control trial meta-analysis found that the pooled post-intervention malaria case incidence among children (aged 6 months to 10 years using Pyriproxyfen long-lasting insecticidal nets (LLINs over different durations (12, 24, and 36 months) was 69 per 100 children years (95% CI: 0.46, 0.89), and in the control group/ pyrethroid-only LLINs was 46 per 100 children years ) (95% CI: 0.28 , 0.63) in Africa (see details in Figure 53 and Figure 57).

**Figure 53:**
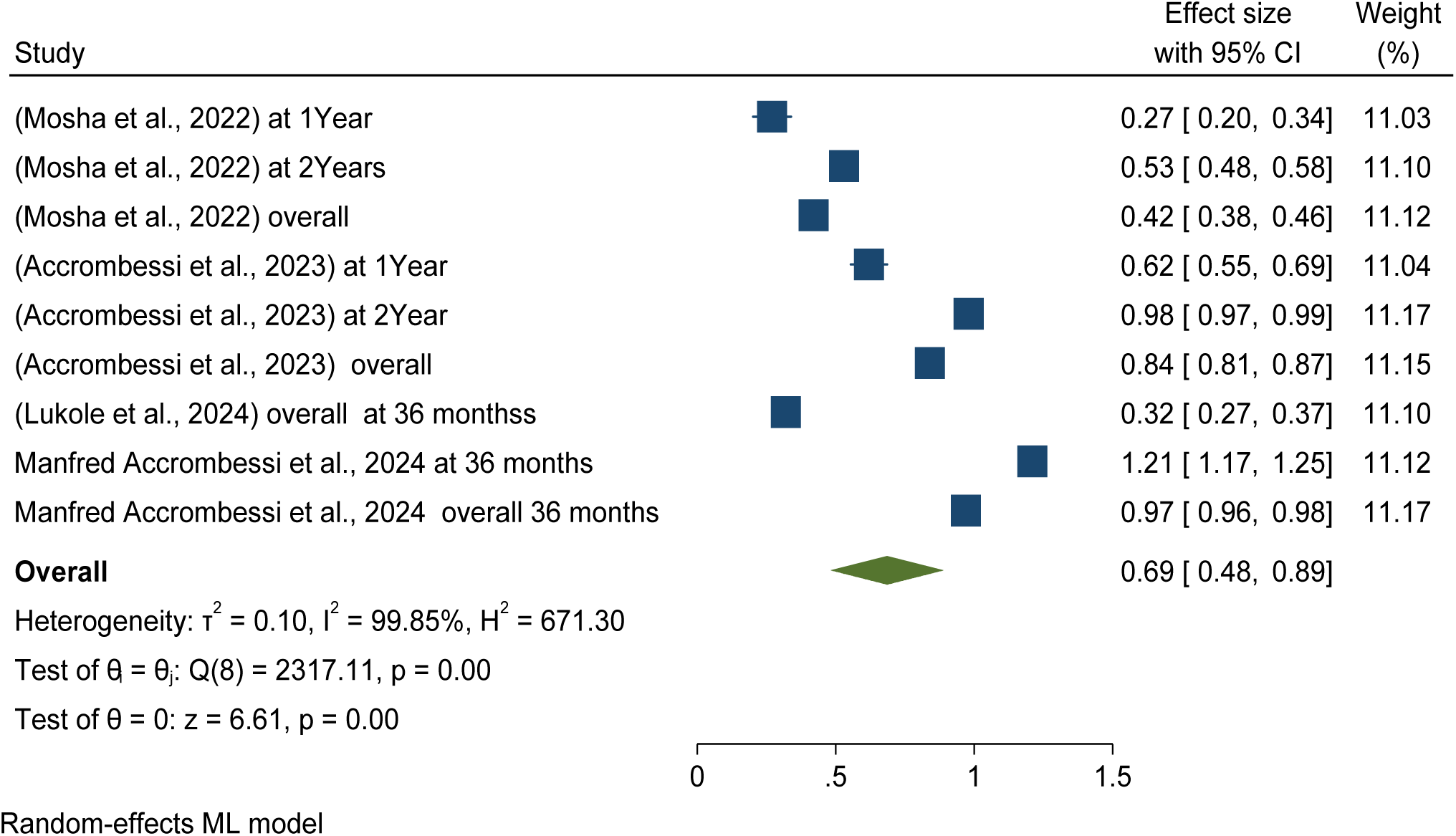
*Forest* plots shows *pooled post-intervention Malaria case incidence reduction in children (aged 6 months to 10 years using Pyriproxyfen long-lasting insecticidal nets (LLINs) versus pyrethroid-only LLINs in Africa 2024*

### Pooled post-intervention effectiveness and efficacy of chlorfenapyr long-lasting insecticidal nets (LLINs) versus pyrethroid-only LLINs malaria case incidence reduction in children aged 6 months to 10 years in Africa

This study evaluated the post-intervention effectiveness and efficacy of chlorfenapyr long-lasting insecticidal nets (LLINs) in malaria case incidence reduction in children aged 6 months to 10 years over different durations, one year to 2 years, and overall reduction by 4% as compared to the standard or control of the pyrethroid-only group (ARR = -0.04, 95% CI = - 0.33, 0.26). See details in Figure 54.

**Figure 54:**
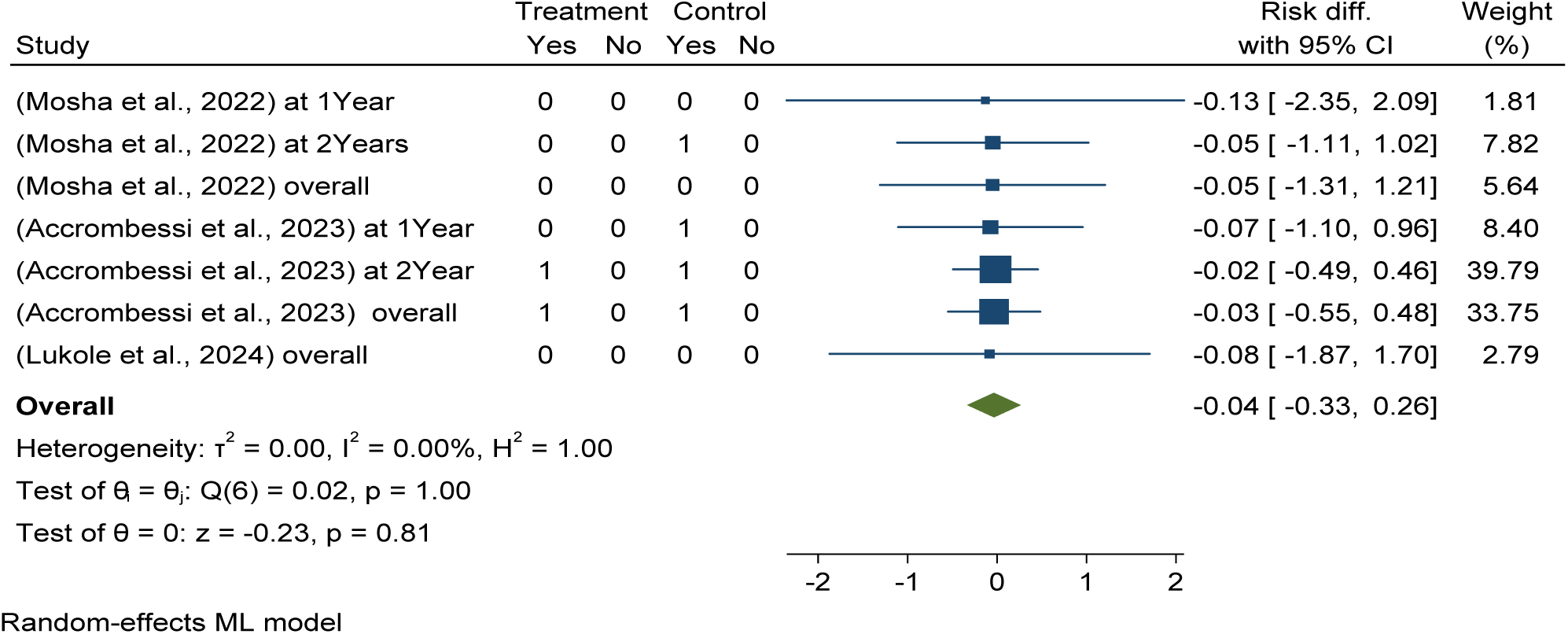
*Forest* plots shows *pooled post-intervention effectiveness and efficacy of chlorfenapyr long-lasting insecticidal nets (LLINs) versus pyrethroid-only LLINs malaria case incidence reduction in children aged 6 months to 10 years in Africa 2024*

This randomized control trial meta-analysis found that the pooled post-intervention malaria case incidence among children (aged 6 months to 10 years using chlorfenapyr long-lasting insecticidal nets (LLINs) and control group/pyrethroid-only LLINs over different durations (12, 24, and 36 months) was the same 46 per 100 children years (95% CI: 0.28, 0.63) in Africa (see details in Figure 55 and Figure 58).

**Figure 55:**
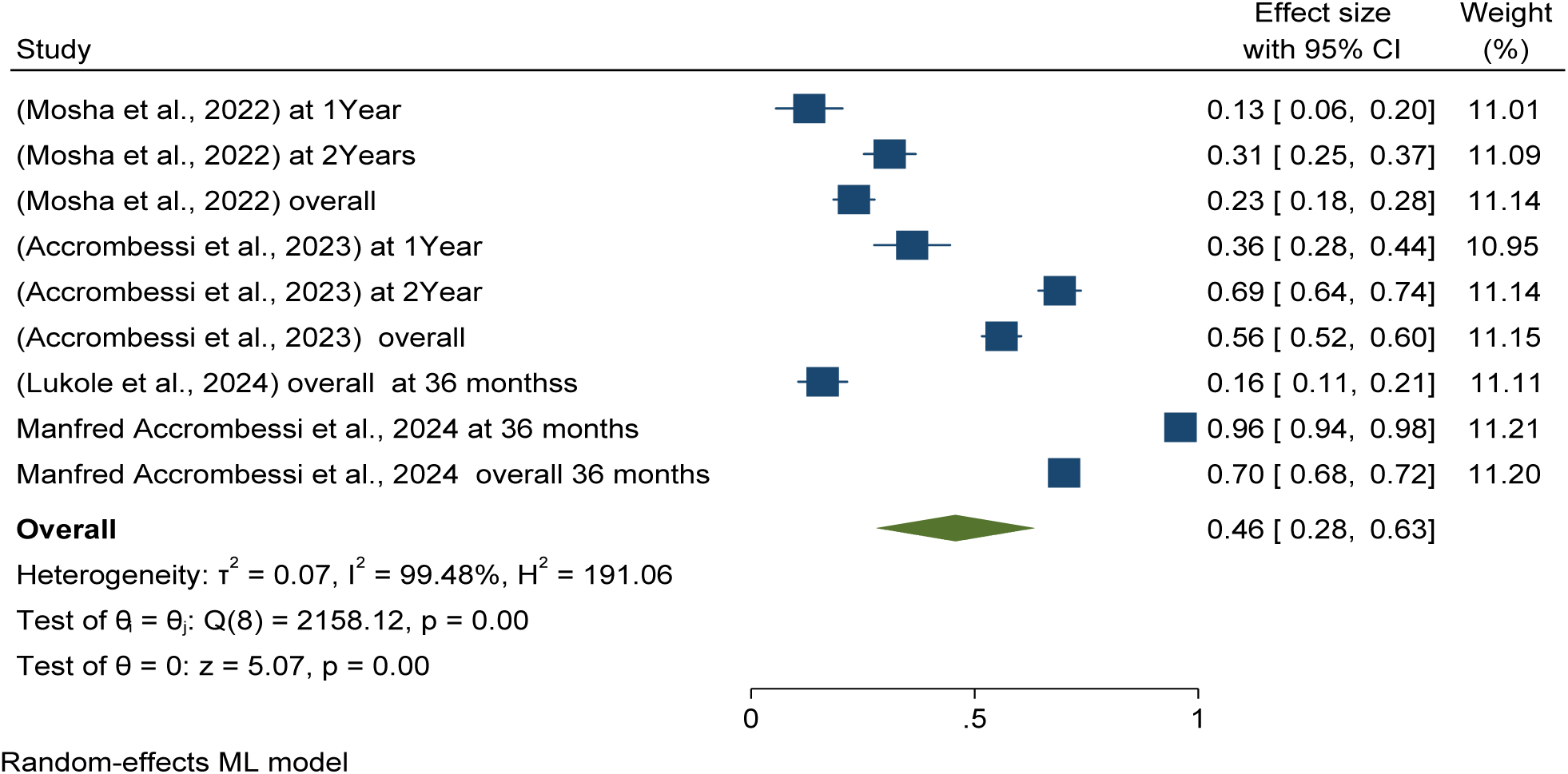
*Forest* plots shows *pooled post-intervention Malaria case incidence reduction in children (aged 6 months to 10 years using chlorfenapyr long-lasting insecticidal nets (LLINs) for malaria control in Africa 2024*

**Figure 56:**
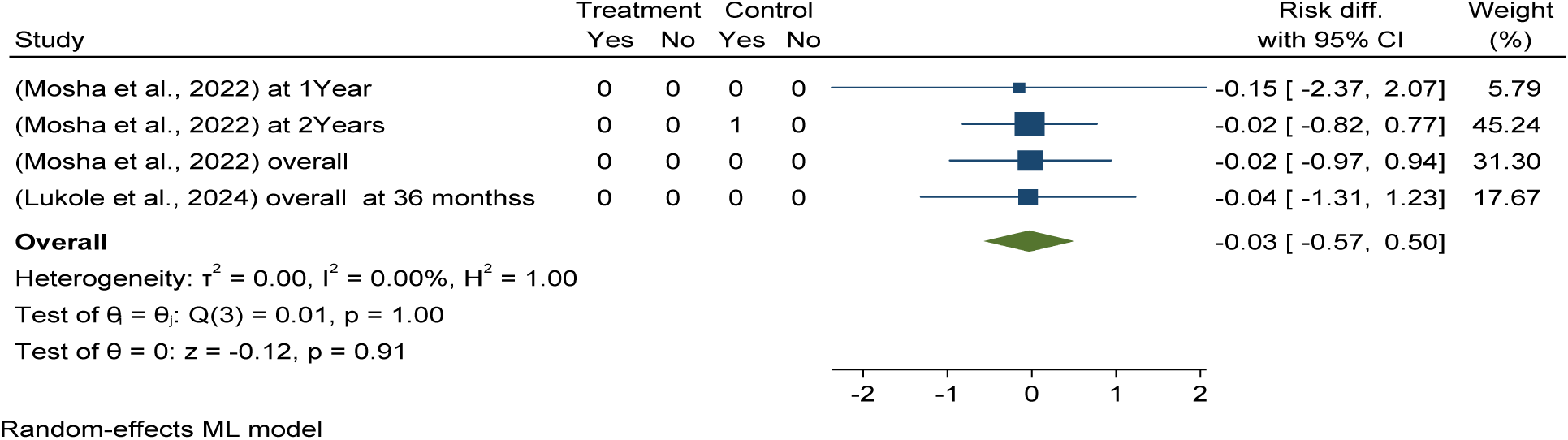
Forest *plots shows p*ooled post-intervention effectiveness and efficacy of Piperonyl butoxide long-lasting insecticidal nets (LLINs) versus pyrethroid-only LLINs malaria case incidence reduction in children aged 6 months to 10 years in Africa 2024

**Figure 57:**
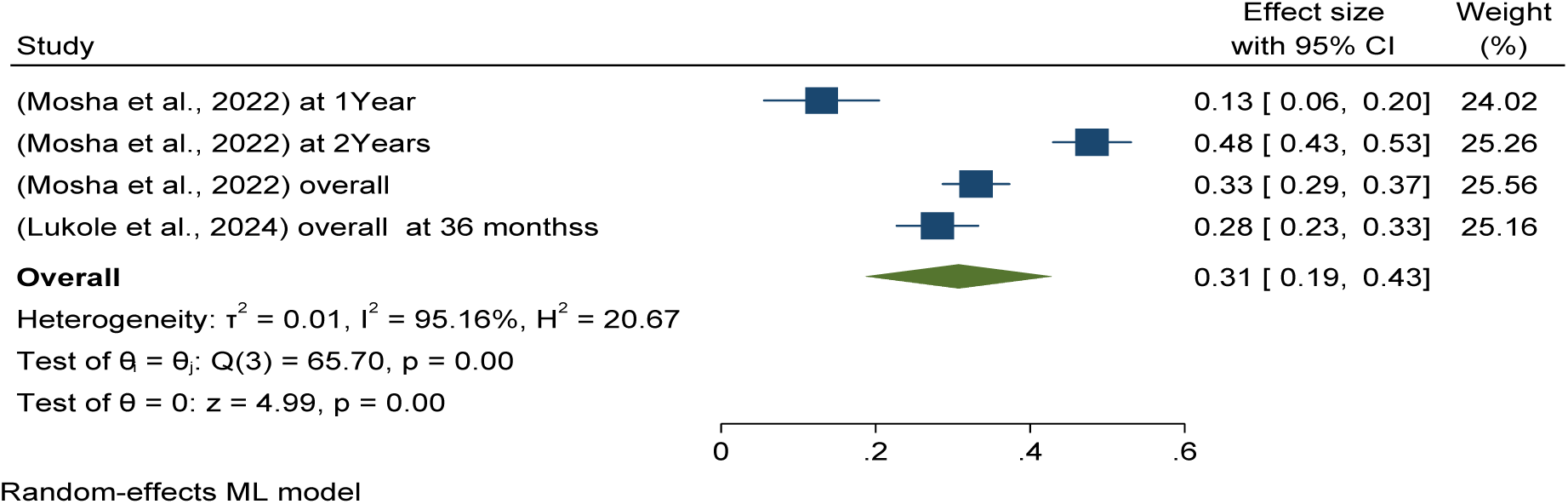
*Forest* plots shows *pooled post-intervention Malaria case incidence reduction in children (aged 6 months to 10 years using piperonyl butoxide long-lasting insecticidal nets (LLINs) for malaria control in Africa 2024*

**Figure 58:**
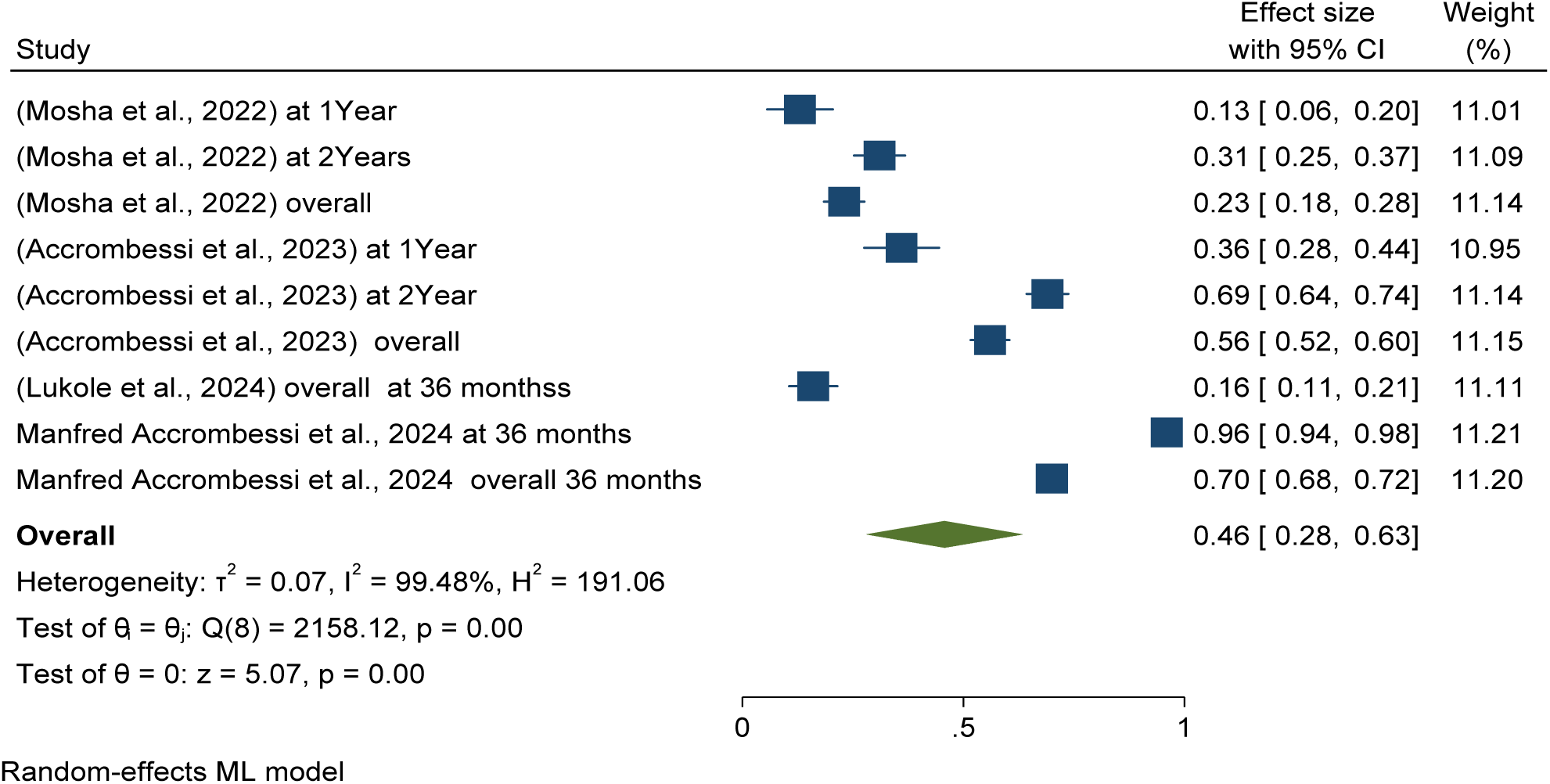
*Forest* plots shows *pooled post-intervention Malaria case incidence reduction in children (aged 6 months to 10 years using pyrethroid-only long-lasting insecticidal nets (LLINs) for malaria control in Africa 2024*

### Pooled post-intervention effectiveness and efficacy of Piperonyl butoxide long-lasting insecticidal nets (LLINs) versus pyrethroid-only LLINs malaria case incidence reduction in children aged 6 months to 10 years in Africa

This study determined that post intervention effectiveness and efficacy of piperonyl butoxide long-lasting insecticidal nets (LLINs) in malaria case incidence reduction in children aged 6 months to 10 years over different durations, one year to 2 years, and overall reduced by 3% as compared to the standard or control of the pyrethroid-only group (ARR = -0.03, 95% CI = - 0.57, 0.5). See details in Figure 56.

This randomized control trial meta-analysis found that the pooled post-intervention malaria case incidence among children (aged 6 months to 10 years using piperonyl butoxide long-lasting insecticidal nets (LLINs) over different durations (12, 24, and 36 months) was 31per 100 children years (95% CI: 0.19, 0.43), and in the control group/ pyrethroid-only LLINs was 46 per 100 children years ) (95% CI: 0.28 , 0.63) in Africa (see details in Figure 57 and Figure 58).

This randomized control trial meta-analysis found that the pooled prevalence of post-intervention malaria case incidence among children (aged 6 months to 10 years using pyrethroid-only LLINs or control standard care over different durations (12, 24, and 36 months) was 46 per 100 children years (95% CI: 0.28 , 0.63) in Africa (see details in Figure 37).

### Post-intervention: pooled mean indoor vector density per household per night

This meta-analysis determined entomological outcomes effectiveness and efficacy in terms of mean indoor vector density per household per night reduction by using different long-lasting insecticidal nets (LLINs) in Africa.

The forest plot demonstrates that pyriproxyfen long-lasting insecticidal nets significantly reduce pooled mean indoor vector density in Africa by 1% as compared to standard care/pyrethroid-only LLINs treatment in Africa (ARR = 0.01, 95% CI = -0.05, 0.08), with the included studies were homogeneous (I2 = 0.00%). See details in Figure 59.

**Figure 59:**
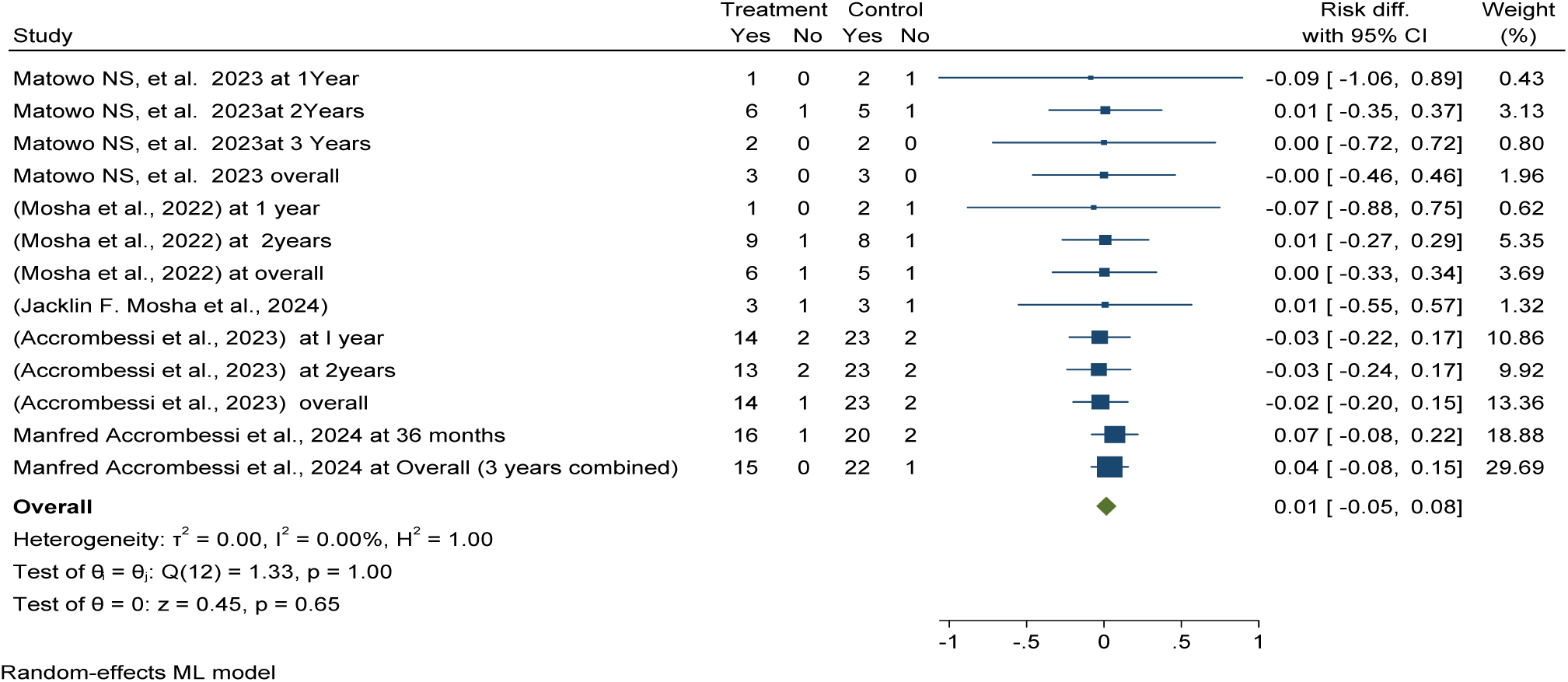
Forest plot shows the effectiveness and efficacy of pyriproxyfen long-lasting insecticidal nets (LLINs) in pooled mean indoor vector density reduction compared to pyrethroid-only LLINs in Africa in 2024.

### Post-intervention pooled mean indoor vector density per household per nights among pyriproxyfen long-lasting insecticidal nets intervention

The study revealed that the pooled mean indoor vector density per household per night in the pyriproxyfen LLIN intervention was 7.74, which slightly lower than pyrethroid-only LLINs or control groups, with a mean of 8.04 per household per night as shown in Figures 60 and 68.

**Figure 60:**
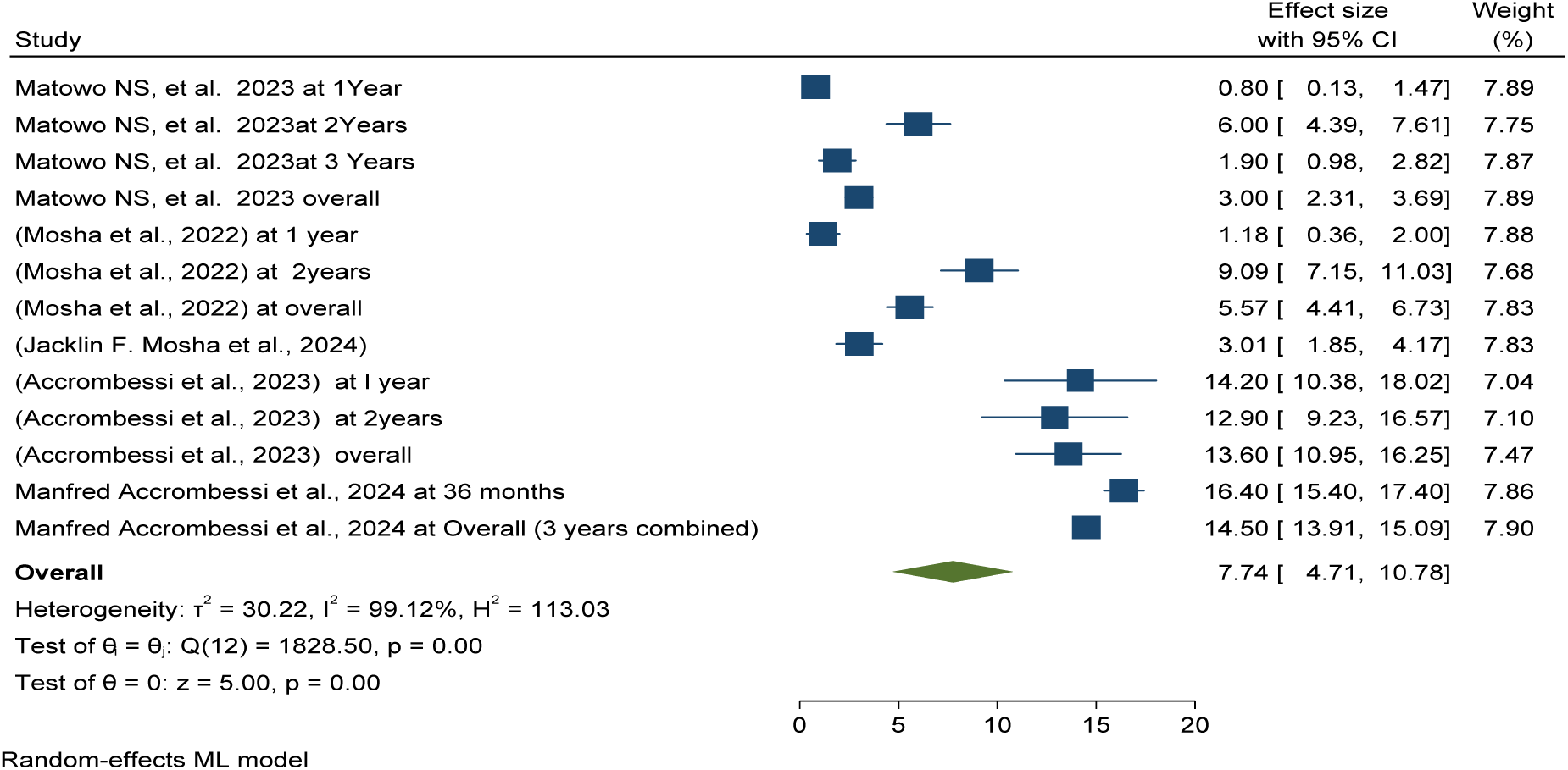
Forest plot shows post-intervention pooled mean indoor vector density per household per nights among pyriproxyfen long-lasting insecticidal nets intervention in Africa 2024

Publication bias was checked using funnel plots looking at symmetrical distribution, and it was objectively verified using Egger’s regression test, which revealed that there was no publication bias (p < 0.847) (Figure 61).

**Figure 61:**
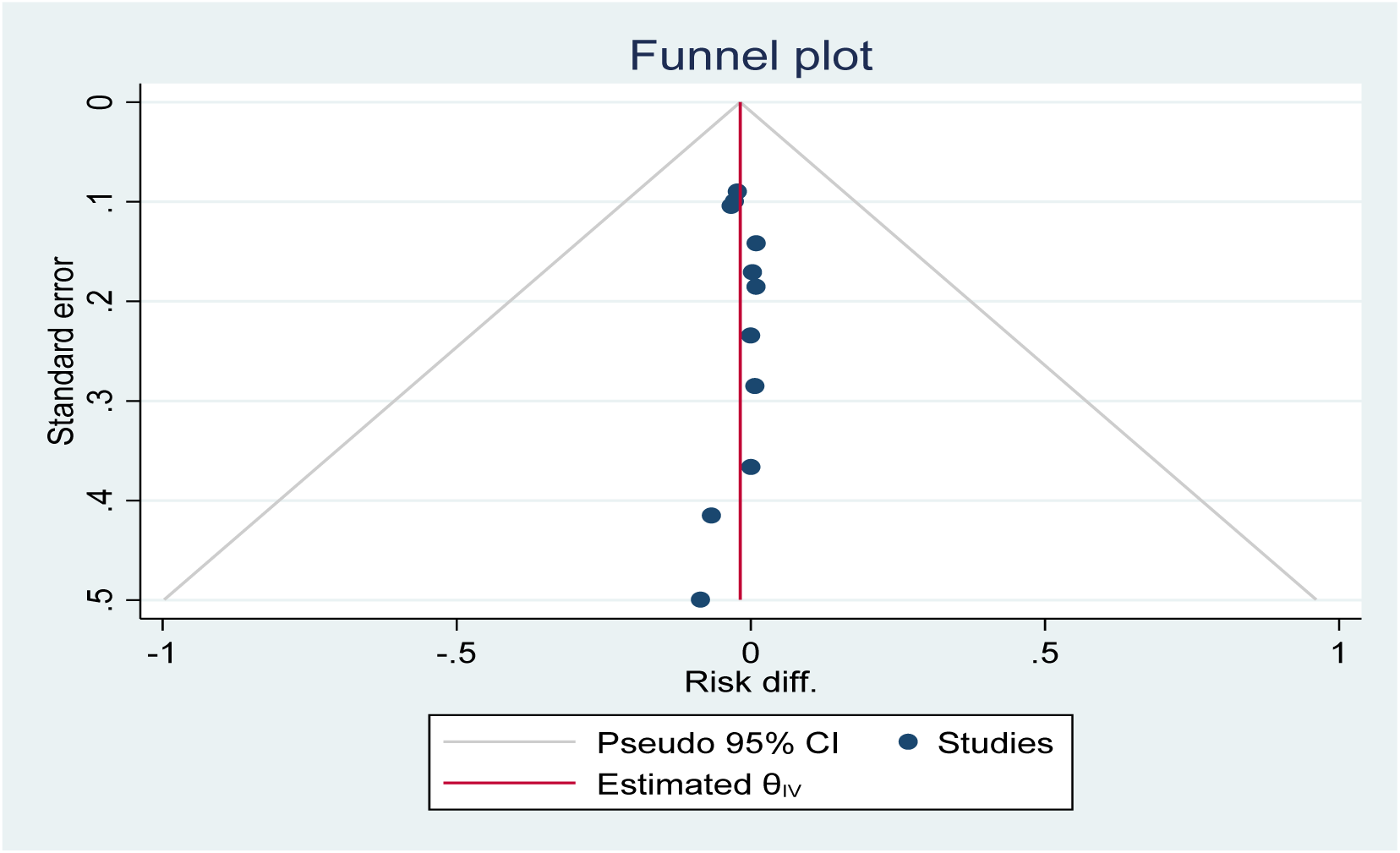
*Funnel plot showing the distribution of included studies post-intervention pooled mean indoor vector density per household per nights among pyriproxyfen long-lasting insecticidal nets intervention in Africa 2024*

This meta-analysis found that chlorfenapyr long-lasting insecticidal nets are effectively and efficiently reduced mean vector density per household per night by 4% compared to pyrethroid-only LLNs, demonstrating their effectiveness and efficacy over varying durations from one to three years post-distribution in Africa (Figure 62).

**Figure 62:**
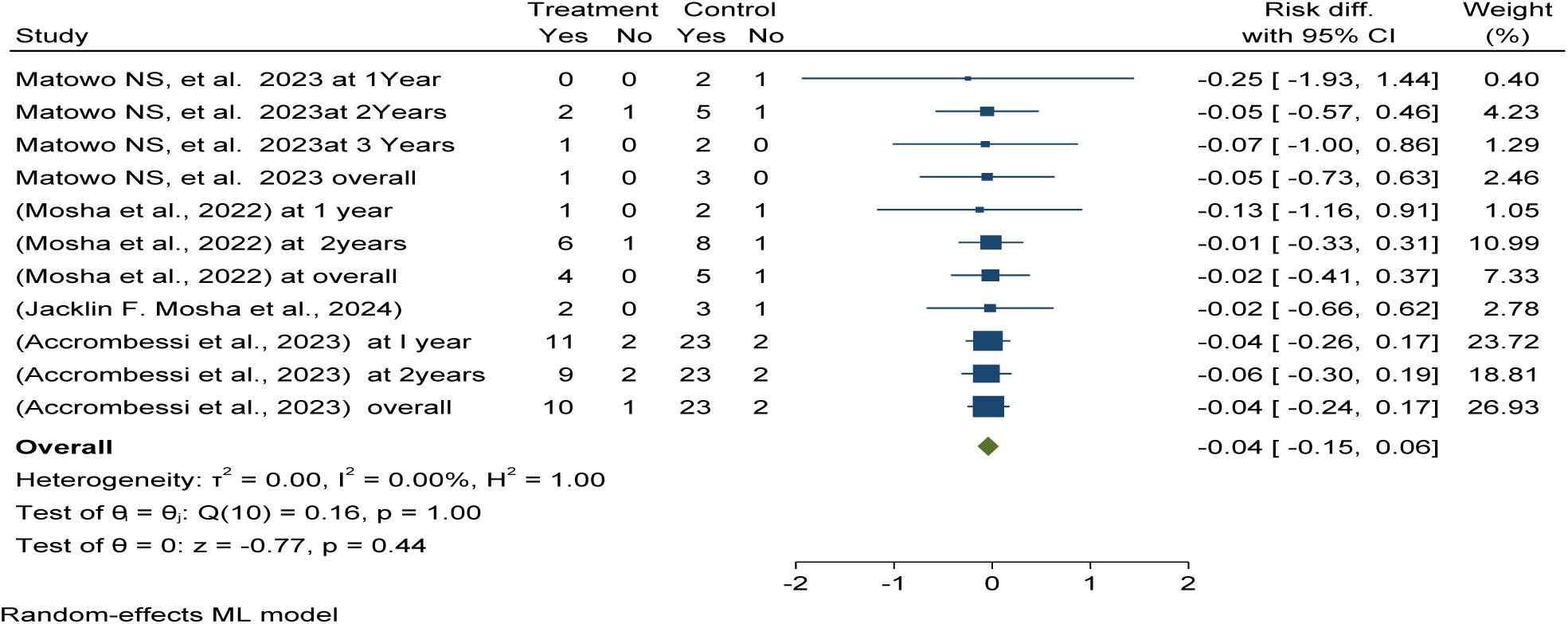
Forest plot shows the effectiveness and efficacy of chlorfenapyr long-lasting insecticidal nets (LLINs) in pooled mean indoor vector density reduction compared to pyrethroid-only LLINs in Africa in 2024.

### Post-intervention pooled mean indoor vector density per household per nights among chlorfenapyr long-lasting insecticidal nets

The study revealed that the pooled mean indoor vector density per household per night in the chlorfenapyr long-lasting insecticidal nets intervention was 5.53, which significantly lower than pyrethroid-only LLINs or control groups, with a mean of 8.04 per household per night as shown in Figures 63 and 68.

**Figure 63:**
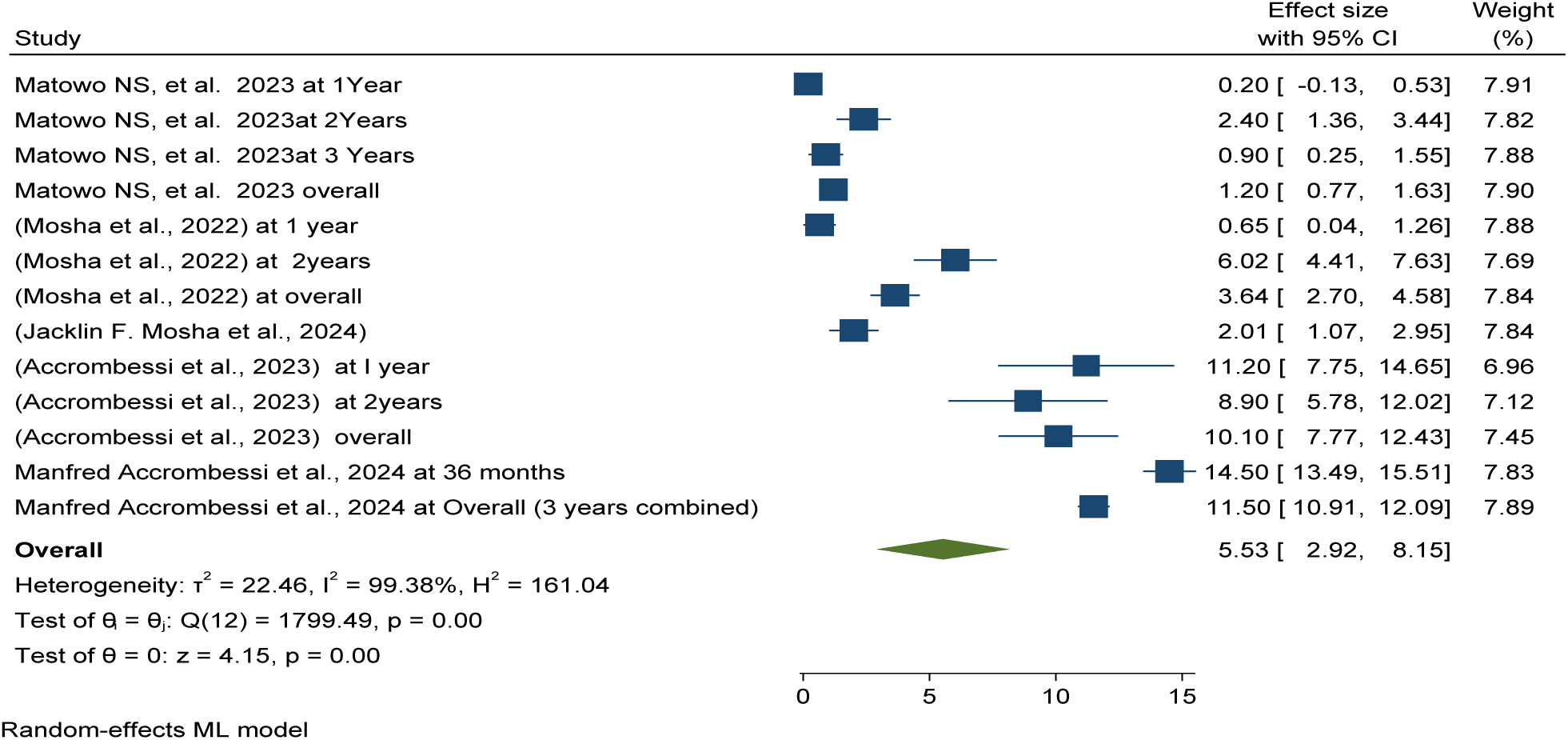
*Forest plot shows post-intervention pooled mean indoor vector density per household per nights among* chlorfenapy *long-lasting insecticidal nets intervention in Africa 2024*

Publication bias was checked using funnel plots looking at symmetrical distribution, and it was objectively verified using Egger’s regression test, which revealed that there was no publication bias (p < 0.98) (Figure 64).

**Figure 64:**
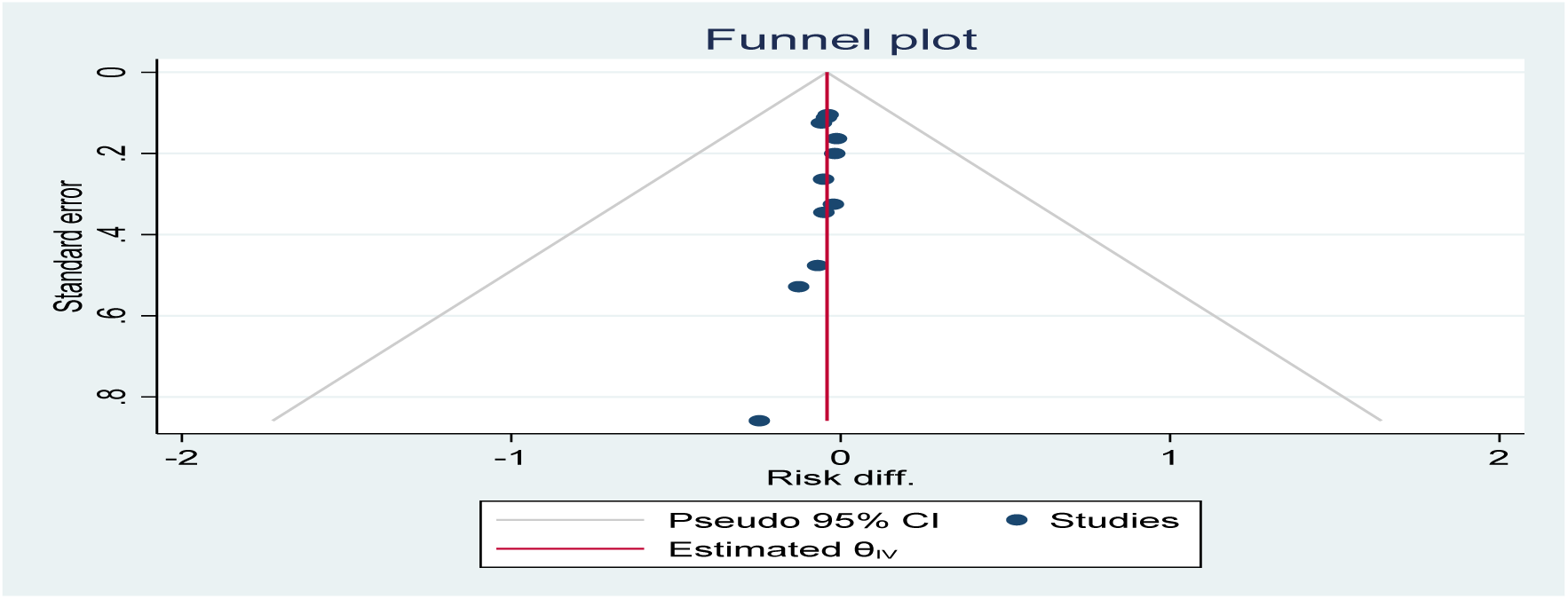
*Funnel plot showing the distribution of included studies post-intervention pooled mean indoor vector density per household per nights among* chlorfenapy *long-lasting insecticidal nets intervention in Africa 2024*

This meta-analysis determined entomological outcomes of Piperonyl butoxide long-lasting insecticidal nets are effectively and efficiently reduced mean vector density per household per night by 3% compared to pyrethroid-only LLNs, which demonstrating their effectiveness and efficacy over varying durations from one to three years post-distribution in Africa

The study revealed that the pooled mean indoor vector density per household per night in the Piperonyl butoxide long-lasting insecticidal nets intervention group was 1.9, which significantly lower than pyrethroid-only LLINs or control groups, with a mean of 8.04 per household per night as shown in Figures 65 and 68.

**Figure 65:**
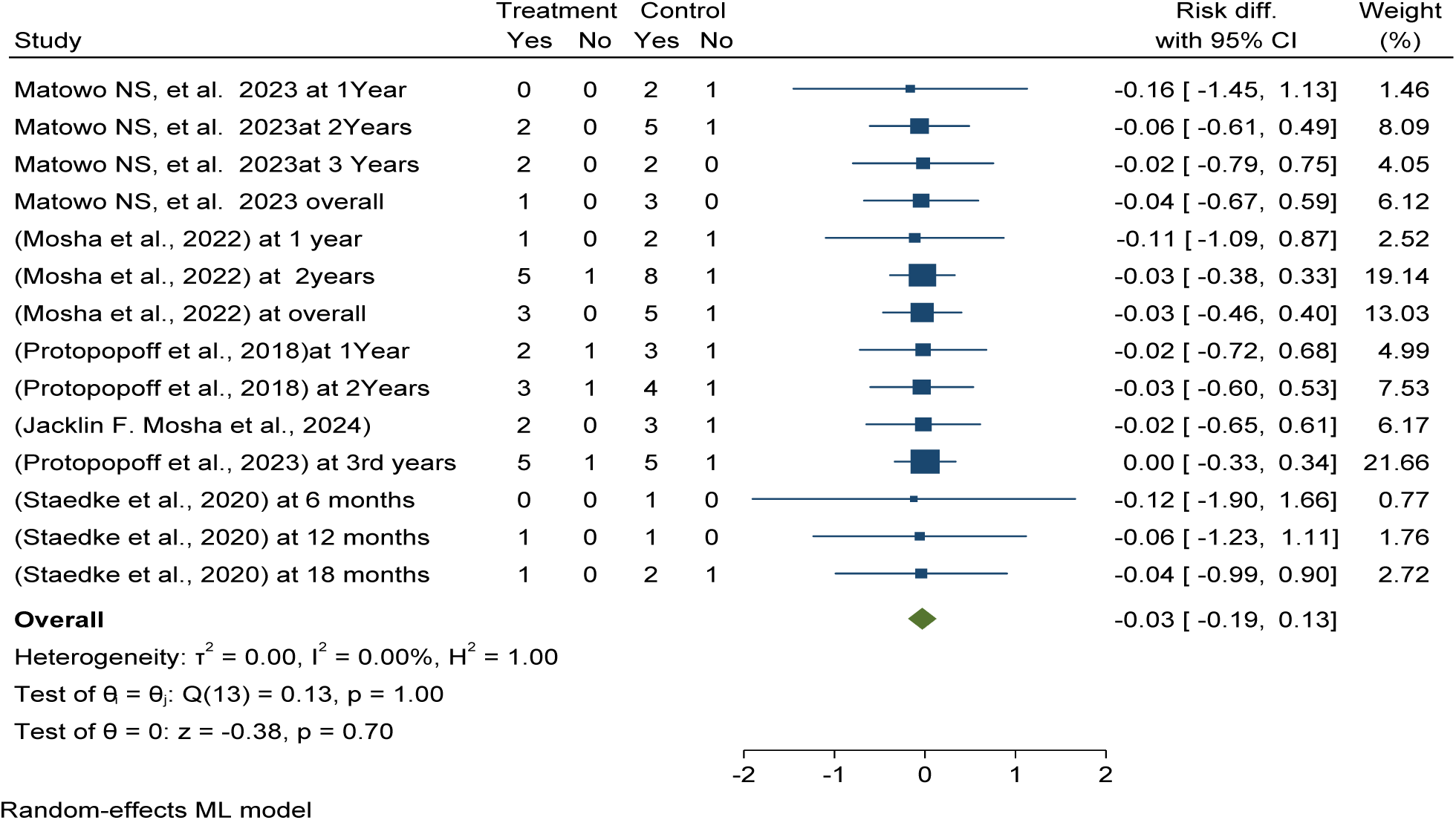
Forest plot shows the effectiveness and efficacy of Piperonyl butoxide long-lasting insecticidal nets (LLINs) in pooled mean indoor vector density reduction compared to pyrethroid-only LLINs in Africa in 2024.

Publication bias was checked using funnel plots looking at symmetrical distribution, and it was objectively verified using Egger’s regression test, which revealed that there was no publication bias (p < 0.148) (Figure 66).

**Figure 66:**
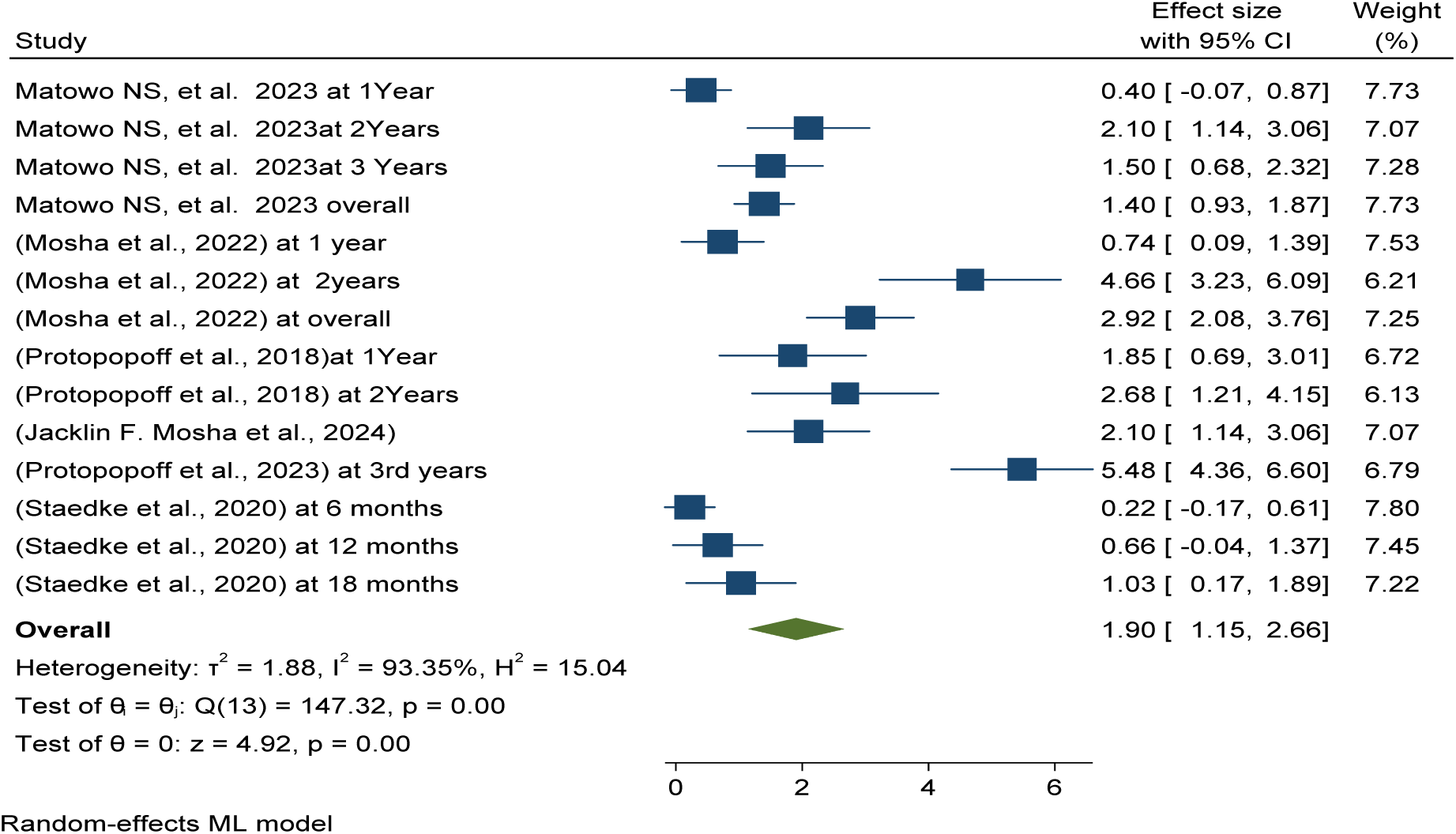
*Forest plot shows post-intervention pooled mean indoor vector density per household per nights among* Piperonyl butoxide *long-lasting insecticidal nets intervention in Africa 2024*

**Figure 67:**
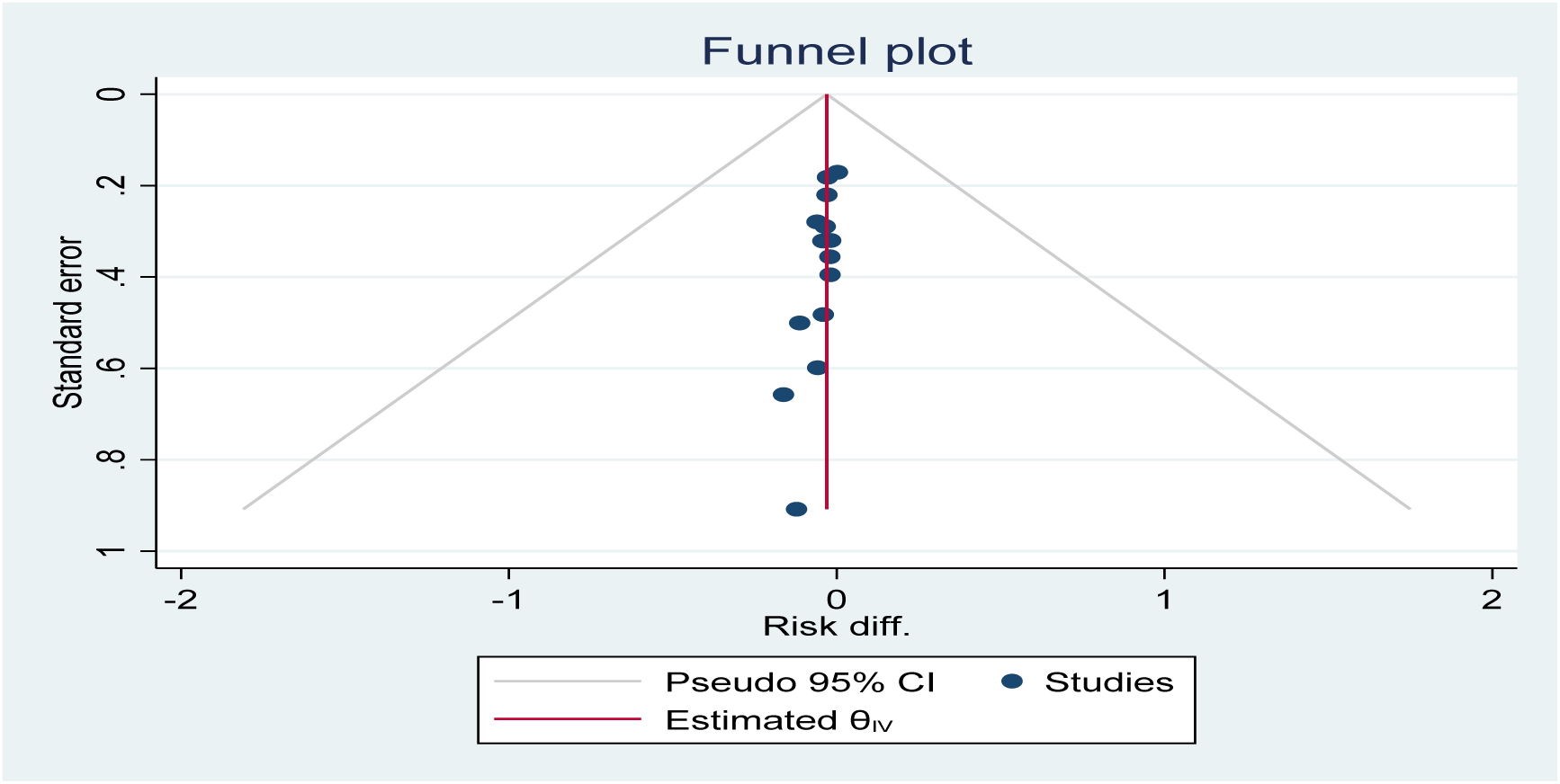
*Funnel plot showing the distribution of included studies post-intervention pooled mean indoor vector density per household per nights among* Piperonyl butoxide *long-lasting insecticidal nets intervention in Africa 2024*

**Figure 68:**
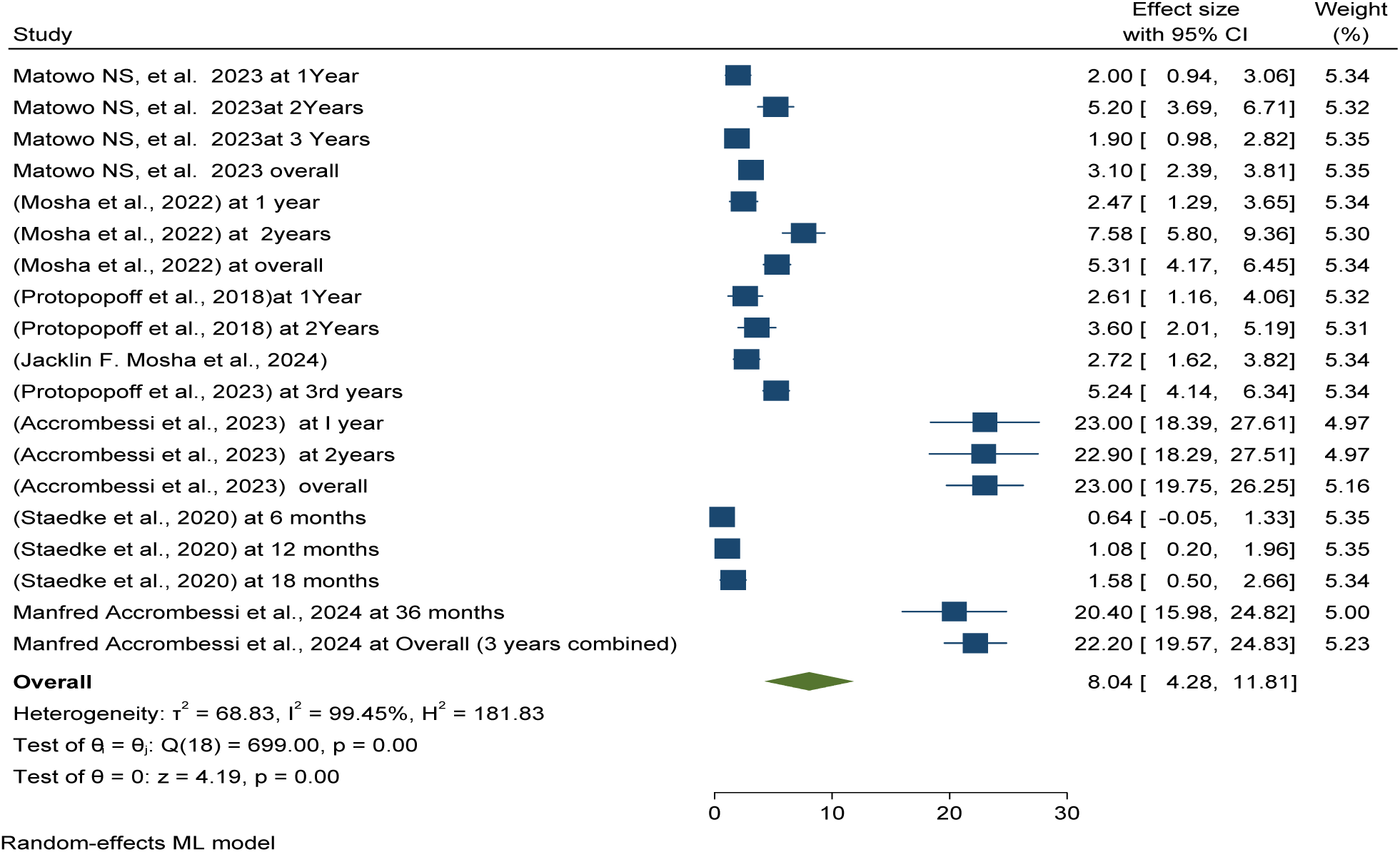
*Forest plot shows post-intervention pooled mean indoor vector density per household per nights among* pyrethroid-only *long-lasting insecticidal nets intervention in Africa 2024*

This meta-analysis determined that the pooled mean indoor vector density per household per night in the pyrethroid-only long-lasting insecticidal nets or control group was 8.04 per household per night as shown in Figures 68.

### Post-intervention pooled effectiveness and efficacy of pyriproxyfen long-lasting insecticidal nets (LLINs) versus pyrethroid-only LLINs for sporozoite rate reduction

Forest plots showed that pyriproxyfen long-lasting insecticidal nets (LLINs) post-intervention effectively and efficiently reduces the sporozoite rate by 15% compared to standard or pyrethroid-only LLINs (RR = -0.02 with a 95%CI of -0.08, 0.37) which demonstrating their effectiveness and efficacy over varying durations from one to three years post-distribution in Africa See details in Figure 69.

**Figure 69:**
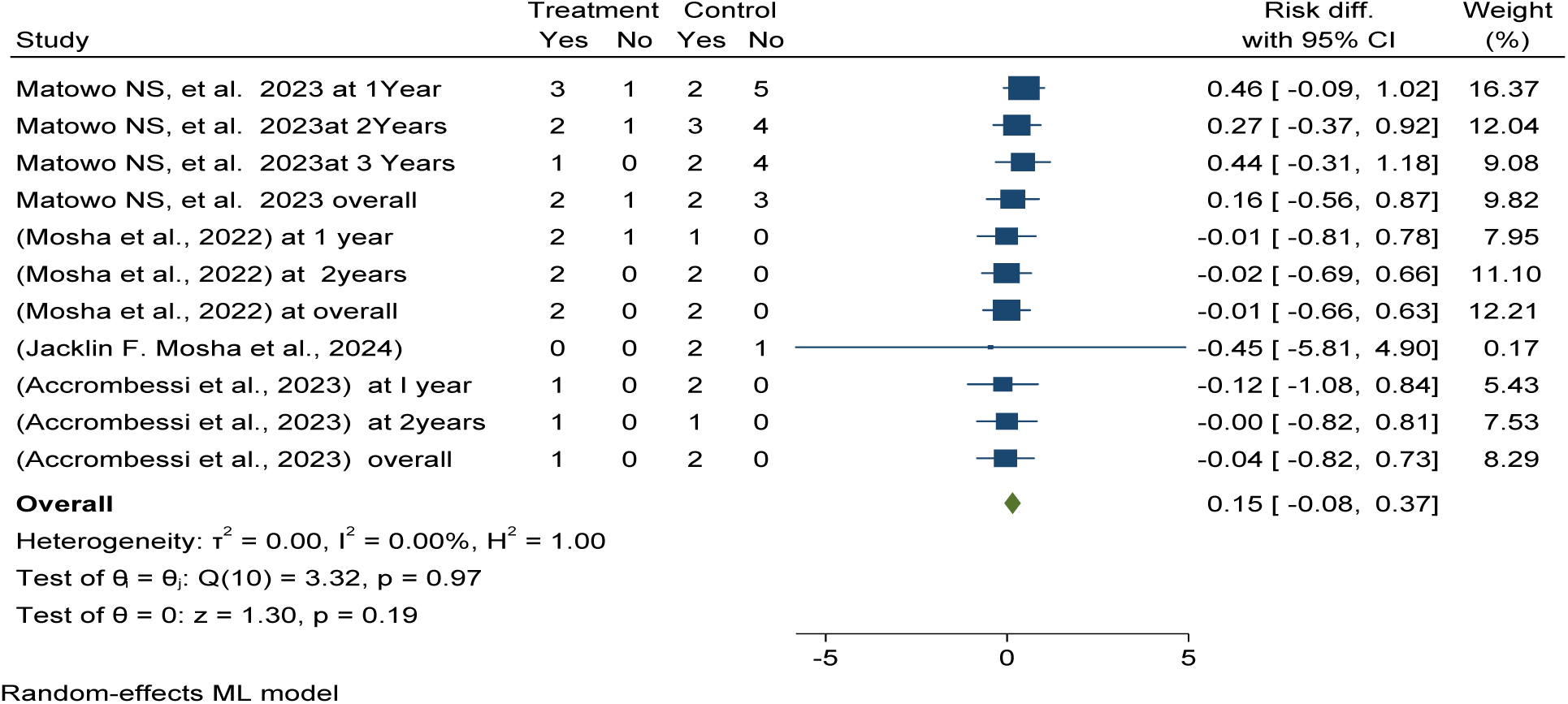
*Forest plot shows* post-intervention pooled effectiveness and efficacy of pyriproxyfen long-lasting insecticidal nets (LLINs) versus pyrethroid-only LLINs for sporozoite rate reduction in Africa 2024

### Pooled prevalence of sporozoite rate among pyriproxyfen LLINs intervention

The pooled prevalence of sporozoite rate was 165 per 100 mosquitoes (95% CI 1.13–2.18) in pyriproxyfen long-lasting insecticidal nets (LLINs) versus 227 per 100 mosquitoes in pyrethroid-only LLINs (95% CI 1.59–2.95) in post-intervention over varying durations from one to three years post-distribution in Africa (see details in Figures 70 and 75).

**Figure 70:**
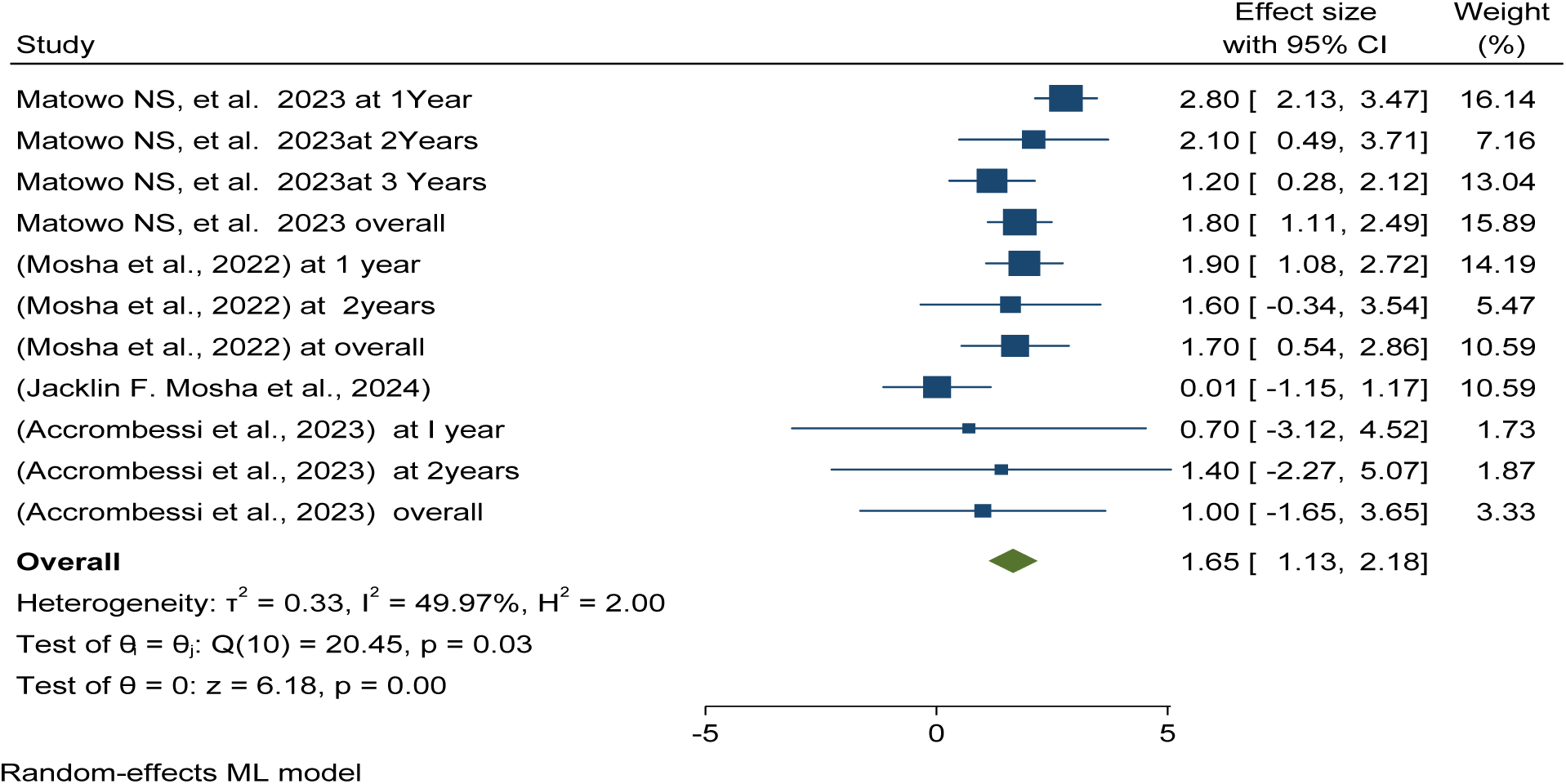
*Forest plot shows* post-intervention pooled prevalence of sporozoite rate among using pyriproxyfen long-lasting insecticidal nets (LLINs) versus pyrethroid-only LLINs for sporozoite rate reduction in Africa 2024

This meta-analysis determined entomological outcomes effectiveness and efficacy in terms of sporozoite rate reducing status reported that in the chlorfenapyr group 9% effective and efficacy in reducing as compared to standard/control of Pyrethroid-only group which demonstrating their effectiveness and efficacy over varying durations from one to three years post-distribution in Africa See details in Figure 71.

**Figure 71:**
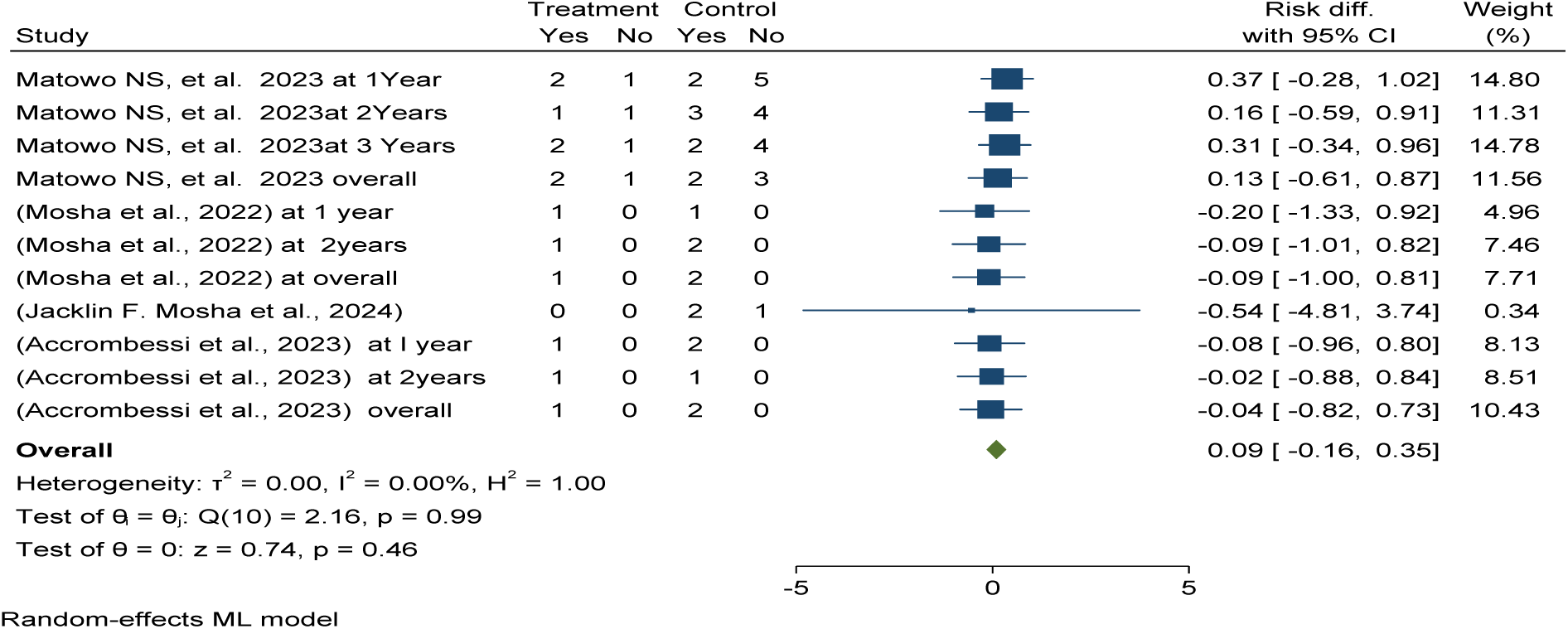
*Forest plot shows* post-intervention pooled effectiveness and efficacy of chlorfenapyr long-lasting insecticidal nets (LLINs) versus pyrethroid-only LLINs for sporozoite rate reduction in Africa 2024

The pooled prevalence of sporozoite rate was 79 per 100 mosquitoes (95% CI 0.49, 1.09 ) in chlorfenapyr long-lasting insecticidal nets (LLINs) versus 227 per 100 mosquitoes in pyrethroid-only LLINs (95% CI 1.59–2.95) in post-intervention over varying durations from one to three years post-distribution in Africa (see details in Figures 72 and 75).

**Figure 72:**
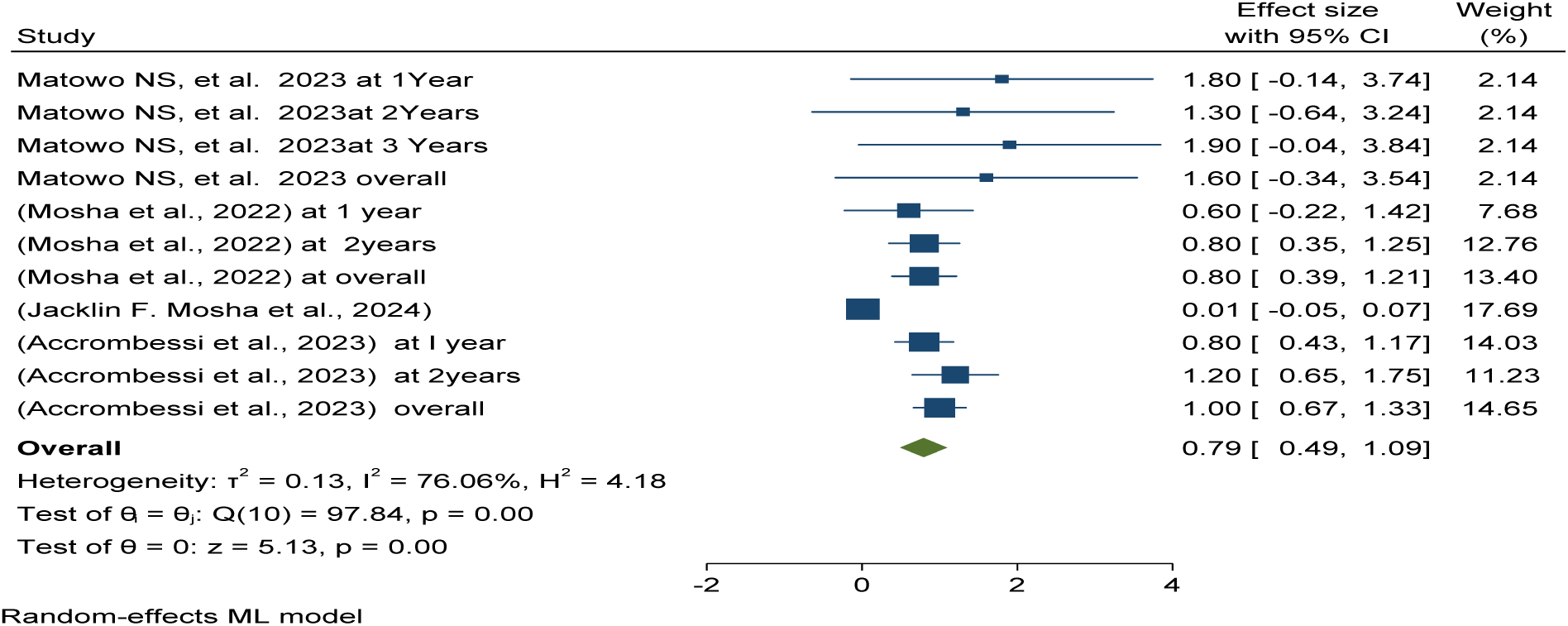
*Forest plot shows* post-intervention pooled prevalence of sporozoite rate among using chlorfenapyr long-lasting insecticidal nets (LLINs) versus pyrethroid-only LLINs for sporozoite rate reduction in Africa 2024

This meta-analysis determined entomological outcomes effectiveness and efficacy in terms of sporozoite rate reducing status reported that in the Piperonyl butoxide group 10% effective and efficacy in reducing as compared to standard/control of Pyrethroid-only group in post-intervention over varying durations from one to three years post-distribution in Africa (see details in Figures 73).

**Figure 73:**
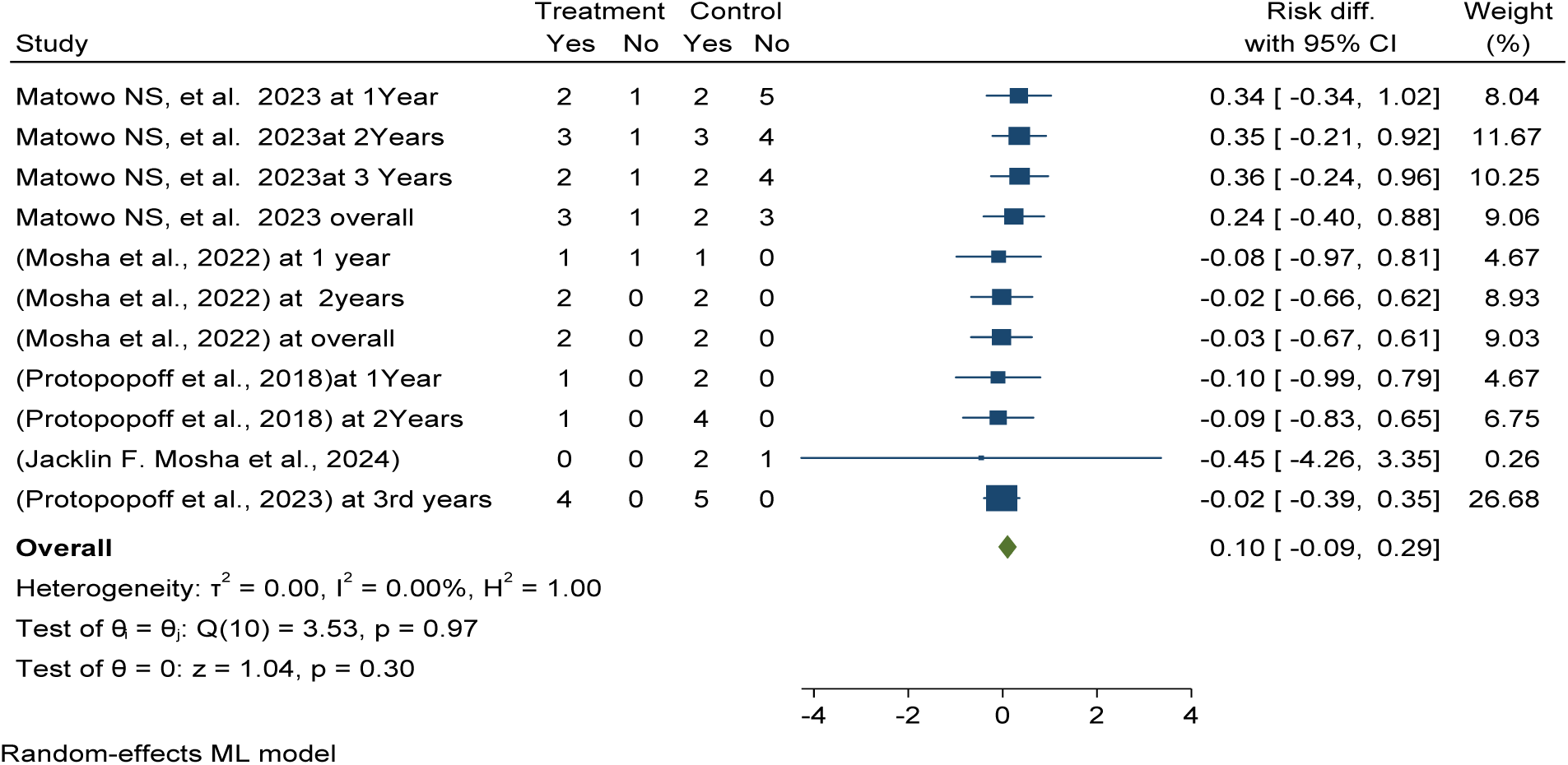
*Forest plot shows* post-intervention pooled effectiveness and efficacy of Piperonyl butoxide long-lasting insecticidal nets (LLINs) versus pyrethroid-only LLINs for sporozoite rate reduction in Africa 2024

The pooled prevalence of sporozoite rate was 172 per 100 mosquitoes with 95% CI (1.06 , 2.38 ) in Piperonyl butoxide long-lasting insecticidal nets (LLINs) versus 227 per 100 mosquitoes in pyrethroid-only LLINs (95% CI 1.59–2.95) in post-intervention over varying durations from one to three years post-distribution in Africa (see details in Figures 74 and 75).

**Figure 74:**
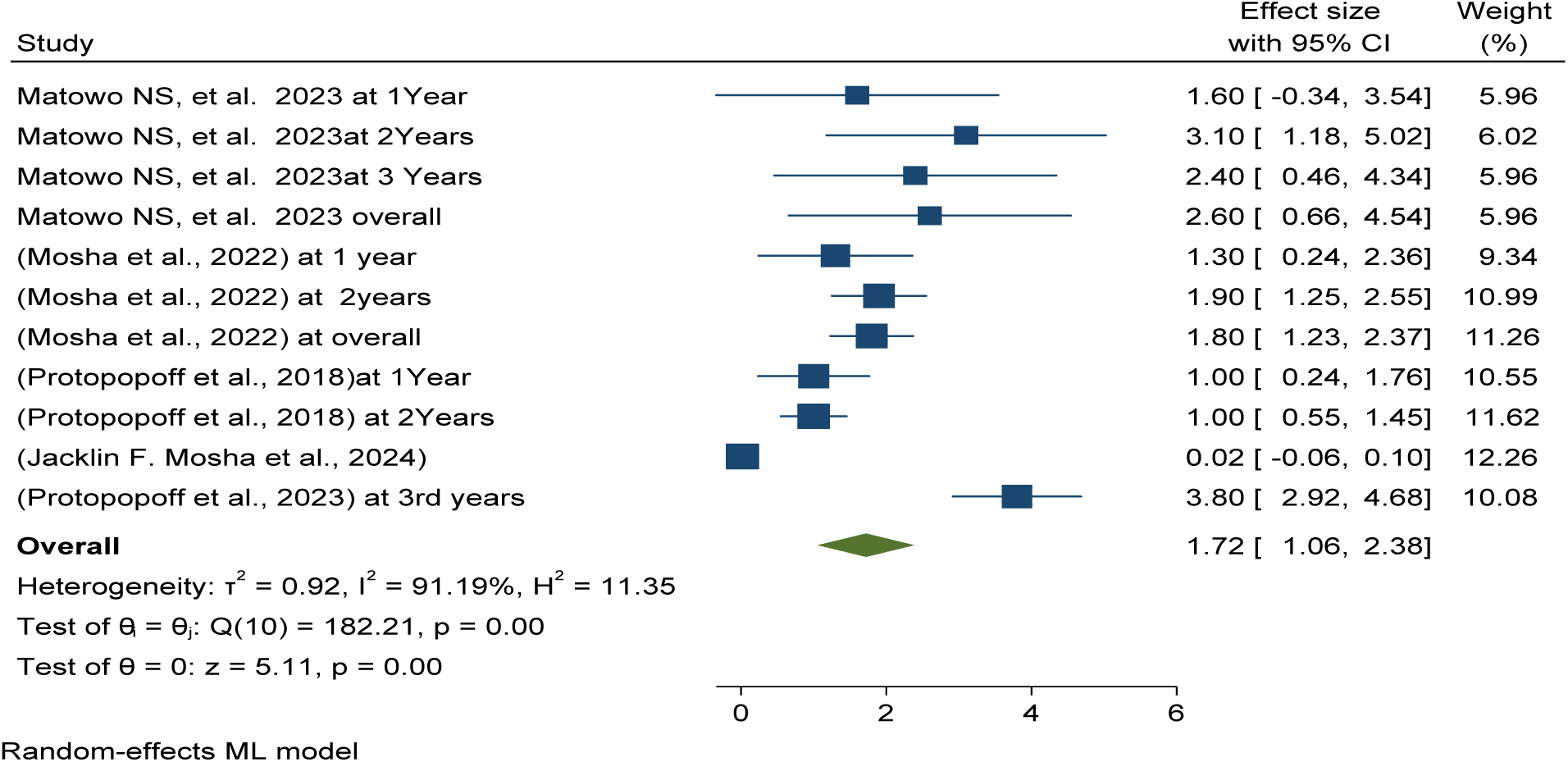
*Forest plot shows* post-intervention pooled prevalence of sporozoite rate among using Piperonyl butoxide long-lasting insecticidal nets (LLINs) versus pyrethroid-only LLINs for sporozoite rate reduction in Africa 2024

**Figure 75:**
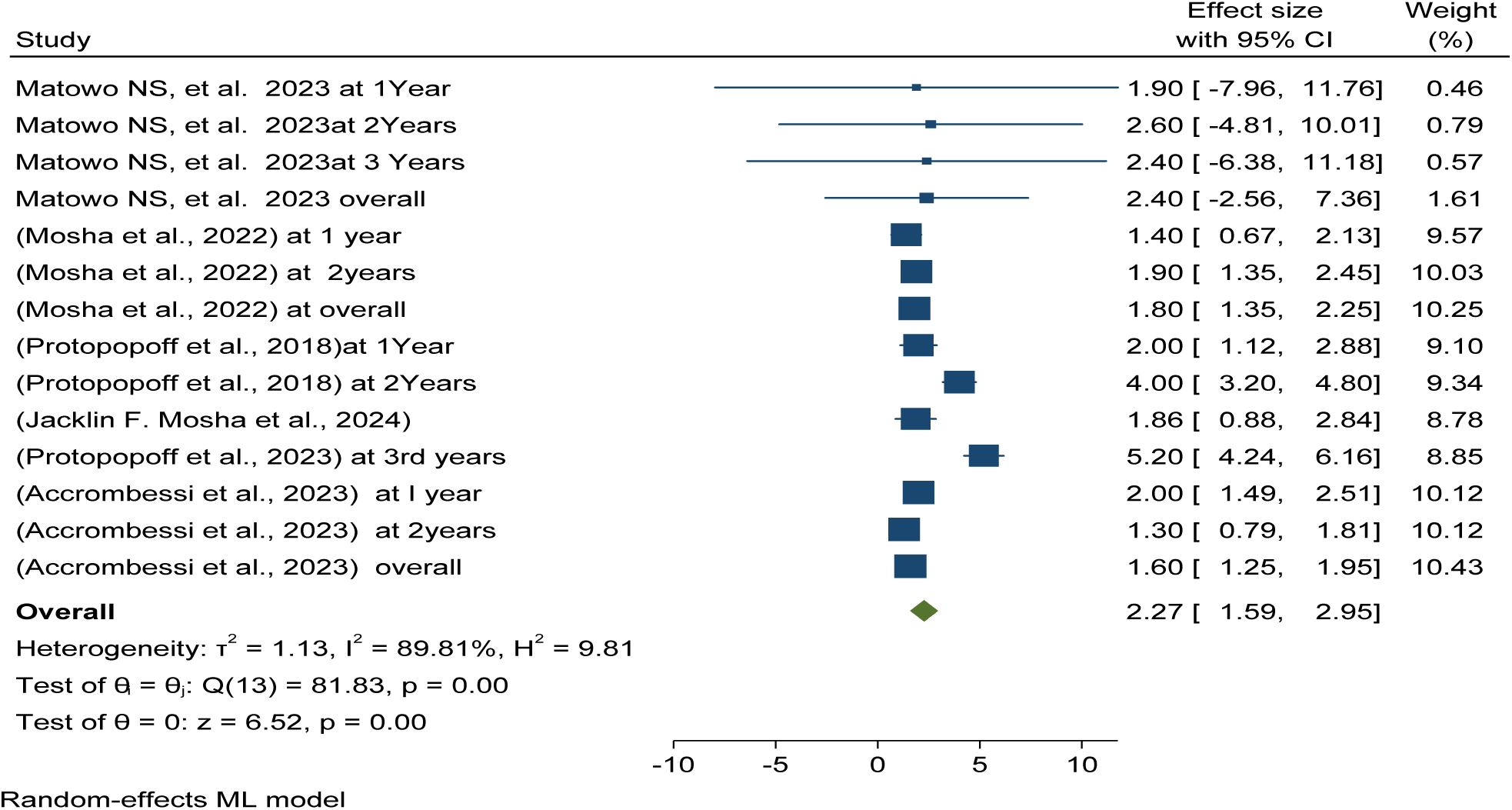
*Forest plot shows* post-intervention pooled prevalence of sporozoite rate among using pyrethroid-only long-lasting insecticidal nets (LLINs) for sporozoite rate reduction in Africa 2024

The pooled prevalence of sporozoite rate was 227 per 100 mosquitoes in pyrethroid-only long-lasting insecticidal nets or control group (95% CI 1.59–2.95) in post-intervention over varying durations from one to three years post-distribution in Africa (see details in Figure 75).

### Post-intervention entomological inoculation rate among different long-lasting insecticidal nets

This meta-analysis determined entomological outcomes effectiveness and efficacy in inoculation rate terms of mean per household per night reduction reported that in the Pyriproxyfen group were in reduced in 7% as compared to standard/control of Pyrethroid-only group in post-intervention over varying durations from one to three years post-distribution in Africa (see details in Figure 76).

**Figure 76:**
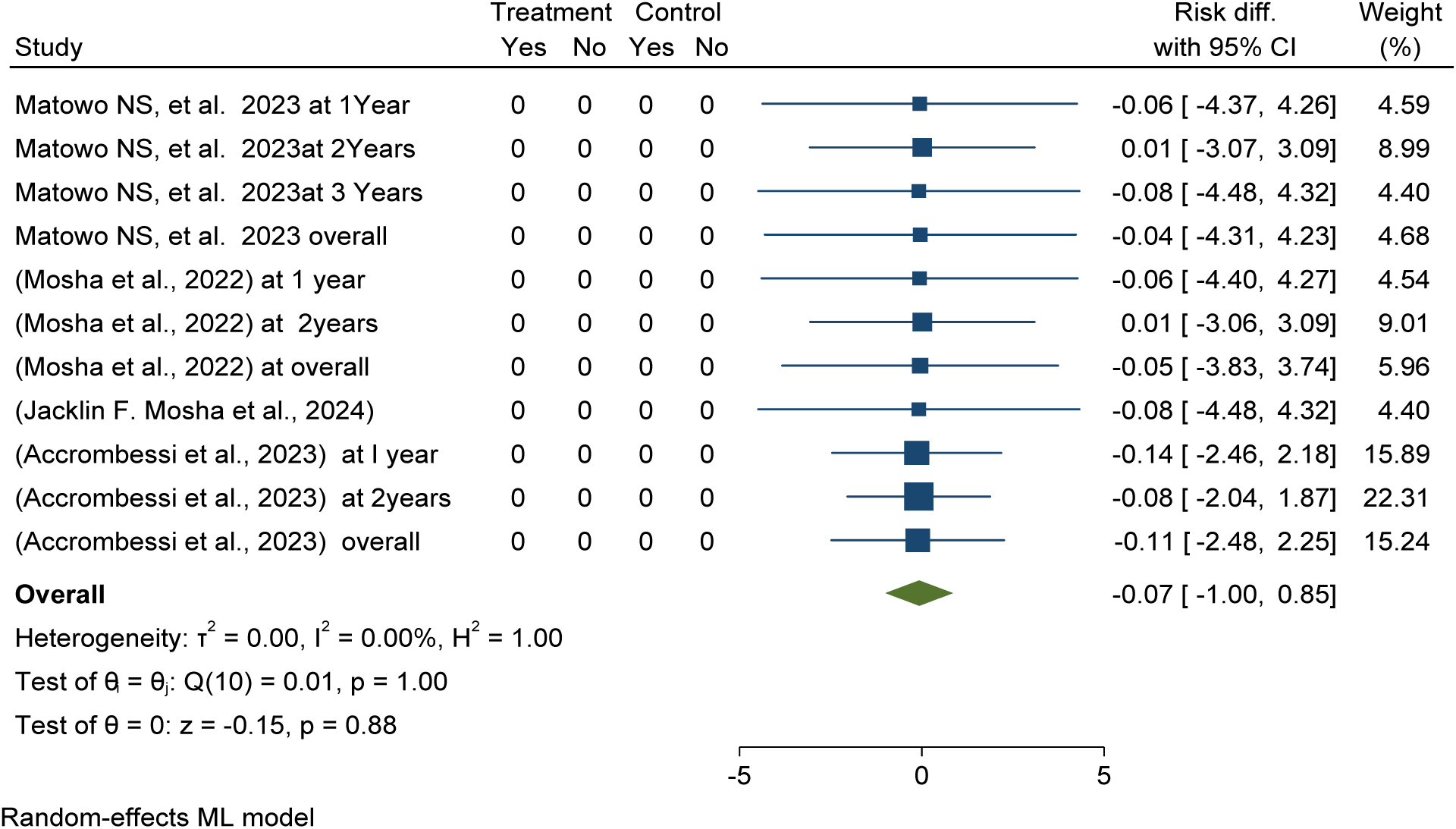
*Forest plot shows* post-intervention pooled effectiveness and efficacy of Pyriproxyfen long-lasting insecticidal nets (LLINs) versus pyrethroid-only LLINs for mean entomological inoculation rate reduction in Africa 2024

This meta-analysis determined pooled mean entomological inoculation rate per household per night was 4per 100 household per night with 95% CI (-0.00, 0.08) in the Pyriproxyfen group compared to 7per 100 household per night in Pyrethroid-only group with 95% CI 0.03,0.12) in post-intervention over varying durations from one to three years post-distribution in Africa (see details in Figure 77).

**Figure 77:**
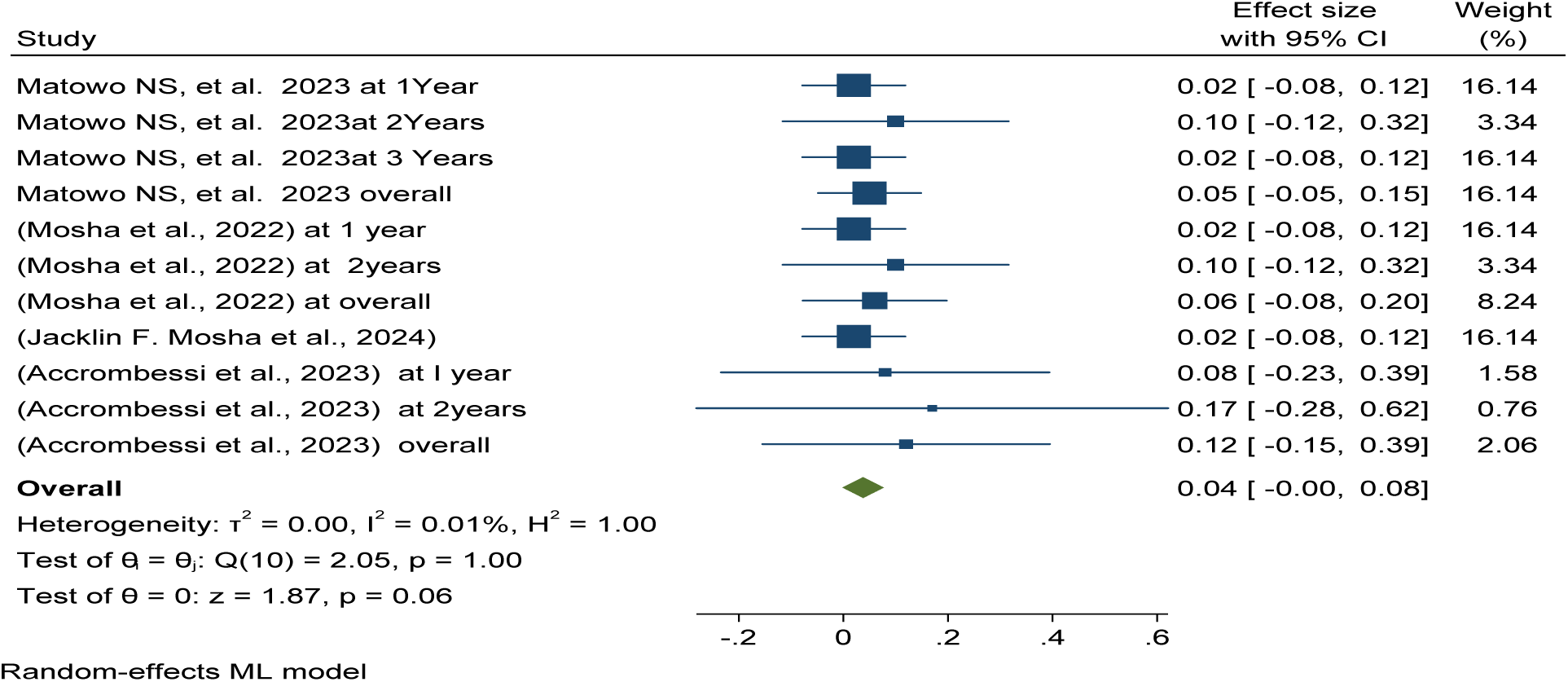
*Forest plot shows* post-intervention pooled mean entomological inoculation rate per household per night among using the Pyriproxyfen long-lasting insecticidal nets (LLINs) for sporozoite rate reduction in Africa 2024.

Publication bias was checked using funnel plots looking at symmetrical distribution, and it was objectively verified using Egger’s regression test, which revealed that there was no publication bias (p < 0.38) (Figure 78).

**Figure 78:**
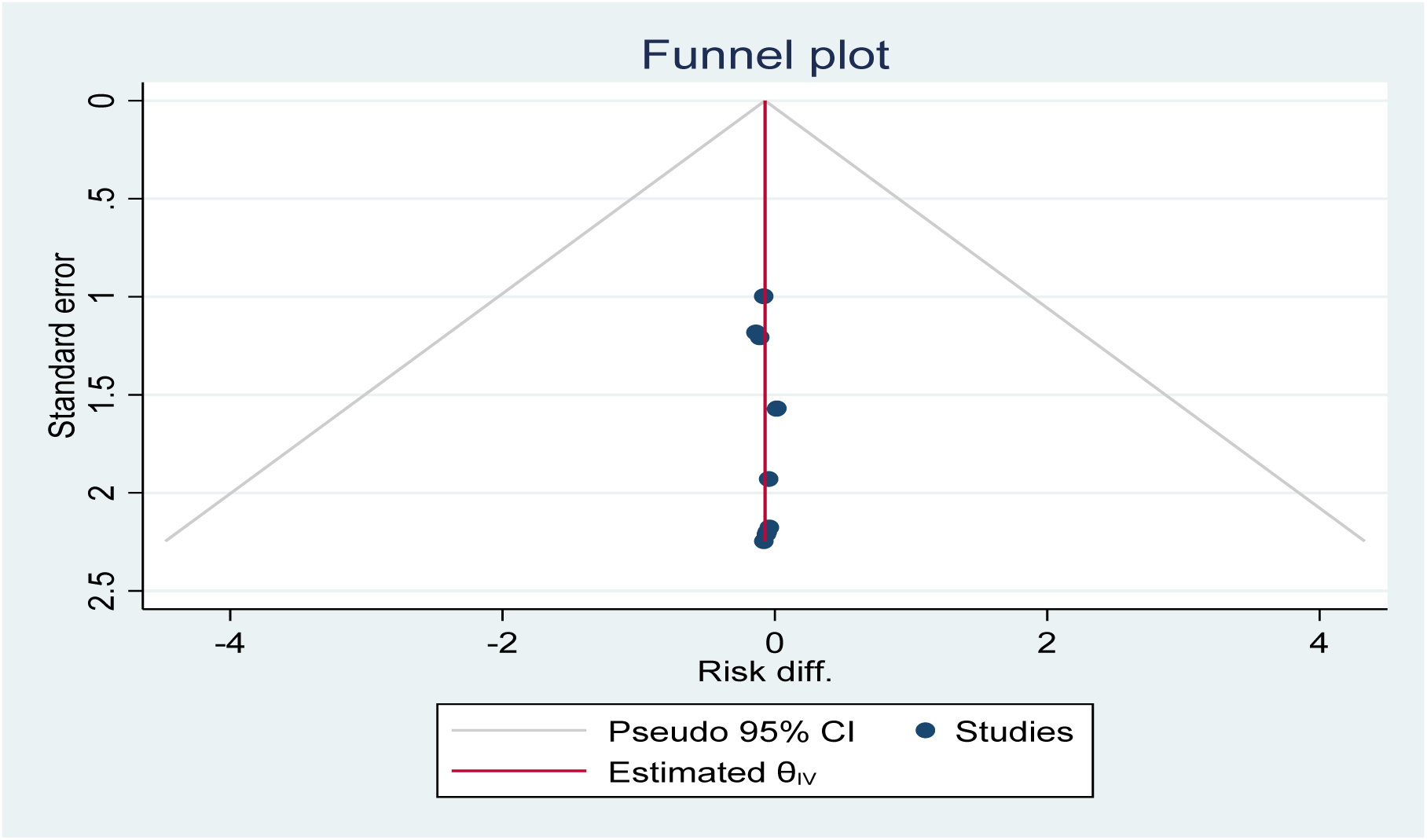
Funnel plot showing the distribution of included studies post-intervention pooled mean entomological inoculation rate per household per nights among using Pyriproxyfen long-lasting insecticidal nets intervention in Africa 2024

This meta-analysis determined entomological outcomes effectiveness and efficacy in terms of mean per household per night inoculation rate reduction reported that in the chlorfenapyr group were in reduced in 23% as compared to standard/control of Pyrethroid-only group in post-intervention over varying durations from one to three years post-distribution in Africa (see details in Figure 79).

**Figure 79:**
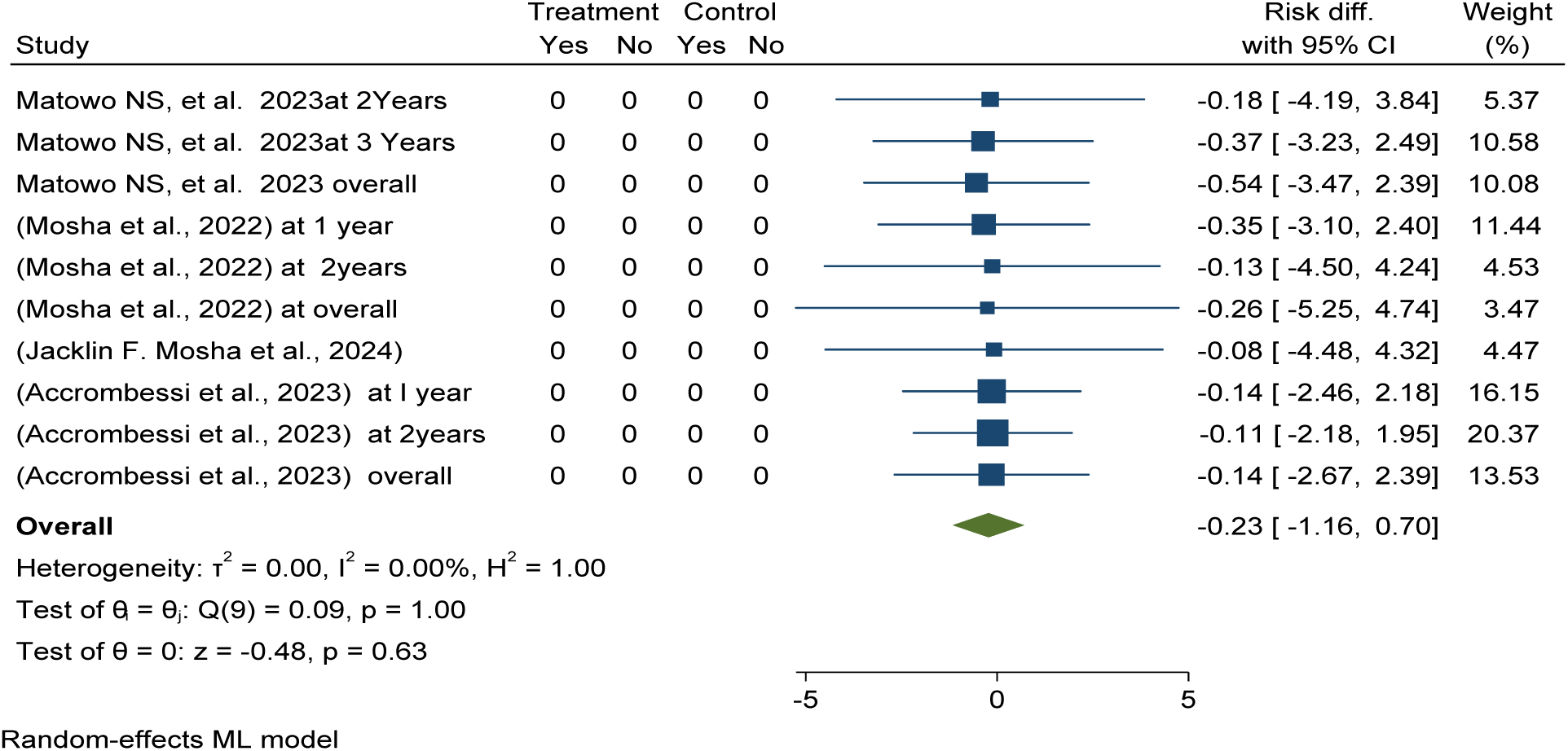
*Forest plot shows* post-intervention pooled effectiveness and efficacy of chlorfenapyr long-lasting insecticidal nets (LLINs) versus pyrethroid-only LLINs for mean entomological inoculation rate reduction in Africa 2024

This meta-analysis determined pooled mean inoculation rate per household per night 4per 100 household per night in chlorfenapyr LLNs intervention with 95% CI (-0.00, 0.08) versus 7per 100 household per night with 95% CI 0.03, 0.12) in post-intervention over varying durations from one to three years post-distribution in Africa (see details in Figure 80 ).

**Figure 80:**
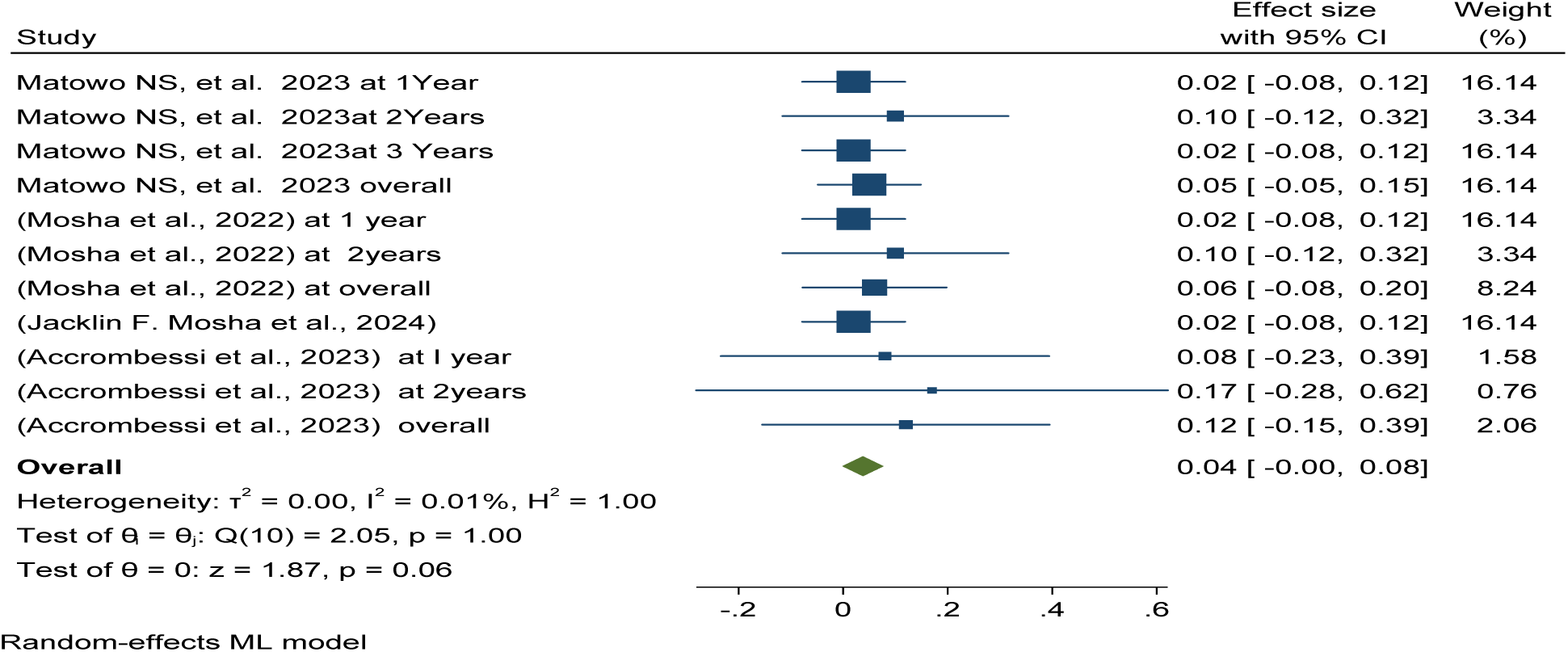
*Forest plot shows* post-intervention pooled mean entomological inoculation rate per household per night among using the chlorfenapyr long-lasting insecticidal nets (LLINs) for sporozoite rate reduction in Africa 2024.

This meta-analysis determined entomological outcomes effectiveness and efficacy in terms of mean per household per night inoculation rate reduction reported that in the Piperonyl butoxide group were in reduced in 12% as compared to standard/control of Pyrethroid-only group in post-intervention over varying durations from one to three years post-distribution in Africa (see details in Figure 81).

**Figure 81:**
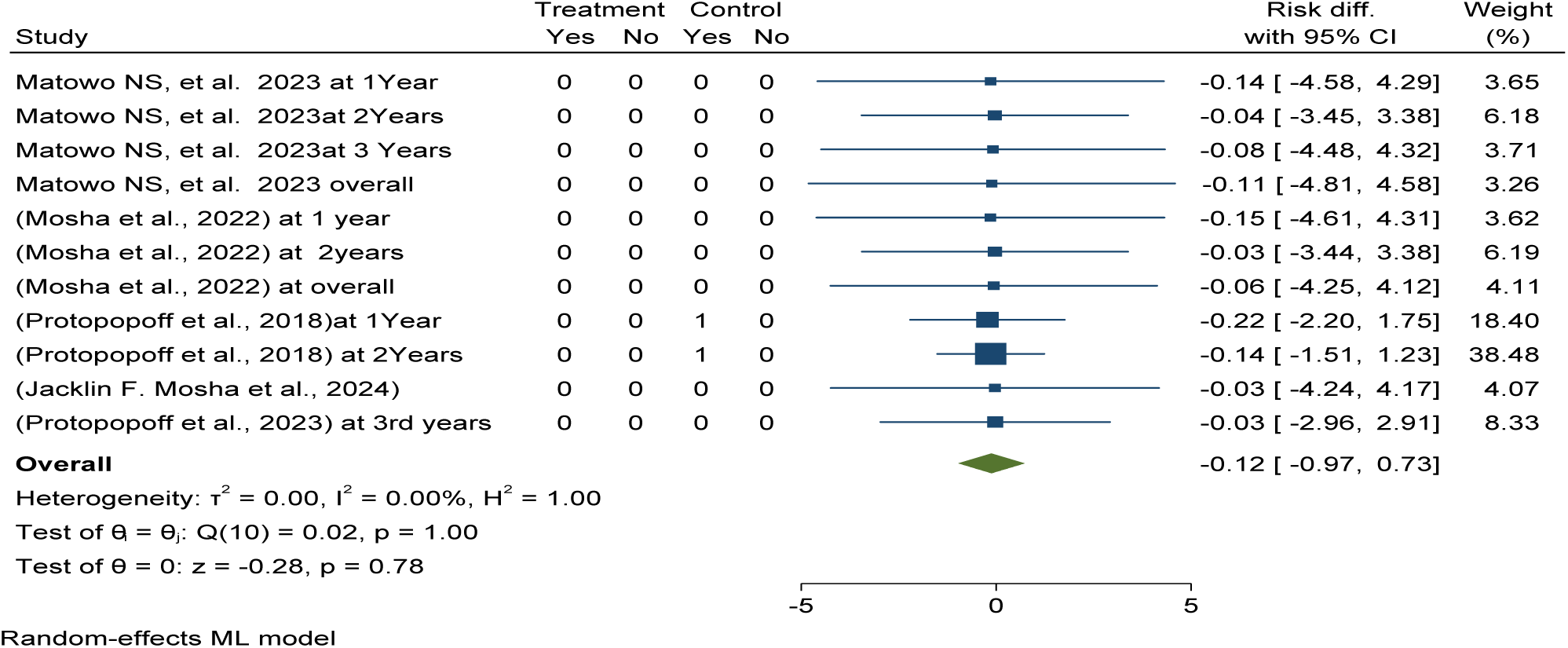
*Forest plot shows* post-intervention pooled effectiveness and efficacy of Piperonyl butoxide long-lasting insecticidal nets (LLINs) versus pyrethroid-only LLINs for mean entomological inoculation rate reduction in Africa 2024

This meta-analysis determined pooled mean inoculation rate per household per night 3per 100 household per night with 95% CI (-0.00, 0.06) in Piperonyl butoxide versus 7per 100 household per night with 95% CI 0.03, 0.12) pyrethroid-only LLINs in post-intervention over varying durations from one to three years post-distribution in Africa (see details in Figure 82).

**Figure 82:**
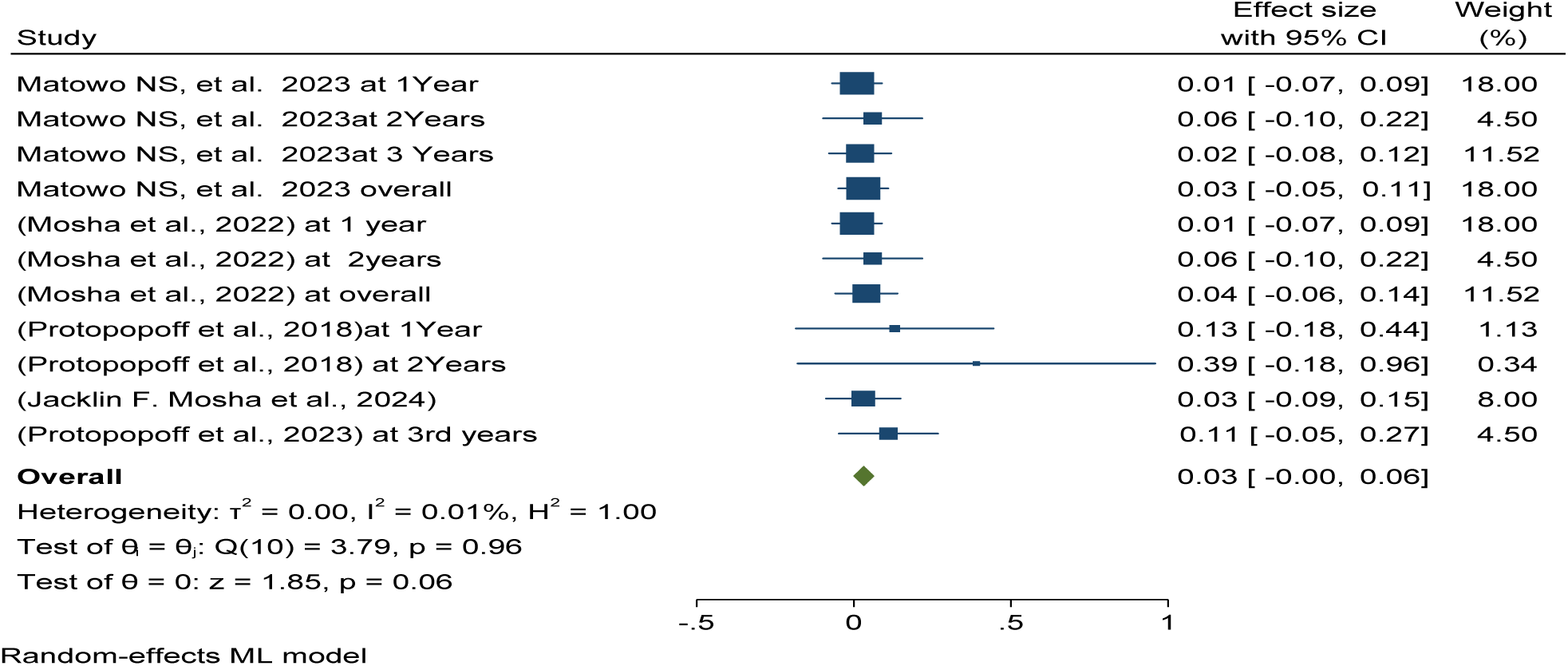
*Forest plot shows* post-intervention pooled mean entomological inoculation rate per household per night among using the Piperonyl butoxide long-lasting insecticidal nets (LLINs) in Africa 2024.

Publication bias was checked using funnel plots looking at symmetrical distribution, and it was objectively verified using Egger’s regression test, which revealed that there was no publication bias (p < 0.83) (Figure 83).

**Figure 83:**
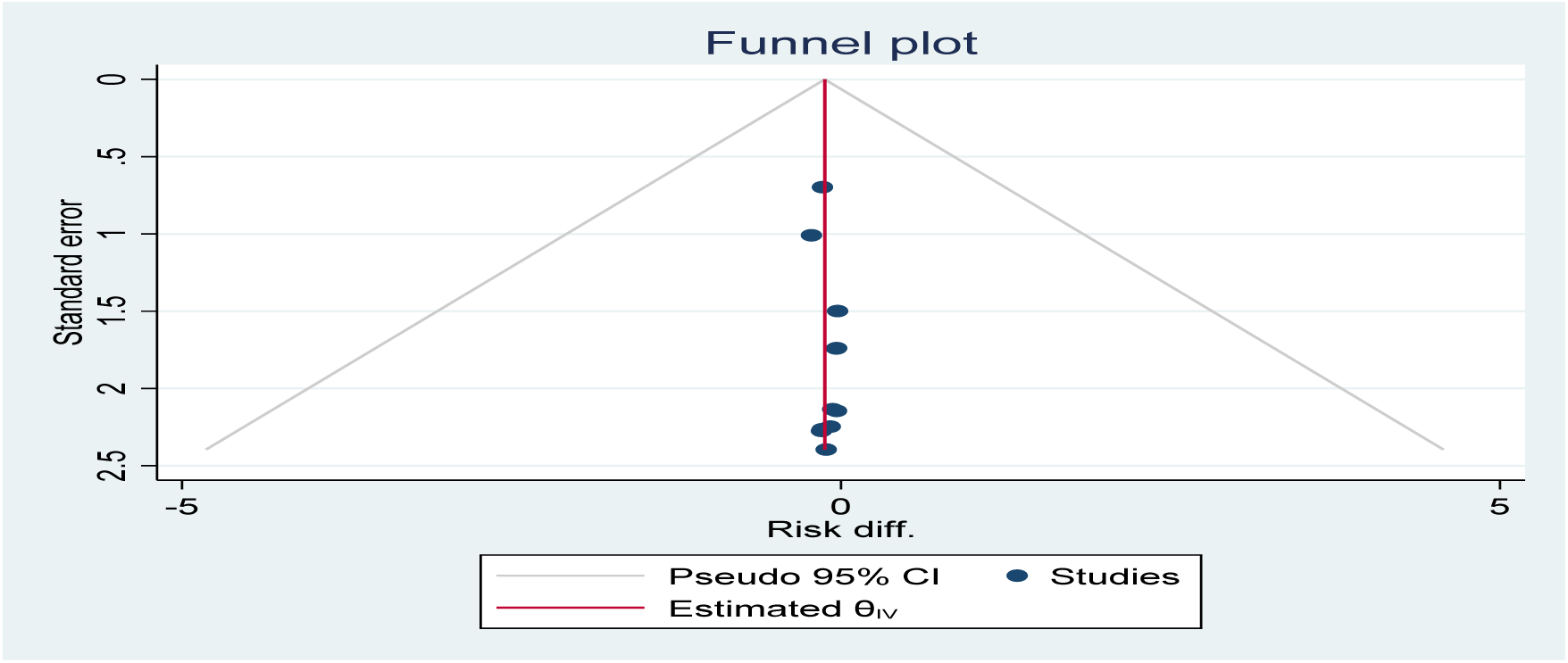
*Funnel plot showing the distribution of included studies pooled mean entomological inoculation rate per household per night among using* piperonyl butoxide *long-lasting insecticidal nets (LLINs) versus pyrethroid-only LLINs for malaria control in Africa in 2024*.

This review determined pooled mean anopheles mosquito inoculation rate using Pyrethroid-only group long-lasting insecticidal nets was 7per 100 household per night with a 95% CI of - 0.03, 0.12 in post-intervention over varying durations from one to three years post-distribution in Africa (see details in Figure 84).

**Figure 84:**
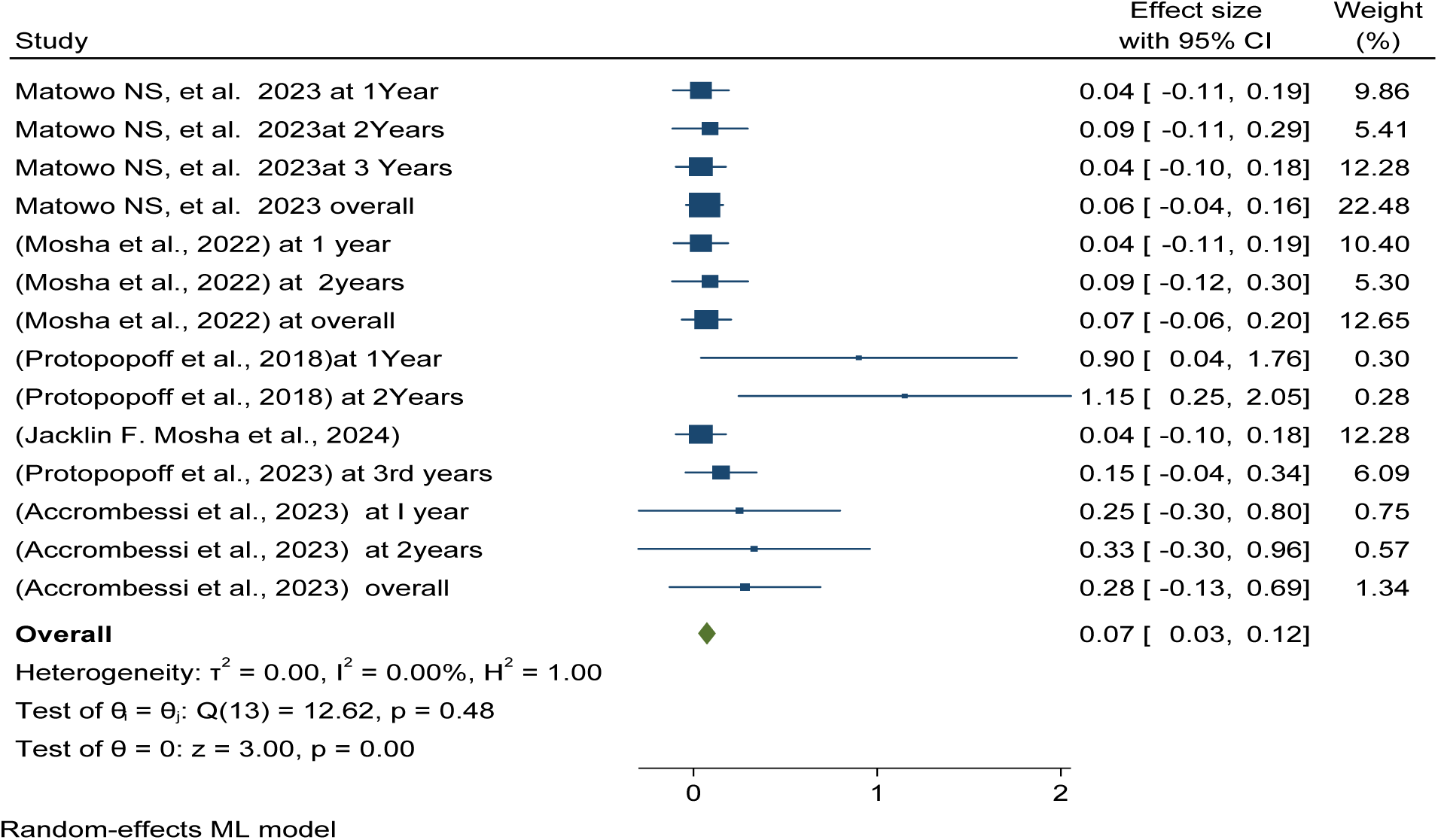
*Forest plot shows* post-intervention pooled mean entomological inoculation rate per household per night among using the Pyrethroid-only long-lasting insecticidal nets (LLINs) in Africa 2024.

The adopted GRADE (Grading of Recommendations Assessment, Development and Evaluation) as the method for assessing the quality of a body of evidence and for determining the direction and strength of the resulting recommendations. This generated evidence was evaluated the effectiveness and efficacy of pyriproxyfen, chlorfenapyr, and piperonyl butoxide long-lasting insecticidal nets against the pyrethroid-only LLINs.

This study found that PYR-only LLINs (control arm) had higher pooled prevalence of malaria infection, case incidence, anemia, mean indoor vector density, inoculation rate, and sporozoite rate as compared to intervention group (PPF, CFP, and PBO LLINs see details in Table 1.

**Table 1.**
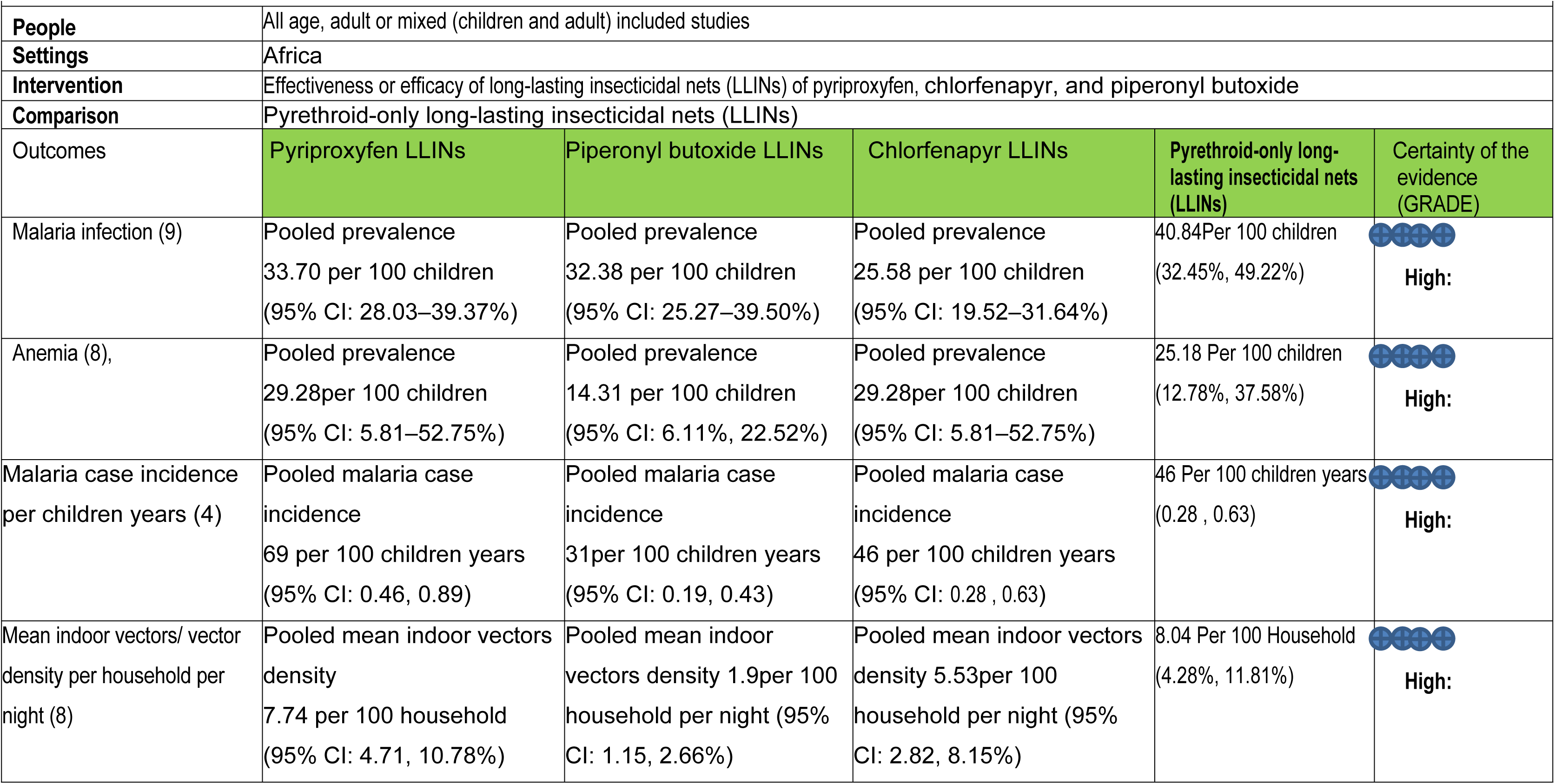
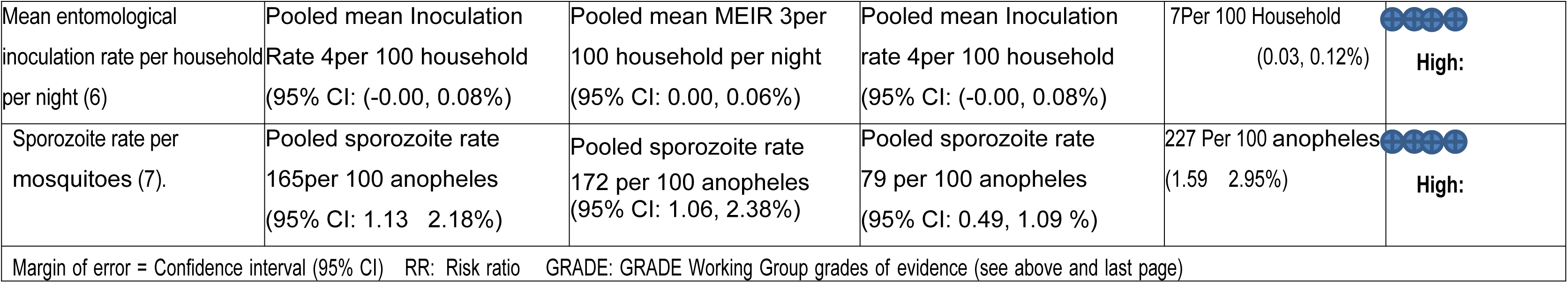
Evidence profile compared the effectiveness and efficacy of pyriproxyfen, chlorfenapyr, and piperonyl butoxide LLINs with pyrethroid-only LLINs for malaria control in Africa.

The evidence generated from this meta-analysis reveals that pyriproxyfen (PPF) long-lasting insecticidal nets (LLINs) have no significant difference in malaria infection, case incidence, or anemia reduction among children as compared to pyrethroid-only LLINs. However, this study found that Pyriproxyfen (PPF) LLINs effectively and efficaciously reduce indoor vector density, entomological inoculation rate, and sporozoite rate of malaria parasites compared to pyrethroid-only LLINs.

The study found that chlorfenapyr (CFP) and piperonyl butoxide (PBO) long-lasting insecticidal nets (LLINs) are highly effective and efficacious in reducing malaria infection, case incidence, and anemia among children, as well as reducing indoor vector density, inoculation rate, and sporozoite rate in Africa as compared to pyrethroid-only LLINs.

Critical appraisal of individual randomized control trials revealed that 100% of the studies scored high quality, and Cochrane methodology was used to assess the risk of bias and evaluate evidence quality, which was graded as high. This research provides a very good indication of the likely effect. The likelihood that the effect will be substantially different is low see details in Table 2.

**Table 2.**
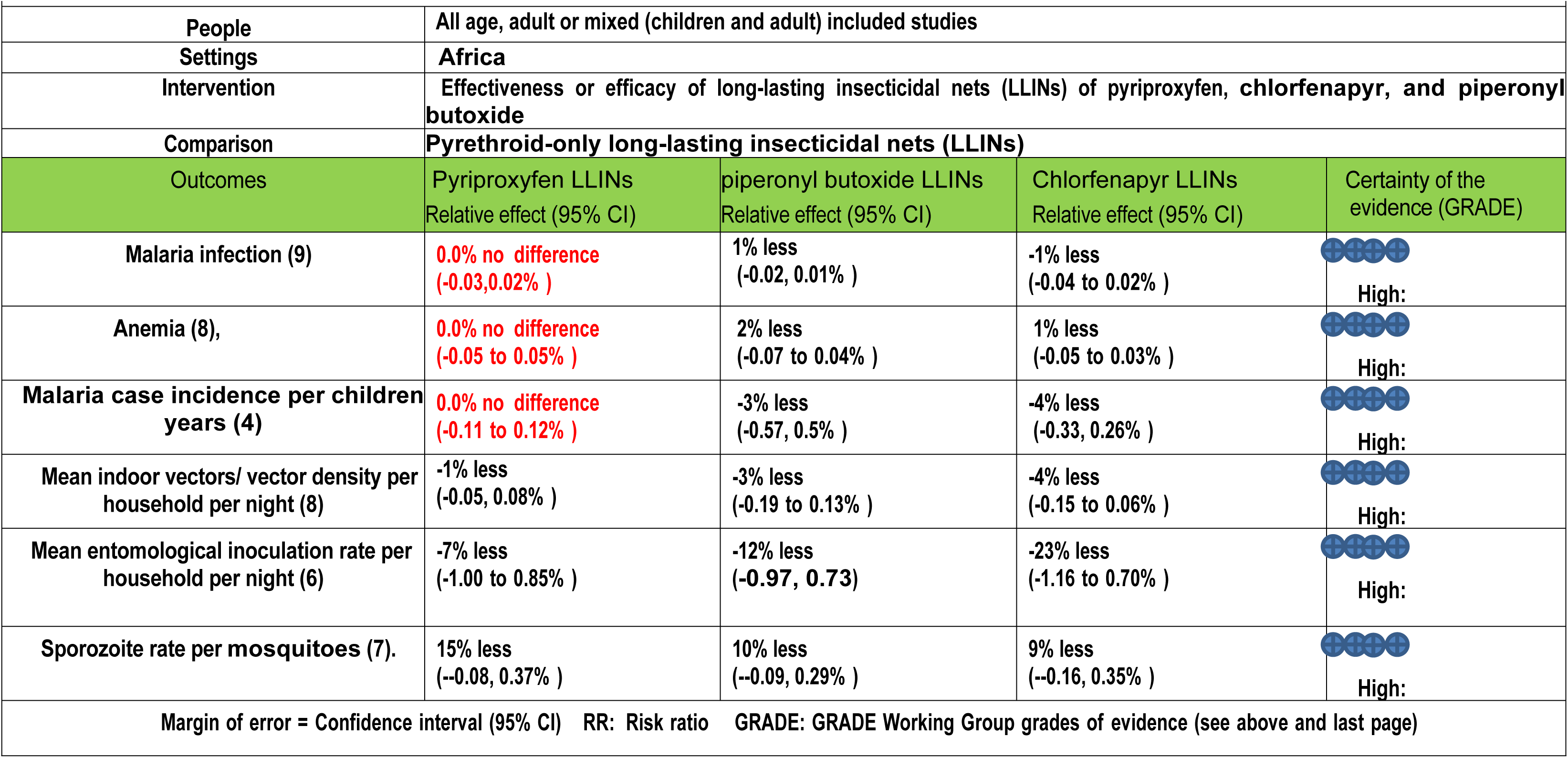
Evidence profile compared the effectiveness and efficacy of pyriproxyfen, chlorfenapyr, and piperonyl butoxide LLINs with pyrethroid-only LLINs for malaria control in Africa.

|  |  |  |  |  |
| --- | --- | --- | --- | --- |
| People | All age, adult or mixed (children and adult) included studies |  |  |  |
| Settings | Africa |  |  |  |
| Intervention | Effectiveness or efficacy of long-lasting insecticidal nets (LLINs) of pyriproxyfen, chlorfenapyr, and piperonyl butoxide |  |  |  |
| Comparison | Pyrethroid-only long-lasting insecticidal nets (LLINs) |  |  |  |
| Outcomes | Pyriproxyfen LLINs<br>Relative effect (95% CI) | piperonyl butoxide LLINs<br>Relative effect (95% CI) | Chlorfenapyr LLINs<br>Relative effect (95% CI) | Certainty of the evidence (GRADE) |
| Malaria infection (9) | 0.0% no difference<br>(-0.03, 0.02% ) | 1% less<br>(-0.02, 0.01% ) | -1% less<br>(-0.04 to 0.02% ) | High: |
| Anemia (8), | 0.0% no difference<br>(-0.05 to 0.05% ) | 2% less<br>(-0.07 to 0.04% ) | 1% less<br>(-0.05 to 0.03% ) | High: |
| Malaria case incidence per children years (4) | 0.0% no difference<br>(-0.11 to 0.12% ) | -3% less<br>(-0.57, 0.5% ) | -4% less<br>(-0.33, 0.26% ) | High: |
| Mean indoor vectors/ vector density per household per night (8) | -1% less<br>(-0.05, 0.08% ) | -3% less<br>(-0.19 to 0.13% ) | -4% less<br>(-0.15 to 0.06% ) | High: |
| Mean entomological inoculation rate per household per night (6) | -7% less<br>(-1.00 to 0.85% ) | -12% less<br>(-0.97, 0.73) | -23% less<br>(-1.16 to 0.70% ) | High: |
| Sporozoite rate per mosquitoes (7). | 15% less<br>(--0.08, 0.37% ) | 10% less<br>(--0.09, 0.29% ) | 9% less<br>(--0.16, 0.35% ) | High: |
| Margin of error = Confidence interval (95% CI) RR: Risk ratio GRADE: GRADE Working Group grades of evidence (see above and last page) |  |  |  |  |

The evidence generated from this meta-analysis reveals that pyriproxyfen (PPF) long-lasting insecticidal nets (LLINs) have no significant difference in malaria infection, case incidence, or anemia reduction among children as compared to pyrethroid-only LLINs. Based on above finding as input this study also plan evaluated the effectiveness and efficacy of chlorfenapyr, and piperonyl butoxide long-lasting insecticidal nets compared to pyriproxyfen LLINs for malaria control in Africa

### Pooled post-intervention effectiveness and efficacy of chlorfenapyr long-lasting insecticidal nets (LLINs) versus pyriproxyfen LLINs malaria infection reduction in Africa

This study determined the overall pooled post-intervention effectiveness and efficacy of chlorfenapyr long-lasting insecticidal nets (LLINs) in malaria infection reduction in children, which was reduced by 1% as compared to pyriproxyfen LLINs (ARR = -0.01, 95% CI = -0.04, 0.03). See details in Figure 85.

**Figure 85:**
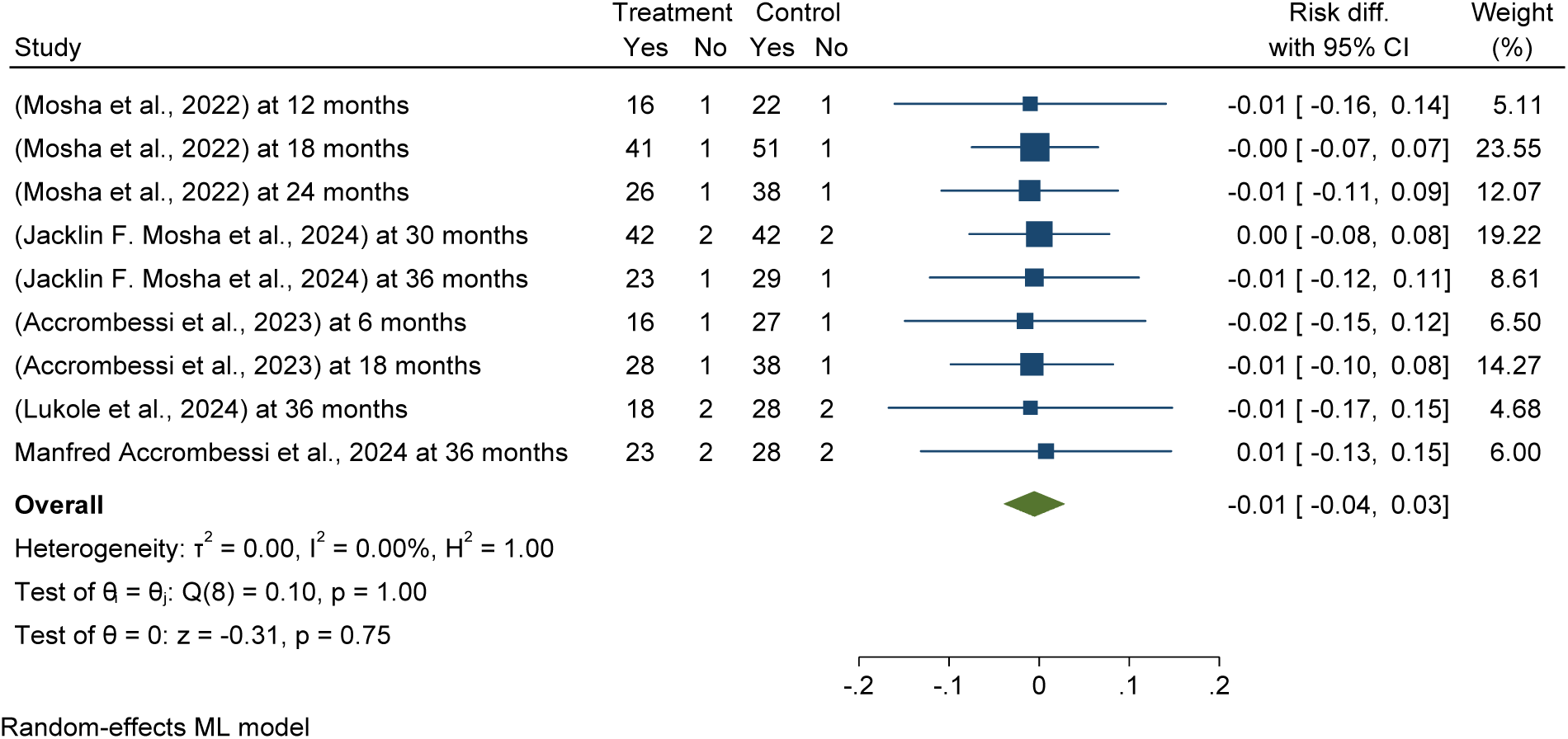
*Forest plot shows* pooled post-intervention effectiveness and efficacy of chlorfenapyr long-lasting insecticidal nets (LLINs) versus pyriproxyfen LLINs malaria infection reduction in Africa 2024

The subgroup analysis of post-intervention follow up effectiveness and efficacy of chlorfenapyr long-lasting insecticidal nets (LLINs) showed a significant reduction in malaria infection among children over different durations, with a 2% reduction at six months and 1% at 36 months post-distribution of LLINs, compared to pyriproxyfen LLINs (ARR = -0.01, 95% CI = -0.04, 0.03). See details in Figure 86.

**Figure 86:**
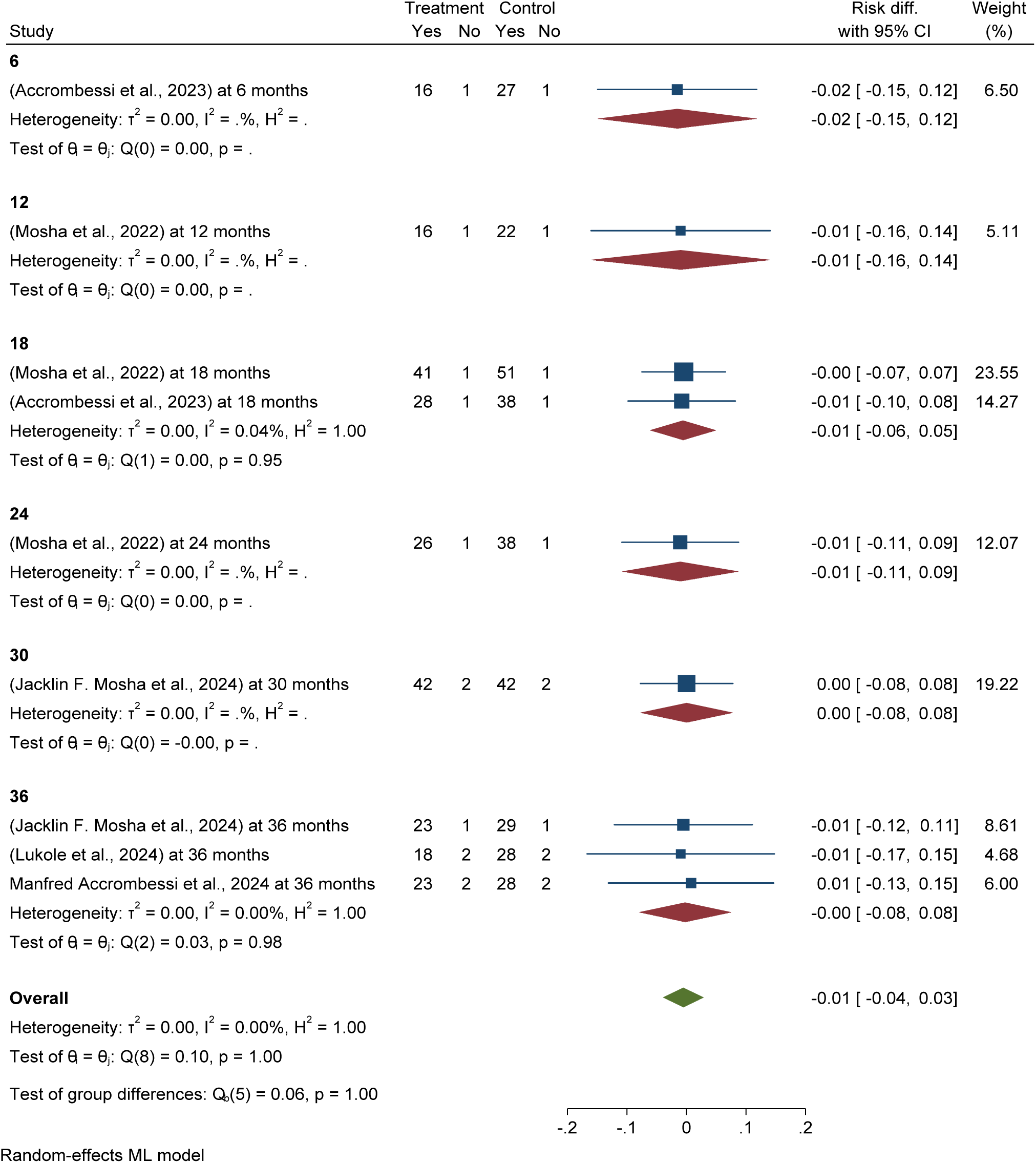
*Forest plot shows* subgroup analysis of post-intervention follow up effectiveness and efficacy of chlorfenapyr long-lasting insecticidal nets (LLINs) versus pyriproxyfen LLINs malaria infection reduction in Africa 2024

Publication bias was checked using funnel plots looking at symmetrical distribution, and it was objectively verified using Egger’s regression test, which revealed that there was no publication bias (p < 0.174) (Figure 87).

**Figure 87:**
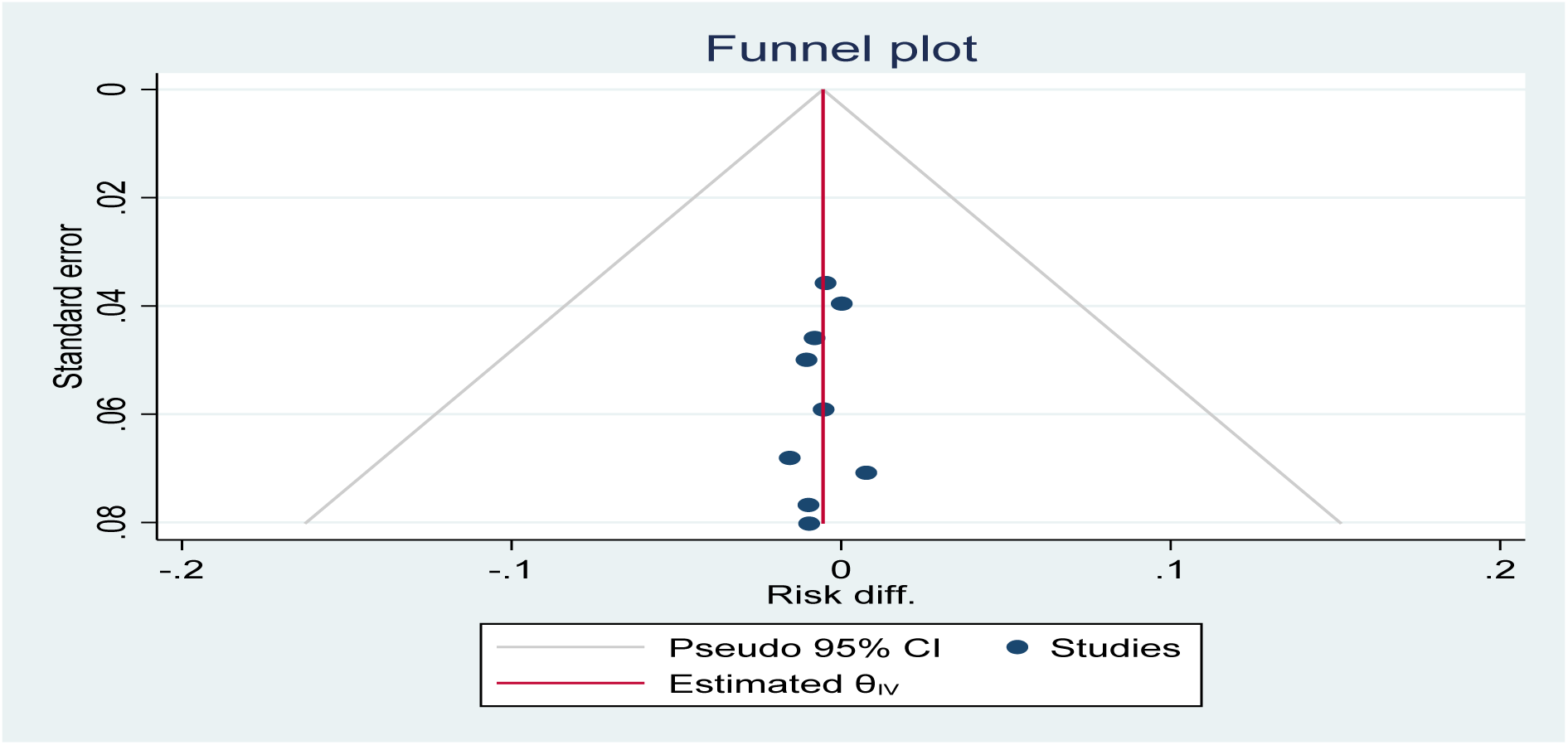
Funnel plot showing the distribution of included studies pooled malaria infection risk reduction among children using chlorfenapyr long-lasting insecticidal nets (LLINs) versus pyriproxyfen LLINs for malaria control in Africa in 2024.

The study found no difference in the effectiveness and efficacy of Piperonyl butoxide long-lasting insecticidal nets (LLINs) in reducing malaria infection risk in children over different durations compared to Pyriproxyfen LLINs *for malaria control( see figure 88)*.

**Figure 88:**
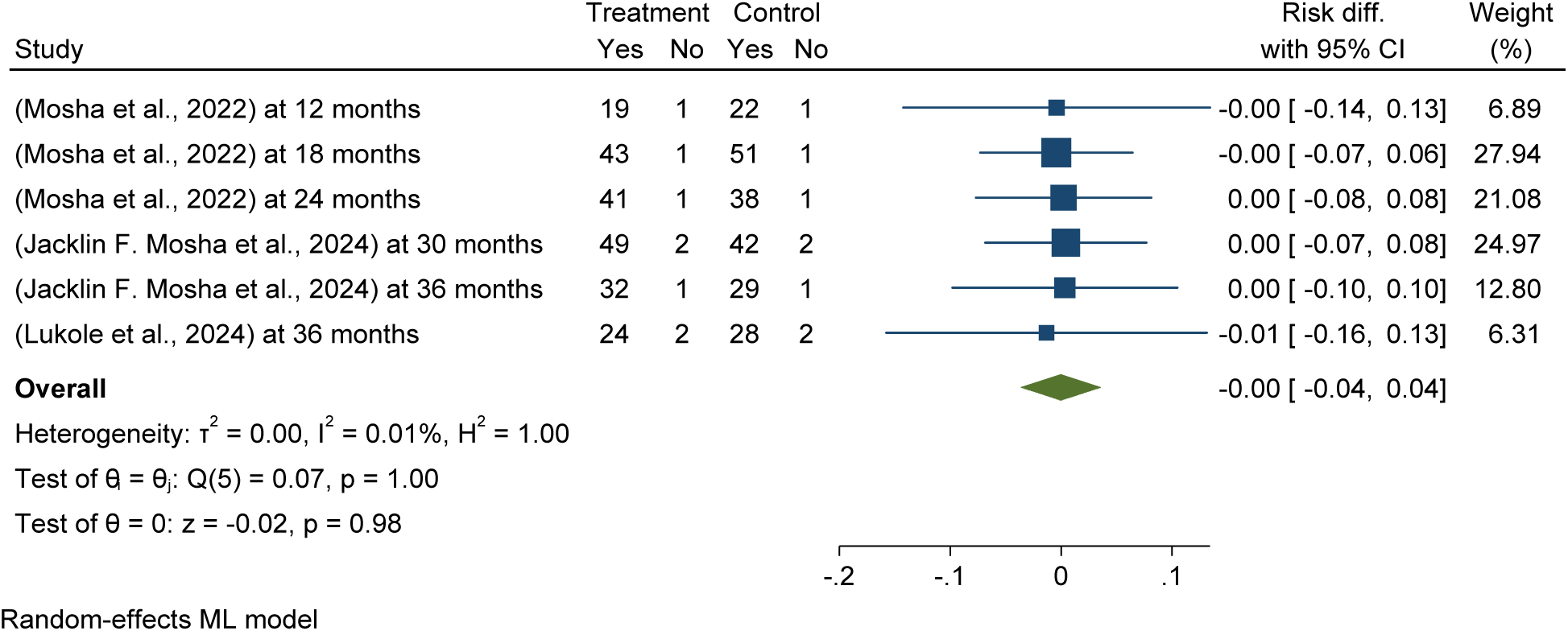
*Forest plot shows* pooled post-intervention effectiveness and efficacy of Piperonyl butoxide long-lasting insecticidal nets (LLINs) versus pyriproxyfen LLINs malaria infection reduction in Africa 2024

This meta-analysis determined entomological outcomes effectiveness and efficacy of chlorfenapyr versus pyriproxyfen LLINs, the forest plot shows that chlorfenapyr long-lasting insecticidal nets significantly reduce pooled mean indoor vector density per household per night by 1% compared to pyriproxyfen LLINs treatment in Africa. The included studies were homogeneous (I2 = 0.00%). See details in Figure 89.

**Figure 89:**
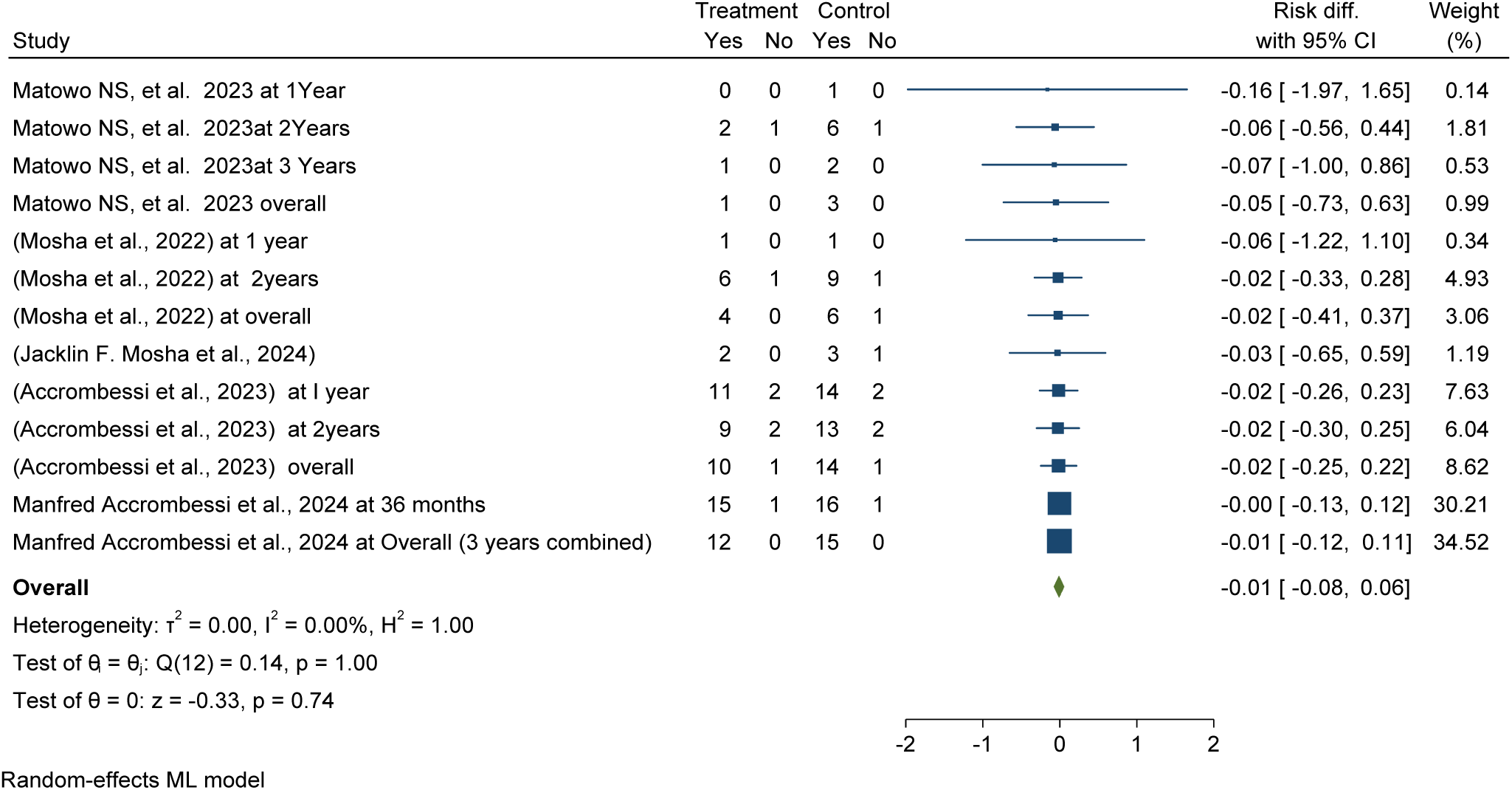
*Forest plot shows* pooled post-intervention effectiveness and efficacy of chlorfenapyr versus pyriproxyfen long-lasting insecticidal nets (LLINs) reduce mean indoor vector density per household per night in Africa 2024

Subgroup analysis with post-intervention follow-up effectiveness and efficacy of chlorfenapyr long-lasting insecticidal nets (LLINs) showed a significant reduction in pooled mean indoor vector density per household per night over different durations, with a 16% reduction at twelve months versus only 1% at 36 months post-distribution of LLINs, compared to pyriproxyfen LLINs (ARR = -0.01, 95% CI = -0.08, 0.06). See details in Figure 90.

**Figure 90:**
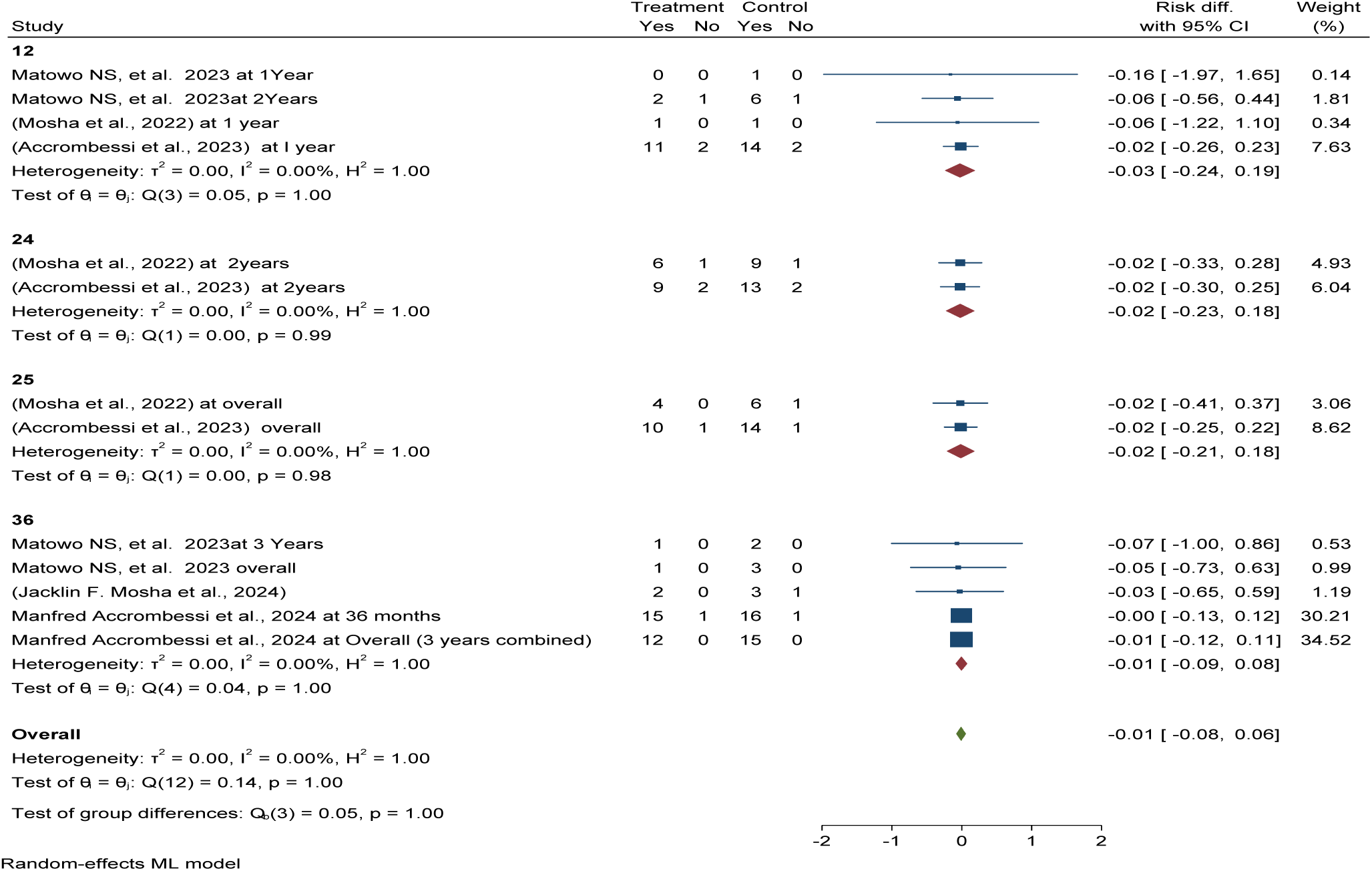
*Forest plot shows* subgroup analysis post-intervention follow-up effectiveness and efficacy of chlorfenapyr versus pyriproxyfen long-lasting insecticidal nets (LLINs) reduce mean indoor vector density per household per night in Africa 2024

Publication bias was checked using funnel plots looking at symmetrical distribution, and it was objectively verified using Egger’s regression test, which revealed that there was no publication bias (p < 0.272) (Figure 91).

**Figure 91:**
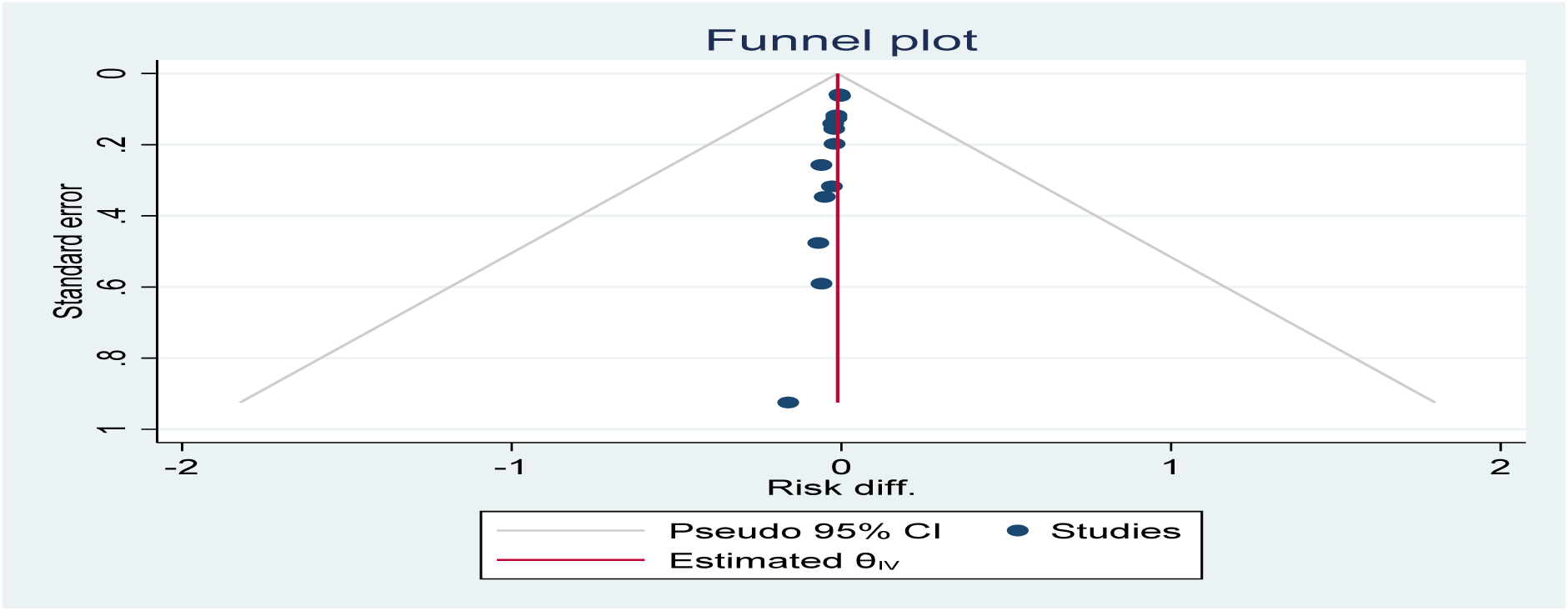
Funnel plot showing the distribution of included studies pooled mean indoor vector density per household per night among using chlorfenapyr long-lasting insecticidal nets (LLINs) versus pyriproxyfen LLINs for malaria control in Africa in 2024

This meta-analysis determined the entomological outcomes, effectiveness, and efficacy of Piperonyl butoxide versus pyriproxyfen long-lasting insecticidal nets (LLINs). The forest plot shows that Piperonyl butoxide long-lasting insecticidal nets significantly reduce indoor vector density per household per night by 4% compared to pyriproxyfen LLINs treatment in Africa. The included studies were homogeneous (I2 = 0.00%). See details in Figure 92.

**Figure 92:**
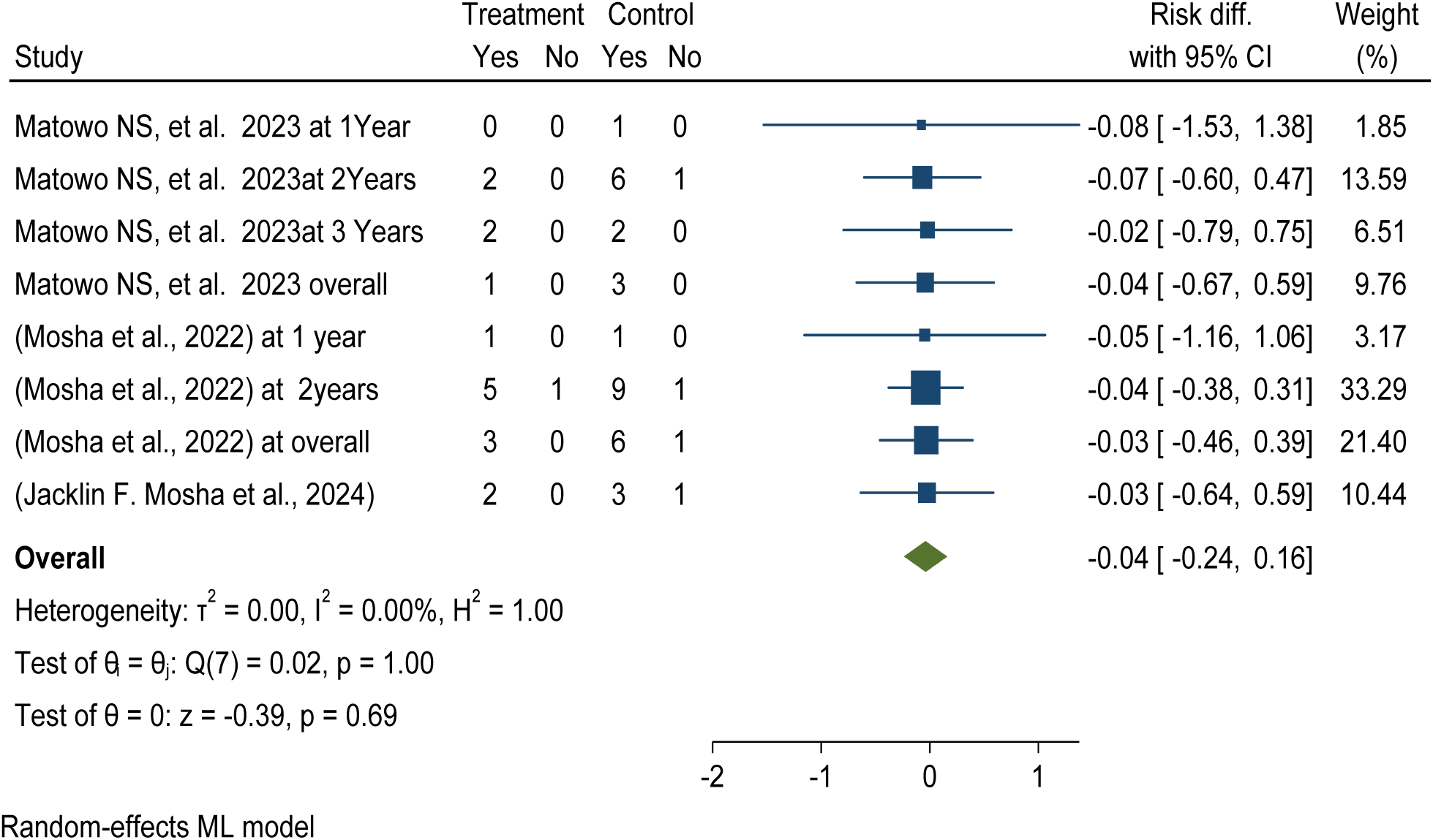
*Forest plot shows* post-intervention effectiveness and efficacy of Piperonyl butoxide versus pyriproxyfen long-lasting insecticidal nets (LLINs) reduce mean indoor vector density per household per night in Africa 2024

Subgroup analysis with post-intervention follow-up effectiveness and efficacy of Piperonyl butoxide long-lasting insecticidal nets (LLINs) showed a significant reduction in pooled mean indoor vector density per household per night over different durations, with a 8% reduction at twelve months versus only 4% at 36 months post-distribution of LLINs, compared to pyriproxyfen LLINs (ARR = -0.01, 95% CI = -0.24, 0.16). See details in Figure 93.

**Figure 93:**
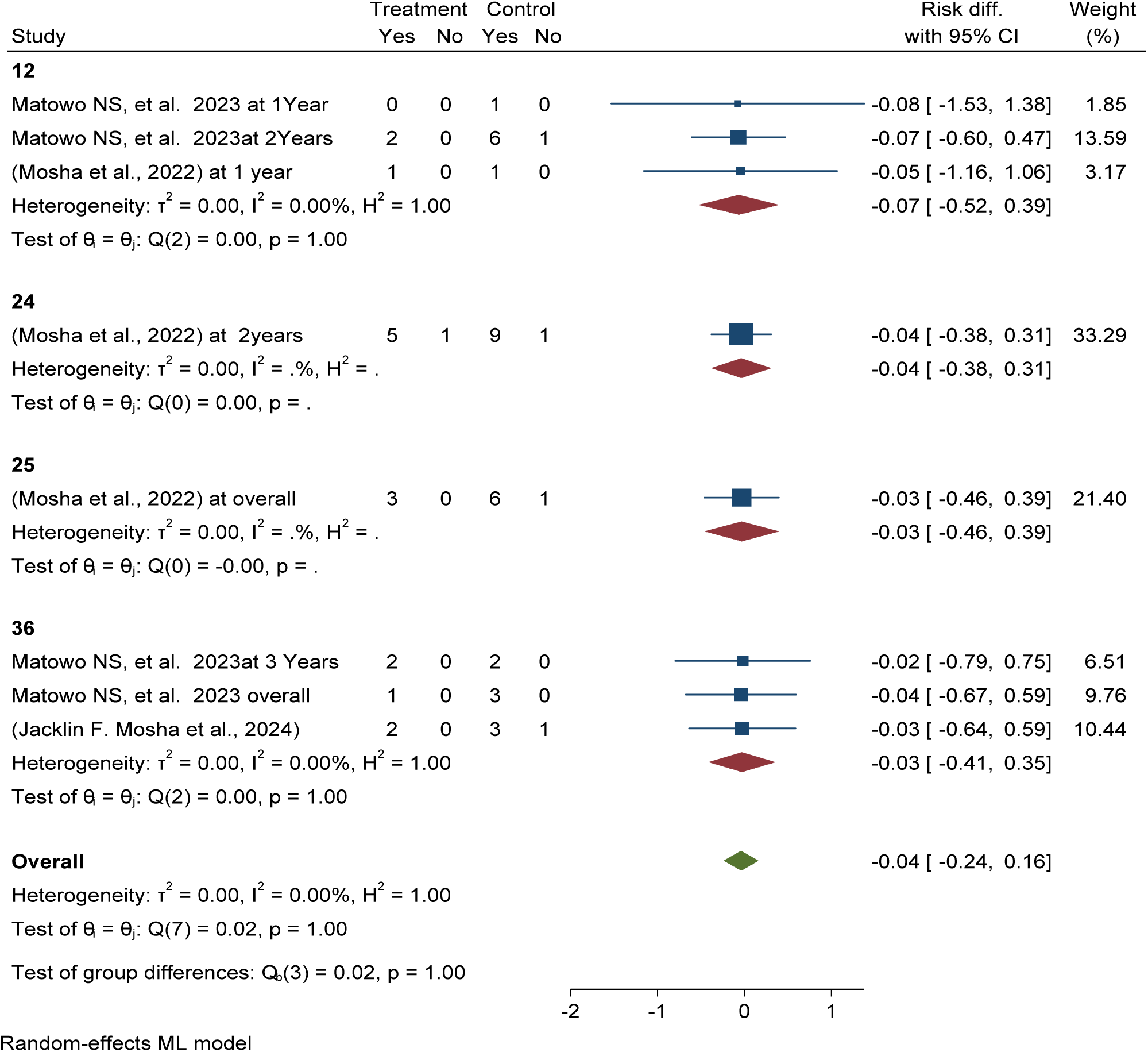
*Forest plot shows* subgroup analysis post-intervention follow-up effectiveness and efficacy of Piperonyl butoxide versus pyriproxyfen long-lasting insecticidal nets (LLINs) reduce mean indoor vector density per household per night in Africa 2024

Publication bias was checked using funnel plots looking at symmetrical distribution, and it was objectively verified using Egger’s regression test, which revealed that there was no publication bias (p < 0.974) (Figure 94).

**Figure 94:**
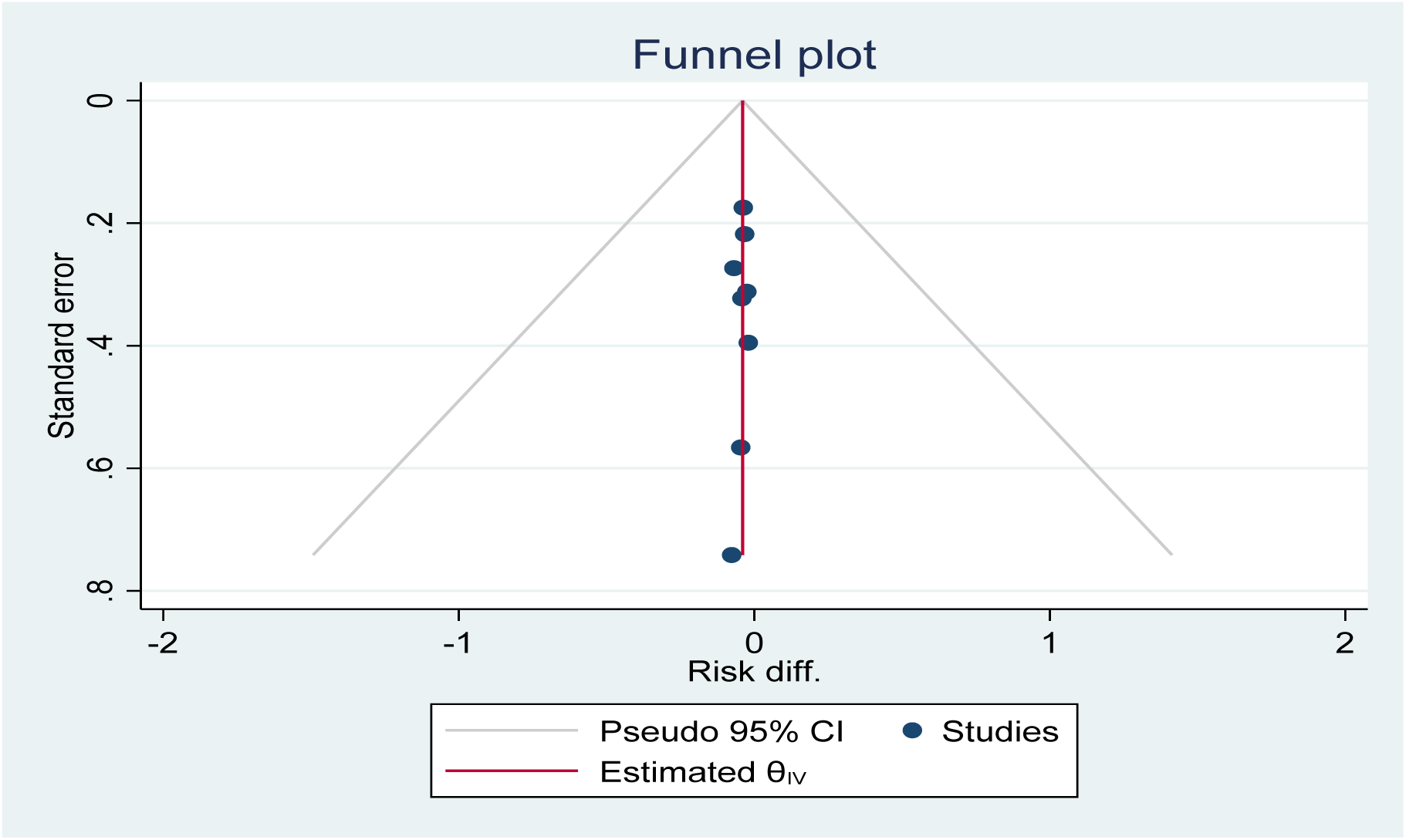
*Funnel plot showing the distribution of included studies pooled mean indoor vector density per household per night among using* Piperonyl butoxide *long-lasting insecticidal nets (LLINs) versus pyriproxyfen LLINs for malaria control in Africa in 2024*

The study reveals that chlorfenapyr LLINs effectively reduce sporozoite rate by 7% compared to pyriproxyfen LLINs, demonstrating its superior efficacy in long-lasting insecticidal nets (LLINs) in Figure 95.

**Figure 95:**
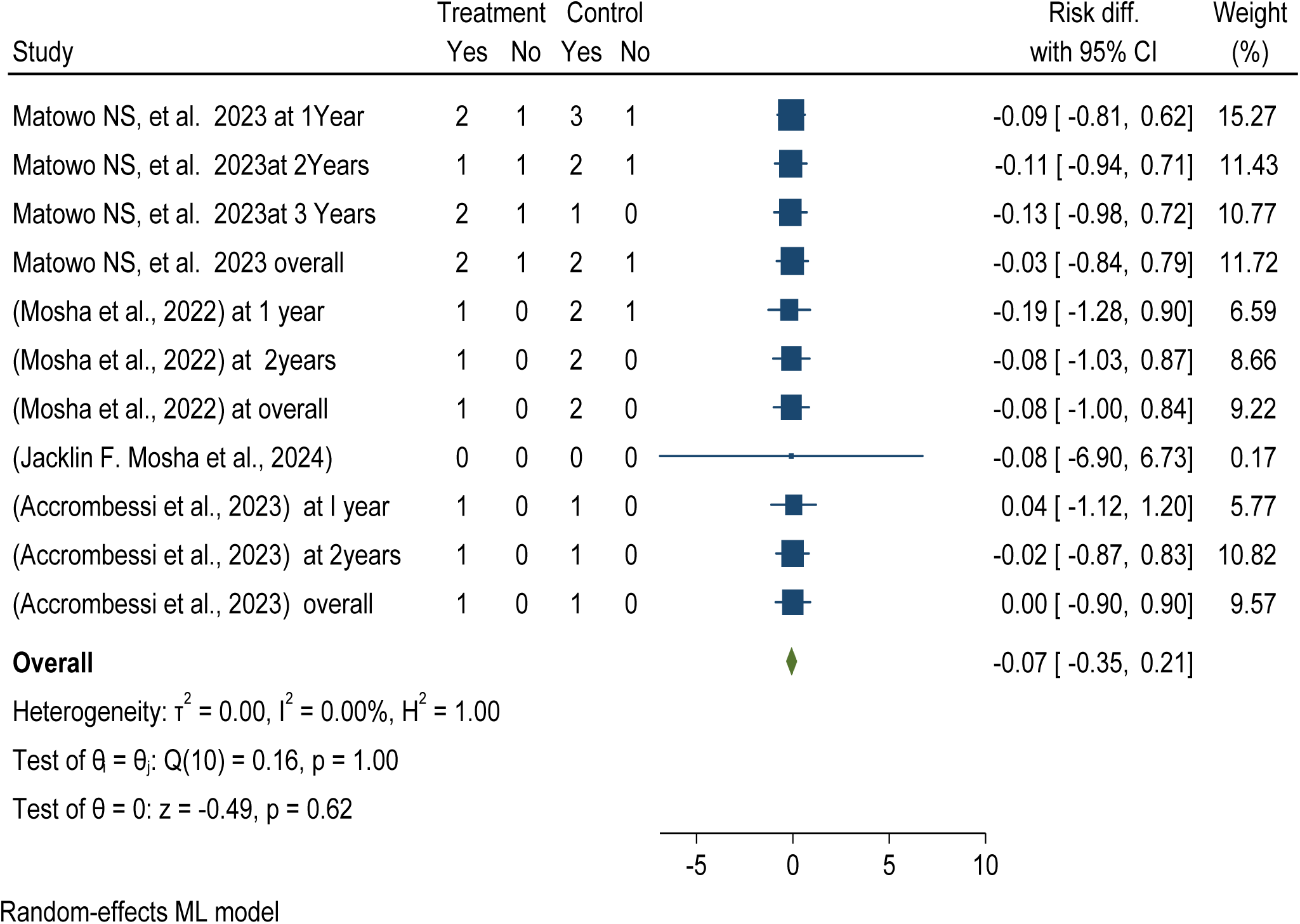
*Forest plot shows* pooled post-intervention effectiveness and efficacy of chlorfenapyr versus pyriproxyfen long-lasting insecticidal nets (LLINs) reduce pooled sporozoite rate in Africa 2024

The subgroup analysis based on a follow-up duration study analyzing the effectiveness and efficacy of chlorfenapyr long-lasting insecticidal nets (LLINs) showed a significant reduction in pooled sporozoite rate per anopheles mosquito over different durations, with a 10% reduction at twelve months and 8% at 36 months post-distribution compared to pyriproxyfen LLINs. See details in Figure 96

**Figure 96:**
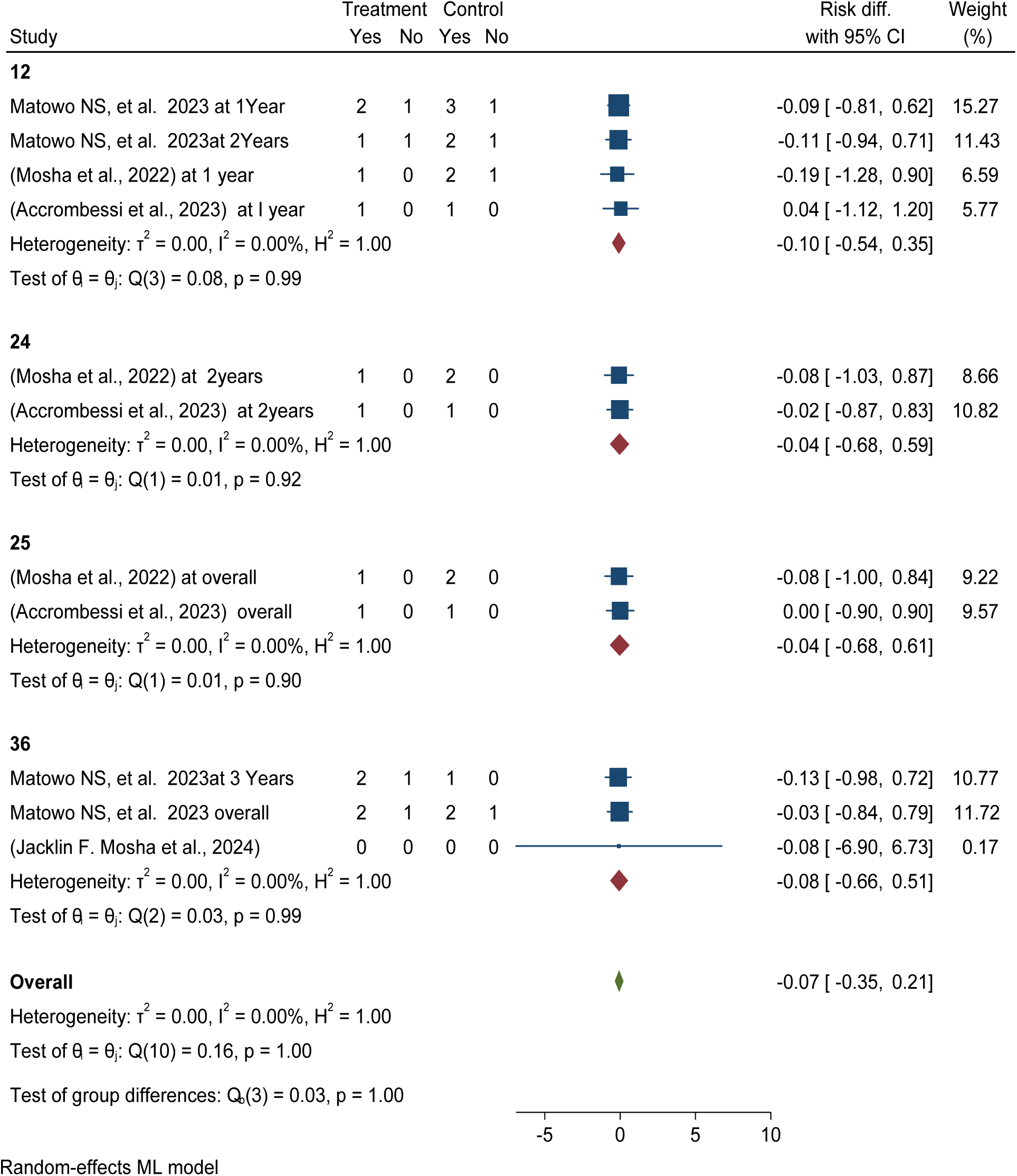
*Forest plot shows* subgroup analysis post-intervention follow-up pooled post-intervention effectiveness and efficacy of chlorfenapyr versus pyriproxyfen long-lasting insecticidal nets (LLINs) reduce pooled sporozoite rate in Africa 2024

Publication bias was checked using funnel plots looking at symmetrical distribution, and it was objectively verified using Egger’s regression test, which revealed that there was no publication bias (p < 0.293) (Figure 97).

**Figure 97:**
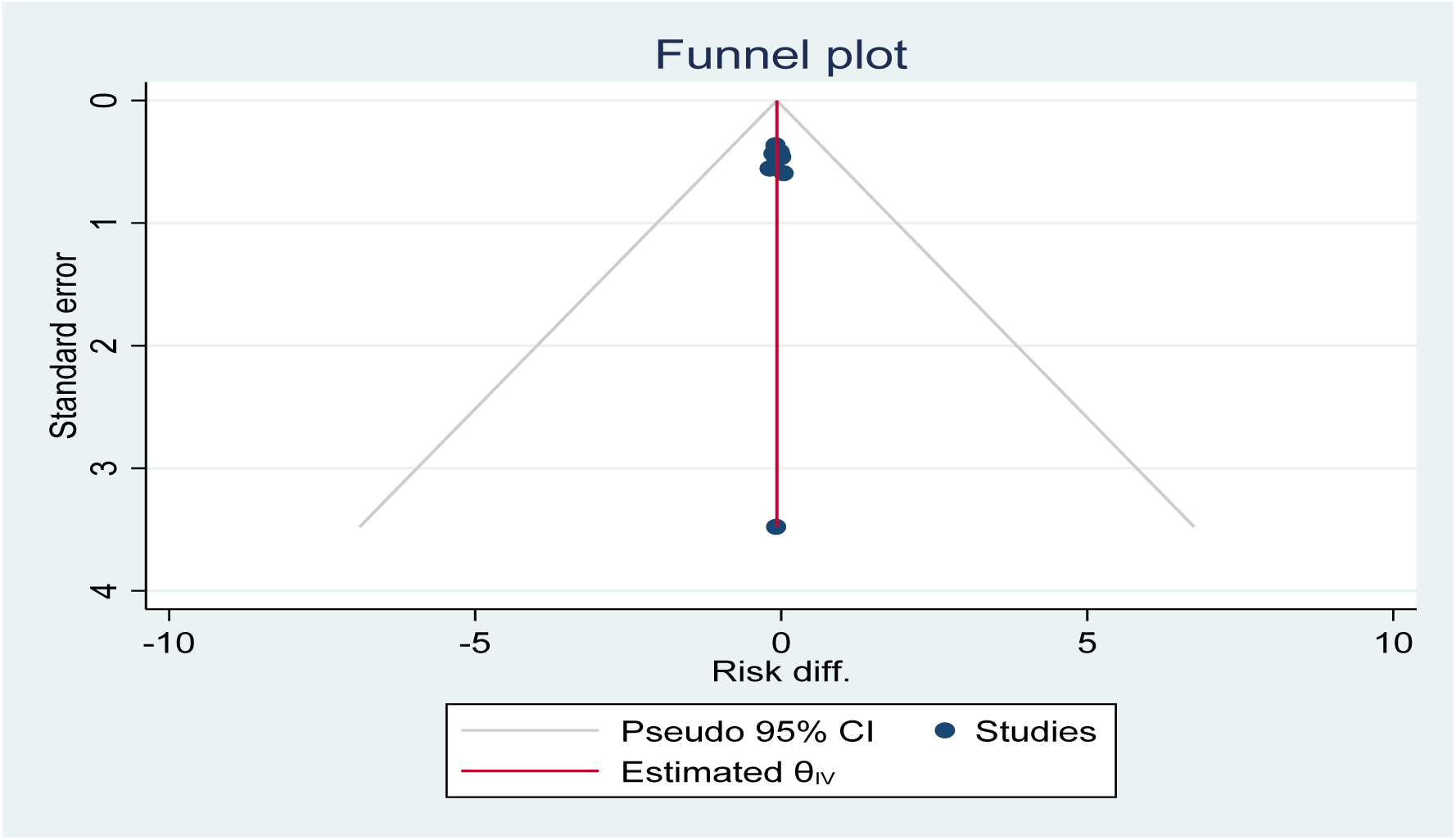
Funnel plot showing the distribution of included studies pooled sporozoite rate among using piperonyl butoxide long-lasting insecticidal nets (LLINs) versus pyriproxyfen LLINs for malaria control in Africa in 2024.

The study reveals that Piperonyl butoxide long-lasting insecticidal nets (LLINs) effectively and efficaciously reduce sporozoite rate by 1% compared to pyriproxyfen LLINs in Figure 98.

**Figure 98:**
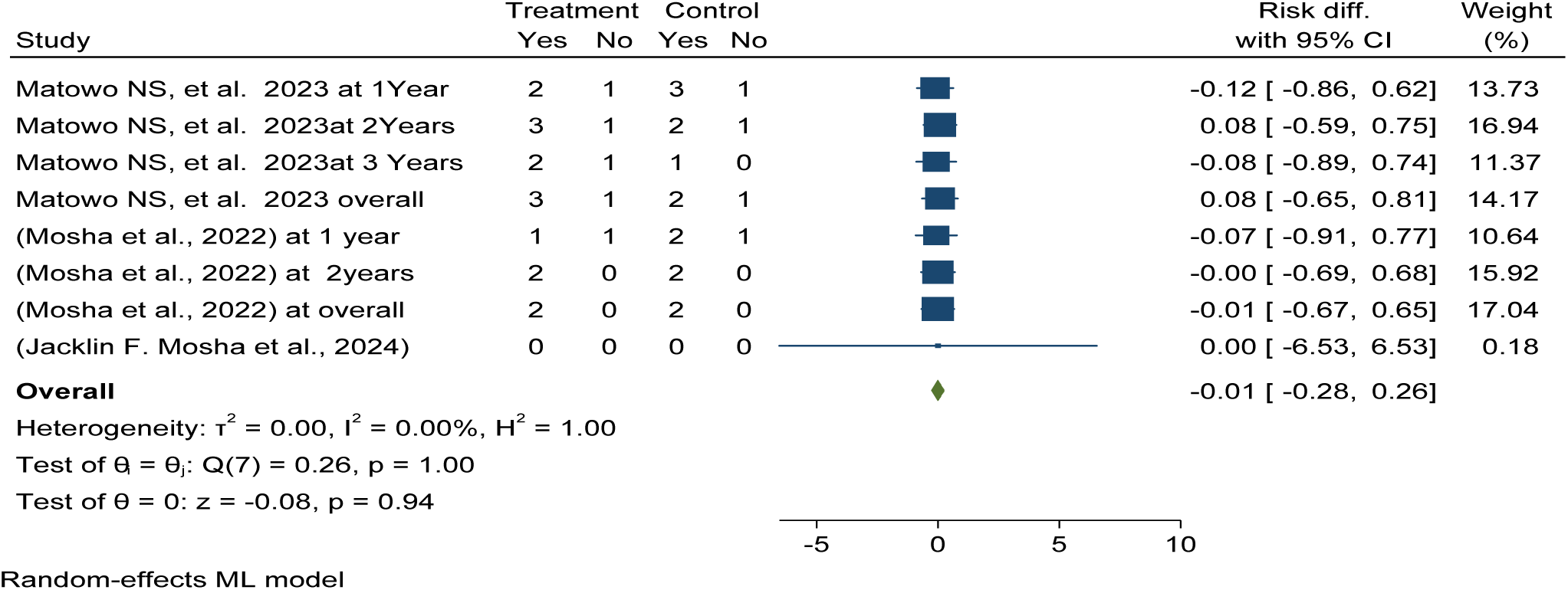
*Forest plot shows* pooled post-intervention effectiveness and efficacy of Piperonyl butoxid versus pyriproxyfen long-lasting insecticidal nets (LLINs) reduce pooled sporozoite rate in Africa 2024

The subgroup analysis based on a follow-up duration study analyzing the effectiveness and efficacy of Piperonyl butoxide long-lasting insecticidal nets (LLINs) showed a significant reduction in pooled sporozoite rate per anopheles mosquito over different durations, with a 3% reduction at twelve months and 1% at 36 months post-distribution compared to pyriproxyfen LLINs. See details in Figure 99.

**Figure 99:**
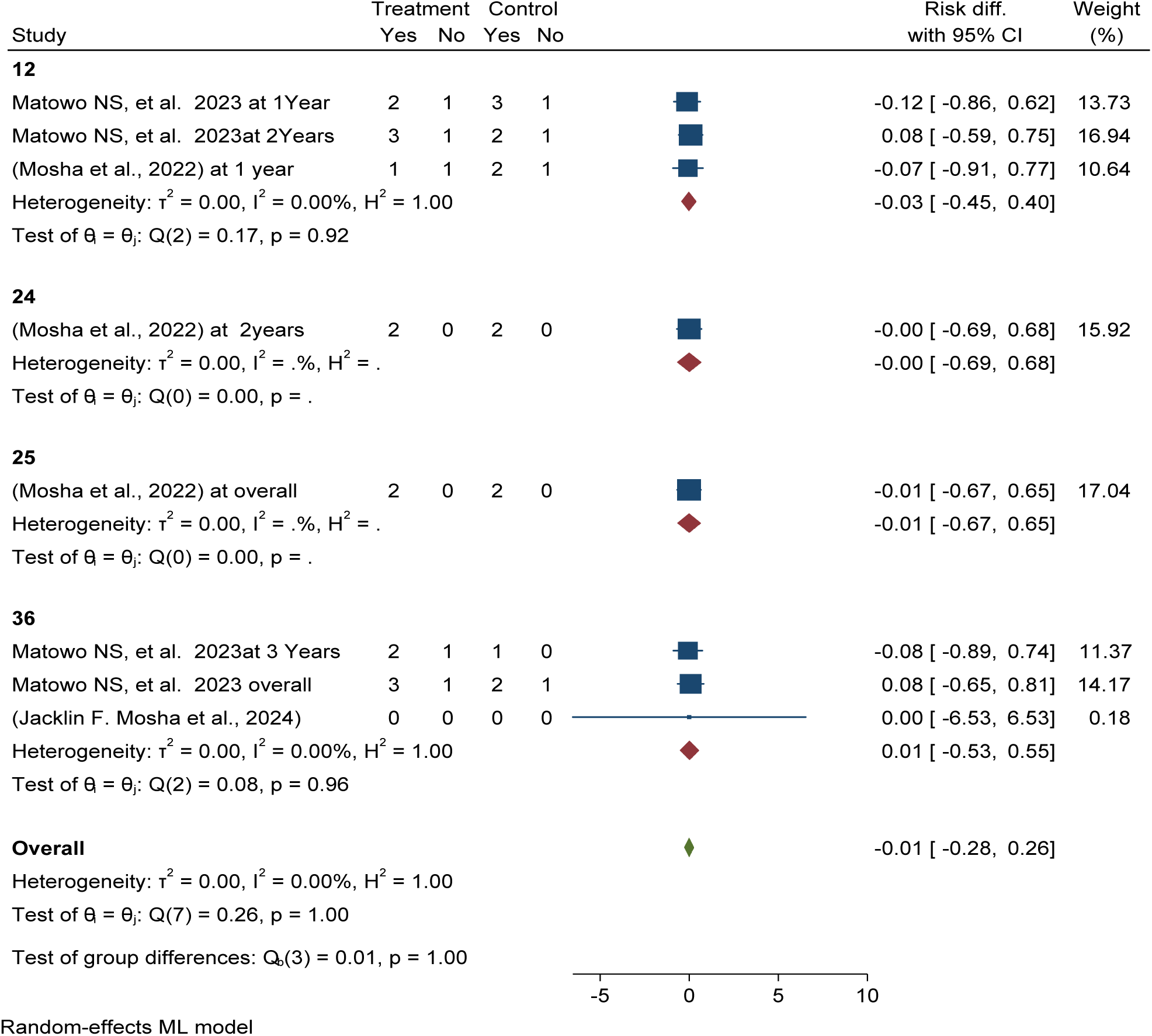
*Forest plot shows* subgroup analysis of post-intervention follow-up effectiveness and efficacy of Piperonyl butoxid versus pyriproxyfen long-lasting insecticidal nets (LLINs) reduce pooled sporozoite rate in Africa 2024

The study reveals that chlorfenapyr LLINs effectively reduce mean entomological inoculations rate per household per night by 15% compared to pyriproxyfen LLINs, demonstrating its superior efficacy in mean entomological incubations rate reduction see details in Figure 100.

**Figure 100:**
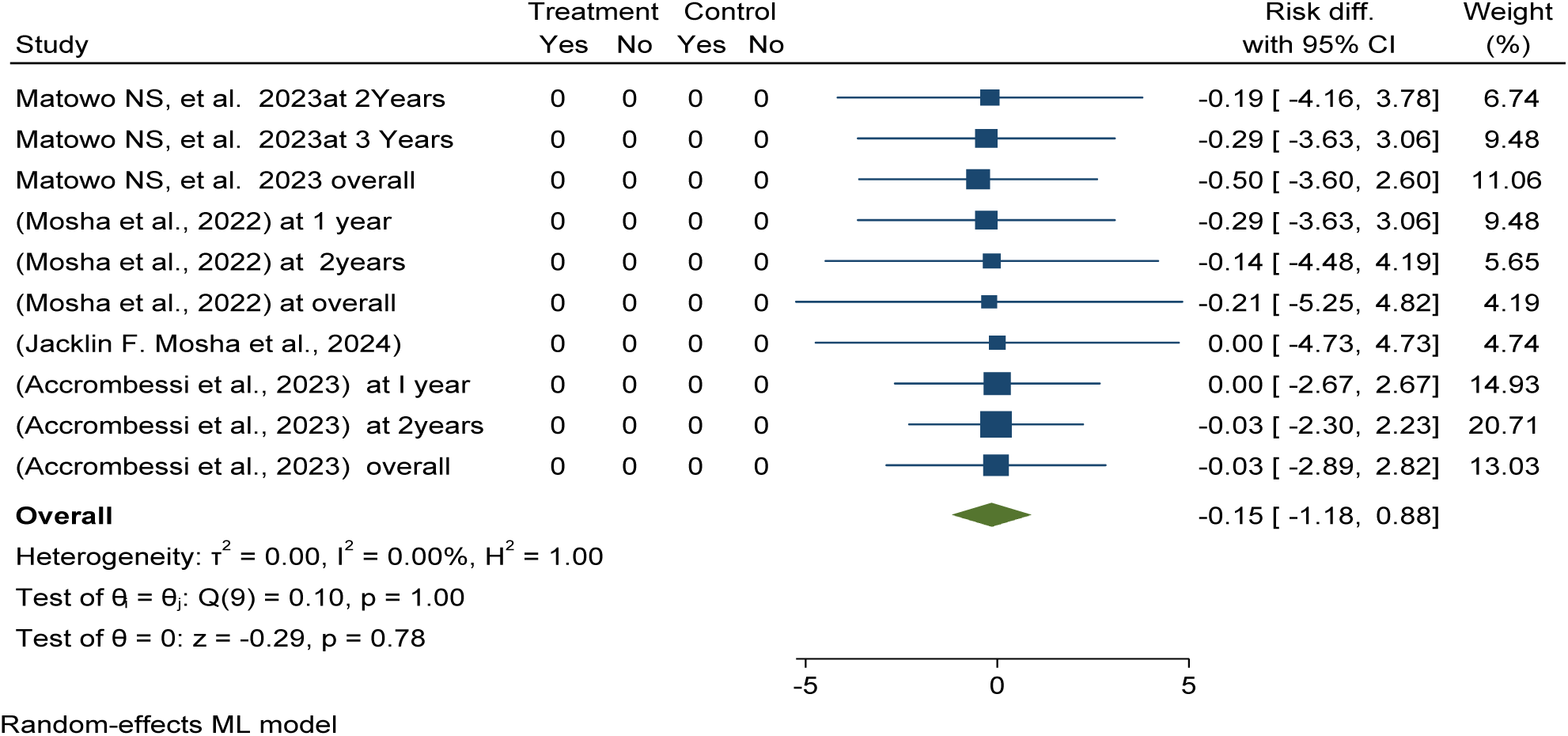
*Forest plot shows* pooled post-intervention effectiveness and efficacy of chlorfenapyr versus pyriproxyfen long-lasting insecticidal nets (LLINs) reduce pooled mean entomological inoculation rate per household per night in Africa 2024

The subgroup analysis based on a follow-up duration study analyzing the effectiveness and efficacy of chlorfenapyr long-lasting insecticidal nets (LLINs) showed a significant reduction in pooled mean entomological inoculation rate per household per night over different durations, with a 13% reduction at twelve months and 33% at 36 months post-distribution compared to pyriproxyfen LLINs. See details in Figure 101.

**Figure 101:**
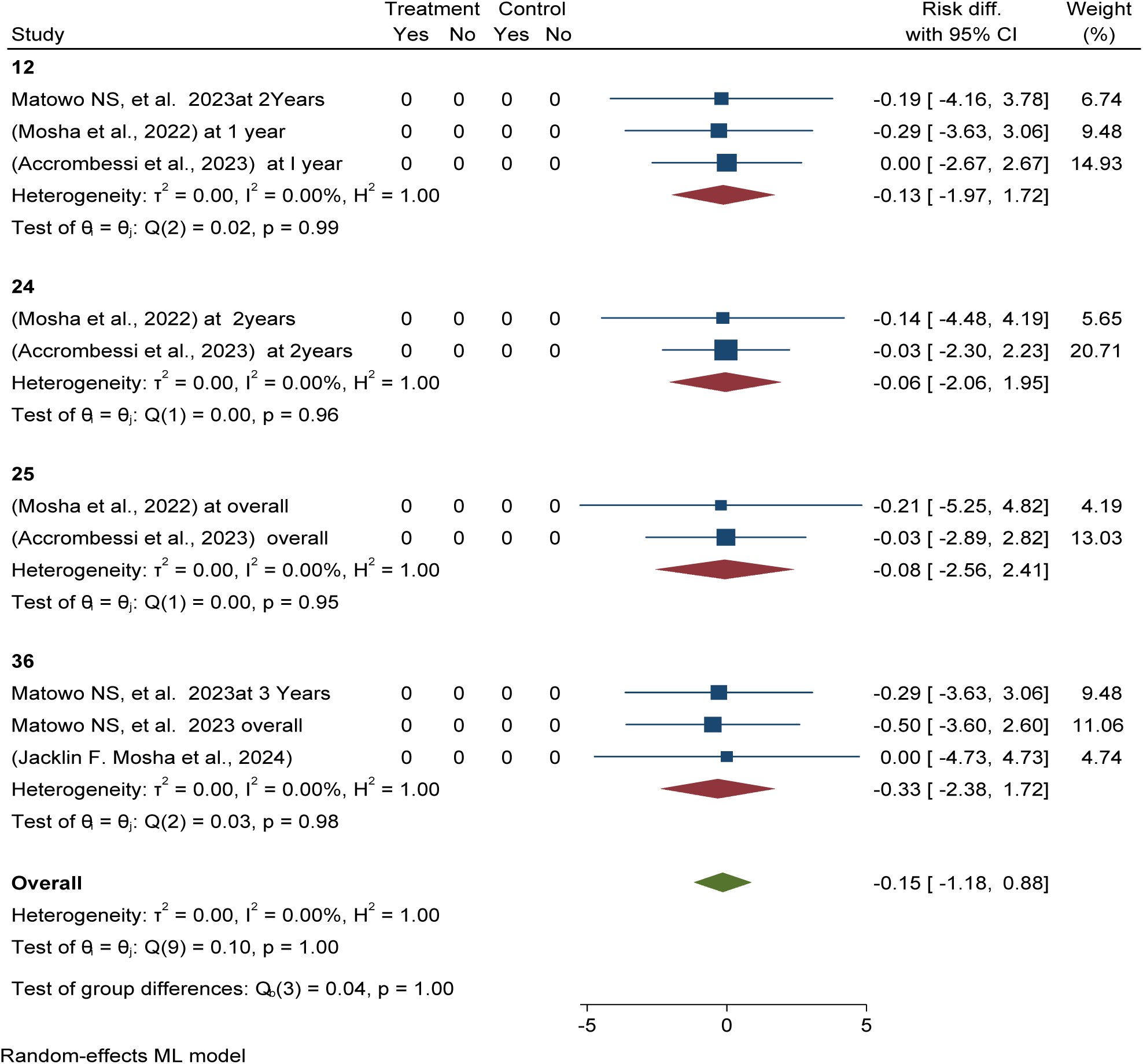
*Forest plot shows* subgroup analysis of post-intervention follow-up effectiveness and efficacy of chlorfenapyr versus pyriproxyfen long-lasting insecticidal nets (LLINs) reduce pooled mean entomological inoculation rate per household per night in Africa 2024

Publication bias was checked using funnel plots looking at symmetrical distribution, and it was objectively verified using Egger’s regression test, which revealed that there was no publication bias (p < 0.38) (Figure 102).

**Figure 102:**
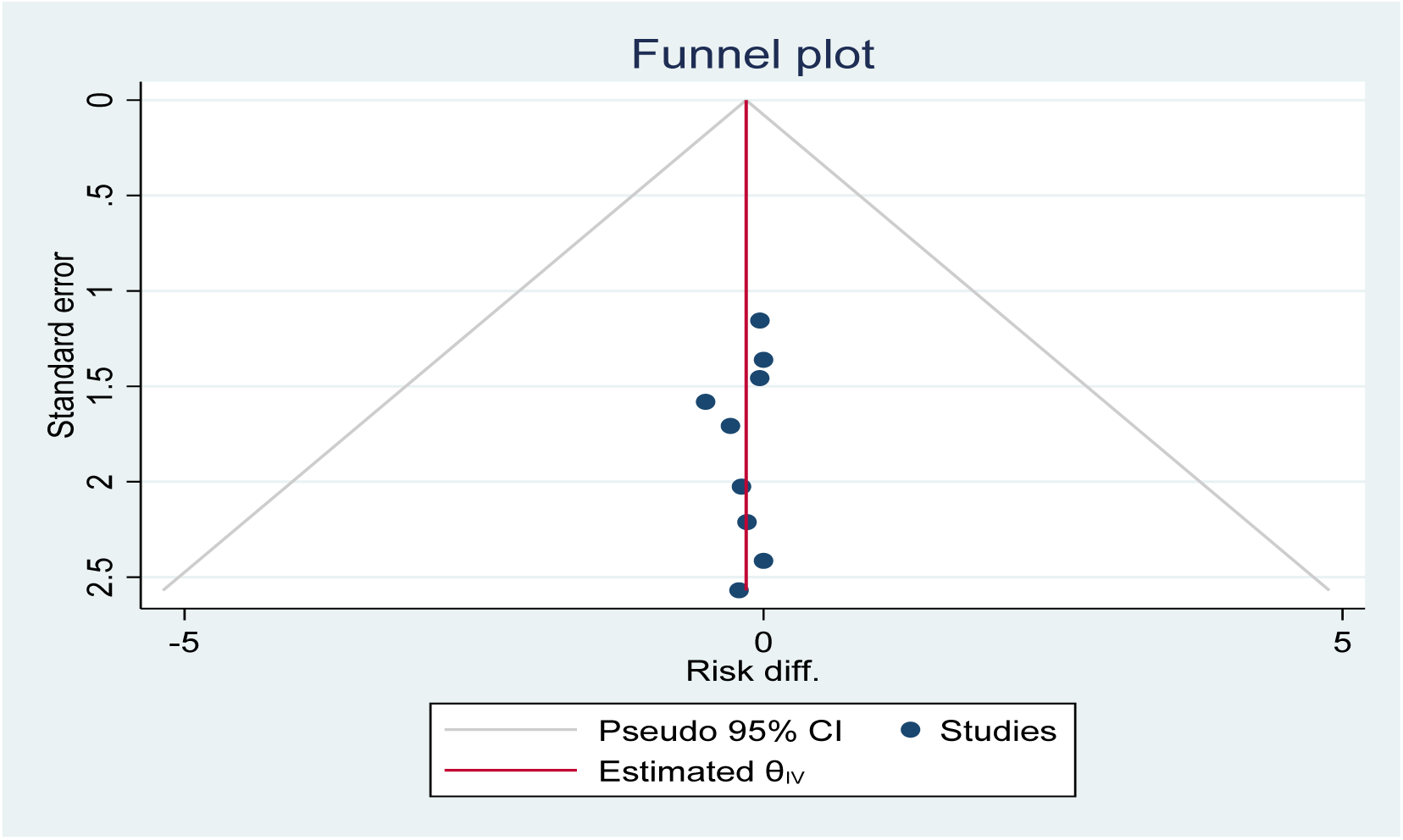
Funnel plot showing the distribution of included studies pooled mean entomological inoculation rate per household per night among using chlorfenapyr long-lasting insecticidal nets (LLINs) versus pyriproxyfen LLINs for malaria control in Africa in 2024.

The study reveals that Piperonyl butoxide LLINs effectively reduce mean entomological inoculations rate per household per night by 5% compared to pyriproxyfen LLINs, demonstrating its superior efficacy in mean entomological inoculations rate reduction see details in Figure 103.

**Figure 103:**
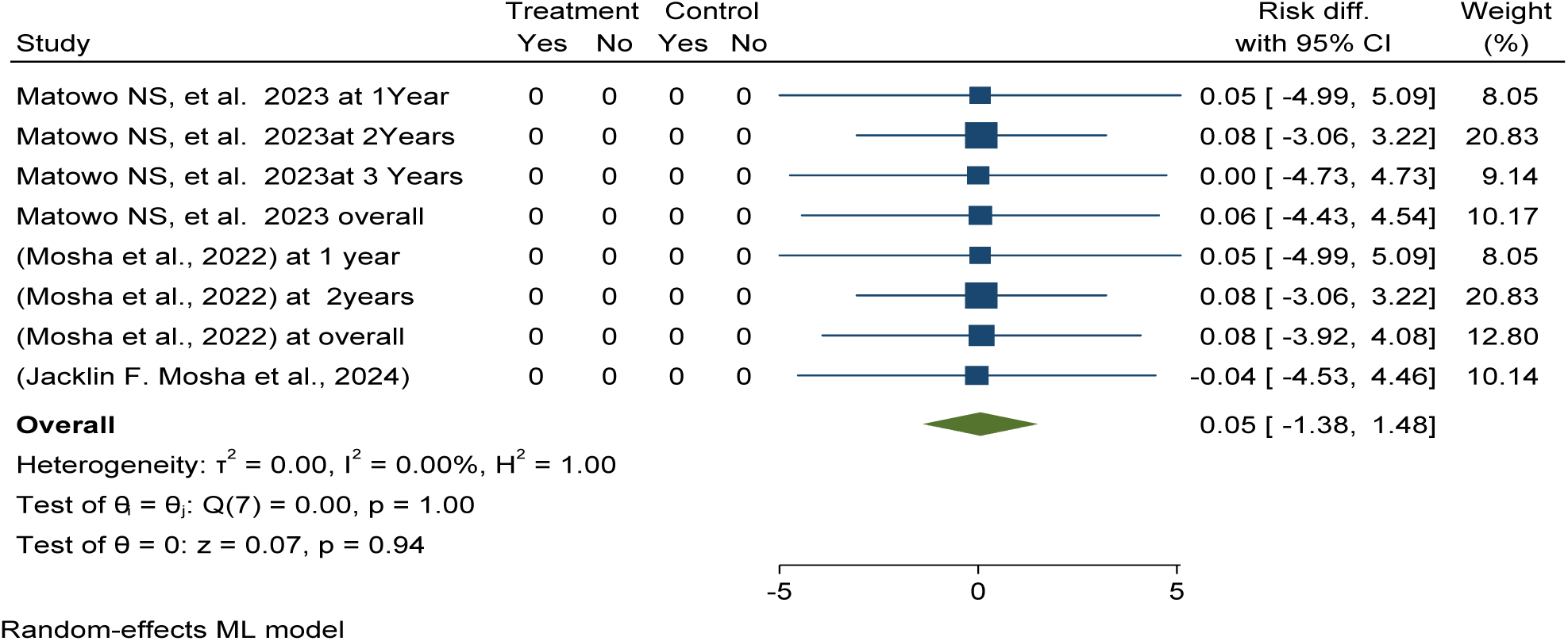
*Forest plot shows* pooled post-intervention effectiveness and efficacy of Piperonyl butoxide versus pyriproxyfen long-lasting insecticidal nets (LLINs) reduce pooled mean entomological inoculation rate per household per night in Africa 2024

The subgroup analysis based on a follow-up duration study analyzing the effectiveness and efficacy of Piperonyl butoxide long-lasting insecticidal nets (LLINs) showed a significant reduction in entomological inoculation rate per household per night over different durations, with a 7% reduction at twelve months and 1% at 36 months post-distribution compared to pyriproxyfen LLINs. See details in Figure 104.

**Figure 104:**
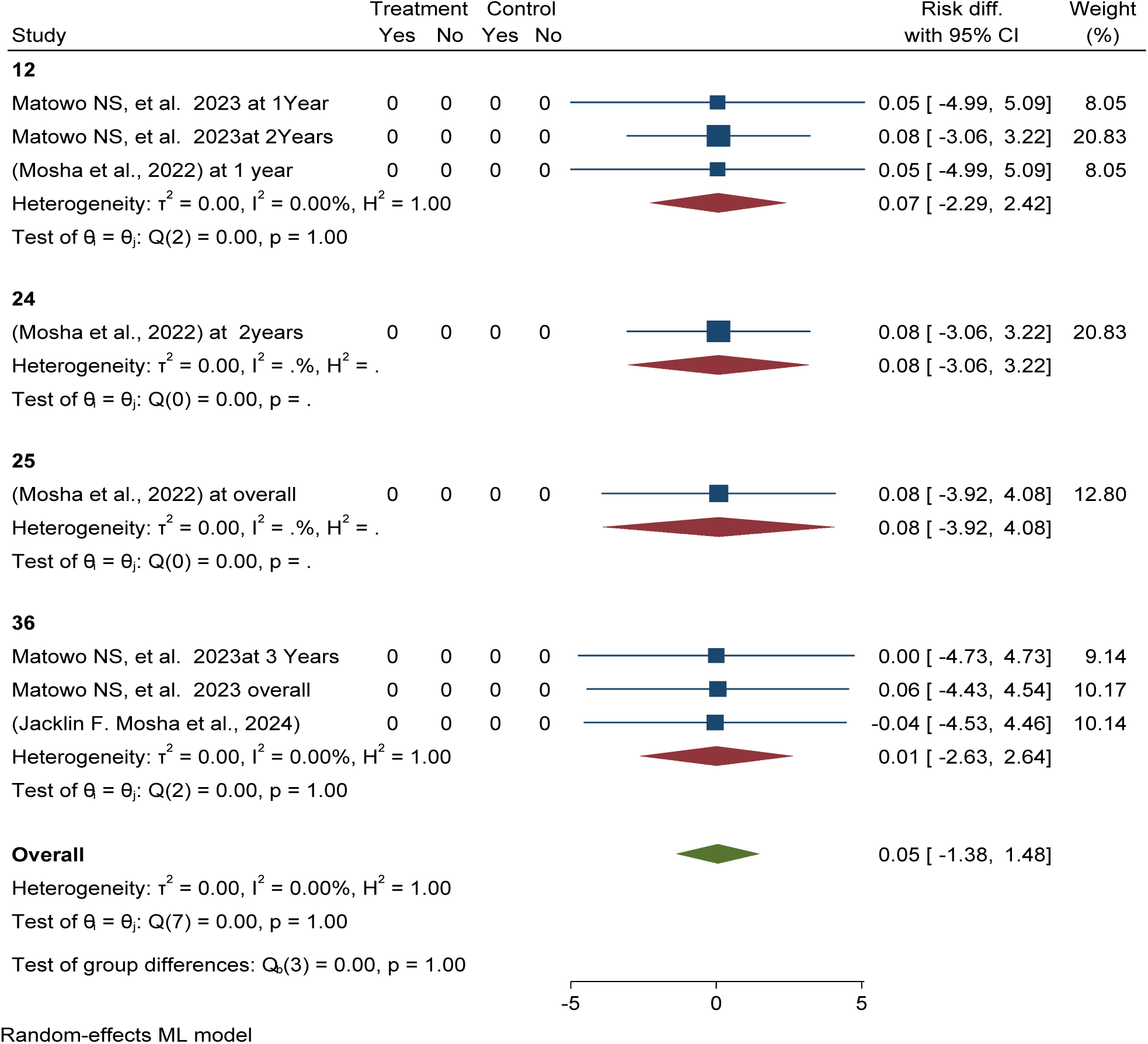
*Forest plot shows* subgroup analysis of post-intervention follow-up effectiveness and efficacy of Piperonyl butoxide versus pyriproxyfen long-lasting insecticidal nets (LLINs) reduce pooled mean entomological inoculation rate per household per night in Africa 2024

## Malaria incidence CFP VS PPF

This study evaluated the post-intervention effectiveness and efficacy of chlorfenapyr long-lasting insecticidal nets (LLINs) in malaria case incidence reduction in children aged 6 months to 10 years over different durations, one year to 2 years, and overall reduction by 1% as compared to pyriproxyfen LLINs (ARR = -0.01, 95% CI = -0.19, 0.17). See details in Figure 105.

**Figure 105:**
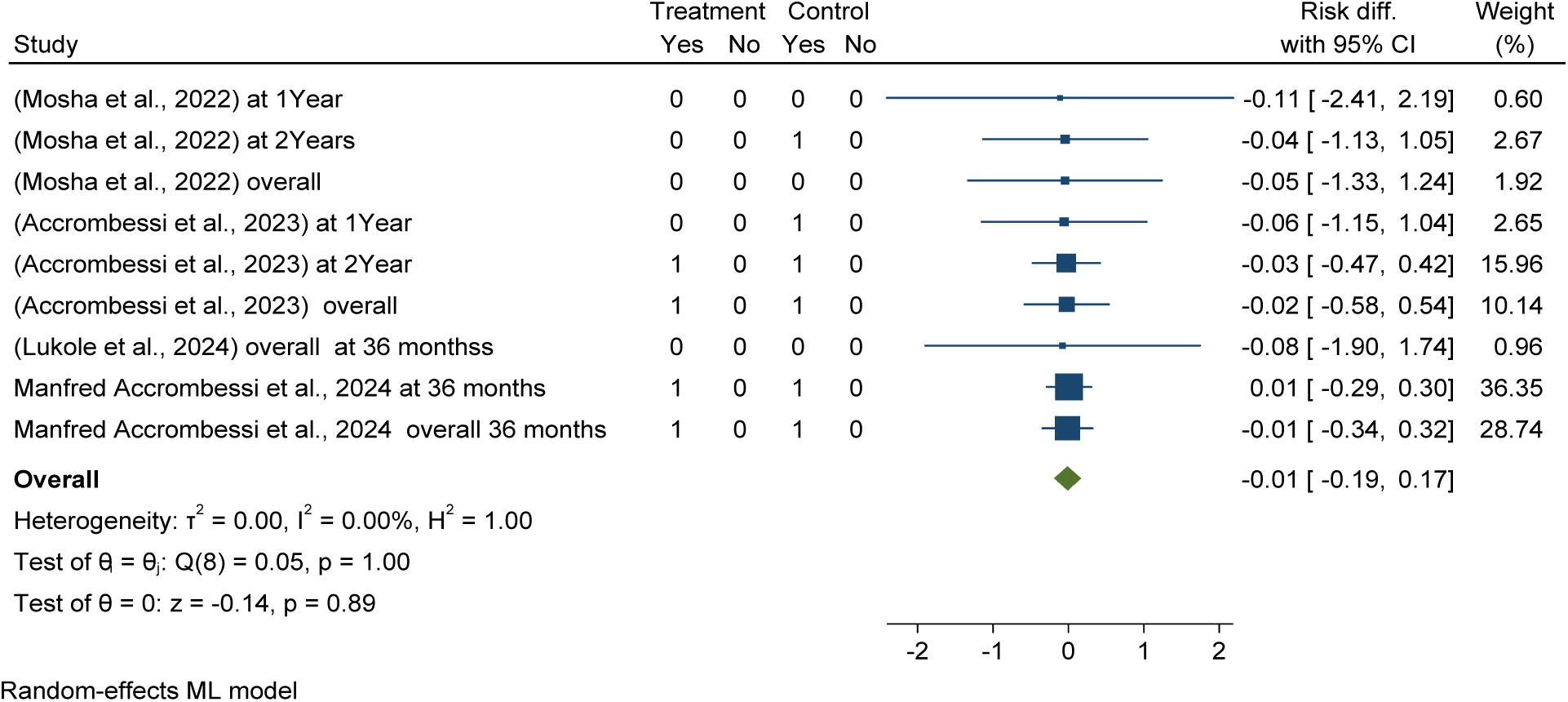
*Forest plot shows* post-intervention effectiveness and efficacy of chlorfenapyr long-lasting insecticidal nets (LLINs) in malaria case incidence reduction in children aged 6 months to 10 years over different durations in Africa 2024.

The subgroup analysis based on a follow-up duration study analyzing the effectiveness and efficacy of chlorfenapyr long-lasting insecticidal nets (LLINs) showed a significant reduction in entomological inoculationrate per household per night over different durations, with a 7% reduction at twelve months and 3% at 25 months post-distribution compared to pyriproxyfen LLINs. See details in Figure 106.

**Figure 106:**
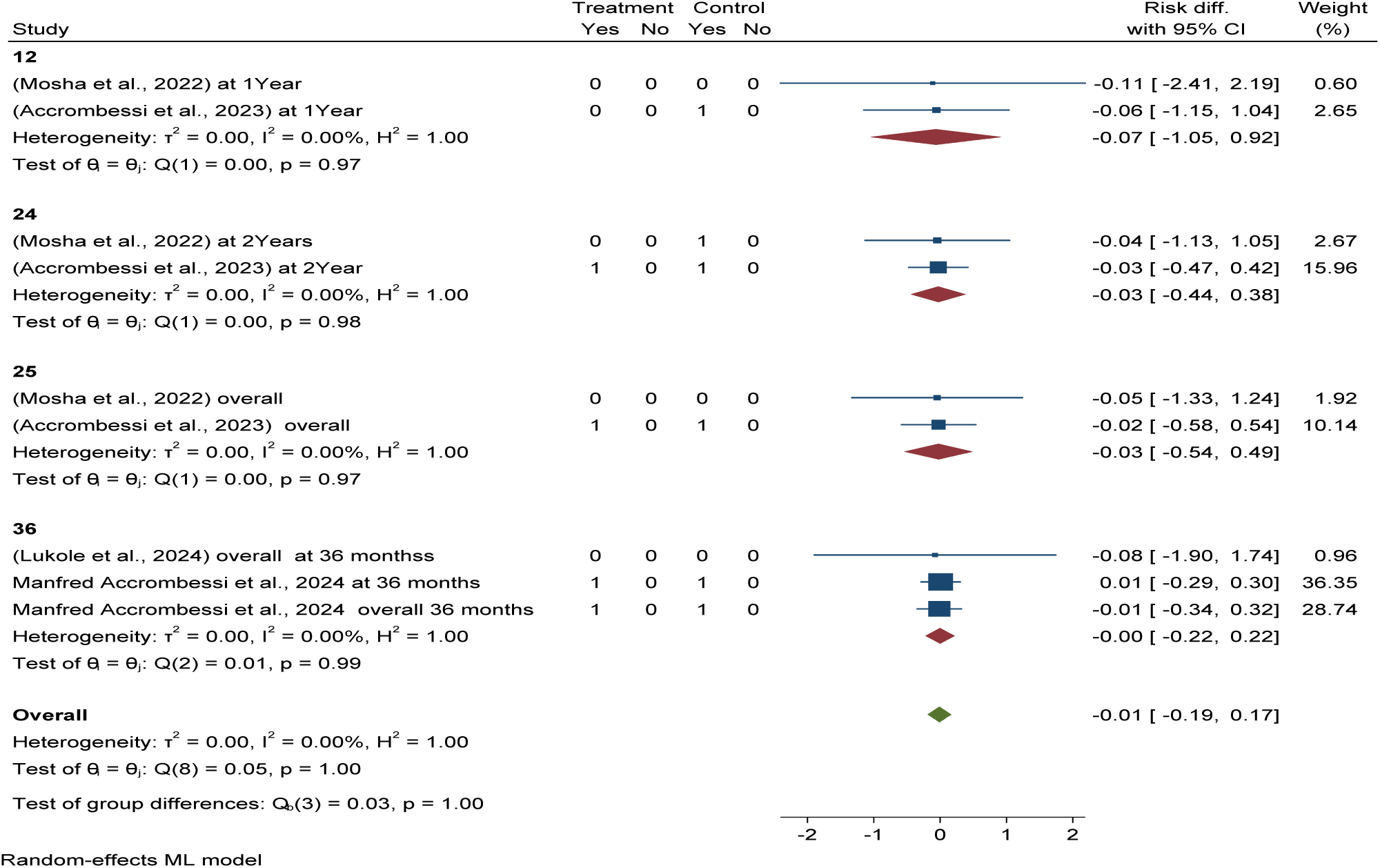
*Forest plot shows* subgroup analysis of post-intervention follow-up effectiveness and efficacy of chlorfenapyr long-lasting insecticidal nets (LLINs) in malaria case incidence reduction in children aged 6 months to 10 years over different durations in Africa 2024.

## Malaria incidence PBO VS PPF

This study determined that post intervention effectiveness and efficacy of piperonyl butoxide long-lasting insecticidal nets (LLINs) in malaria case incidence reduction in children aged 6 months to 10 years over different durations, one year to 2 years, and overall reduced by 2% as compared to pyriproxyfen LLINs (ARR = -0.02, 95% CI = -0.57, 0.54). See details in Figure 107.

**Figure 107:**
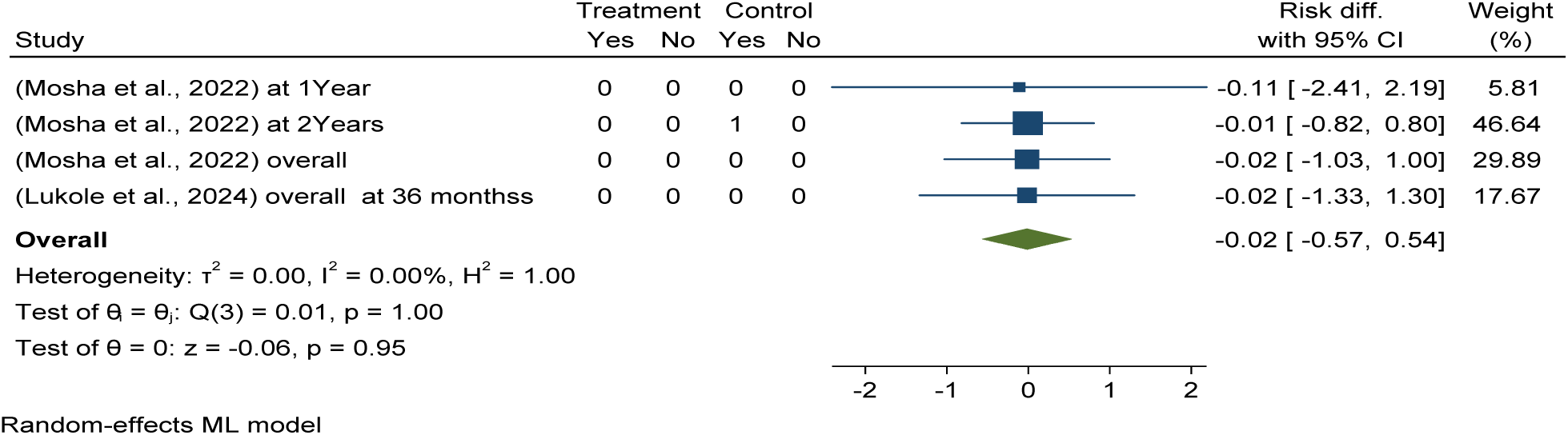
*Forest plot shows* post-intervention follow-up effectiveness and efficacy of piperonyl butoxide long-lasting insecticidal nets (LLINs) in malaria case incidence reduction in children aged 6 months to 10 years over different durations in Africa 2024.

The adopted GRADE (Grading of Recommendations Assessment, Development and Evaluation) as the method for assessing the quality of a body of evidence and for determining the direction and strength of the resulting recommendations. This generated evidence was evaluated the effectiveness and efficacy of chlorfenapyr, and piperonyl butoxide long-lasting insecticidal nets compared with pyriproxyfen long-lasting insecticidal nets for malaria control. The evidence generated found that piperonyl butoxide (PBO) long-lasting insecticidal nets effectively and efficaciously reduce indoor vector density, entomological inoculation rate, and sporozoite rate of malaria parasites compared to Pyriproxyfen (PPF) LLINs, but no significant difference was found in malaria infection reduction among children who use piperonyl butoxide (PBO) versus Pyriproxyfen (PPF) long-lasting insecticidal nets in Africa.

The study found that chlorfenapyr (CFP) long-lasting insecticidal nets (LLINs) are highly effective and superiorly efficacious in reducing malaria infection, case incidence, and anemia among children, as well as reducing mean indoor vector density, mean entomological inoculationrate, and sporozoite rate compared to pyriproxyfen (PPF) long-lasting insecticidal nets (LLINs) in Africa.

Critical appraisal of individual randomized control trials revealed that 100% of the studies scored high quality, and Cochrane methodology was used to assess the risk of bias and evaluate evidence quality, which was graded as high. This research provides a very good indication of the likely effect. The likelihood that the effect will be substantially different is low see details in Table 3.

**Table 3.**
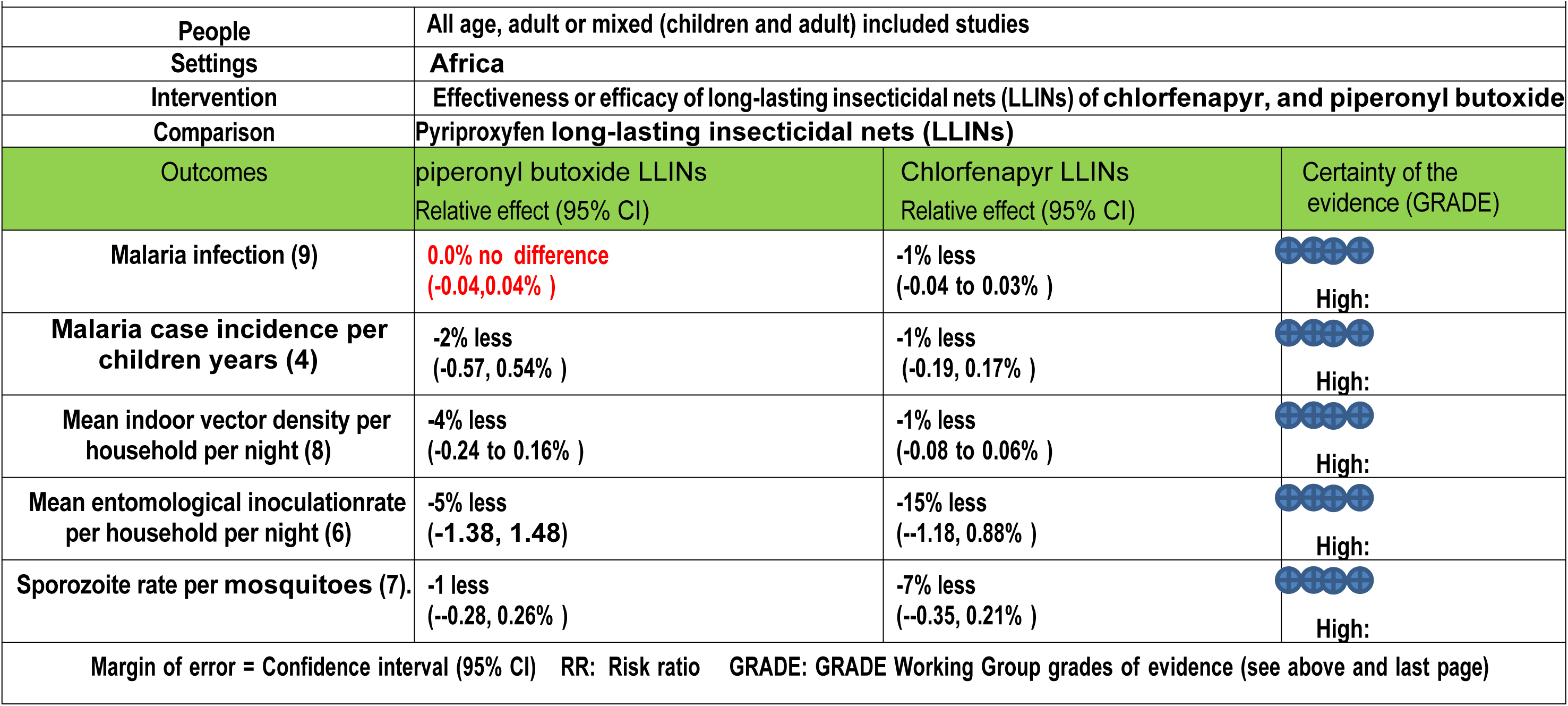
Evidence profile compared the effectiveness and efficacy of chlorfenapyr, and piperonyl butoxide long-lasting insecticidal nets with pyriproxyfen long-lasting insecticidal nets for malaria control in Africa.

|  |  |  |  |
| --- | --- | --- | --- |
| People | All age, adult or mixed (children and adult) included studies |  |  |
| Settings | Africa |  |  |
| Intervention | Effectiveness or efficacy of long-lasting insecticidal nets (LLINs) of chlorfenapyr, and piperonyl butoxide |  |  |
| Comparison | Pyriproxyfen long-lasting insecticidal nets (LLINs) |  |  |
| Outcomes | piperonyl butoxide LLINs<br>Relative effect (95% CI) | Chlorfenapyr LLINs<br>Relative effect (95% CI) | Certainty of the evidence (GRADE) |
| Malaria infection (9) | 0.0% no difference<br>(-0.04, 0.04%) | -1% less<br>(-0.04 to 0.03%) | High: |
| Malaria case incidence per children years (4) | -2% less<br>(-0.57, 0.54%) | -1% less<br>(-0.19, 0.17%) | High: |
| Mean indoor vector density per household per night (8) | -4% less<br>(-0.24 to 0.16%) | -1% less<br>(-0.08 to 0.06%) | High: |
| Mean entomological inoculation rate per household per night (6) | -5% less<br>(-1.38, 1.48) | -15% less<br>(-1.18, 0.88%) | High: |
| Sporozoite rate per mosquitoes (7). | -1 less<br>(-0.28, 0.26%) | -7% less<br>(-0.35, 0.21%) | High: |
| Margin of error = Confidence interval (95% CI) RR: Risk ratio GRADE: GRADE Working Group grades of evidence (see above and last page) |  |  |  |

## Discussion

This systematic review and meta-analysis aims to inform governments, policymakers, health professionals, and populations at risk of malaria about the effectiveness and efficacy of long-lasting insecticidal nets (LLINs) and evaluate changes in LLIN effectiveness and efficacy compared to pyrethroid-only LLINs. The study also compares the effectiveness of chlorfenapyr and piperonyl butoxide LLINs compared to pyriproxyfen LLINs for malaria control over time. Authors identified 11 RCTs that evaluated effectiveness and efficacy of different long-lasting insecticidal nets (LLINs) as malaria prevention and control among children in Africa. This evidence synthesis adopted GRADE (Grading of Recommendations Assessment, Development and Evaluation) as the method for assessing the quality of a body of evidence and for determining the direction and strength of the resulting recommendations. Included studies compared the performance of pyriproxyfen, chlorfenapyr, and piperonyl butoxide long-lasting insecticidal nets against the pyrethroid-only LLINs.

The study revealed that chlorfenapyr long-lasting insecticidal nets (LLINs) among children in the treatment groups provided significantly better protection against malaria compared to the pyrethroid-only LLINs. On the other hand malaria infection risk reduction among children using the Pyriproxyfen or Piperonyl butoxide LLINs resulted no significant difference compared to the pyrethroid-only LLINs.

More specifically, unlike the Pyriproxyfen or Piperonyl butoxide, the chlorfenapyr long-lasting insecticidal nets (LLINs) provided a significant advantage including reduction in malaria infection, the sporozoites rate and associated anemia compared to the standard pyrethroid-only group. The intervention resulted protection against malaria by reducing the risk of malaria infection by 10%. Besides, the intervention reduced the sporozoite rate by 5% and anemia by 1% as compared to standard or pyrethroid-only. Moreover, this study determined that post intervention effectiveness and efficacy of chlorfenapyr long-lasting insecticidal nets (LLINs) in malaria infection risk reduction in children over different durations (6, 12, 18, 24, and 36 months) was reduced by 1% as compared to the standard or control of the pyrethroid-only group.

Evidence shows that prevention and reduction of Malaria transmission can be done by vector control strategies which include insecticide-treated mosquito nets (74). Bed nets treated with an insecticide are believed to be much more protective than untreated nets (4). However, we found that not all LLINs- Pyriproxyfen or Piperonyl butoxide LLINs are effective. The finding is crucial as the vast majority of malaria cases and deaths are found in the WHO African Region and the malaria impact continues to be a global challenge (75).

The reason why some of the LLINs appeared less effective than the standard could be explained in several ways. There is a reason to believe that resistant malaria species can infect children despite the bed nets (74). Outdoor malaria transmission or poor utilization of the net could compromise the effectiveness and efficacy of the LLINs (76). This implies that the distribution of these LLINs should be carefully thought and further studies need to be carried out to uncover the main reasons.

As mosquitoes in many areas are now resistant to Pyriproxyfen or Piperonyl butoxide, nets treated with other active ingredients are needed to control malaria. On the other hand the recent WHO recommendation encourage the use of the new classes of dual ingredient ITNs with different modes of action- Pyrethroid-chlorfenapyr nets combine a pyrethroid and a pyrrole insecticide to enhance the killing effect of the net and Pyrethroid-pyriproxyfen nets combine a pyrethroid with an insect growth regulator (IGR) that disrupts mosquito growth and reproduction[6]. Based on our study the chlorfenapyr long-lasting insecticidal nets (LLINs) can still be considered.

Evolutionary theory reveals widespread resistance against insecticides, particularly pyrethroids, which were once used in bed nets to kill mosquitoes. This resistance was first detected in Côte d’Ivoire in 1993 and is now widespread throughout the region (74).

Some net products use piperonyl butoxide (PBO) in combination with pyrethroid insecticide to manage resistance, but the WHO does not consider these nets effective tools (4)

The WHO has released recommendations for two new insecticide-treated nets: Pyrethroid-chlorfenapyr nets, which combine pyrethroid and chlorfenapyr insecticides to enhance net killing, and Pyrethroid-pyriproxyfen nets, which use a pyrethroid and an insect growth regulator to disrupt mosquito growth and reproduction.

The World Health Organization (WHO) has issued a strong recommendation for the use of pyrethroid-chlorfenapyr ITNs over pyrethroid-only nets to prevent malaria in adults and children in areas with pyrethroid resistance. However, the WHO has also issued a conditional recommendation for the use of pyrethroid-chlorfenapyr ITNs instead of pyrethroid-PBO nets. Additionally, the WHO has issued a conditional recommendation against the use of pyrethroid-pyriproxyfen nets over pyrethroid-PBO nets (4). Thus our finding further justify and support the recommendation made by WHO.

The evidence generated found that piperonyl butoxide (PBO) long-lasting insecticidal nets effectively and efficaciously reduce indoor vector density, entomological inoculation rate, and sporozoite rate of malaria parasites compared to Pyriproxyfen (PPF) LLINs, but no significant difference was found in malaria infection reduction among children who use piperonyl butoxide (PBO) versus Pyriproxyfen (PPF) long-lasting insecticidal nets in Africa.These findings are strongly supported by previous study done revealed that in highly pyrethroid-resistant areas, unwashed pyrethroid-PBO nets led to higher mosquito mortality compared to unwashed standard-LLINs (risk ratio (RR) 1.84, 95% CI 1.60 to 2.11; 14,620 mosquitoes, 5 trials, 9 comparisons; high-certainty evidence) and lower blood feeding success (RR 0.60, 95% CI 0.50 to 0.71; 14,000 mosquitoes, 4 trials, 8 comparisons; high-certainty evidence)(77).

The study found that chlorfenapyr (CFP) long-lasting insecticidal nets (LLINs) are highly effective and superiorly efficacious in reducing malaria infection, case incidence, and anemia among children, as well as reducing mean indoor vector density, mean entomological inoculation rate, and sporozoite rate compared to pyriproxyfen (PPF) long-lasting insecticidal nets (LLINs) in Africa. This is supported by previous evidence sleeping under a Chlorfenapyr- PY LLIN offered superior protection against malaria infection (12). The U.S. President’s Malaria Initiative, one of its five intervention is to scale up access to and use of Long- lasting insecticide-treated nets in 27 countries in sub-Saharan Africa (78). Thus this finding may inform the selection of types of LLITNs and the need to focus on monitoring.

Sahel Malaria Elimination Initiative (SaME Initiative)-aim to accelerate towards the attainment of malaria elimination goals by 2030 in the Sub-region (79). This could be realized when latest evidences are considered and this study could serve that and help planners to effectively base their plan on evidence like both CFP & PBO effectively and efficaciously reduce malaria infection, case incidence, and anemia among children, indoor vector density, entomological incubation rate, and sporozoite rate reduction in Africa.

## Conclusion

The adopted GRADE (Grading of Recommendations Assessment, Development and Evaluation) as the method for assessing the quality of a body of evidence and for determining the direction and strength of the resulting recommendations. Critical appraisal of individual randomized control trials revealed that 100% of the studies scored high quality, and Cochrane methodology was used to assess the risk of bias and evaluate evidence quality, which was graded as high. This research provides a very good indication of the likely effect. The likelihood that the effect will be substantially different is low.

This generated evidence was evaluated the effectiveness and efficacy of pyriproxyfen, chlorfenapyr, and piperonyl butoxide long-lasting insecticidal nets against the pyrethroid-only LLINs.

This study found that PYR-only LLINs (control arm) had higher pooled prevalence of malaria infection, case incidence, anemia, mean indoor vector density, inoculation rate, and sporozoite rate as compared to intervention group (PPF, CFP, and PBO LLINs

This study also evaluated the effectiveness and efficacy of chlorfenapyr, and piperonyl butoxide long-lasting insecticidal nets compared to pyriproxyfen LLINs.

The study found that chlorfenapyr (CFP) long-lasting insecticidal nets (LLINs) are highly effective and superiorly efficacious in reducing malaria infection, case incidence, and anemia among children, as well as reducing mean indoor vector density, mean entomological inoculation rate, and sporozoite rate compared to pyriproxyfen (PPF) long-lasting insecticidal nets (LLINs) in Africa.

### Recommendation and policy implication

In conclusion this study found that PYR-only LLINs (control arm) had higher pooled prevalence of malaria infection, case incidence, anemia, mean indoor vector density, inoculation rate, and sporozoite rate as compared to intervention group (PPF, CFP, and PBO) LLINs. This research provides a very good indication of the likely effect. The likelihood that the effect will be substantially different is low. Therefore, policymakers and health planners should give a great deal of emphasis on addressing the effectiveness, efficacy, and resistance management of long-lasting insecticidal nets (LLINs) as part of their current public health agenda to eliminate malaria by 2030.

Policymakers and health planners should prioritizing the effectiveness of long-lasting insecticidal nets (LLINs) in malaria prevention strategies, particularly for vulnerable populations, and resistance management strategies to maintain their efficacy in resource-limited countries.

Policymakers and health planners should give a great deal of emphasis on addressing the effectiveness, efficacy, and resistance management of long-lasting insecticidal nets (LLINs) as part of their current public health agenda to eliminate malaria by 2030.

Policymakers and health planners should prioritize evidence-based effectiveness and efficacy of long-lasting insecticidal nets (LLINs) for malaria control strategies particularly for vulnerable populations, and resistance management strategies to maintain their efficacy in resource-limited countries.

The policy recommendation based on this study is that net re-distribution cycles should be aligned with the lifespan of long-lasting insecticidal nets (LLINs) to maintain maximum coverage and impact, even though usage was low in this study.

Policymakers should replace pyrethroid-only long-lasting insecticidal nets with chlorfenapyr or piperonyl butoxide long-lasting insecticidal combinations and achieve universal access to more effective and efficacious long-lasting insecticidal nets preventing malaria.

For researchers- the finding may spark a number of research questions including the factors that leads to the resistance and the net effect introduced due to the resistance

Future research should focus on evaluating the effectiveness of pyriproxyfen long-lasting insecticidal nets as compared to chlorfenapyr and piperonyl butoxide long-lasting insecticidal nets in more realistic field conditions to better understand their true efficacy against mosquitoes.

## Data Availability

All data generated or analyzed during this study are included in this article and its supplementary information files.

## Ethical approval and consent to participate

Not applicable.

## Acknowledgement

Our acknowledgement goes to the authors of the primary studies included in this study.

## Author contributions

DBD conceptualized, designed, and drafted the manuscript. DBD and GFH performed article search, data extraction, and quality assessment. DBD and FTT conducted data analysis and wrote the manuscript.TD and FTT have reviewed the manuscript. All authors read, reviewed, and approved the final manuscript.

